# Genome-wide assessment identifies novel runs of homozygosity linked to Parkinson’s disease etiology across diverse ancestral populations

**DOI:** 10.1101/2025.05.21.25328076

**Authors:** Kathryn Step, Carlos F. Hernández, Esraa Eltaraifee, Ana Jimena Hernández-Medrano, Pin-Jui Kung, Miriam Ostrožovičová, Alexandra Zirra, Eduardo Pérez-Palma, Niccolò E. Mencacci, Ignacio J. Keller Sarmiento, Huw R. Morris, Ignacio F. Mata, Global Parkinson’s Genetics Program (GP2), Juliana Acosta-Uribe, Zih-Hua Fang, Sara Bandres-Ciga

## Abstract

**Objective:** We conducted the first large-scale, multi-ancestral investigation of Parkinson’s disease (PD) to examine the impact of genome-wide homozygosity on disease risk and age at onset. Using genotyping, imputed, and whole-genome sequencing (WGS) data from 16,599 PD cases and 13,585 controls across nine ancestral populations from the Global Parkinson’s Genetics Program, we aimed to identify novel regions of homozygosity contributing to PD heritability.

**Methods:** We analyzed runs of homozygosity (ROHs) for total length (S_ROH_), number (N_ROH_), average length (AV_ROH_), and genomic inbreeding coefficient (F_ROH_). ROHs were intersected with known PD, pallido-pyramidal syndrome, and atypical parkinsonism gene regions and risk loci to assess pleomorphic or pleiotropic contributions. Homozygosity mapping identified ROH overlaps in families, consanguineous individuals, and early-onset PD (EOPD) cases.

**Results:** Significant differences in S_ROH_, AV_ROH_, N_ROH_, and F_ROH_ were observed between cases and controls across multiple ancestral groups, persisting after excluding known PD-associated recessive genes. Our analysis revealed distinct patterns of ROH enrichment associated with age at onset, suggesting recessive genetic modifiers of PD across diverse ancestral backgrounds. Homozygosity mapping identified 672 case-exclusive ROH pools, 21 prioritized variants, and 1,300 ROHs enriched in cases. Finally, 167 ROHs in consanguineous individuals and EOPD overlapped known PD and risk loci.

**Interpretation:** Our findings suggest that ROH regions contribute to PD heritability in a global context, with a portion attributed to recessive allelic architecture. We developed an open-science framework for unbiased homozygosity mapping. Future studies should use larger, diverse cohorts and WGS data to uncover rare recessive variants linked to PD susceptibility.

## Introduction

Historically, Parkinson’s disease (PD) was viewed as a sporadic condition attributed to aging or environmental factors. However, growing evidence points to PD as a multifactorial disorder (Bandres-Ciga *et al*., 2020) resulting from complex gene-gene and gene-environment interactions (Khani *et al*., 2024). There is an increased global large-scale effort to diversify PD research by investigating the genetics underlying disease etiology across ancestral groups (Foo *et al*., 2020; Global Parkinson’s Genetics Program, 2021; Loesch *et al*., 2021; Rizig *et al*., 2023; Kim *et al*., 2024).

The genetic risk for PD spans the etiological spectrum, ranging from rare, highly penetrant disease-causing variants to common genetic variation that increases disease risk. At least 21 genes have been reported to cause monogenic forms of PD, but many have yet to be confirmed through replication and functional validation studies (Blauwendraat, Nalls and Singleton, 2020; Lange *et al*., 2022). In cases where PD is caused by pathogenic variants in single genes (monogenic PD), the disease can be inherited in either an autosomal dominant or recessive manner, and it is commonly familial (Jia, Fellner and Kumar, 2022). However, monogenic PD mutations may also occur in sporadic cases, though a significant portion of the disease heritability is still unexplained. Over 100 susceptibility loci have been associated with an increased risk for PD (Kim *et al*., 2024). Additionally, approximately 3-5% of sporadic PD cases have been linked to an autosomal recessive mode of inheritance through disease-causing variants in the *PRKN*, *PINK1*, and *DJ-1* genes (Klein and Westenberger, 2012; Ahfeldt *et al*., 2020). Autosomal recessive pathogenic variants are particularly associated with early-onset PD (EOPD), defined as onset before 45 or up to 50 years of age (Lesage *et al*., 2020; Mehanna *et al*., 2022).

Pleomorphism is commonly observed in genetics, where genetic risk within a specific region often involves a spectrum of allele frequency and effect sizes, ultimately encompassing high-risk rare variants and low-risk common variants potentially linked to disease (Singleton and Hardy, 2011). For instance, common and rare genetic variation in *VPS13C*, have been found to be associated with PD through both GWAS as well as sequencing approaches aimed to unravel patterns of recessive inheritance (Lesage *et al*., 2016; Nalls *et al*., 2019). Typical pleomorphic loci exhibit structural, coding, and non-coding variants, each contributing to different levels of PD risk. Further investigation within reported PD risk loci is needed to uncover additional pleomorphic effects with potential genes involved in recessive patterns of inheritance. In contrast, pleiotropy occurs when a single gene influences multiple phenotypic traits or functions, such as PD, atypical parkinsonism, and pallido-pyramidal syndrome.

The presence of long regions of consecutive homozygous genotypes, known as runs of homozygosity (ROHs) (Szpiech *et al*., 2013), is the result of inbreeding and recessive modes of inheritance (Moreno-Grau *et al*., 2021). Although ROHs arise from shared common ancestors, they are found commonly in outbred populations and are more common in older populations (Nalls *et al*., 2009). ROHs can result from population bottlenecks, cultural practices that promote consanguineous marriages, and natural selection resulting in elevated frequencies of haplotypes surrounding a favorable allele (Broman and Weber, 1999; Hildebrandt *et al.*, 2009; Wang *et al*., 2009). Characteristics of ROH, such as their number and burden, provide insights into population structure and history. Larger and admixed populations tend to have shorter and fewer ROHs, whereas bottlenecked, consanguineous, and isolated populations have longer ROHs (Ceballos *et al*., 2018), as highlighted in **Figure 1**. This may reflect the contribution to disease risk of specific recessive loci and risk haplotypes or the cumulative effect of multiple regions spanning ROHs (Ghani *et al*., 2015). Detecting regions where individuals share the same alleles can be indicative of genetic relatedness or common ancestry. This is achieved by analyzing allele segregation within families and identifying shared segments that may harbor potential disease-causing variants across different ancestries. To further identify and investigate genes and/or variants underlying autosomal recessive diseases, approaches like homozygosity mapping using closely related populations can be employed (Vahidnezhad *et al*., 2018).

**Figure 1:**
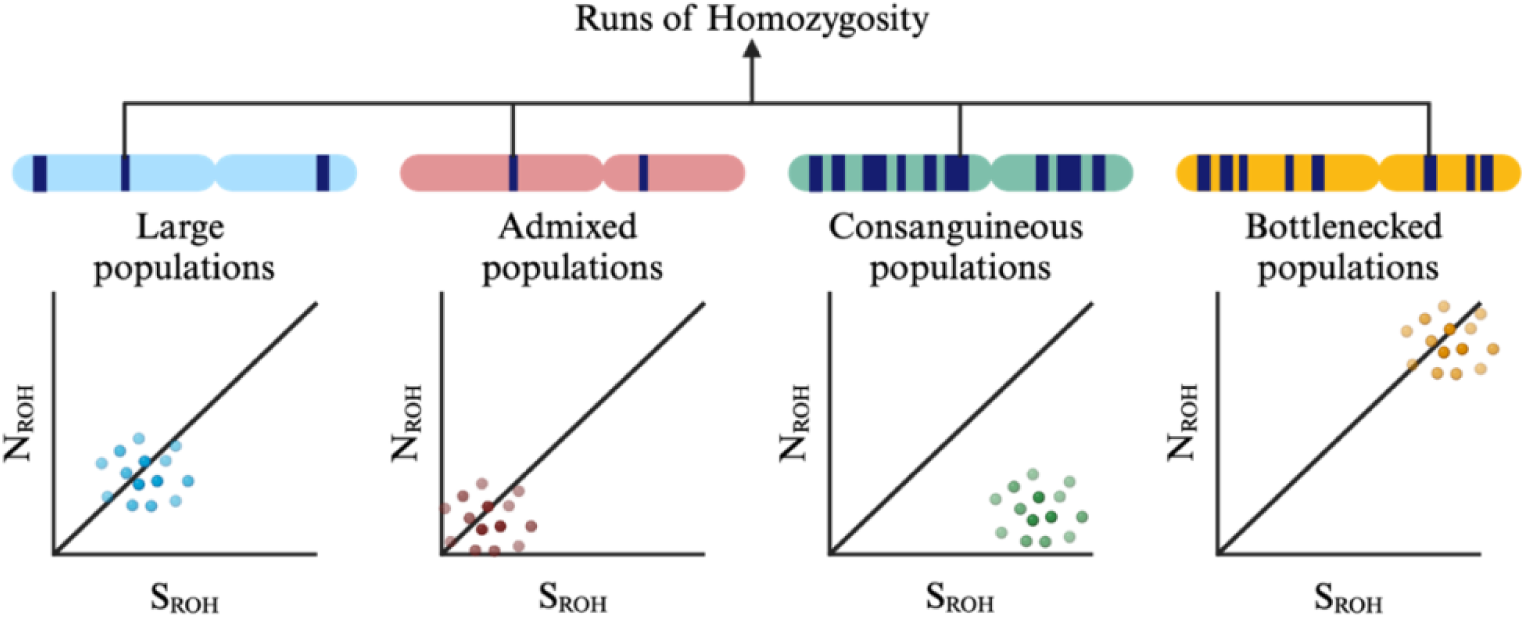
Overview of runs of homozygosity trends observed in different populations. N_ROH_ = total number of runs of homozygosity, S_ROH_ = total length of runs of homozygosity.

Here, we aim to conduct the first large-scale multi-ancestral study in PD to investigate the impact of genome-wide homozygosity on disease risk and age at onset (AAO). We comprehensively assess the homozygosity spectrum across nine diverse ancestry populations by leveraging the largest multi-ancestry resources of genotyping imputed and WGS individual-level data available in the PD field, to potentially nominate novel regions of homozygosity in a global setting. Our goal is to investigate regions of ROHs enriched in PD cases to uncover novel recessive effects that may explain a proportion of PD heritability.

## Methods

A rationale overview of our extensive ROH characterization and subsequent prioritization is illustrated in **Figure 2**.

**Figure 2:**
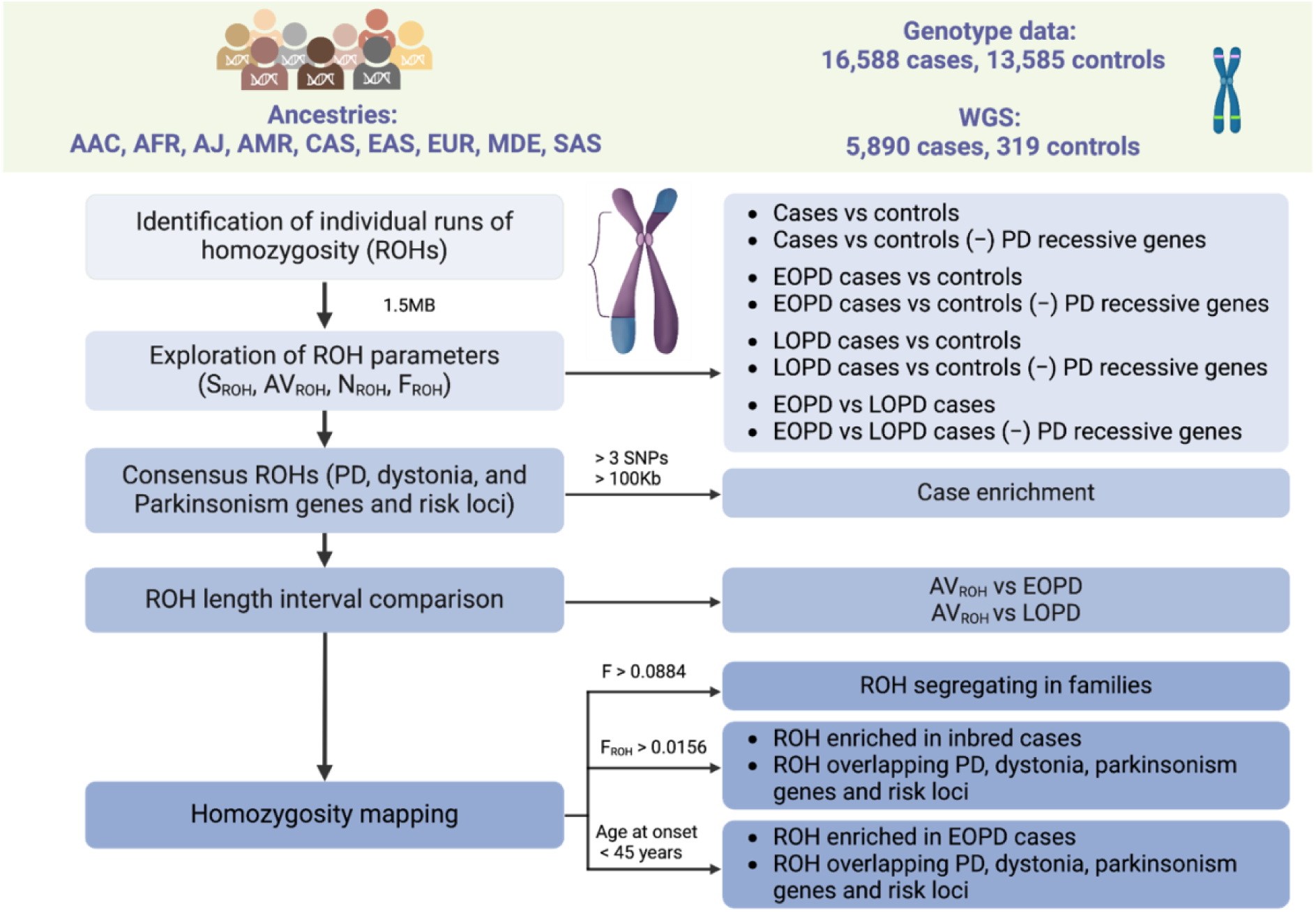
Workflow and rationale summary. AAC = African Admixed, AFR = African, AJ = Ashkenazi Jewish, AMR = American Admixed, AVROH = average runs of homozygosity length, CAS = Central Asian, EAS = East Asian, EOPD = early-onset Parkinson’s disease, EUR = European, F= inbreeding coefficient, FROH = run of homozygosity based estimates for the inbreeding coefficient, KB = kilobase, LOPD = late-onset Parkinson’s disease, MB = megabase, MDE = Middle-Eastern, NROH, number of runs of homozygosity, ROH = runs of homozygosity, SAS = South Asian, SNPs = single nucleotide polymorphisms, SROH = total length of runs of homozygosity, - = excluding.

### Demographic and clinical characteristics

Study participants were recruited as part of the Global Parkinson’s Genetics Program (GP2), as previously reported (Global Parkinson’s Genetics Program, 2021). Individual-level data was obtained from GP2’s Data Release 6 (Leonard *et al*., 2023). We performed data analysis on nine ancestral groups, including African Admixed (AAC), African (AFR), Ashkenazi Jewish (AJ), American Admixed (AMR), Central Asian (CAS), East Asian (EAS), European (EUR), Middle-Eastern (MDE), and South Asian (SAS) populations. The Finnish group was excluded due to the small sample size. The data analysis included 30,173 individuals comprising 16,588 PD cases and 13,585 controls, including related individuals (**Supplementary Table 1**). GP2’s Data Release 8 includes additional WGS data for the nine ancestral groups, which we utilized to further explore and prioritize ROHs (Leonard *et al*., 2024). The WGS data analysis included 6,209 individuals comprising 5,890 PD cases and 319 controls (**Supplementary Table 2**). We prioritized WGS data from GP2 over that from the Accelerating Medicines Partnership program for PD (AMP-PD) because the first is enriched with familial and early-onset cases, whereas AMP-PD predominantly includes sporadic late-onset cases.

### Genotyping, quality control, ancestry predictions, and imputation

Genotyping was performed by GP2 using the NeuroBooster Array (v.1.0, Illumina, San Diego, CA) <u>(Bandres-Ciga *et al*., 2023)</u>. Raw genotyping data was processed using the GenoTools pipeline (https://github.com/GP2code/GenoTools), as previously described (Koretsky *et al*., 2022). Briefly, all samples underwent standardized quality control (QC), where samples were excluded if the call rate was less than 95%, the genetically determined sex did not match the clinical records, or an excess heterozygosity rate was detected (|F| >0.25) (Vitale *et al*., 2025). As previously described, ancestry estimates were performed using a standardized protocol. Samples were subsetted according to ancestry estimates using reference panels from the 1000 Genomes Project (1000 Genomes Project Consortium *et al*., 2015), an AJ population dataset (Bray *et al*., 2010), and the Human Genome Diversity Project (Siva, 2008). Imputation was performed using the TOPMed-r3 panel (Das *et al*., 2016). The imputed files were pruned for an imputation R^2^ of 0.3, and no minor allele count threshold was applied.

### Inbreeding coefficient and identity by descent analyses

Genotyping data was used to calculate the kinship coefficient with KING v.2.3 (Manichaikul *et al*., 2010). Post-imputed data, separated by chromosomes, underwent further QC using PLINK v.2.0 (Chang *et al*., 2015). This included removing related individuals with a kinship coefficient threshold of 0.0884 or higher, indicating second-degree relatives or closer (Manichaikul *et al*., 2010). After relatedness analysis, our dataset contained 29,673 unrelated individuals, of which 16,349 were PD cases, and 13,324 were controls. Variants were filtered for a minor allele frequency of <5%, and genotypes were pruned using the --indep-pairwise flag at a window size of 50kb, step size of 5, and r^2^ threshold of 0.5. Thereafter, chromosome binary files were merged into one post-QC file. Finally, the level of inbreeding was investigated using the --het flag, which was used to compare F coefficients across ancestries. Identity-by-descent was computed using the -- genome flag.

### Estimation of runs of homozygosity

#### Individual runs of homozygosity calling

Individual calling of ROHs was carried out using PLINK v.1.9 (Purcell *et al*., 2007). Detection of ROH was performed for each of the nine ancestral populations. To investigate homozygosity across the genome, we implemented a sliding window in a stepwise approach with predefined parameters. For our study, we used a sliding window of 50 SNPs and 1,500kb length as previously described (Simón-Sánchez *et al*., 2012). The minimum number of SNPs in ROH regions was 100, with an allowed threshold of one heterozygous SNP and five missing SNPs. A region was considered a potential ROH if each SNP was covered by at least 5% of the homozygous sliding window (Simón-Sánchez *et al*., 2012; Moreno-Grau *et al*., 2021), allowing a shorter ROH to be captured. The ROHs cut-off was set at >1.5Mb, as longer ROHs are more informative of inbreeding and disease association (Moreno-Grau *et al*., 2021), and ROHs <1.5Mb tend to reflect linkage disequilibrium patterns and population substructure (McQuillan *et al*., 2008). We set a maximum threshold of 1Mb distance between two consecutive SNPs and a minimum density threshold of one SNP in the 50kb length of the genome as described in **Supplementary Table 3**.

#### General homozygosity metrics assessment

To study ROHs between cases and controls for each ancestral group, we examined the following homozygosity metrics: (1) the total length of ROHs (S_ROH_), (2) the number of ROHs (N_ROH_), (3) the average ROHs length (AV_ROH_), and (4) ROHs-based estimates for the inbreeding coefficient (F_ROH_). The S_ROH_ divided by the N_ROH_ per individual gives the AV_ROH_, while the proportion of ROH in the autosomal genome per individual is represented by F_ROH_. For this, the S_ROH_ was divided by the total length of the autosomal genome (2,875,001.522kb), according to the GRCh38.p14 assembly (National Institutes of Health, 2022). The subset of individuals with consanguinity was detected using F_ROH_. The threshold to indicate individuals with consanguinity and outbred individuals corresponds to a second-degree relation set at F_ROH_ >0.0156 <u>(Bittles and Black, 2010; Ceballos, Hazelhurst, and Ramsay, 2019)</u>. The four metric equations and additional information can be found in **Supplementary Material**.

#### Association of overlapping runs of homozygosity parameters with Parkinson’s disease risk and onset

Firstly, we assessed the sample distribution of each ancestry for the four homozygosity parameters (S_ROH_, N_ROH_, AV_ROH_, and F_ROH_). A Scree plot for each ancestral group was generated to determine the number of principal components (PCs) to retain as covariates for the analysis (**Supplementary Figure 1**). Secondly, PD cases were subsetted into early onset (EOPD onset <45 years old) and late-onset (LOPD onset ≥45 years old) according to their reported AAO. Using the calculated mean age, the age was imputed for each ancestral group separately for cases missing AAO in their clinical information (**Supplementary Table 4**).

Logistic regression models were conducted for (1) cases *versus* controls, (2) EOPD *versus* controls, (3) LOPD *versus* controls, (4) EOPD cases *versus* LOPD cases, (5) cases *versus* controls excluding known recessive PD genes (*PRKN*, *DJ-1*, and *PINK1*), (6) EOPD cases *versus* controls excluding known recessive PD genes, (7) LOPD cases *versus* controls excluding known recessive PD genes, and (8) EOPD cases *versus* LOPD cases excluding known recessive PD genes. Linear regression models were performed for ROH *versus* AAO, of which one model included known recessive PD genes, while the other excluded them. Further analysis was performed using the four homozygosity parameters comparing controls to cases stratified by the following groups: <35 years, 35-44 years, 45-54 years, 55-65 years, and >65 years. Subsequently, a logistic regression model was used to examine the association between PD risk and consensus ROHs. The model was adjusted for age, sex, and five PCs, as indicated by the scree plots, to ensure the majority of variance was adequately captured while minimizing the risk of overfitting (**Supplementary Figure 1**). Bonferroni correction was used to adjust the regression-based results for multiple testing (0.05/9, the number of ranges).

#### Runs of homozygosity spanning Parkinson’s disease, pallido-pyramidal syndrome, and atypical parkinsonism genes and risk loci

Subsequently, ROHs were further investigated for known PD, pallido-pyramidal syndrome, and atypical parkinsonism gene regions and risk loci defined from GWAS loci (Nalls *et al*., 2019; Foo *et al*., 2020; Loesch *et al*., 2021; Rizig *et al*., 2023; Kim *et al*., 2024) using an approximate 1Mb window upstream or downstream from the GWAS hits and genes (**Supplementary Table 5**) to account for overlapping genetic etiologies. The equal proportions test was utilized to assess whether the proportions (probabilities of success) in both the case and control groups were identical or as specified by the null hypothesis.

#### Interval comparison for runs of homozygosity lengths

Thresholds ranging from >2Mb to >10Mb in 1Mb increments were used to further investigate various ROH lengths for each ancestry. This was done to provide insight into the origin and timing of ROHs in different populations (Ceballos *et al*., 2018). A logistic regression model was used to examine the association between AV_ROH_ and case status. The model was adjusted for age at sampling, sex, and five PCs. Bonferroni correction was used to correct the regression-based results for multiple testing by dividing 0.05 by nine (the number of ranges). Moreover, this analysis was performed to examine the association between AV_ROH_ and EOPD case status as well as AV_ROH_ and LOPD case status.

### Homozygosity mapping

We implemented homozygosity mapping to identify known and potentially novel extended genomic regions that differed between cases and controls. Individual calling of ROHs was carried out using PLINK v.1.9 (Purcell *et al*., 2007) using the aforementioned parameters. To identify highly penetrant recessive variants, we prioritized ROHs not carried by controls and nominated rare coding variants in the ROH region detected in families, PD cases with consanguinity, and EOPD on those individuals with WGS data available. Variants were annotated using Ensembl’s Variant Effect Predictor v.110 (McLaren *et al*., 2016).

#### Runs of homozygosity overlaps segregating in families

We performed QC on the genotyping data, including related individuals, to ensure high-quality markers were retained by filtering for genotype data missingness at 5% and Hardy-Weinberg Equilibrium at 1×10^−10^. Relationship inference was performed using KING v.2.3 (Manichaikul *et al*., 2010), where relatedness was defined with a threshold of 0.0884. We searched for overlapping ROH that have pairwise allelic matches shared by PD cases within the same families based on kinship inference and using genotyping data (**Supplementary Table 6**). Variants in ROHs were extracted from WGS and prioritized based on the credible genetic ancestry group allele frequency (Chen *et al*., 2024) in the gnomAD v.4.1.0 database, restricting minor allele frequency to ≤0.01 as well as variant consequence.

#### Runs of homozygosity overlaps enriched in Parkinson’s disease cases with consanguinity

Individuals with consanguinity were subsetted using F_ROH_ >0.0156. Here, the individual calling for ROHs was conducted using the post-QC imputed data. We examined overlapping ROHs and prioritized unique consensus regions greater than 100kb and 100 SNPs to find ROHs enriched in individuals with consanguinity. These thresholds were chosen to identify real ROHs, as sparse marker density may lead to false positives. Variants were prioritized using the above mentioned method for those individuals with WGS data available. Subsequently, a logistic model was used to examine the association of ROHs enriched in individuals with consanguinity as well as ROHs enriched in these individuals overlapping known PD, pallido-pyramidal syndrome, and atypical parkinsonism gene regions and risk loci. The model was adjusted for age, sex, and five PCs. Bonferroni correction was used to adjust the regression-based results for multiple testing.

#### Runs of homozygosity overlaps enriched in early-onset Parkinson’s disease cases

EOPD cases defined by age at onset <45 years old were subsetted from the post-QC imputed data, and ROH mapping was performed. The overlapping ROHs were examined, and unique consensus regions greater than 100kb and 100 SNPs were prioritized to find ROHs enriched in EOPD cases. Variants were prioritized using the above mentioned method for those individuals with WGS data available. Subsequently, a logistic regression model was used to examine the association of ROH enriched in EOPD cases as well as regions enriched in EOPD cases that overlap known PD, pallido-pyramidal syndrome, and atypical parkinsonism gene regions and risk loci. The model was adjusted for age, sex, and five PCs. Bonferroni correction was used to adjust the regression-based results for multiple testing by dividing 0.05 by the number of enriched ROHs, as indicated in **Table 5**.

## Results

### Multi-ancestry genome-wide assessment shows increased homozygosity in Parkinson’s disease

Four ROH parameters (S_ROH_, N_ROH_, AV_ROH_, and F_ROH_) were examined in 29,673 unrelated individuals from the GP2 initiative (**Table 1, Supplementary Table 7, Supplementary Figure 2**), covering nine ancestries. We detected 102,681 ROHs greater than 1.5Mb where approximately 90% of individuals—both cases and controls—had at least one ROH of 1.5Mb or longer that met the calling criteria, indicating a high prevalence of valid ROH calls, with only 10% lacking ROHs of this length. The overall mean N_ROH_ for the study cohort was 5.5 ± 3. **Supplementary Figure 3** illustrates the relationships between the mean N_ROH_ and S_ROH_ for each ancestral group and each individual included in the study. The MDE group exhibited the highest mean N_ROH_ (10.0 ± 14.0), and the AAC group showed the lowest mean N_ROH_ (1.8 ± 3.0). For S_ROH_, SAS showed the highest mean with 25.624 ± 48.145Mb, and the lowest mean S_ROH_ was observed in the AAC group (4.842 ± 12.144Mb). Moreover, relationships between N_ROH_ and the S_ROH_ for each ancestry are shown in **Supplementary Figure 4**, while relationships between N_ROH_ and AV_ROH_ are shown in **Supplementary Figure 5**.

**Table 1:** Regression results for measures of genome-wide homozygosity in Parkinson’s disease.

| Ancestry | $S_{ROH}$ | | $AV_{ROH}$ | | $N_{ROH}$ | | $F_{ROH}$ | |
| --- | --- | --- | --- | --- | --- | --- | --- | --- |
| | Beta<br>(mean ± SE) | p-value | Beta<br>(mean ± SE) | p-value | Beta<br>(mean ± SE) | p-value | Beta<br>(mean ± SE) | p-value |
| AAC | 1.31E-05 ±<br>6.16E-06 | **0.034 \*** | 1.76E-04 ±<br>6.23e-05 | **0.005 \*\*** | 0.048 ±<br>0.027 | 0.077 | 37.8 ±<br>17.8 | **0.034 \*** |
| AFR | 7.16E-06 ±<br>3.45E-06 | **0.038 \*** | 1.19E-04 ±<br>5.1E-05 | **0.019 \*** | 0.022 ±<br>0.012 | 0.054 | 20.7 ±<br>9.96 | **0.038 \*** |
| AJ | 2.03E-06 ±<br>2.66E-06 | 0.445 | 6.8E-05 ±<br>6.77E-05 | 0.316 | 2.65E-03 ±<br>0.013 | 0.836 | 5.85 ±<br>7.67 | 0.445 |
| AMR | 4.35E-06 ±<br>4.61E-06 | 0.345 | -8.6E-05 ±<br>7.64E-05 | 0.256 | 0.023 ±<br>0.022 | 0.296 | 12.6 ±<br>13.3 | 0.345 |
| CAS | -2.69E-07 ±<br>2.86E-06 | 0.925 | -1.57E-04 ±<br>1.21E-04 | 0.194 | -8.86E-04 ±<br>0.010 | 0.930 | -0.775 ±<br>8.26 | 0.925 |
| EAS | -9.94E-06 ±<br>5.41E-06 | 0.066 | -4.94E-05 ±<br>6.26E-05 | 0.430 | -0.0255 ±<br>0.014 | 0.059 | -28.7 ±<br>15.6 | 0.066 |
| EUR | -8.48E-06 ±<br>1.68E-06 | **4.44E-07**<br>*** | -6.04E-05 ±<br>2E-05 | **2.57E-03**<br>** | -0.0227 ±<br>0.005 | **1.93E-05**<br>*** | -24.5 ±<br>4.85 | **4.44E-07**<br>*** |
| MDE | 4.04E-05 ±<br>2.1E-05 | 0.055 | 4.53E-04 ±<br>3.02E-04 | 0.134 | 0.115 ±<br>0.057 | **0.041 \*** | 117 ±<br>60.7 | 0.055 |
| SAS | -1.67E-05 ±<br>4.83E-06 | **5.44E-04**<br>** | -2.61E-04 ±<br>1.52E-04 | 0.086 | -0.0449 ±<br>0.013 | **7.81E-04**<br>** | -48.2 ±<br>13.9 | **5.44E-04**<br>*** |

Legend: AAC = African Admixed, AFR = African, AJ = Ashkenazi Jewish, AMR = American Admixed, $AV_{ROH}$ = average length of runs of homozygosity, CAS = Central Asian, EAS = East Asian, EUR = European, $F_{ROH}$ = runs of homozygosity-based estimates for inbreeding coefficient, MDE = Middle-Eastern, $N_{ROH}$ = number of runs of homozygosity, SAS = South Asian, SD = standard deviation, $S_{ROH}$ = total length of runs of homozygosity. p-values indicate the statistical significance as following $p < 0.001$ : \*\*\*, $p < 0.01$ : \*\*, $p < 0.05$ : \*, $p \geq 0.05$ : not significant

Cases had longer ROHs, longer average ROHs, and a higher proportion of consanguineous individuals as compared to controls in the AAC, AFR, EUR, and SAS groups. For N_ROH_, significant associations between cases and controls were seen in the EUR, MDE, and SAS groups. The statistically significant results remained unchanged after excluding the known recessive PD genes: *PRKN*, *DJ-1*, and *PINK1* (**Supplementary Table 8**). As expected, populations with smaller sample sizes display wider confidence intervals and less reliable results. The SAS (F_ROH_= 0.0088), MDE (F_ROH_= 0.0087), and AJ (F_ROH_= 0.0076) populations showed the highest F_ROH_, while AAC showed the lowest F_ROH_ at 0.0017 (**Supplementary Figures 6 to 8**).

We conducted linear regression analyses to examine the association between PD AAO and the four ROH parameters across the different ancestral groups. Significant associations were found in the AMR group across all four ROH parameters: S_ROH_, AV_ROH_, N_ROH_, and F_ROH_. Additionally, significant enrichment was observed for S_ROH_ in the EUR group, for AV_ROH_ in the AAC group, and for F_ROH_ in the EUR group. Detailed beta coefficients and p-values for each parameter and ancestral group are provided in **Supplementary Table 9**. No significance was observed for EUR across these parameters after excluding known recessive PD genes (**Supplementary Table 10**). Additional analysis assessed the four parameters in cases subsetted for the following age ranges: <35, 35-44, 45-54, 55-65, >65, as shown in **Supplementary Table 11**.

### Increased genomic homozygosity, excluding known recessive genes, suggests unknown recessive genetic factors contribute to Parkinson’s disease heritability

Additionally, we investigated the relationship between AAO for cases and the four ROH parameters to identify potential genetic onset modifiers, acknowledging that cases with AAO could also carry genetic mutations in known EOPD genes like *PRKN*, *PINK1*, and *DJ-1*. PD cases were subsetted into EOPD and LOPD where a logistic regression analysis was performed for each status *versus* controls (**Table 2**, **Supplementary Table 12**, **Supplementary Figure 9**). In summary, for S_ROH_, there was a statistically significant difference in the AFR group between EOPD cases and controls; moreover, significant associations between LOPD cases and controls were observed in the EUR, and MDE groups. A significant enrichment of AV_ROH_ was observed between EOPD cases and controls in the AAC group. An overrepresentation of N_ROH_ was seen in the AFR group between EOPD cases and controls, and statistically significant differences between LOPD cases and controls were seen in the EUR and MDE groups. For F_ROH_, the AFR group showed a statistically significant difference between EOPD cases and controls. In contrast, a significant enrichment between LOPD cases and controls was seen in the EUR, and MDE groups. There was no statistical significance for the AJ, AMR, and CAS groups. The association analysis was repeated after excluding known recessive PD genes (*PRKN*, *DJ-1*, and *PINK1*) from the dataset (**Supplementary Table 13**).

**Table 2:** Regression results for runs of homozygosity in cases subsetted for early-onset Parkinson’s disease and late-onset Parkinson’s disease versus controls.

| Ancestry | AAO | Number of cases (controls) | SROH |  | AVROH |  | NROH |  | FROH |  |
| --- | --- | --- | --- | --- | --- | --- | --- | --- | --- | --- |
| | | | Beta (mean ± SE) | p-value | Beta (mean ± SE) | p-value | Beta (mean ± SE) | p-value | Beta (mean ± SE) | p-value |
| AAC | EOPD | 21 (804) | 2.66E-05 ± 1.63E-05 | 0.103 | 6.27E-04 ± 2.05E-04 | **0.002** ** | 0.081 ± 0.073 | 0.263 | 76.7 ± 47 | 0.103 |
| | LOPD | 128 (804) | 1.26E-05 ± 8.02E-06 | 0.115 | 1.69E-04 ± 9.06E-05 | 0.062 | 0.036 ± 0.043 | 0.408 | 36.5 ± 23.2 | 0.115 |
| AFR | EOPD | 21 (1,631) | 1.54E-05 ± 7.45E-06 | **0.039 \*** | 3.46E-04 ± 2.71E-04 | 0.201 | 0.059 ± 0.028 | **0.036 \*** | 44.4 ± 21.5 | **0.039 \*** |
| | LOPD | 137 (1,631) | -1.34E-05 ± 1.26E-05 | 0.288 | 7.56E-07 ± 1.14E-04 | 0.995 | -0.024 ± 0.033 | 0.469 | -38.6 ± 36.4 | 0.289 |
| AJ | EOPD | 83 (458) | 6.7E-06 ± 8E-06 | 0.403 | 9.7E-05 ± 1.89E-04 | 0.607 | 0.026 ± 0.039 | 0.508 | 19.3 ± 23.1 | 0.402 |
| | LOPD | 719 (458) | 2.21E-06 ± 2.73E-06 | 0.418 | 8.38E-05 ± 7.22E-05 | 0.246 | 0.003 ± 0.012 | 0.780 | 6.38 ± 7.88 | 0.418 |
| AMR | EOPD | 91 (139) | 1.19E-05 ± 8E-06 | 0.138 | 5.73E-05 ± 1.21E-04 | 0.637 | 0.067 ± 0.041 | 0.103 | 34.3 ± 23.1 | 0.138 |
| | LOPD | 163 (139) | 3.31E-06 ± 6.43E-06 | 0.607 | -4.77E-05 ± 9.29E-05 | 0.608 | 0.022 ± 0.029 | 0.446 | 9.57 ± 18.6 | 0.606 |
| CAS | EOPD | 53 (267) | -4.73E-06 ± 7.69E-06 | 0.538 | -3.3E-04 ± 2.11E-04 | 0.118 | -0.016 ± 0.027 | 0.553 | -13.6 ± 22.2 | 0.538 |
|  | LOPD | 57 (267) | -3.34E-05 ± 2.43E-05 | 0.170 | -2.93E-05 ± 2.45E-04 | 0.905 | -0.097 ± 0.062 | 0.115 | -96.3 ± 70.1 | 0.170 |
| EAS | EOPD | 243 (2,302) | 4.36E-06 ± 1.29E-05 | 0.736 | 2.98E-05 ± 1.57E-04 | 0.849 | 0.015 ± 0.031 | 0.637 | 12.6 ± 37.3 | 0.735 |
|  | LOPD | 455 (2,302) | -2.16E-05 ± 1.21E-05 | 0.074 | -1.96E-04 ± 1.06E-04 | 0.066 | -0.050 ± 0.028 | 0.074 | -62.3 ± 34.8 | 0.074 |
| EUR | EOPD | 1,042 (7,505) | -1.67E-06 ± 3.55E-06 | 0.638 | -9.24E-05 ± 5.12E-05 | 0.071 | -0.009 ± 0.013 | 0.476 | -4.82 ± 10.3 | 0.638 |
|  | LOPD | 6,400 (7,505) | -7.98E-06 ± 2.01E-06 | **7.33E-05 \*\*\*** | -4.46E-05 ± 2.31E-05 | 0.053 | -0.021 ± 0.006 | **9.69E-04 \*\*\*** | -23 ± 5.81 | **7.34E-05 \*\*\*** |
| MDE | EOPD | 31 (20) | 3.46E-05 ± 2.61E-05 | 0.184 | -2.73E-04 ± 7.44E-04 | 0.713 | 0.096 ± 0.074 | 0.191 | 99.8 ± 75.2 | 0.184 |
|  | LOPD | 128 (20) | 3.33E-05 ± 1.69E-05 | **0.049 \*** | 2.86E-04 ± 3.03E-04 | 0.346 | 0.096 ± 0.047 | **0.040 \*** | 96.1 ± 48.8 | **0.049 \*** |
| SAS | EOPD | 22 (198) | -2.14E-05 ± 1.25E-05 | 0.087 | -1.84E-04 ± 3.06E-04 | 0.546 | -0.048 ± 0.03 | 0.106 | -61.7 ± 36 | 0.087 |
|  | LOPD | 40 (198) | -1.44E-05 ± 7.55E-06 | 0.057 | -4.44E-05 ± 2.5E-04 | 0.859 | -0.041 ± 0.023 | 0.067 | -41.5 ± 21.8 | 0.057 |

Legend: AAC = African Admixed, AFR = African, AJ = Ashkenazi Jewish, AMR = American Admixed, AV<sub>ROH</sub> = average length of runs of homozygosity, CAS = Central Asian, EAS = East Asian, EOPD = early-onset Parkinson's disease (< 45 years), EUR = European, F<sub>ROH</sub> = runs of homozygosity based estimates for inbreeding coefficient, LOPD = late-onset Parkinson's disease (≥ 45 years), MDE = Middle-Eastern, N<sub>ROH</sub> = number of runs of homozygosity, SAS = South Asian, SD = standard deviation, SROH = total length of runs of homozygosity. p-values indicate the statistical significance as following p < 0.001: \*\*\*, p < 0.01: \*\*, p < 0.05: \*, p ≥ 0.05: not significant

We analyzed the burden of ROH for EOPD *versus* LOPD cases by investigating the four parameters previously defined (**Table 3**, **Supplementary Table 14**). Here, significant enrichment was present in the AAC group across all four ROH parameters: S_ROH_, AV_ROH_, N_ROH_, and F_ROH_. No significant differences were observed in the other ancestral groups. The analysis was repeated after excluding known recessive PD genes (*PRKN*, *DJ-1*, and *PINK1*) from the dataset (**Supplementary Table 15**). Despite this adjustment, the statistically significant results remained unchanged, indicating that additional yet to be unraveled, recessive genes contribute to PD heritability in diverse ancestral populations.

**Table 3:** Regression results for runs of homozygosity in cases early-onset Parkinson’s disease versus late-onset Parkinson’s disease.

| Ancestry | Number of EOPD (LOPD) | S<sub>ROH</sub> |  | AV<sub>ROH</sub> |  | N<sub>ROH</sub> |  | F<sub>ROH</sub> |  |
| --- | --- | --- | --- | --- | --- | --- | --- | --- | --- |
|  |  | Beta (mean ± SE) | p-value | Beta (mean ± SE) | p-value | Beta (mean ± SE) | p-value | Beta (mean ± SE) | p-value |
| AAC | 21 (128) | -3.88E-05 ± 1.9E-05 | **0.041 \*** | -8.61E-04 ± 3.6E-04 | **0.017 \*** | -0.296 ± 0.138 | **0.032 \*** | -112 ± 54.7 | **0.041 \*** |
| AFR | 21 (137) | -4.31E-04 ± 2.42E-04 | 0.075 | 0.002 ± 0.001 | 0.105 | -0.557 ± 0.364 | 0.127 | -1240 ± 697 | 0.075 |
| AJ | 83 (719) | 6.05E-07 ± 4.97E-06 | 0.903 | 1.06E-05 ± 1.85E-04 | 0.954 | -0.003 ± 0.028 | 0.911 | 1.74 ± 14.4 | 0.903 |
| AMR | 91 (163) | -9.39E-06 ± 5.53E-06 | 0.090 | -2.24E-05 ± 1.42E-04 | 0.875 | -0.051 ± 0.028 | 0.073 | -27.1 ± 16 | 0.090 |
| CAS | 53 (57) | -2.25E-05 ± 2.54E-05 | 0.376 | 5.28E-04 ± 4.47E-04 | 0.237 | -0.075 ± 0.079 | 0.346 | -64.9 ± 73.2 | 0.376 |
| EAS | 243 (455) | -2.52E-05 ± 1.75E-05 | 0.151 | -1.57E-04 ± 1.39E-04 | 0.259 | -0.067 ± 0.044 | 0.127 | -72.6 ± 50.5 | 0.151 |
| EUR | 1,042 (6,400) | -8.37E-06 ± 4.36E-06 | 0.055 | -8.56E-06 ± 5.92E-05 | 0.885 | -0.016 ± 0.015 | 0.260 | -24.1 ± 12.6 | 0.055 |
| MDE | 31 (128) | 1.48E-06 ± 6.7E-06 | 0.825 | 3.2E-04 ± 5.1E-04 | 0.530 | 0.006 ± 0.021 | 0.764 | 4.27 ± 19.3 | 0.825 |
| SAS | 22 (40) | -7.97E-06 ± 4.15E-05 | 0.848 | -2E-04 ± 9.99E-04 | 0.841 | -0.052 ± 0.105 | 0.621 | -23 ± 120 | 0.848 |

Legend: AAC = African Admixed, AFR = African, AJ = Ashkenazi Jewish, AMR = American Admixed, AV<sub>ROH</sub> = average length of runs of homozygosity, CAS = Central Asian, EAS = East Asian, EOPD = early-onset Parkinson's disease (< 45 years), EUR = European, F<sub>ROH</sub> = runs of homozygosity based estimates for inbreeding coefficient, LOPD = late-onset Parkinson's disease (≥ 45 years), MDE = Middle-Eastern, N<sub>ROH</sub> = number of runs of homozygosity, SAS = South Asian, SD = standard deviation, S<sub>ROH</sub> = total length of runs of homozygosity. p-values indicate the statistical significance as following p < 0.001: \*\*\*, p < 0.01: \*\*, p < 0.05: \*, p ≥ 0.05: not significant

### Runs of homozygosity intersecting with known Parkinson’s disease, pallido-pyramidal syndrome, and atypical parkinsonism genes and risk loci suggest potential pleomorphic and pleiotropic effects in disease etiology across diverse ancestries

To explore the possibility of previously reported pleomorphic risk loci harboring recessive variants, we investigated ROHs intersecting with known PD, pallido-pyramidal syndrome, and atypical parkinsonism genes and risk loci. Specifically, we assessed the number of ROH segments overlapping these 297 gene regions (*N*=2,637) and the number of ROHs enriched in cases (*N*=638). Bonferroni correction was applied separately for each ancestry group, using a significance threshold of 0.05 divided by the number of ROH segments overlapping PD loci within each group (**Table 4**). Although ROHs were not found to be enriched in cases after Bonferroni correction due to the expected low frequency of potentially hidden recessive variants, we identified promising ROHs spanning known PD genes/loci (**Supplementary Figure 10**).

**Table 4:** Runs of homozygosity intersecting with known Parkinson’s disease, dystonia, and atypical parkinsonism known gene regions and risk loci.

| Ancestry | Number of ROH segments overlapping PD loci | Number of ROH segments enriched in cases | Number of ROH enriched segments with $p < 0.05$: | Number of ROH that Pass Bonferroni | Bonferroni Correction |
| --- | --- | --- | --- | --- | --- |
| AAC | 69 | 43 | 0 | 0 | 7.25E-04 |
| AFR | 256 | 164 | 7 | 0 | 1.95E-04 |
| AJ | 436 | 25 | 0 | 0 | 1.15E-04 |
| AMR | 107 | 56 | 0 | 0 | 4.67E-04 |
| CAS | 128 | 61 | 2 | 0 | 3.91E-04 |
| EAS | 277 | 78 | 0 | 0 | 1.81E-04 |
| EUR | 1,102 | 124 | 0 | 0 | 4.54E-05 |
| MDE | 118 | 63 | 0 | 0 | 4.24E-04 |
| SAS | 144 | 24 | 0 | 0 | 3.47E-04 |
| Legend: AAC = African Admixed, AFR = African, AJ = Ashkenazi Jewish, AMR = American Admixed, CAS = Central Asian, EAS = East Asian, EUR = European, MDE = Middle-Eastern, PD = Parkinson's disease, ROH = runs of homozygosity, SAS = South Asian. The Bonferroni threshold was adjusted by dividing 0.05 by the number of enriched ROHs to account for multiple comparisons. |  |  |  |  |  |

### Homozygosity length interval analysis reveals distinct patterns based on age at onset and ancestry

The AV_ROH_ was assessed from 2Mb to 10Mb in 1Mb increments (**Supplementary Table 16**). A statistically significant difference between cases and controls was observed for certain ROH lengths. Specifically, within the AAC ancestral group, nominal significance was found when assessing risk for ROHs at >2Mb, >3Mb, >4Mb, and >5Mb intervals. Within the EUR ancestral group, multiple lengths of ROHs exhibited significance. Similarly, within the SAS ancestral group, significance was observed for ROHs >3Mb to >10Mb. No other groups showed a statistically significant difference between cases and controls.

The analysis was repeated after cases were subsetted for EOPD and LOPD where a regression analysis was run for each age group *versus* controls (**Supplementary Table 17**). This analysis comparing EOPD and controls revealed significant differences in the frequency of ROH at specific lengths. Notably, within the AAC ancestral group, ROH >2Mb exhibited significance, along with ROH >3Mb and >4Mb. Notable differences in ROH frequency were observed between LOPD and controls across ancestral groups. In the AAC group, ROH >3Mb and ROH >4Mb were significant. In the AFR group, ROH >7Mb showed a notable difference, while in the EAS group, ROH >2Mb also differed. Within the EUR ancestral group, multiple lengths of ROH were significant, including those >2Mb to >10Mb. Finally, in the SAS ancestral group, significance was observed for ROH >4Mb to >7Mb. There was no significant enrichment for the AJ, AMR, and CAS groups.

### Homozygosity mapping identifies runs of homozygosity overlaps segregating within families

We examined regions of ROHs that segregate with PD within families (**Table 5**). Our analysis identified 11 ROHs segregating within MDE families present in cases following a potential recessive model of inheritance and absent in controls. WGS data was used to further investigate 151,995 variants identified in the ROHs segregating within MDE families. After filtering for variants based on allele frequency and variant consequence, eight homozygous variants remained (**Table 6**) and were present in more than one individual. Seven variants (rs773266496, rs41285470, rs369145807, rs144512254, rs145062084, rs2338627, and rs2338626) were reported as likely benign. One variant (rs45539432) was identified as a stop-gain variant in PTEN-induced kinase 1 (*PINK1*), with a clinical classification of pathogenic and likely causal for EOPD. After assessing overall ROHs, we did not detect ROHs that segregate within families and were present in cases but absent in controls for any other ancestral group.

**Table 5:** Homozygosity mapping results for runs of homozygosity segregating in families.

| Ancestry | Cases (controls) related individuals | Cases (controls) unrelated individuals | Related individuals (Percentage) | Number of families (Average members) | Age at onset (mean ± SD) | Has family history (Percentage) | ROHs in cases only segregating in families |
| --- | --- | --- | --- | --- | --- | --- | --- |
| AAC | 257 (808) | 256 (804) | 11 (1) | 5 (2) | 53 ± NA | 18.2 | 0 |
| AFR | 920 (1,683) | 917 (1,631) | 87 (3.3) | 30 (3) | 40 ± 8.7 | 2.3 | 0 |
| AJ | 1,011 (459) | 1,001 (458) | 19 (1.3) | 9 (2) | 61.7 ± 12 | 68.4 | 0 |
| AMR | 367 (139) | 363 (139) | 7 (1.4) | 3 (2) | 48.3 ± 16 | 85.7 | 0 |
| CAS | 286 (294) | 265 (267) | 79 (13.6) | 26 (3) | 33 ± 13 | 38 | 0 |
| EAS | 1,577 (2,378) | 1,537 (2,302) | 215 (5.4) | 102 (2) | 57.6 ± 18 | 27 | 0 |
| EUR | 11,841<br>(7,596) | 11,679<br>(7,505) | 411 (2.1) | 190 (2) | 55.4 ± 15 | 57.2 | 0 |
| MDE | 218 (21) | 214 (20) | 12 (5) | 5 (2) | 41.2 ± 13 | 66.7 | 11 |
| SAS | 122 (207) | 117 (198) | 32 (9.7) | 13 (2) | 52.8 ± 9 | 15.6 | 0 |

Legend: AAC = African Admixed, AFR = African, AJ = Ashkenazi Jewish, AMR = American Admixed, CAS = Central Asian, EAS = East Asian, EUR = European, MDE = Middle-Eastern, PD = Parkinson's disease, ROHs = runs of homozygosity, SAS = South Asian. The Bonferroni threshold was adjusted by dividing 0.05 by the number of enriched ROHs to account for multiple comparisons.

**Table 6:**
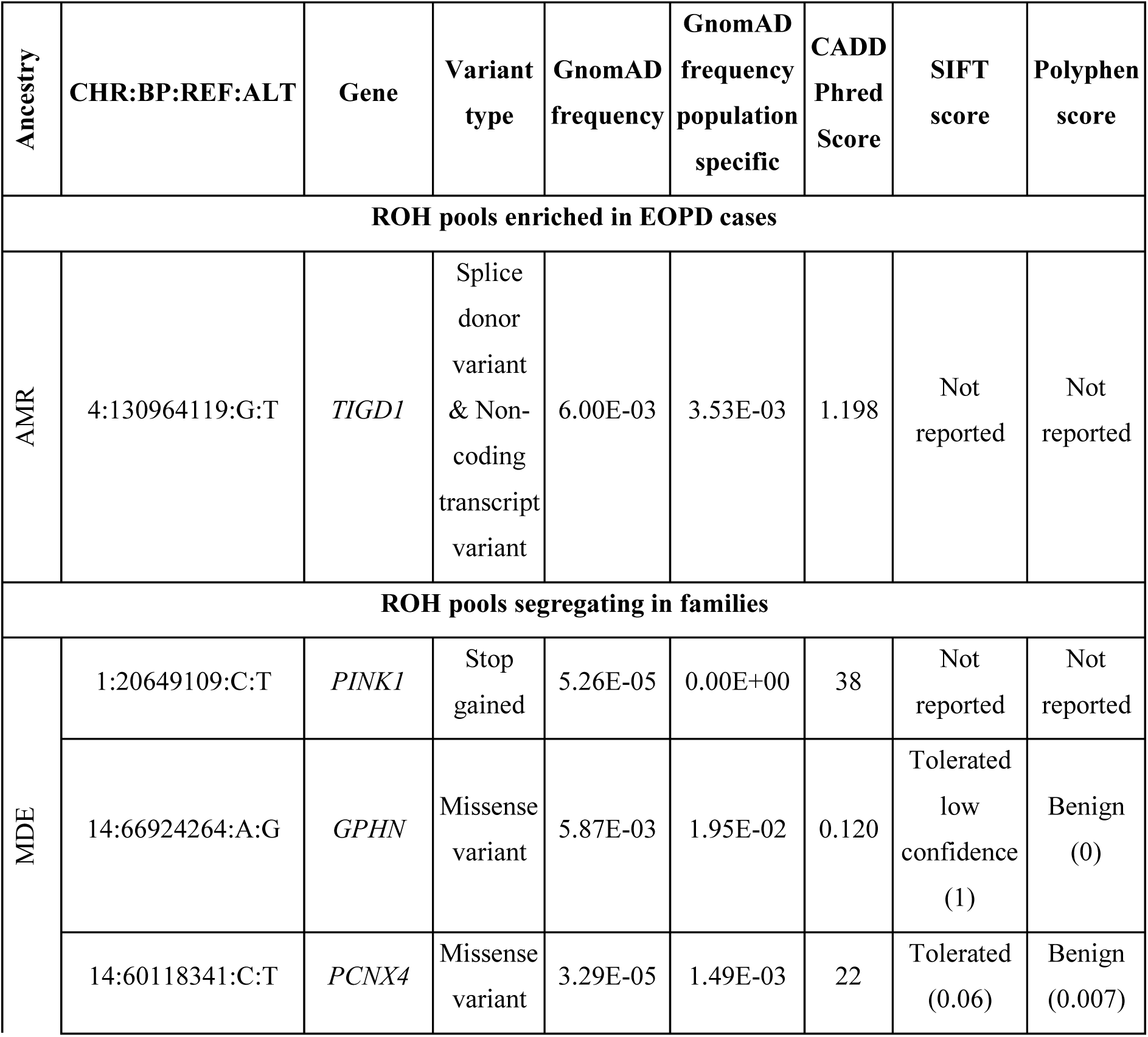
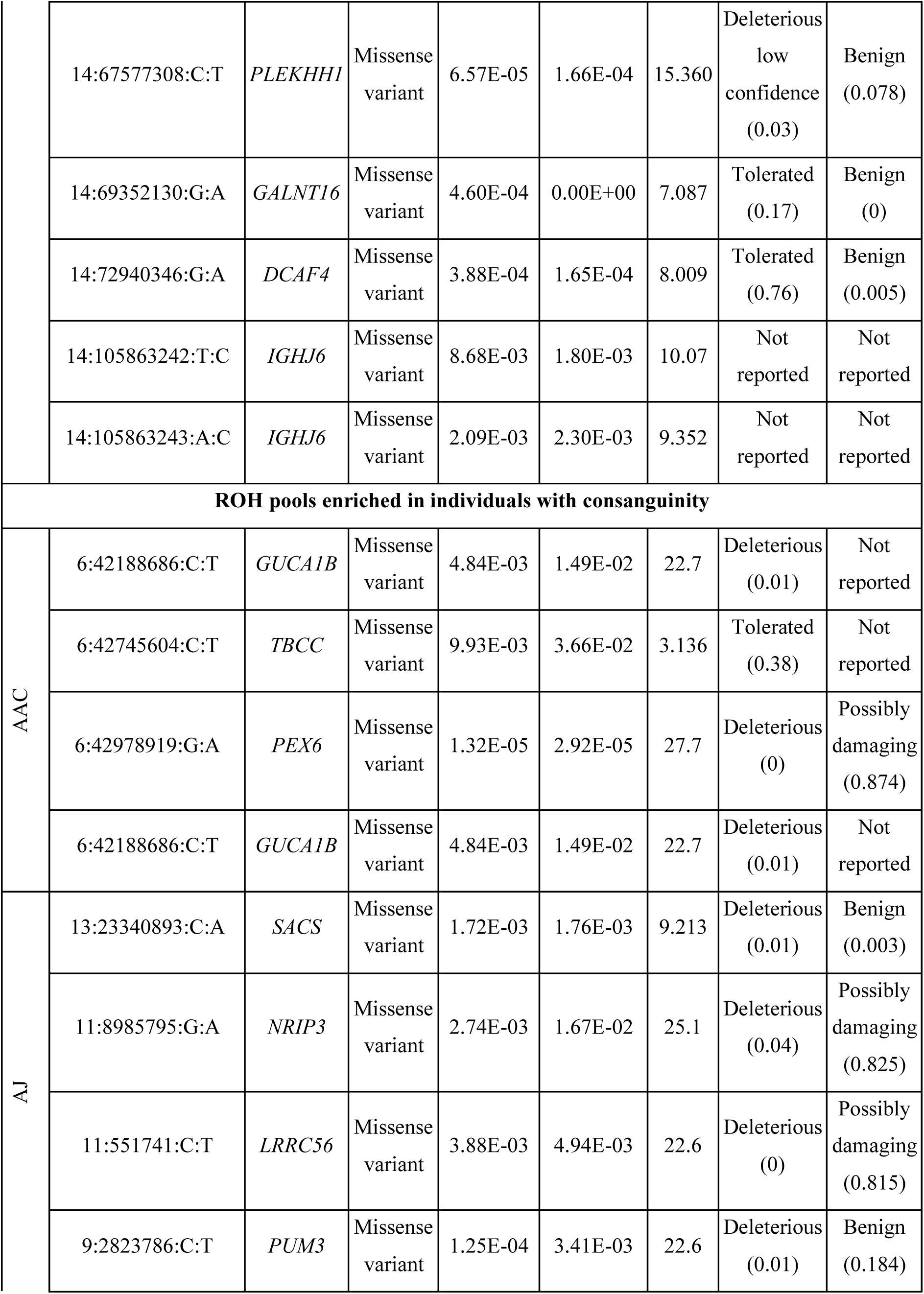
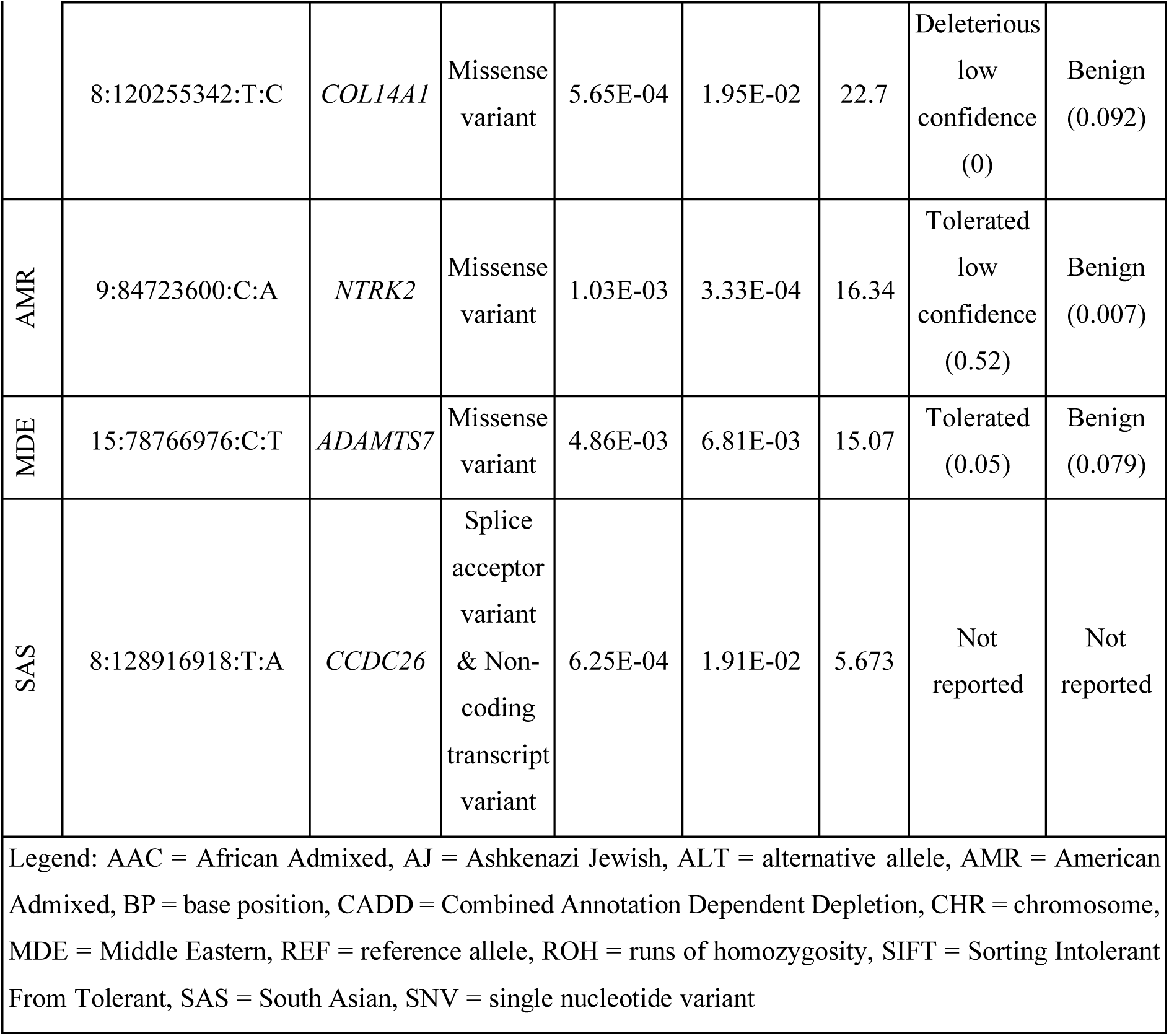
Annotated variants only present in Parkinson’s disease cases prioritized from homozygosity mapping.

### Homozygosity mapping identifies runs of homozygosity enriched in individuals with consanguinity

Among the total sample (*N*=29,673), we identified 235 PD cases presenting with a F_ROH_ >0.0156, corresponding to a second-degree relation or closer between parents as compared to all controls (*N*=13,324) (**Supplementary Table 18**, **Supplementary Figure 11**). For the 235 PD cases, 72 had a positive family history, 105 had no family history, and 53 had no family history reported. After investigating ROHs present in these cases and absent in controls, we found the following counts across ancestries: AAC=3, AFR=1, AJ=10, AMR=86, CAS=17, EAS=2, EUR=1, MDE=298, and SAS=2. WGS data (*N*=54) was used to further investigate the ROHs exclusive to the individuals with consanguinity. We retained a total of 12 variants with the maximum credible genetic ancestry group allele frequency ≤0.01, each in a different case (AAC=4, AJ=5, AMR=1, MDE=1, and SAS=1; **Table 6**, **Supplementary Table 19**).

Our analysis also identified 7,816 ROH overlaps across the nine ancestral groups, of which 5,274 were enriched in the cases and 1,283 passed Bonferroni correction (**Supplementary Table 20**). Moreover, we investigated ROHs that overlap known recessive PD, pallido-pyramidal syndrome, and atypical parkinsonism genes and risk loci (*N*=811), as well as those overlapping PD genes that were enriched in individuals with consanguinity (*N*=565), and those passing Bonferroni correction (*N*=167), as seen in **Supplementary Table 21**. Notably, the MDE group did not have any significant ROH overlapping known recessive PD, pallido-pyramidal syndrome, and atypical parkinsonism genes and risk loci, suggesting novel genetic causes might contribute to PD susceptibility in this group, or the cases sharing the same genetic cause is low, e.g., variants in PINK1.

### Homozygosity mapping identifies runs of homozygosity enriched in early-onset Parkinson’s disease cases

Homozygosity mapping was used to further investigate ROHs enriched in EOPD. We identified 1,607 EOPD cases across the nine ancestral groups (**Supplementary Table 18**, **Supplementary Figure 12**). Our analysis revealed ROH pools present exclusively in EOPD cases, specifically in the AJ (*N*=3), AMR (*N*=60), EUR (*N*=3), and MDE (*N*=175) groups. WGS data was used to further investigate the ROHs exclusive to the EOPD individuals. The variant rs78589926, located in tigger transposable element derived 1 (*TIGD1*) pseudogene, was found to be carried in a homozygous state in four individuals and remained in the dataset after filtering out variants with the maximum credible genetic ancestry group allele frequency >0.01 in gnomAD v4.1.0 genomes (**Table 6**). Additionally, we investigated ROH pools enriched in EOPD cases (*N*=2,466), with 17 passing Bonferroni correction (**Supplementary Table 22**). Finally, we examined ROHs overlapping PD genes (*N*=661) and those enriched in EOPD cases (*N*=259), but no ROHs passed Bonferroni correction as expected due to the low frequency of the ROH under study.

## Discussion

This study is the most extensive screening of homozygosity in PD across diverse populations. We successfully investigated the burden of ROHs across four parameters in nine ancestral groups. We screened ROHs intersecting with known recessive PD genes and risk loci, in addition to pallido-pyramidal syndrome, and atypical parkinsonism genes. We further nominated and prioritized novel consensus ROHs in families, individuals with consanguinity, and EOPD cases; and further validated our findings using WGS data.

Complex phenotypes, such as PD, can be linked to long ROHs that have been recessively inherited and are detectable using genotyping arrays and sequencing approaches (Clark *et al*., 2019). In the neurodegenerative space, previous studies have reported significant overrepresentation of ROHs associated with increased AD risk in European populations (Moreno-Grau *et al*., 2021). Additionally, in an AD African American cohort, a range of ROHs regions greater than 2 Mb in length were significantly associated with disease risk (Ghani *et al*., 2015). In the PD field, a study conducted over a decade ago examined ROHs in a European PD cohort and identified increased levels of homozygosity in EOPD cases in comparison to controls (Simón-Sánchez *et al*., 2012), failing to explore the role of homozygosity across diverse ancestry populations.

In this study, our multi-ancestry genome-wide assessment revealed increased homozygosity in PD. Larger values for S_ROH_, AV_ROH_, and N_ROH_ were seen in more populations with consanguinity, such as the AJ (F_ROH_=0.008), MDE (F_ROH_=0.009), and CAS (F_ROH_=0.005) groups. Individuals from these populations are more likely to share recent common ancestors (Bittles, 2008). As a result, regions of the genome tend to be homozygous over longer stretches compared to outbred and more admixed populations, such as the AAC (F_ROH_=0.002), AFR (F_ROH_=0.003), and EAS (F_ROH_=0.002) groups. In an attempt to define recessive modifiers of PD onset, a significant overrepresentation of ROH burden was observed in EOPD and LOPD. These findings highlight the relevance of ROH parameters in understanding the genetic architecture of PD. Furthermore, the results showing statistical significance remained the same after removing known recessive PD genes (*PRKN*, *DJ-1*, and *PINK1*) on (1) cases *versus* controls, (2) EOPD *versus* controls, (3) LOPD *versus* controls, and (4) EOPD cases *versus* LOPD cases. Ultimately, increased genomic homozygosity, excluding known recessive genes, suggests unknown genetic factors contribute to PD heritability.

We took a step forward by characterizing increased homozygosity in a granular manner and potentially nominated regions harboring novel and rare recessive variants for further study. ROH intersecting with known recessive PD, pallido-pyramidal syndrome, and atypical parkinsonism genes and risk loci suggest putative pleiotropic effects in disease etiology or the presence of misdiagnosed cases across diverse ancestries that warrant further investigation. The limited availability of WGS data constrained the analysis of overlapping ROHs within known gene regions. However, in the AAC group, individuals shared ROHs in *PLA2G6* and *PINK1*, while overlapping ROHs were observed in both the AAC and AFR groups in *GCH1, MAP4KA,* and *ELOVL7*. Additionally, overlapping ROHs in *PRKN* were identified in both the AJ and MDE groups. Notably, two carriers from the SAS group had ROHs in *CHCHD2*, and overlapping ROHs in *SV2C* were found in both the SAS and EAS groups. Interestingly, *SV2C* had been previously flagged in a GWAS study on an Asian cohort (Foo *et al*., 2020). The few *PRKN* and *PINK1* carriers likely reflect the pre-selection of WGS samples negative for known genetic causes.

We observed a similar trend as previously reported for EOPD (Simón-Sánchez *et al*., 2012), where the AV_ROH_ decreased as the ROH interval increments increased. The homozygosity length interval analysis reveals distinct genetic architecture patterns based on AAO and ancestry. This analysis highlighted that populations with higher levels of admixture, typically viewed as ‘older’ populations (Ragsdale *et al*., 2023), tend to exhibit shorter ROH segments (Ceballos *et al*., 2018). These shorter segments have likely been present for longer periods, suggesting ancient admixture. In contrast, longer ROHs reflect a more recent relatedness, possibly from founder effects or consanguinity (Kirin *et al*., 2010). This distinction serves as a proof of concept for investigating family-specific ROHs *versus* ROHs as a common population haplotype. Moreover, the AV_ROH_ was consistently greater in EOPD cases compared to LOPD cases for most ancestral groups. However, in the AJ, EUR, and SAS populations, LOPD cases exhibited a slightly higher AV_ROH_ than EOPD cases. These differences between ancestries further underscore the importance of including diverse ancestral groups in PD genetic research to fully understand the genetic architecture of risk and onset.

The present study identified homozygosity overlaps segregated within families, regions enriched in individuals with consanguinity, and regions enriched in EOPD cases. Among the 21 variants prioritized using WGS, one variant (rs45539432) was identified as being the genetic cause underlying PD in one family, with an AAO of 38, 44, and 41 across the affected members, as well as in one unrelated individual with an AAO of 47 in the MDE group. This variant was previously shown to co-segregate in all affected members in a Sudanese family (Bakhit *et al*., 2023). Functional studies revealed that the encoded protein was poorly expressed, unstable, and minimally stabilized upon mitochondrial depolarization, resulting in its failure to activate parkin and initiate substrate ubiquitination (Siuda *et al*., 2014). The remaining 20 prioritized variants were missense or splice-site variants classified as either likely benign or not reported. While the EUR group had the most EOPD cases, the MDE group had the most ROHs present in cases and absent in controls.

Notably, the MDE group also had the highest ROH presence in individuals with consanguinity but was the only group with no ROHs in known PD regions that passed Bonferroni correction. This suggests that novel genetic causes might contribute to PD susceptibility in this group.

Conversely, all groups showed ROHs in EOPD cases that overlapped with known PD gene regions. This indicates that while some ancestral groups like MDE may harbor unique genetic factors not yet associated with known PD regions, other groups show a direct overlap with established PD loci. This would support our hypothesis regarding novel pleomorphic effects. The presence of ROHs in known PD regions across different groups highlights the complex genetic architecture of PD, involving both common and potentially novel genetic contributions to disease susceptibility. Although homozygosity mapping did not identify any new gene regions associated with PD, the findings demonstrate the potential of this analysis approach for exploring the genetic etiology of the disease. Here, we have developed an open-science framework to conduct homozygosity mapping in an unbiased and large-scale manner. Future research should focus on larger sample sizes across diverse ancestries and include comprehensive WGS data to further identify rare variants contributing to disease susceptibility.

Despite successfully performing a genome-wide assessment of homozygosity across nine ancestral groups, our study has several limitations. Firstly, the predominant representation of EUR populations among the participants may introduce a bias in our findings. Since the analysis was conducted separately for each ancestral group, the overrepresentation of EUR populations could potentially skew the overall interpretation and generalizability of the results across diverse genetic backgrounds. Additionally, certain ancestral groups, such as the MDE group, had limited number of controls, resulting in an unequal ratio of cases to controls and potentially affecting the power to detect ROH associations in these groups. We were also underpowered to detect rare variations in some populations due to sample size constraints, limiting our ability to capture the full spectrum of genetic diversity and potentially underrepresenting rare variants significant in non-European populations. Due to the absence of some common data elements, we were required to impute the missing ages to incorporate them as a covariate in the analysis. This imputation process introduces potential biases and uncertainties, which may affect the reliability and accuracy of our results. Furthermore, WGS data was yet to be available for the majority of ROH pools we had prioritized through genotyping, restricting our ability to further explore the nominated regions. Future data releases are expected to significantly increase the number of WGS, particularly for populations previously underrepresented in PD genetics research. Finally, we acknowledge the possibility of overestimating the presence of potential ROHs as a result of heterozygous deletions, effectively mimicking the behavior of homozygosity. The analysis of both homozygous and heterozygous structural variants is not included in the scope of this project.

Our findings highlight the potential contribution of homozygosity to the genetic etiology of PD, providing compelling evidence that an additional portion of PD heritability may be attributed to a recessive pattern of inheritance outside the known recessive PD genes. Our comprehensive approach nominated several novel ROHs enriched in PD across diverse ancestries, paving the way for further discoveries contributing to our understanding of PD heritability on a global scale.

## Data and code availability

Data used in the preparation of this article were obtained from Global Parkinson’s Genetics Program (GP2). GP2 is funded by the Aligning Science Across Parkinson’s (ASAP) initiative and implemented by The Michael J. Fox Foundation for Parkinson’s Research (https://gp2.org). All GP2 data for these analyses is available through collaboration with Accelerating Medicines Partnership in Parkinson’s disease and is available through application on the AMP-PD website (https://amp-pd.org/register-for-amp-pd). For a complete list of GP2 members see https://gp2.org. GenoTools version 10 (https://github.com/GP2code/GenoTools) was used for quality control, imputation, and ancestry prediction. A secure workspace on the online Terra platform (https://app.terra.bio/) was created to analyze the data using GP2’s release 6. Finally, all scripts used for the data analysis can be found in the public domain on GitHub (https://github.com/GP2code/ROH_genomewide/, (Makarious, 2025)).

## Author contributions

S.B-C. conceptualized the study and Z.H., K.S., C.F.H., A.J.H-M., P-J.K., M.O., E.E., and A.Z. designed the study. The data analysis was performed by C.F.H. (AAC, AMR, & EUR), K.S. (AFR & CAS), A.J.H-M. (AJ), M.O. (CAS), P-J.K. (EAS), E.E. (MDE), and A.Z. (SAS). Z-H.F and C.F.H. wrote the homozygosity mapping script. K.S. wrote the first draft of the manuscript. All the authors reviewed, edited, and approved the manuscript.

## Acknowledgments

This work was supported and guided by the ‘GP2 Trainee Network’ and formed part of the Global Parkinson’s Genetics Program (GP2) Hackathon 2023. The data used for the analysis was obtained from the GP2 (https://gp2.org) which is funded by the Aligning Science Across Parkinson’s (ASAP) initiative and implemented by The Michael J. Fox Foundation for Parkinson’s Research (https://michaeljfox.org/). A complete list of GP2 members is available at https://gp2.org. All figures were created using BioRender (https://www.biorender.com/). We thank Caroline Pantazis for her work as scientific project manager for this project. We thank Paige Brown Jarreau for her meticulous editing of this manuscript. Research reported in this publication was partly supported by the South African Medical Research Council. *For open access, the author has applied a CC BY public copyright license to all Author Accepted Manuscripts arising from this submission*.

