## Supplementary Material for "Genome-wide assessment identifies novel runs of homozygosity linked to Parkinson’s disease etiology across diverse ancestral populations"

**Supplementary Figure 1:** Scree plots illustrating eigenvalues for principal component analysis (PCA). The eigenvalues of each principal component indicate the proportion of variance explained by each component. The point at which the eigenvalues level off suggests the optimal number of components to retain as covariates in the analysis. AAC = African Admixed, AFR = African, AJ = Ashkenazi Jewish, AMR = American Admixed, CAS = Central Asian, EAS = East Asian, EUR = European, MDE = Middle-Eastern, SAS = South Asian ........ 1

**Supplementary Figure 2**: Box plots illustrating the distribution of four ROH parameters assessed colored by phenotype for each ancestral group. The x-axis represents the ancestry and the y-axis represents the ROH parameter. AAC = African Admixed, AFR = African, AJ = Ashkenazi Jewish, AMR = American Admixed, ANC = ancestry, $AV_{ROH}$ = average length of runs of homozygosity, CAS = Central Asian, EAS = East Asian, EUR = European, $F_{ROH}$ = runs of homozygosity-based estimates for inbreeding coefficient, MDE = Middle-Eastern, $N_{ROH}$ = number of runs of homozygosity, SAS = South Asian, $S_{ROH}$ = total length of runs of homozygosity. ........ 2

........ 2

**Supplementary Figure 3:** (A) Mean $N_{ROH}$ versus $S_{ROH}$ in Mb for each of the nine ancestral groups. (B) Mean $N_{ROH}$ versus the $S_{ROH}$ in Mb for each individual included in the analysis. AAC = African Admixed, AFR = African, AJ = Ashkenazi Jewish, AMR = American Admixed, CAS = Central Asian, EAS = East Asian, EUR = European, MDE = Middle-Eastern, ROH = runs of homozygosity, SAS = South Asian, $S_{ROH}$ = total length of runs of homozygosity. ........ 3

**Supplementary Figure 4:** Comparison of the runs of homozygosity versus length of runs of homozygosity for cases and controls across the ancestral groups. For phenotype, 1 indicates control and 2 indicates case. AAC = African Admixed, AFR = African, AJ = Ashkenazi Jewish, AMR = American Admixed, CAS = Central Asian, EAS = East Asian, EUR = European, KB = kilobase ($S_{ROH}$), MDE = Middle-Eastern, NSEG = number of runs of homozygosity ($N_{ROH}$), PHE = phenotype, SAS = South Asian ........ 4

**Supplementary Figure 5:** Comparison of the runs of homozygosity versus average length of runs of homozygosity for cases and controls across the ancestral groups. For phenotype, 1 indicates control and 2 indicates case. AAC = African Admixed, AFR = African, AJ = Ashkenazi Jewish, AMR = American Admixed, CAS = Central Asian, EAS = East Asian, EUR = European, KBAVG = average number of runs of homozygosity ($AV_{ROH}$), MDE = Middle-Eastern, NSEG = number of runs of homozygosity ($N_{ROH}$), SAS = South Asian ........ 5

**Supplementary Figure 6:** Density plot illustrating the distribution of inbreeding coefficients ($F_{ROH}$) across the ancestry groups for (A) cases and controls, (B) cases only, and (C) controls only. The x-axis represents the inbreeding coefficient values ranging from -0.5 to 0.25, while the y-axis represents the density of observations. AAC = African Admixed, AFR = African, AJ = Ashkenazi Jewish, AMR = American Admixed, CAS = Central Asian, EAS = East Asian, EUR = European, MDE = Middle-Eastern, SAS = South Asian. ..6

**Supplementary Figure 7:** Density plots illustrating the distribution of inbreeding coefficients ($F_{ROH}$) across the ancestral groups. The x-axis represents the $F_{ROH}$ values, while the y-axis represents the density of observations at each F value. AAC = African Admixed, AFR = African, AJ = Ashkenazi Jewish, AMR =

American Admixed, CAS = Central Asian, EAS = East Asian, EUR = European, MDE = Middle-Eastern, SAS = South Asian..........7

**Supplementary Figure 8:** Density plots illustrating the distribution of inbreeding coefficients ($F_{ROH}$) colored by phenotype for each ancestral group. For phenotype, 0 indicates control and 1 indicates case. Each curve represents the density of observations for a specific phenotype value. The x-axis represents the values of F, while the y-axis represents the density of observations at each F value. AAC = African Admixed, AFR = African, AJ = Ashkenazi Jewish, AMR = American Admixed, CAS = Central Asian, EAS = East Asian, EUR = European, MDE = Middle-Eastern, SAS = South Asian..........8

**Supplementary Figure 9:** Box plots illustrating the distribution of four ROH parameters assessed colored by phenotype including early and late onset PD for each ancestral group. The y-axis represents the ancestral group. AAC = African Admixed, AFR = African, AJ = Ashkenazi Jewish, AMR = American Admixed, ANC = ancestry, CAS = Central Asian, EAS = East Asian, EUR = European, $F_{ROH}$ = run of homozygosity based estimates for the inbreeding coefficient, KB = kilobases, KBAVG = average length of runs of homozygosity, MDE = Middle-Eastern, NSEG = number of runs of homozygosity, SAS = South Asian. ..........9

**Supplementary Figure 10**: Homozygosity mapping plots displaying runs of homozygosity enriched in cases across diverse ancestries. Highlighted in red are the runs of homozygosity present only in cases. The chromosome is along the x-axis and the runs of homozygosity size in base positions in along the y-axis. AAC = African Admixed, AFR = African, AJ = Ashkenazi Jewish, AMR = American Admixed, CAS = Central Asian, EAS = East Asian, EUR = European, MDE = Middle-Eastern, SAS = South Asian..........10

**Supplementary Figure 11:** Heatmap illustrating the number of individuals or runs of homozygosity obtained from the homozygosity mapping analysis for individuals with consanguinity. AAC = African Admixed, AFR = African, AJ = Ashkenazi Jewish, AMR = American Admixed, CAS = Central Asian, EAS = East Asian, EUR = European, MDE = Middle-Eastern, SAS = South Asian. The Bonferroni threshold was adjusted by dividing 0.05 by the number of enriched ROHs to account for multiple comparisons. ..........11

**Supplementary Figure 12:** Heatmap illustrating the number of individuals or runs of homozygosity obtained from the homozygosity mapping analysis for early-onset Parkinson's disease patients. AAC = African Admixed, AFR = African, AJ = Ashkenazi Jewish, AMR = American Admixed, CAS = Central Asian, EAS = East Asian, EUR = European, MDE = Middle-Eastern, SAS = South Asian. The Bonferroni threshold was adjusted by dividing 0.05 by the number of enriched ROHs to account for multiple comparisons..........12

### Supplementary Materials

*ROH calculations*

*Sum of ROH ($S_{ROH}$)*

The mean length of ROH is calculated by dividing the sum of ROH lengths by number of ROH segments, as follows:

$$Mean\ ROH\ =\ \frac{Sum\ of\ ROH\ lengths}{Number\ of\ ROH\ segments}$$

$$SROH\ =\ \Sigma_{i=1}^{n}\ Length\ of\ ROH_i$$

Where $n$ is the number of ROH segments and the length of ROH and $Length\ of\ ROH_i$ is the number of consecutive homozygous markers in a segment.

*Average length of ROH (AVROH)*

The $AV_{ROH}$ provides the average measure of length of homozygous regions in an individual's genome. This is calculated by dividing the sum of the lengths of all the individual ROH by the total number of ROH, as follows:

$$Average\ ROH\ =\ \frac{\Sigma_{i=1}^{n} Length\ of\ ROH_i}{n}$$

Where $n$ is the total number of ROH segments and $Length\ of\ ROH_i$ is the length of the $i^{th}$ ROH.

*Number of ROH ($N_{ROH}$)*

The $N_{ROH}$ is the count of the total number of occurrences of ROH segments in an individual's genome. Here each ROH segment is considered one ROH occurrence and the total number of these segments is the $N_{ROH}$ in the genome, as follows:

$$NROH\ =\ Total\ number\ of\ ROH\ segments$$

*ROH-based estimates of the inbreeding coefficient per individual ($F_{ROH}$)*

The inbreeding coefficient ($F_{ROH}$) indicates the degree of relatedness due to shared ancestry. It is the probability of an individual inheriting two alleles at a locus from a common ancestor without a recombination event. In a ROH context, $F_{ROH}$ estimates F based on the number of length of ROH segments in an individual's genome. It is the ratio of the sum of the lengths of ROH segments and the total length of the autosomal genome containing ROH segments. The $F_{ROH}$ provides the proportion of the genome that is homozygous due to recent common ancestry and is calculated as follows:

$$FROH\ =\ \frac{\Sigma_{i=1}^{n} Length\ of\ ROH_i}{Lgenome}$$

Where $n$ is the total number of ROH segments, $Length\ of\ ROH_i$ is the length of the $i^{th}$ ROH segment, and $L_{genome}$ is the total length of the autosomal genome containing ROH segments.

### Supplementary Figures

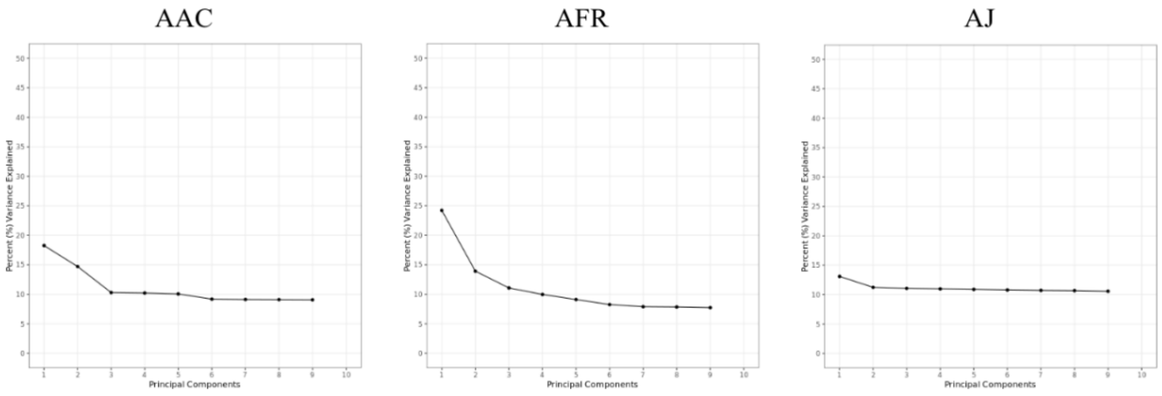

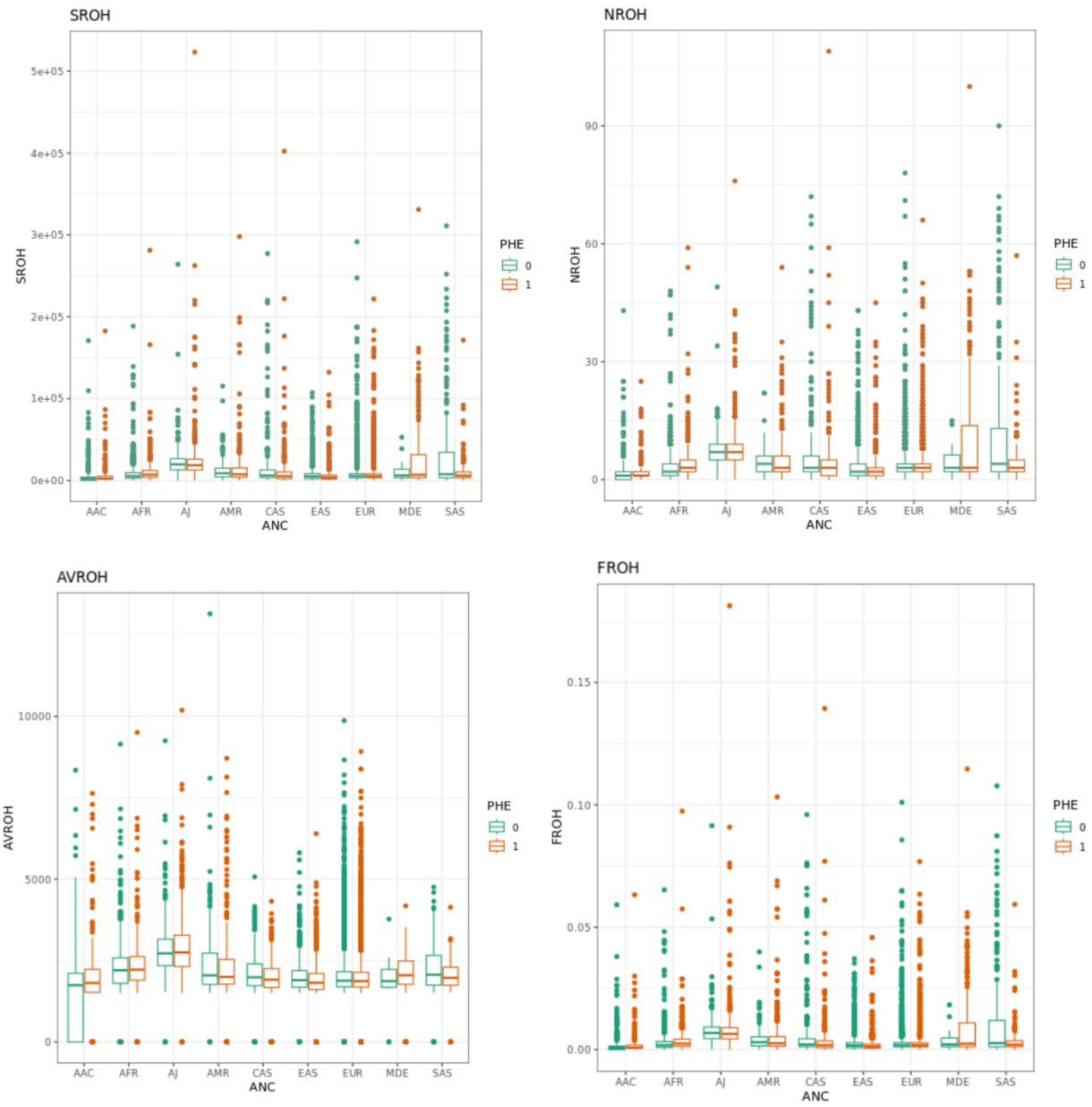

***Supplementary Figure 2****: Box plots illustrating the distribution of four ROH parameters assessed colored by phenotype for each ancestral group. The x-axis represents the ancestry and the y-axis represents the ROH parameter. AAC = African Admixed, AFR = African, AJ = Ashkenazi Jewish, AMR = American Admixed, ANC = ancestry, $AV_{ROH}$ = average length of runs of homozygosity, CAS = Central Asian, EAS = East Asian, EUR = European, $F_{ROH}$ = runs of homozygosity-based estimates for inbreeding coefficient, MDE = Middle-Eastern, $N_{ROH}$ = number of runs of homozygosity, SAS = South Asian, $S_{ROH}$ = total length of runs of homozygosity.*

(A) Per ancestral group (B) Per individual

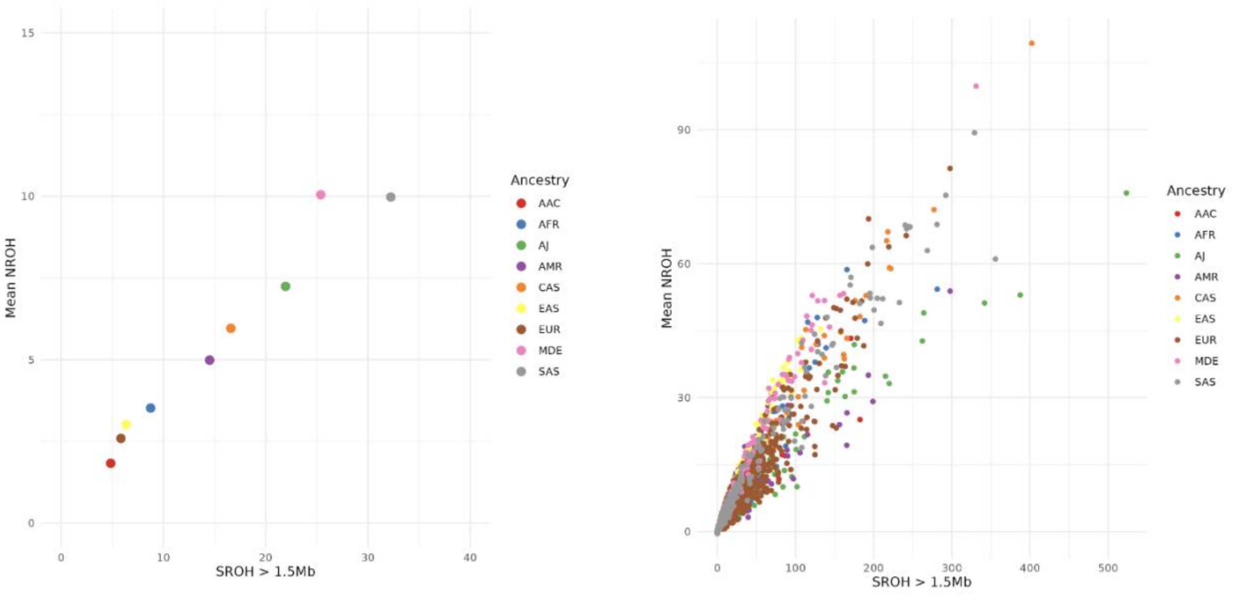

***Supplementary Figure 3:*** *(A) Mean* $N_{ROH}$ *versus* $S_{ROH}$ *in Mb for each of the nine ancestral groups. (B) Mean* $N_{ROH}$ *versus the* $S_{ROH}$ *in Mb for each individual included in the analysis. AAC = African Admixed, AFR = African, AJ = Ashkenazi Jewish, AMR = American Admixed, CAS = Central Asian, EAS = East Asian, EUR = European, MDE = Middle-Eastern, ROH = runs of homozygosity, SAS = South Asian,* $S_{ROH}$ *= total length of runs of homozygosity.*

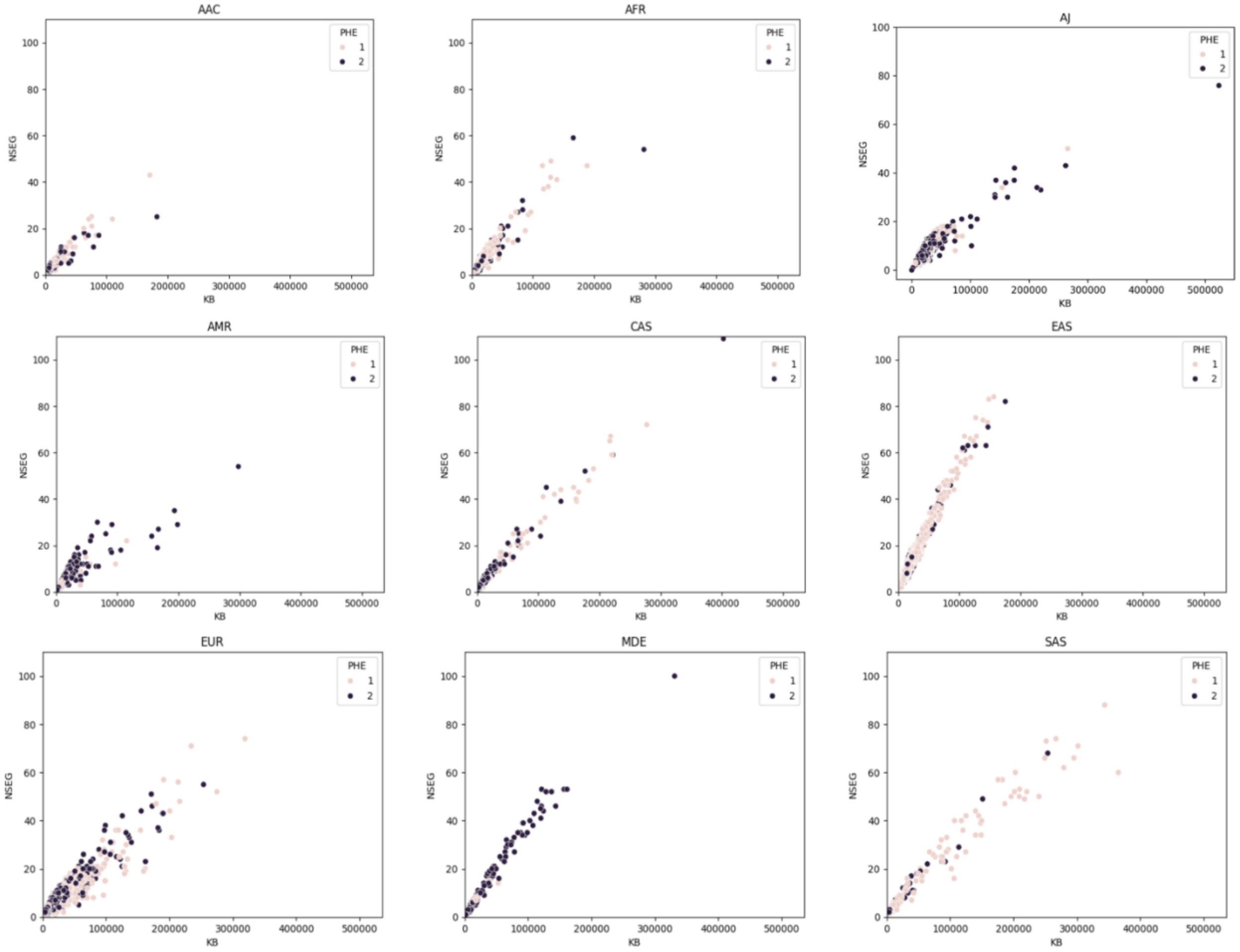

***Supplementary Figure 4:*** *Comparison of the runs of homozygosity versus length of runs of homozygosity for cases and controls across the ancestral groups. For phenotype, 1 indicates control and 2 indicates case. AAC = African Admixed, AFR = African, AJ = Ashkenazi Jewish, AMR = American Admixed, CAS = Central Asian, EAS = East Asian, EUR = European, KB = kilobase ($S_{ROH}$), MDE = Middle-Eastern, NSEG = number of runs of homozygosity ($N_{ROH}$), PHE = phenotype, SAS = South Asian*

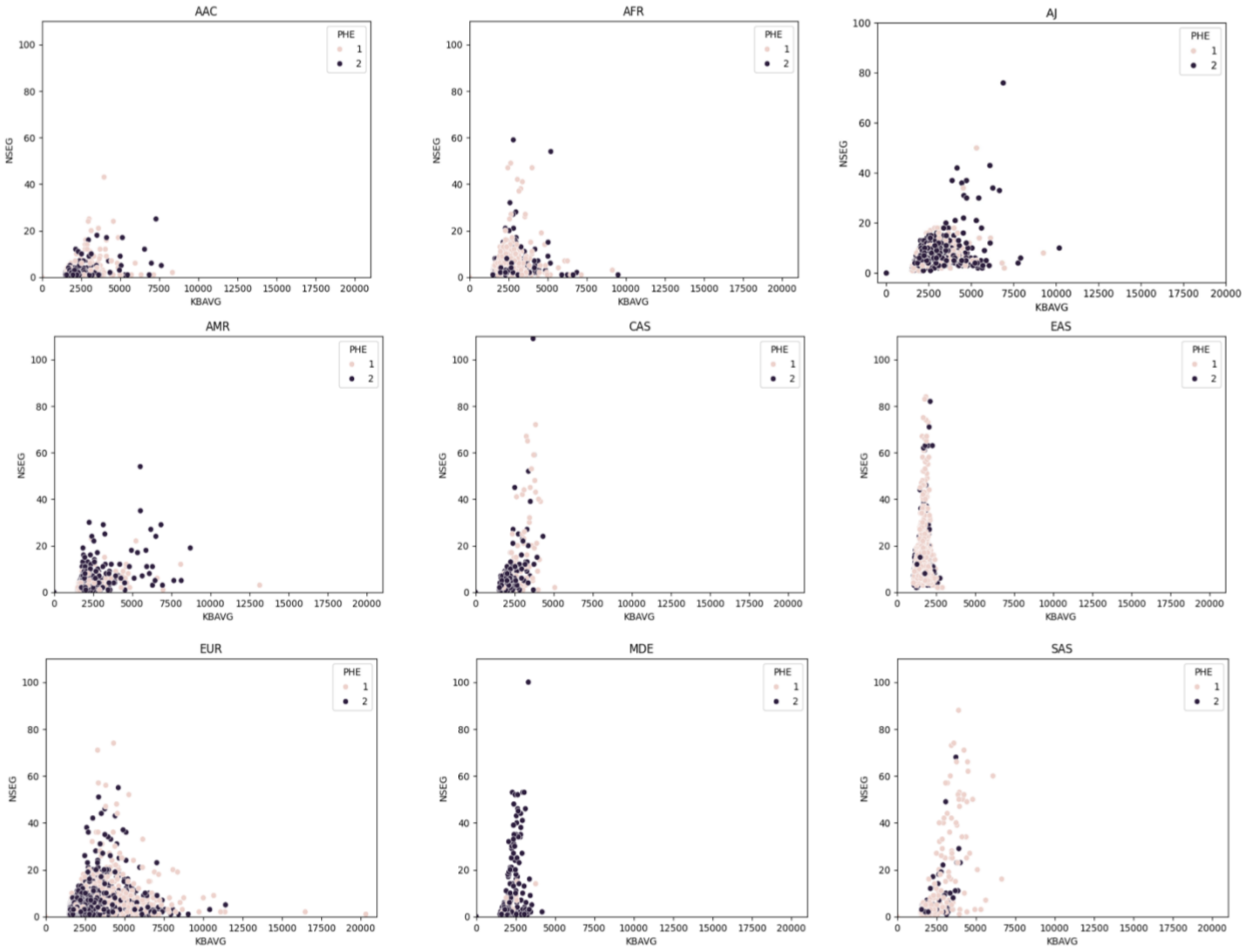

***Supplementary Figure 5:*** *Comparison of the runs of homozygosity versus average length of runs of homozygosity for cases and controls across the ancestral groups. For phenotype, 1 indicates control and 2 indicates case. AAC = African Admixed, AFR = African, AJ = Ashkenazi Jewish, AMR = American Admixed, CAS = Central Asian, EAS = East Asian, EUR = European, KBAVG = average number of runs of homozygosity ($AV_{ROH}$), MDE = Middle-Eastern, NSEG = number of runs of homozygosity ($N_{ROH}$), SAS = South Asian*

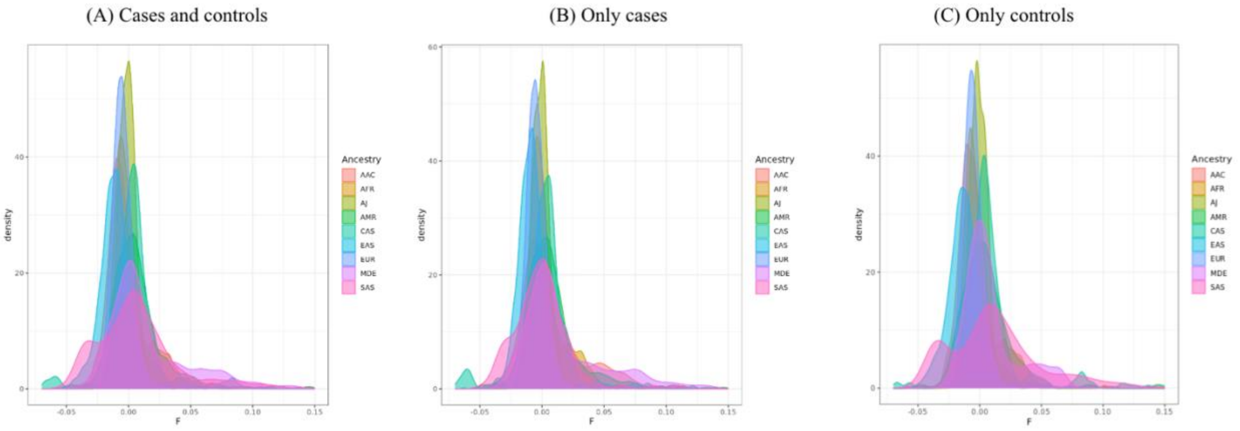

***Supplementary Figure 6:*** *Density plot illustrating the distribution of inbreeding coefficients ($F_{ROH}$) across the ancestry groups for (A) cases and controls, (B) cases only, and (C) controls only. The x-axis represents the inbreeding coefficient values ranging from -0.5 to 0.25, while the y-axis represents the density of observations. AAC = African Admixed, AFR = African, AJ = Ashkenazi Jewish, AMR = American Admixed, CAS = Central Asian, EAS = East Asian, EUR = European, MDE = Middle-Eastern, SAS = South Asian.*

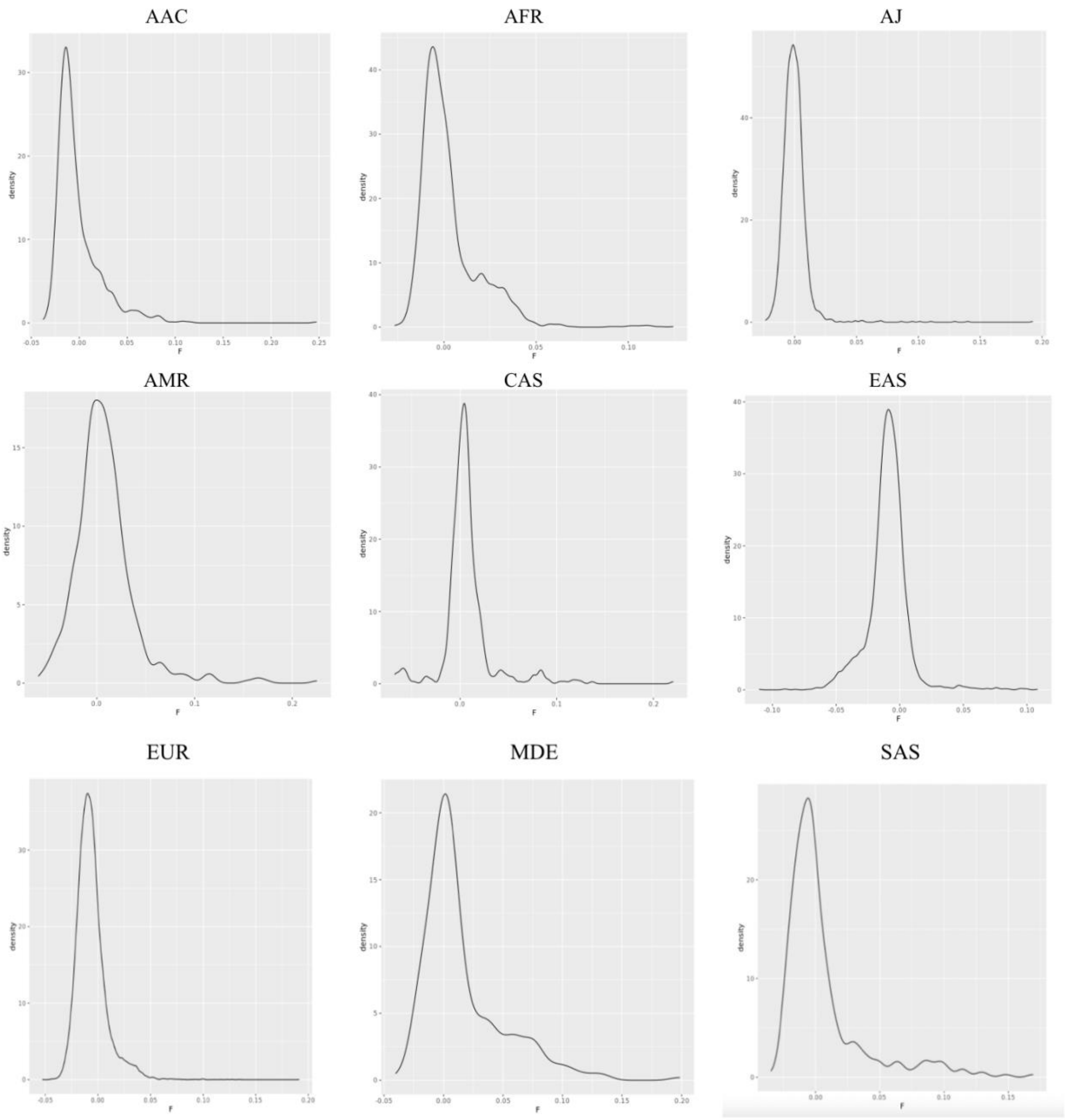

***Supplementary Figure 7:*** *Density plots illustrating the distribution of inbreeding coefficients ($F_{ROH}$) across the ancestral groups. The x-axis represents the $F_{ROH}$ values, while the y-axis represents the density of observations at each F value. AAC = African Admixed, AFR = African, AJ = Ashkenazi Jewish, AMR = American Admixed, CAS = Central Asian, EAS = East Asian, EUR = European, MDE = Middle-Eastern, SAS = South Asian*

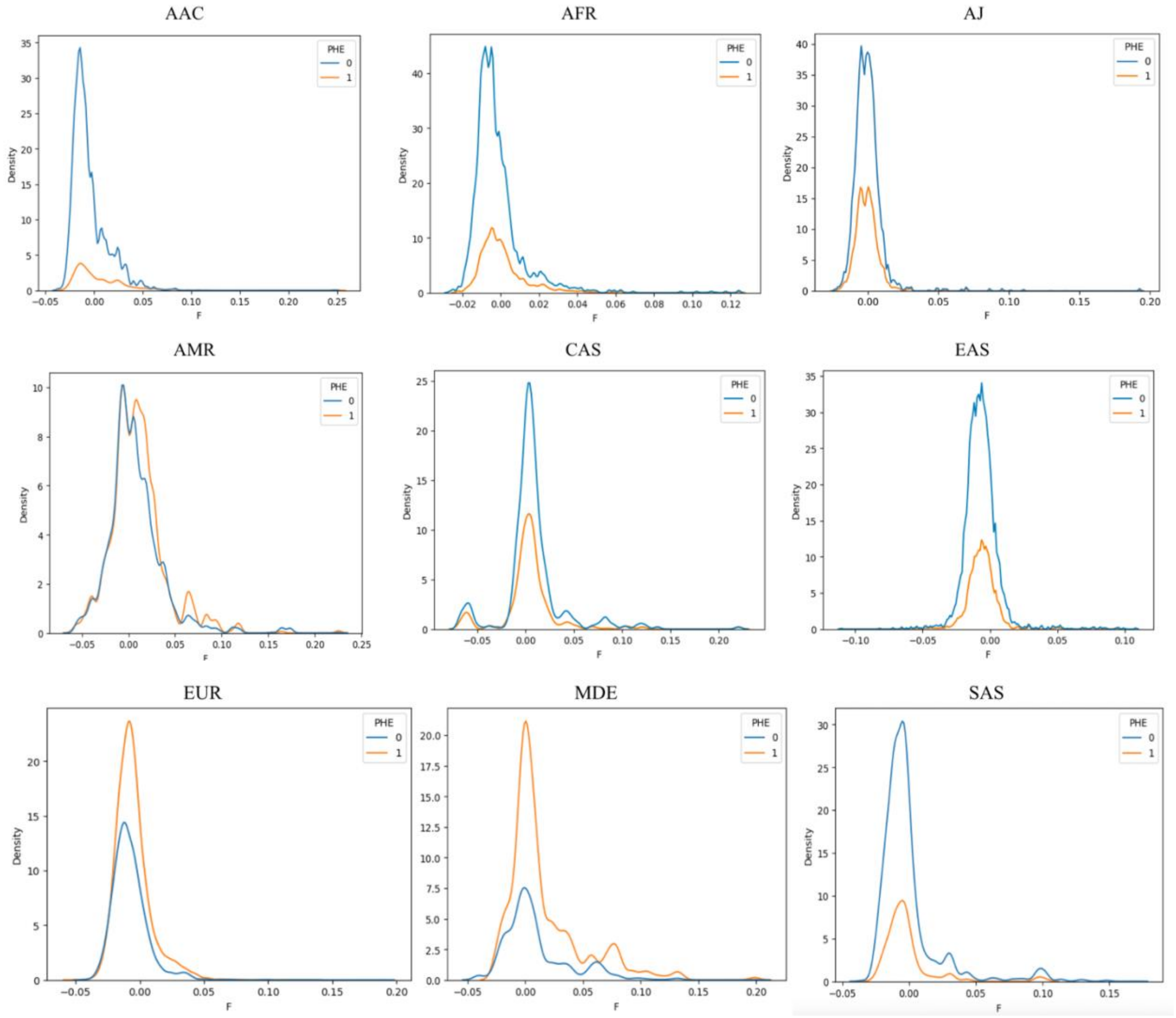

***Supplementary Figure 8:*** *Density plots illustrating the distribution of inbreeding coefficients ($F_{ROH}$) colored by phenotype for each ancestral group. For phenotype, 0 indicates control and 1 indicates case. Each curve represents the density of observations for a specific phenotype value. The x-axis represents the values of F, while the y-axis represents the density of observations at each F value. AAC = African Admixed, AFR = African, AJ = Ashkenazi Jewish, AMR = American Admixed, CAS = Central Asian, EAS = East Asian, EUR = European, MDE = Middle-Eastern, SAS = South Asian.*

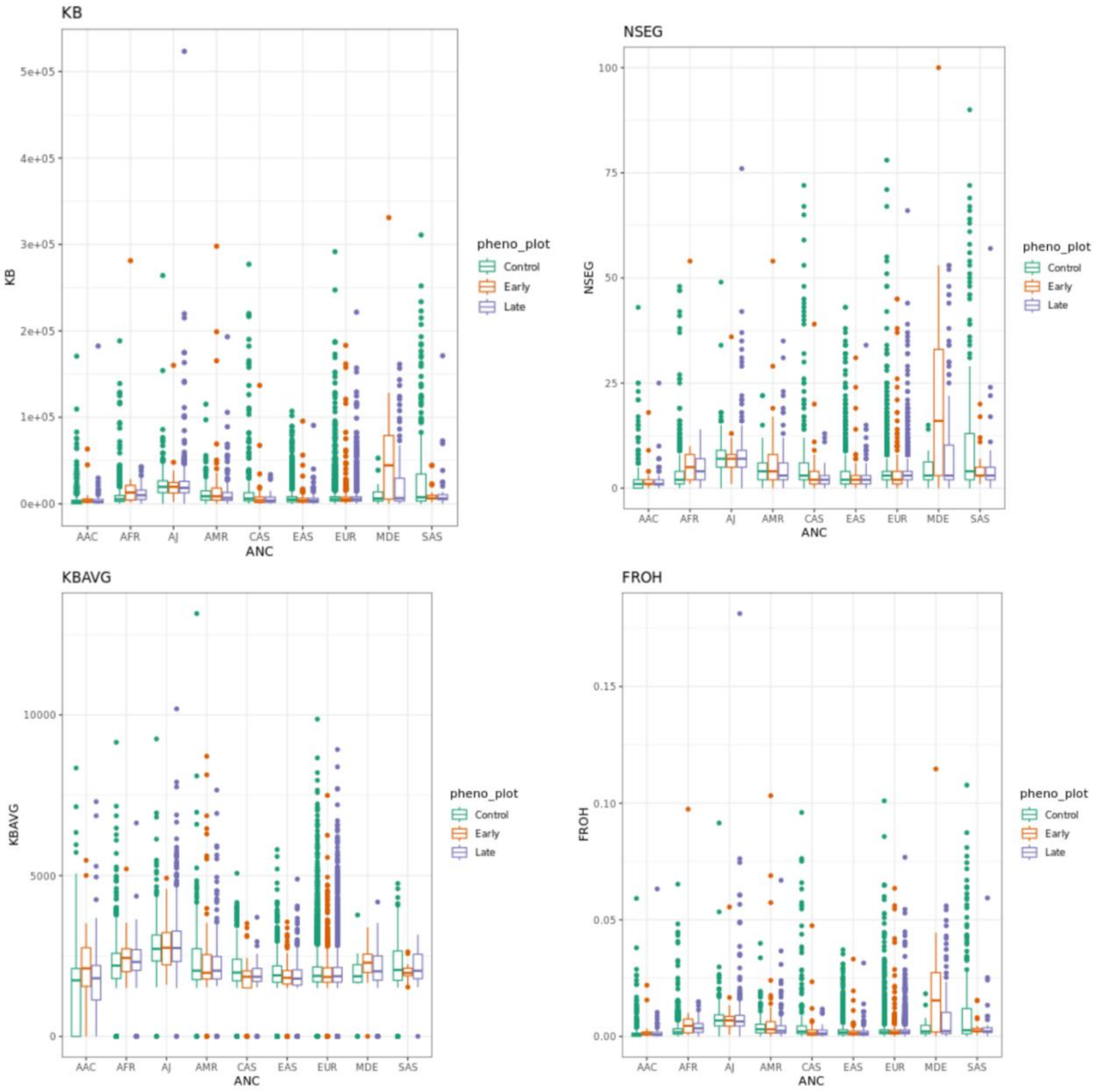

***Supplementary Figure 9:*** *Box plots illustrating the distribution of four ROH parameters assessed colored by phenotype including early and late onset PD for each ancestral group. The y-axis represents the ancestral group. AAC = African Admixed, AFR = African, AJ = Ashkenazi Jewish, AMR = American Admixed, ANC = ancestry, CAS = Central Asian, EAS = East Asian, EUR = European,* $F_{ROH}$ *= run of homozygosity based estimates for the inbreeding coefficient, KB = kilobases, KBAVG = average length of runs of homozygosity, MDE = Middle-Eastern, NSEG = number of runs of homozygosity, SAS = South Asian.*

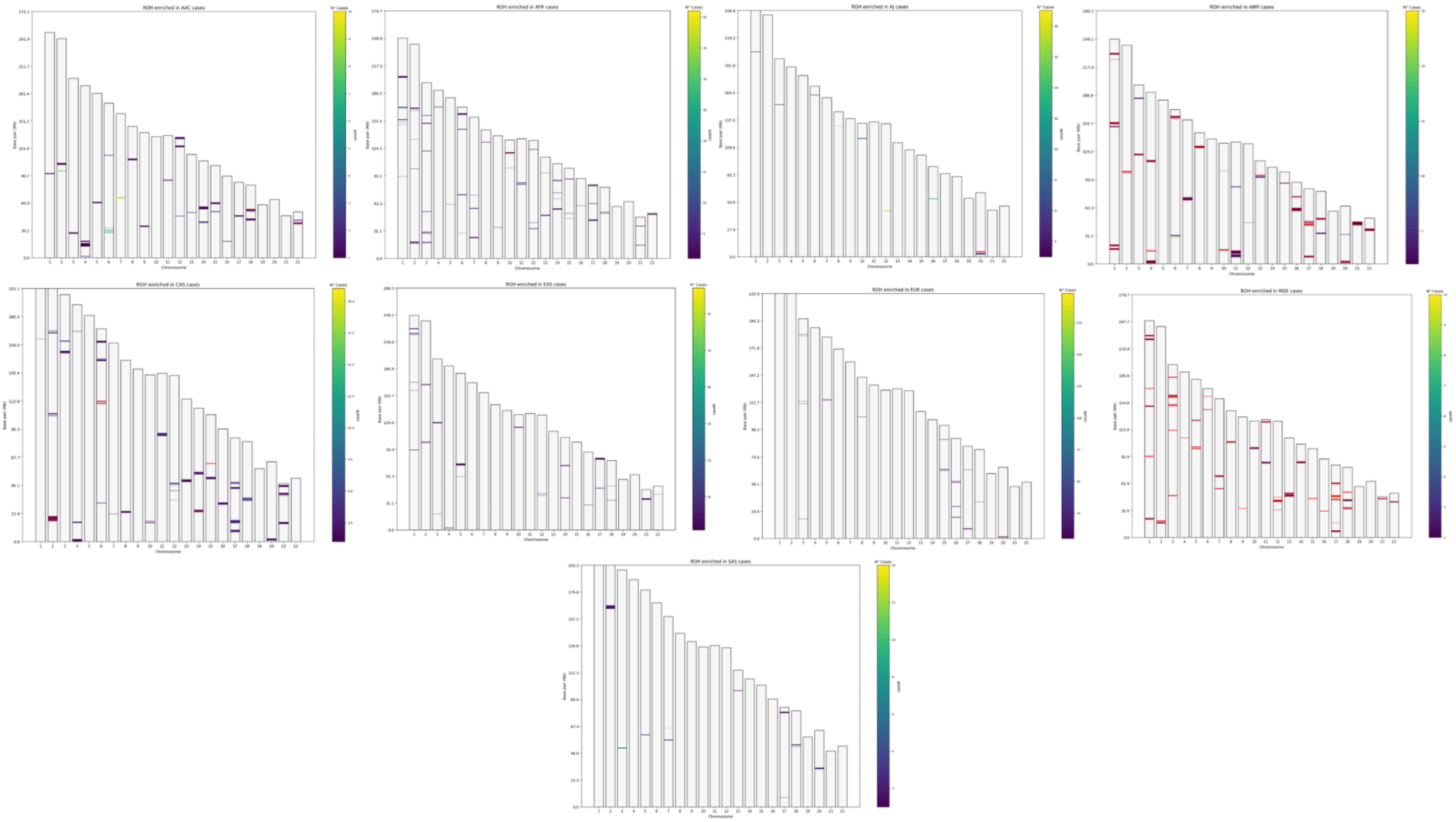

***Supplementary Figure 10**: Homozygosity mapping plots displaying runs of homozygosity enriched in cases across diverse ancestries. Highlighted in red are the runs of homozygosity present only in cases. The chromosome is along the x-axis and the runs of homozygosity size in base positions in along the y-axis. AAC = African Admixed, AFR = African, AJ = Ashkenazi Jewish, AMR = American Admixed, CAS = Central Asian, EAS = East Asian, EUR = European, MDE = Middle-Eastern, SAS = South Asian.*

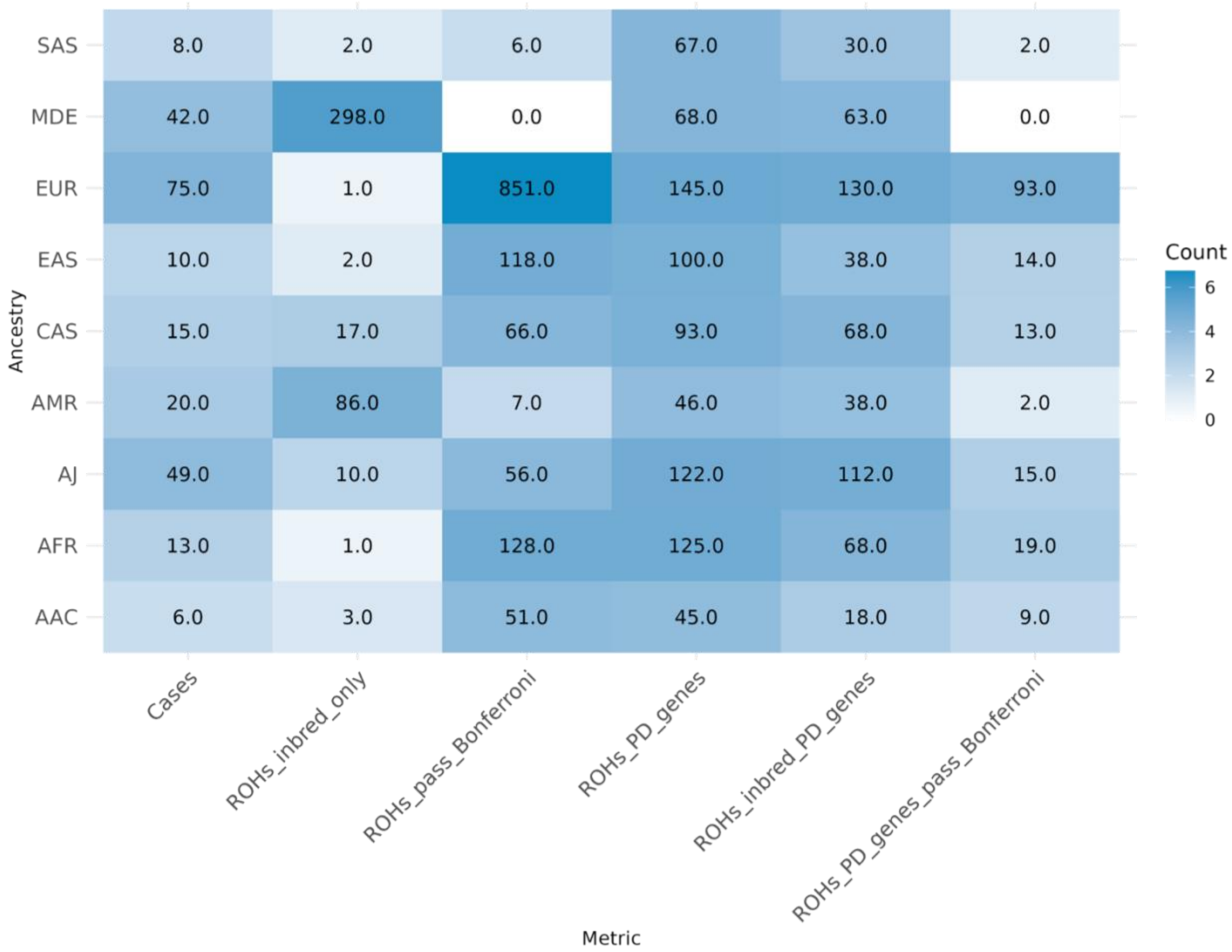

***Supplementary Figure 11:*** *Heatmap illustrating the number of individuals or runs of homozygosity obtained from the homozygosity mapping analysis for individuals with consanguinity. AAC = African Admixed, AFR = African, AJ = Ashkenazi Jewish, AMR = American Admixed, CAS = Central Asian, EAS = East Asian, EUR = European, MDE = Middle-Eastern, SAS = South Asian. The Bonferroni threshold was adjusted by dividing 0.05 by the number of enriched ROHs to account for multiple comparisons.*

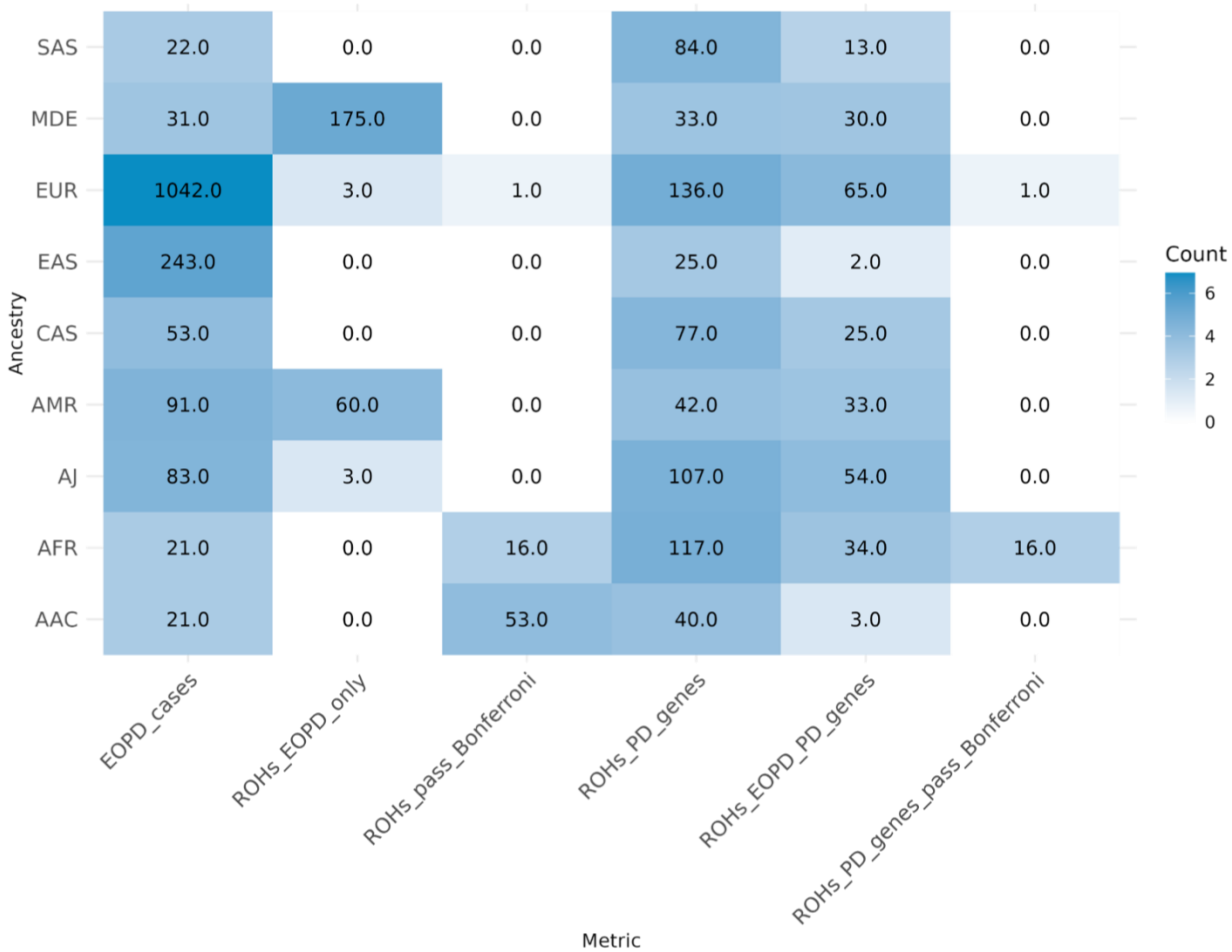

***Supplementary Figure 12:*** *Heatmap illustrating the number of individuals or runs of homozygosity obtained from the homozygosity mapping analysis for early-onset Parkinson's disease patients. AAC = African Admixed, AFR = African, AJ = Ashkenazi Jewish, AMR = American Admixed, CAS = Central Asian, EAS = East Asian, EUR = European, MDE = Middle-Eastern, SAS = South Asian. The Bonferroni threshold was adjusted by dividing 0.05 by the number of enriched ROHs to account for multiple comparisons.*

### Supplementary Tables

***Supplementary Table 1:*** *Study participant demographic information for NeuroBooster array genotyping imputed data*

| Ancestry | Total participants | Total PD cases | Total controls | Total unrelated participants | Total unrelated cases | Total unrelated controls |
|---|---|---|---|---|---|---|
| **AAC** | 1,065 | 257 | 808 | 1,060 | 256 | 804 |
| **AFR** | 2,603 | 920 | 1,683 | 2,548 | 917 | 1,631 |
| **AJ** | 1,470 | 1,011 | 459 | 1,459 | 1,001 | 458 |
| **AMR** | 506 | 367 | 139 | 502 | 363 | 139 |
| **CAS** | 580 | 286 | 294 | 532 | 265 | 267 |
| **EAS** | 3,955 | 1,577 | 2,378 | 3,839 | 1,537 | 2,302 |
| **EUR** | 19,437 | 11,841 | 7,596 | 19,184 | 11,679 | 7,505 |
| **MDE** | 239 | 218 | 21 | 234 | 214 | 20 |
| **SAS** | 318 | 111 | 207 | 315 | 117 | 198 |
| **Total** | **30,173** | **16,588** | **13,585** | **29,673** | **16,349** | **13,324** |
| Legend: AAC = African Admixed, AFR = African, AJ = Ashkenazi Jewish, AMR = American Admixed, CAS = Central Asian, EAS = East Asian, EUR = European, MDE = Middle-Eastern, PD = Parkinson's disease, SAS = South Asian | | | | | | |

***Supplementary Table 2:*** *Study participant demographic information for whole-genome sequencing data*

| Ancestry | Total participants | Total PD cases | Total controls |
|---|---|---|---|
| **AAC** | 41 | 36 | 5 |
| **AFR** | 208 | 149 | 59 |
| **AJ** | 165 | 163 | 2 |
| **AMR** | 155 | 152 | 3 |
| **CAS** | 71 | 71 | 0 |
| **EAS** | 1,234 | 1,219 | 15 |
| **EUR** | 4,005 | 3,770 | 235 |
| **MDE** | 110 | 110 | 0 |
| **FIN** | 11 | 11 | 0 |
| **SAS** | 209 | 209 | 0 |
| **Total** | **6,209** | **5,890** | **319** |
| Legend: AAC = African Admixed, AFR = African, AJ = Ashkenazi Jewish, AMR = American Admixed, CAS = Central Asian, EAS = East Asian, EUR = European, MDE = Middle-Eastern, SAS = South Asian | | | |

***Supplementary Table 3:*** *Parameters used for the genome-wide homozygosity analyses*

| PLINK input flags | Flag description | Default threshold | Threshold used |
|---|---|---|---|
| **--homozyg** | Performs ROH analysis and generates .hom file | N/A | N/A |
| **--homozyg-density** | Minimum SNP density (kb/SNP) per ROH segment | 50 | 50 |
| **--homozyg-gap** | Maximum gap between two SNPs before splitting ROH segment | 1,000 | 1,000 |
| **--homozyg-group** | Generates .hom.overlap file | N/A | N/A |
| **--homozyg-kb** | Minimum length for calling a ROH | 1,000 | 1,500 |
| **--homozyg-snp** | Minimum number of SNPs for calling a ROH | 100 | 100 |
| **--homozyg-window-het** | Maximum number of heterozygous SNPs in a ROH | 1 | 1 |
| **--homozyg-window-missing** | Maximum number of missing SNPs | 5 | 5 |
| **--homozyg-window-snp** | Scanning window size in SNPs | 50 | 50 |
| **--homozyg-window-threshold** | Window threshold for calling a ROH | 0.05 | 0.05 |
| Legend: kb = kilobase, ROH = runs of homozygosity, SNPs = single nucleotide polymorphisms, N/A = not applicable | | | |

***Supplementary Table 4:*** *Missing age information by Ancestry and the corresponding mean age for imputation*

| Ancestry | Number of individuals | Individuals with missing age | Age (mean ± SD) | Case age (mean ± SD) | Control age (mean ± SD) |
|---|---|---|---|---|---|
| **AAC** | 1,060 | 154 | 66 ± 10 | 66 ± 11 | 66 ± 10 |
| **AFR** | 2,548 | 1,493 | 63 ± 15 | 63 ± 12 | 64 ± 15 |
| **AJ** | 1,459 | 267 | 69 ± 10 | 69 ± 10 | 68 ± 10 |
| **AMR** | 502 | 105 | 60 ± 12 | 60 ± 12 | 63 ± 9 |
| **CAS** | 532 | 152 | 57 ± 10 | 59 ± 12 | 55 ± 6 |
| **EAS** | 3,839 | 1,132 | 64 ± 11 | 66 ± 10 | 62 ± 11 |
| **EUR** | 19,184 | 5,438 | 66 ± 11 | 67 ± 11 | 65 ± 11 |
| **MDE** | 234 | 18 | 65 ± 13 | 65 ± 13 | 67 ± 14 |
| **SAS** | 315 | 109 | 57 ± 16 | 62 ± 13 | 54 ± 17 |
| Legend: AAC = African Admixed, AFR = African, AJ = Ashkenazi Jewish, AMR = American Admixed, CAS = Central Asian, EAS = East Asian, EUR = European, MDE = Middle-Eastern, SAS = South Asian, SD = standard deviation | | | | | |

***Supplementary Table 5:*** *List of the known Parkinson's disease, pallido-pyramidal syndrome, and atypical parkinsonism gene regions and risk loci*

| Marker name | Near gene | Source |
|---|---|---|
| chr5:75803383-76803383 | *SV2C* | (Foo et al., 2020) |
| chr7:70785507-71785507 | *GALNT17* | (Foo et al., 2020) |
| chr22:49869588-50869588 | *PPP6R2* | (Kim et al., 2024) |
| chr13:109310821-110310821 | *RN7SKP10* | (Kim et al., 2024) |
| chr1:92586630-93586630 | *MTF2* | (Kim et al., 2024) |
| chr17:15830946-16830946 | *PIGL* | (Kim et al., 2024) |
| chr4:2347441-3347441 | *ADD1* | (Kim et al., 2024) |
| chr21:14940233-15940233 | *AJ009632.2* | (Kim et al., 2024) |
| chr15:49995212-50995212 | *USP8* | (Kim et al., 2024) |
| chr17:81587210-82587210 | *FASN* | (Kim et al., 2024) |
| chr20:31333121-32333121 | *MYLK2* | (Kim et al., 2024) |
| chr8:109132545-110132545 | *SYBU* | (Kim et al., 2024) |

| Marker name | Near gene | Source |
|---|---|---|
| chr3:178643629-179643629 | *AC076966.1* | (Kim et al., 2024) |
| chr22:40538154-41538154 | *RBX1* | (Kim et al., 2024) |
| chr16:52435514-53435514 | *PHBP2* | (Nalls et al., 2019) |
| chr3:182542285-183542285 | *MCCC1* | (Nalls et al., 2019) |
| chr10:101755522-102755522 | *GBF1* | (Nalls et al. 2019) |
| chr9:17227067-18227067 | *SH3GL2* | (Nalls et al. 2019) |
| chr1:232028865-233028865 | *SIPA1L2* | (Nalls et al. 2019) |
| chr12:122342051-123342051 | *HIP1R* | (Nalls et al. 2019) |
| chr16:30466478-31466478 | *SETD1A* | (Nalls et al. 2019) |
| chr14:54382151-55382151 | *GCH1* | (Nalls et al. 2019) |
| chr6:32110995-33110995 | *AHR* | (Nalls et al. 2019) |
| chr1:154425709-155425709 | *PMVK* | (Nalls et al. 2019) |
| chr1:205268611-206268611 | *RAB29* | (Nalls et al. 2019) |
| chr1:171250629-172250629 | *VAMP4* | (Nalls et al. 2019) |
| chr12:131987182-132987182 | *FBRSL1* | (Nalls et al. 2019) |
| chr17:46289439-47289439 | *WNT3* | (Nalls et al. 2019) |
| chr2:101280501-102280501 | *MAP4K4* | (Nalls et al. 2019) |
| chr3:150891177-151891177 | *MED12L* | (Nalls et al. 2019) |
| chr17:45220942-46220942 | *MAPT* | (Nalls et al. 2019) |
| chr10:119276815-120276815 | *INPP5F* | (Nalls et al. 2019) |
| chr5:134363415-135363415 | *AC006077.1* | (Nalls et al. 2019) |
| chr14:37020065-38020065 | *MIPOL1* | (Nalls et al. 2019) |
| chr11:83276234-84276234 | *DLG2* | (Nalls et al. 2019) |
| chr18:42593415-43593415 | *RIT2* | (Nalls et al. 2019) |
| chr3:48211556-49211556 | *IP6K2* | (Nalls et al. 2019) |
| chr6:71278059-72278059 | *RIMS1* | (Nalls et al. 2019) |
| chr17:6952302-7952302 | *MAPT* | (Nalls et al. 2019) |
| chr8:11354934-12354934 | *CTSB* | (Nalls et al. 2019) |
| chr17:42088995-43088995 | *RETREG3* | (Nalls et al. 2019) |

| Marker name | Near gene | Source |
|---|---|---|
| chr4:112947909-113947909 | *CAMK2D* | (Nalls et al. 2019) |
| chr9:17079692-18079692 | *SH3GL2* | (Nalls et al. 2019) |
| chr3:160859842-161859842 | *SPTSSB* | (Nalls et al. 2019) |
| chr2:167753884-168753884 | *STK39* | (Nalls et al. 2019) |
| chr5:60342132-61342132 | *ELOVL7* | (Nalls et al. 2019) |
| chr18:33224354-34224354 | *ASXL3* | (Nalls et al. 2019) |
| chr7:22760430-23760430 | *GPNMB* | (Nalls et al. 2019) |
| chr2:94835195-95835195 | *KCNIP3* | (Nalls et al. 2019) |
| chr8:129389663-130389663 | *FAM49B* | (Nalls et al. 2019) |
| chr21:36980059-37980059 | *DYRK1A* | (Nalls et al. 2019) |
| chr15:61205186-62205186 | *lincRNA* | (Nalls et al. 2019) |
| chr17:43716969-44716969 | *UBTF* | (Nalls et al. 2019) |
| chr8:22168467-23168467 | *BIN3* | (Nalls et al. 2019) |
| chr5:102530090-103530090 | *PAM* | (Nalls et al. 2019) |
| chr16:28433075-29433075 | *CD19* | (Nalls et al. 2019) |
| chr16:52102330-53102330 | *CASC16* | (Nalls et al. 2019) |
| chr4:17467188-18467188 | *LCORL* | (Nalls et al. 2019) |
| chr4:458159-1458159 | *TMEM175* | (Nalls et al. 2019) |
| chr12:39840400-40840400 | *LRRK2* | (Nalls et al. 2019) |
| chr4:89204960-90204960 | *AC097478.1* | (Nalls et al. 2019) |
| chr1:154662560-155662560 | *KRTCAP2* | (Nalls et al. 2019) |
| chr14:74406331-75406331 | *RPS6KL1* | (Nalls et al. 2019) |
| chr11:133417106-134417106 | *IGSF9B* | (Nalls et al. 2019) |
| chr4:75726816-76726816 | *FAM47E* | (Nalls et al. 2019) |
| chr6:27271022-28271022 | *RSL24D1P1* | (Nalls et al. 2019) |
| chr1:226228377-227228377 | *ITPKB* | (Nalls et al. 2019) |
| chr4:15235725-16235725 | *BST1* | (Nalls et al. 2019) |
| chr13:96712767-97712767 | *MBNL2* | (Nalls et al. 2019) |
| chr4:89215479-90215479 | *AC097478.1* | (Nalls et al. 2019) |

| Marker name | Near gene | Source |
|---|---|---|
| chr19:1841049-2841049 | *SPPL2B* | (Nalls et al. 2019) |
| chr3:121978045-122978045 | *KPNA1* | (Nalls et al. 2019) |
| chr2:134207046-135207046 | *TMEM163* | (Nalls et al. 2019) |
| chr17:61340005-62340005 | *BRIP1* | (Nalls et al. 2019) |
| chr8:16340084-17340084 | *AC011586.2* | (Nalls et al. 2019) |
| chr17:45166837-46166837 | *MAPT* | (Nalls et al. 2019) |
| chr4:169162006-170162006 | *CLCN3* | (Nalls et al. 2019) |
| chr9:33546393-34546393 | *UBAP2* | (Nalls et al. 2019) |
| chr16:18766171-19766171 | *SYT17* | (Nalls et al. 2019) |
| chr16:50202745-51202745 | *NOD2* | (Nalls et al. 2019) |
| chr1:160999264-161999264 | *FCGR2A* | (Nalls et al. 2019) |
| chr17:77929399-78929399 | *DNAH17* | (Nalls et al. 2019) |
| chr3:28164199-29164199 | *RBMS3* | (Nalls et al. 2019) |
| chr4:75689212-76689212 | *SCARB2* | (Nalls et al. 2019) |
| chr4:75776901-76776901 | *FAM47E* | (Nalls et al. 2019) |
| chr12:45525303-46525303 | *SLC38A1* | (Nalls et al. 2019) |
| chr10:119156173-120156173 | *BAG3* | (Nalls et al. 2019) |
| chr3:17820267-18820267 | *TBC1D5* | (Nalls et al. 2019) |
| chr6:132389222-133389222 | *HMGB1P13* | (Nalls et al. 2019) |
| chr2:17466582-18466582 | *KCNS3* | (Nalls et al. 2019) |
| chr1:154735843-155735843 | *GBA* | (Nalls et al. 2019) |
| chr12:39720632-40720632 | *LRRK2* | (Nalls et al. 2019) |
| chr7:66044864-67044864 | *AC006001.3* | (Nalls et al. 2019) |
| chr20:5525395-6525395 | *CRLS1* | (Nalls et al. 2019) |
| chr11:10037230-11037230 | *RNF141* | (Nalls et al. 2019) |
| chr18:50657219-51657219 | *SMAD4* | (Nalls et al., 2019) |
| chr1:205254444-206254444 | *RAB29* | (Nalls et al., 2019) |
| chr17:43857262-44857262 | *FAM171A2* | (Nalls et al., 2019) |
| chr4:431588-1431588 | *GAK* | (Nalls et al., 2019) |

| Marker name | Near gene | Source |
|---|---|---|
| chr10:15015407-16015407 | *ITGA8* | (Nalls et al., 2019) |
| chr6:29640906-30640906 | *TRIM40* | (Nalls et al., 2019) |
| chr13:48853596-49853596 | *CAB39L* | (Nalls et al., 2019) |
| chr14:87497920-88497920 | *GPR65* | (Nalls et al., 2019) |
| chr6:111422088-112422088 | *WISP3* | (Nalls et al., 2019) |
| chr1:154735878-155735878 | *GBA1* | (Rizig et al., 2023) |
| chr14:24713480-25147976 | *STXBP6* | (Loesch et al., 2021) |
| chr4:89695550-89866846 | *SNCA* | Known PD genes |
| chr6:161002496-163072863 | *PARK2* | Known PD genes |
| chr4:41253993-41271316 | *UCHL1* | Known PD genes |
| chr1:7955746-7991189 | *DJ-1* | Known PD genes |
| chr1:155264661-155274879 | *GBA* | Known PD genes |
| chr12:40188943-40405353 | *LRRK2* | Known PD genes |
| chr1:20628940-20656025 | *PINK1* | Known PD genes |
| chr1:16979465-17018421 | *ATP13A2* | Known PD genes |
| chr22:38093930-38199319 | *PLA2G6* | Known PD genes |
| chr22:32468692-32504859 | *FBXO7* | Known PD genes |
| chr3:184309279-184340574 | *EIF4G1* | Known PD genes |
| chr16:46652288-46696621 | *VPS35* | Known PD genes |
| chr1:226481196-226488644 | *ITPKB* | Known PD genes |
| chr2:101375382-101412082 | *IL1R2* | Known PD genes |
| chr2:164231020-164347533 | *SCN3A* | Known PD genes |
| chr3:18306448-18397281 | *SATB1* | Known PD genes |
| chr3:48636411-48648494 | *NCKIPSD* | Known PD genes |
| chr3:52164078-52180311 | *ALAS1* | Known PD genes |
| chr3:52187064-52192147 | *TLR9* | Known PD genes |
| chr3:52282303-52366475 | *DNAH1* | Known PD genes |
| chr3:52366997-52376334 | *BAP1* | Known PD genes |
| chr3:52376641-52389625 | *PHF7* | Known PD genes |

| Marker name | Near gene | Source |
|---|---|---|
| chr3:52421102-52459052 | *NISCH* | Known PD genes |
| chr3:52461322-52490479 | *STAB1* | Known PD genes |
| chr3:52760782-52774993 | *ITIH3* | Known PD genes |
| chr3:52789919-52791298 | *ITIH4* | Known PD genes |
| chr4:111896953-112432557 | *ANK2* | Known PD genes |
| chr4:112529876-112840771 | *CAMK2D* | Known PD genes |
| chr5:61455964-61548562 | *ELOVL7* | Known PD genes |
| chr6:27482964-27505339 | *ZNF184* | Known PD genes |
| chr8:11985015-12010720 | *CTSB* | Known PD genes |
| chr8:22694182-22718275 | *SORBS3* | Known PD genes |
| chr8:22721228-22736785 | *PDLIM2* | Known PD genes |
| chr8:22742134-22746637 | *C8orf58* | Known PD genes |
| chr8:22762905-22811607 | *BIN3* | Known PD genes |
| chr9:17579125-17797131 | *SH3GL2* | Known PD genes |
| chr10:15169644-15329060 | *FAM171A1* | Known PD genes |
| chr14:87466670-87526927 | *GALC* | Known PD genes |
| chr16:19056277-19068773 | *COQ7* | Known PD genes |
| chr16:52404096-52513096 | *TOX3* | Known PD genes |
| chr17:44381512-44445214 | *ATP6V0A1* | Known PD genes |
| chr17:44494947-44500427 | *PSMC3IP* | Known PD genes |
| chr17:44581937-44589638 | *TUBG2* | Known PD genes |
| chr3:87178135-87206349 | *CHMP2B* | Known PD genes |
| chr3:183298467-183300020 | *MCCC1* | Known PD genes |
| chr4:75326511-75385910 | *FAM47E* | Known PD genes |
| chr4:15680106-15688816 | *FAM200B* | Known PD genes |
| chr8:17169729-17265135 | *MICU3* | Known PD genes |
| chr10:117891859-117918308 | *BAG3* | Known PD genes |
| chr11:85179700-85179917 | *DLG2* | Known PD genes |
| chr11:134028609-134028686 | *MIR4697* | Known PD genes |

| Marker name | Near gene | Source |
|---|---|---|
| chr12:39831272-39975479 | *LRRK2* | Known PD genes |
| chr12:122490673-122495494 | *OGFOD2* | Known PD genes |
| chr14:54375298-54436134 | *GCH1* | Known PD genes |
| chr14:67003549-67048754 | *TMEM229B* | Known PD genes |
| chr15:61560190-61768274 | *VPS13C* | Known PD genes |
| chr16:31063101-31072875 | *ZNF646* | Known PD genes |
| chr18:45687913-45697685 | *SYT4* | Known PD genes |
| chr19:2321518-2328621 | *LSM7* | Known PD genes |
| chr20:3209704-3224039 | *DDRGK1* | Known PD genes |
| chr17:47952550-48146446 | *KANSL1* | Known PD genes |
| chr17:47921884-47922328 | *STH* | Known PD genes |
| chr17:47767469-47769706 | *SPPL2C* | Known PD genes |
| chr17:47706954-47758460 | *CRHR1* | Known PD genes |
| chr17:47351987-47355550 | *ARHGAP27* | Known PD genes |
| chr16:31104433-31120072 | *KAT8* | Known PD genes |
| chr7:23206944-23235489 | *GPNMB* | Known PD genes |
| chr4:15776655-15851607 | *CD38* | Known PD genes |
| chr2:167097510-167391631 | *STK39* | Known PD genes |
| chr7:23142208-23161390 | *NUPL2* | Known PD genes |
| chr7:23066165-23138295 | *KLHL7* | Known PD genes |
| chr6:32669654-32679285 | *HLA-DQA1* | Known PD genes |
| chr6:32584936-32592225 | *HLA-DRB6* | Known PD genes |
| chr4:965099-979768 | *DGKQ* | Known PD genes |
| chr2:134190255-134195627 | *CCNT2* | Known PD genes |
| chr4:938599-964868 | *TMEM175* | Known PD genes |
| chr2:133698188-133961429 | *TMEM163* | Known PD genes |
| chr1:232262219-232425812 | *SIPA1L2* | Known PD genes |
| chr1:205819965-205844048 | *SLC41A1* | Known PD genes |
| chr7:56101569-56106576 | *CHCHD2* | Known PD genes |

| Marker name | Near gene | Source |
|---|---|---|
| chr20:3888839-3926958 | *PANK2* | Known PD genes |
| chr2:74361154-74392087 | *DCTN1* | Known PD genes |
| chr15:89316303-89334847 | *POLG1* | Known PD genes |
| chr1:151220451-153220451 | *HRNR* | Known PD genes |
| chr1:153925709-155928394 | *PMVK* | Known PD genes |
| chr1:154162560-156165245 | *KRTCAP2* | Known PD genes |
| chr1:154233158-156235843 | *GBAP1* | Known PD genes |
| chr1:154404265-156406949 | *ASH1L* | Known PD genes |
| chr1:155182384-157185070 | *SEMA4A* | Known PD genes |
| chr1:160499264-162499264 | *FCGR2A* | Known PD genes |
| chr1:170750628-172750629 | *VAMP4* | Known PD genes |
| chr1:206009146-206268644 | *NUCKS1* | Known PD genes |
| chr1:206009146-206268644 | *RAB29* | Known PD genes |
| chr1:225728376-227728377 | *ITPKB* | Known PD genes |
| chr1:231528865-233528865 | *SIPA1L2* | Known PD genes |
| chr2:16966581-18966570 | *KCNS3* | Known PD genes |
| chr9:40365644-40365693 | *KCNIP3* | Known PD genes |
| chr2:100780501-102780504 | *MAP4K4* | Known PD genes |
| chr2:133681218-135681219 | *TMEM163* | Known PD genes |
| chr2:167253884-169253884 | *STK39* | Known PD genes |
| chr3:17320267-19320267 | *SATB1* | Known PD genes |
| chr3:27664199-29664199 | *LINC00693* | Known PD genes |
| chr3:47707499-49711556 | *IP6K2* | Known PD genes |
| chr3:121478045-123478045 | *KPNA1* | Known PD genes |
| chr3:150391178-152391176 | *MED12L* | Known PD genes |
| chr3:160359842-162359842 | *SPTSSB* | Known PD genes |
| chr3:182042285-184042285 | *MCCC1* | Known PD genes |
| chr4:849276-932373 | *GAK* | Known PD genes |
| chr4:932387-958656 | *TMEM175* | Known PD genes |

| Marker name | Near gene | Source |
|---|---|---|
| chr4:14735724-16735725 | *BST1* | Known PD genes |
| chr4:16967188-18967188 | *LCORL* | Known PD genes |
| chr4:75185155-77189212 | *SCARB2* | Known PD genes |
| chr4:75222759-77226816 | *FAM47E* | Known PD genes |
| chr4:75272844-77276901 | *FAM47E-STBD1* | Known PD genes |
| chr4:88744890-90685975 | *SNCA* | Known PD genes |
| chr4:112447909-114447909 | *CAMK2D* | Known PD genes |
| chr4:168662006-170662006 | *CLCN3* | Known PD genes |
| chr5:59842133-61842132 | *ELOVL7* | Known PD genes |
| chr5:102030090-104030093 | *PAM* | Known PD genes |
| chr5:123774579-125774580 | *ZNF608* | Known PD genes |
| chr5:133863414-135863416 | *C5orf24* | Known PD genes |
| chr6:26737023-28771024 | *LOC100131289* | Known PD genes |
| chr6:29140906-31140906 | *TRIM40* | Known PD genes |
| chr6:30878457-32878457 | *SLC44A4* | Known PD genes |
| chr6:31610995-33610995 | *HLA-DRB5* | Known PD genes |
| chr6:70778059-72778039 | *RIMS1* | Known PD genes |
| chr6:110922088-112922089 | *FYN* | Known PD genes |
| chr6:131889221-133889223 | *RPS12* | Known PD genes |
| chr7:22260430-24260430 | *GPNMB* | Known PD genes |
| chr7:65544938-67544864 | *GS1-124K5.11* | Known PD genes |
| chr8:10854933-12854934 | *CTSB* | Known PD genes |
| chr8:15840084-17840084 | *FGF20* | Known PD genes |
| chr8:21668468-23668467 | *BIN3* | Known PD genes |
| chr8:128889663-130889663 | *FAM49B* | Known PD genes |
| chr9:16727067-18579692 | *SH3GL2* | Known PD genes |
| chr9:33046393-35046394 | *UBAP2* | Known PD genes |
| chr10:14515407-16515407 | *ITGA8* | Known PD genes |
| chr10:101255522-103255522 | *GBF1* | Known PD genes |

| Marker name | Near gene | Source |
|---|---|---|
| chr10:118656173-120656173 | *BAG3* | Known PD genes |
| chr10:118776815-120776815 | *INPP5F* | Known PD genes |
| chr11:9537230-11537230 | *RNF141* | Known PD genes |
| chr11:82776235-84776234 | *DLG2* | Known PD genes |
| chr11:132917106-134917107 | *IGSF9B* | Known PD genes |
| chr12:31657825-33657824 | *FGD4* | Known PD genes |
| chr12:31826971-33826970 | *PKP2* | Known PD genes |
| chr12:33792942-35252126 | *ALG10* | Known PD genes |
| chr12:39528770-41220632 | *LRRK2* | Known PD genes |
| chr12:40053867-41542837 | *CNTN1* | Known PD genes |
| chr12:40477002-42477002 | *PDZRN4* | Known PD genes |
| chr12:41141601-42901976 | *GXYLT1* | Known PD genes |
| chr12:45025303-47025303 | *SCAF11* | Known PD genes |
| chr12:121888692-123842051 | *HIP1R* | Known PD genes |
| chr12:132241105-132241213 | *FBRSL1* | Known PD genes |
| chr13:48353596-50353596 | *CAB39L* | Known PD genes |
| chr13:96212767-98212767 | *MBNL2* | Known PD genes |
| chr14:36520065-38520066 | *MIPOL1* | Known PD genes |
| chr14:53882151-55882151 | *GCH1* | Known PD genes |
| chr14:73906331-75906691 | *RPS6KL1* | Known PD genes |
| chr14:86997920-88997920 | *GALC* | Known PD genes |
| chr15:60705186-62705186 | *VPS13C* | Known PD genes |
| chr16:18183636-20266171 | *SYT17* | Known PD genes |
| chr16:27933075-29933075 | *CD19* | Known PD genes |
| chr16:29966478-31966478 | *SETD1A* | Known PD genes |
| chr16:49702745-51702745 | *NOD2* | Known PD genes |
| chr16:51602331-53602330 | *CASC16* | Known PD genes |
| chr16:51935514-53935514 | *CHD9* | Known PD genes |
| chr16:57553760-59553768 | *CNOT1* | Known PD genes |

| Marker name | Near gene | Source |
|---|---|---|
| chr17:6452301-8452303 | *CHRNB1* | Known PD genes |
| chr17:41584761-43663645 | *RETREG3* | Known PD genes |
| chr17:43142320-45216970 | *UBTF* | Known PD genes |
| chr17:43357262-45357264 | *FAM171A2* | Known PD genes |
| chr17:44339905-46339907 | *MAP3K14* | Known PD genes |
| chr17:44720940-46666837 | *CRHR1* | Known PD genes |
| chr17:44855939-46855941 | *MAPT-AS1* | Known PD genes |
| chr17:45111699-47111701 | *KANSL1* | Known PD genes |
| chr17:45723580-47723580 | *NSF* | Known PD genes |
| chr17:45789439-47789439 | *WNT3* | Known PD genes |
| chr17:60840005-62840005 | *BRIP1* | Known PD genes |
| chr17:77429398-79429398 | *DNAH17* | Known PD genes |
| chr18:32724355-34724354 | *ASXL3* | Known PD genes |
| chr18:42093416-44093415 | *RIT2* | Known PD genes |
| chr18:50157219-52157219 | *MEX3C* | Known PD genes |
| chr19:1341048-3341049 | *SPPL2B* | Known PD genes |
| chr20:2184040-4184039 | *DDRGK1* | Known PD genes |
| chr20:5025395-7025394 | *CRLS1* | Known PD genes |
| chr21:36480063-38480438 | *DYRK1A* | Known PD genes |
| chr16:74212955-75274831 | *FA2H* | Pallido-pyramidal syndrome (Simón-Sánchez et al., 2012) |
| chr21:43219094-44262307 | *PDXK* | Pallido-pyramidal syndrome (Simón-Sánchez et al., 2012) |
| chr15:44054818-45163688 | *SPG11* | Pallido-pyramidal syndrome (Simón-Sánchez et al., 2012) |
| chr6:146043833-147054953 | *RAB32* | Known PD gene (Hop et al., 2024) |
| chr20:613240-1689415 | *PSMF1* | Known PD gene (Magrinelli et al., 2024) |
| Legend: chr = chromosome, PD = Parkinson's disease. Positions on GRCh38. | | |

***Supplementary Table 6:*** *Putative family structures identified in the Middle-Eastern group for runs of homozygosity segregating in families*

| FID | Phenotype | Family structure/relationship | Sex |
|---|---|---|---|
| KING1 | Case | | M |
| | Case | | M |
| KING2 | Case | | F |
| | Control | | F |
| | Control | | M |
| KING3 | Case | | M |
| | Control | | M |
| KING4 | Case | Sibship | M |
| | Case | | M |
| KING5 | Case | | F |
| | Case | | F |
| | Case | | F |
| Legend: FID = family ID, M = male, F = female | | | |

***Supplementary Table 7:*** *T-test results for measures of genome-wide homozygosity in Parkinson's disease*

| **Ancestry** | **SROH** | | | **AVROH** | | | **NROH** | | | **FROH** | | |
|---|---|---|---|---|---|---|---|---|---|---|---|---|
| | **Cases (mean ± SE)** | **Controls (mean ± SE)** | **p-value** | **Cases (mean ± SE)** | **Controls (mean ± SE)** | **p-value** | **Cases (mean ± SE)** | **Controls (mean ± SE)** | **p-value** | **Cases (mean ± SE)** | **Controls (mean ± SE)** | **p-value** |
| **AAC** | 6330.5 ± 989.3 | 4373.8 ± 378.8 | 0.066 | 1815.6 ± 84.6 | 1580.9 ± 40.8 | **0.013*** | 2.1 ± 0.2 | 1.7 ± 0.1 | 0.094 | 0.0022 ± 3E-04 | 0.0015 ± 1E-04 | 0.066 |
| **AFR** | 9820.3 ± 456.3 | 8158.4 ± 297.7 | **0.00232** | 2252.6 ± 28.4 | 2118.8 ± 23.4 | **2.8E-04*** ** | 4 ± 0.1 | 3.3 ± 0.1 | **4.92E-05*** ** | 0.0034 ± 2E-04 | 0.0028 ± 1E-04 | **0.002**** |
| **AJ** | 22595 ± 816.2 | 21501.1 ± 820 | 0.345 | 2879.5 ± 27.5 | 2826.3 ± 39.7 | 0.270 | 7.3 ± 0.2 | 7.2 ± 0.2 | 0.770 | 0.0078 ± 3E-04 | 0.0074 ± 3E-04 | 0.345 |
| **AMR** | 15162.8 ± 1491.6 | 13186.3 ± 1351 | 0.327 | 2302.8 ± 64.8 | 2483.4 ± 128.7 | 0.212 | 5.1 ± 0.3 | 4.6 ± 0.3 | 0.207 | 0.0053 ± 5E-04 | 0.0046 ± 5E-04 | 0.327 |
| **CAS** | 13658.5 ± 2093.5 | 19719.4 ± 2506.5 | 0.064 | 1885 ± 51.6 | 2087.3 ± 52.6 | **0.006**** | 5.2 ± 0.6 | 6.8 ± 0.7 | 0.073 | 0.0047 ± 7E-04 | 0.0068 ± 9E-04 | 0.064 |
| **EAS** | 5188.6 ± 201.3 | 7134.8 ± 197 | **5.72E-12** | 1735.7 ± 20.4 | 1854 ± 15 | **2.93E-06*** ** | 2.5 ± 0.1 | 3.4 ± 0.1 | **8.47E-14*** ** | 0.0018 ± 7E-04 | 0.0025 ± 7E-04 | **5.68E-12*** ** |
| **EUR** | 6391.1 ± 74.5 | 7053.2 ± 126.2 | **6.29E-06** | 1907.7 ± 6.8 | 1936.8 ± 8.9 | **0.009**** | 3 ± 0 | 3.2 ± 0 | **3.1E-04*** ** | 0.0022 ± 3E-04 | 0.0024 ± 4E-04 | **6.3E-06*** ** |
| **MDE** | 26001.8 ± 2808 | 11143.3 ± 3046 | **6.67E-04** | 2048.5 ± 50.3 | 1814.9 ± 206.5 | 0.284 | 10.4 ± 1 | 4.6 ± 1 | **6.8E-05*** ** | 0.009 ± 0.001 | 0.0039 ± 0.001 | **6.67E-04*** ** |
| **SAS** | 12647.7 ± 2021.5 | 34198.7 ± 4113.8 | **4.08E-06** | 1995.5 ± 62.9 | 2155 ± 66 | 0.081 | 5.3 ± 0.7 | 11.8 ± 1.3 | **1.34E-05*** ** | 0.0044 ± 7E-04 | 0.0118 ± 0.001 | **4.08E-06*** ** |

Legend: AAC = African Admixed, AFR = African, AJ = Ashkenazi Jewish, AMR = American Admixed, $AV_{ROH}$ = average length of runs of homozygosity, CAS = Central Asian, EAS = East Asian, EUR = European, $F_{ROH}$ = runs of homozygosity-based estimates for inbreeding coefficient, MDE = Middle-Eastern, $N_{ROH}$ = number of runs of homozygosity, SAS = South Asian, SE = standard error, $S_{ROH}$ = total length of runs of homozygosity. *p*-values indicate the statistical significance as following $p < 0.001$:***, $p < 0.01$:**, $p < 0.05$:*, $p \geq 0.05$: not significant

***Supplementary Table 8:*** *Measures of genome-wide homozygosity in Parkinson's disease excluding Parkinson's disease recessive genes (PRKN, PINK1, and DJ-1)*

| Ancestry | SROH | | AVROH | | NROH | | FROH | |
|---|---|---|---|---|---|---|---|---|
| | **Beta (mean ± SE)** | **p-value** | **Beta (mean ± SE)** | **p-value** | **Beta (mean ± SE)** | **p-value** | **Beta (mean ± SE)** | **p-value** |
| **AAC** | 1.31E-05 ± 6.16E-06 | **0.034** | 1.76E-04 ± 6.23E-05 | **0.005** | 0.048 ± 0.027 | 0.077 | 37.8 ± 17.8 | **0.034** |
| **AFR** | 7.13E-06 ± 3.45E-06 | **0.039** | 1.19E-04 ± 5.1E-05 | **0.019** | 0.022 ± 0.012 | 0.056 | 20.6 ± 9.97 | **0.039** |
| **AJ** | 2.02E-06 ± 2.66E-06 | 0.448 | 6.51E-05 ± 6.77E-05 | 0.336 | 0.003 ± 0.013 | 0.828 | 5.82 ± 7.67 | 0.448 |
| **AMR** | 4.34E-06 ± 4.61E-06 | 0.347 | -8.66E-05 ± 7.64E-05 | 0.257 | 0.0231 ± 0.022 | 0.300 | 12.5 ± 13.3 | 0.347 |
| **CAS** | -2.69E-07 ± 2.87E-06 | 0.925 | -1.56E-04 ± 1.21E-04 | 0.196 | -9.25E-04 ± 0.010 | 0.927 | -0.775 ± 8.27 | 0.925 |
| **EAS** | -9.94E-06 ± 5.41E-06 | 0.066 | -4.94E-05 ± 6.26E-05 | 0.430 | -0.026 ± 0.014 | 0.059 | -28.7 ± 15.6 | 0.066 |
| **EUR** | -8.48E-06 ± 1.68E-06 | **4.51E-07** | -6.01E-05 ± 2E-05 | **0.003** | -0.023 ± 0.005 | **1.89E-05** | -24.5 ± 4.85 | **4.52E-07** |
| **MDE** | 4.04E-05 ± 2.1E-05 | 0.054 | 4.54E-04 ± 3.02E-04 | 0.133 | 0.115 ± 0.056 | **0.042** | 117 ± 60.6 | 0.054 |
| **SAS** | -1.67E-05 ± 4.83E-06 | **5.44E-04** | -2.61E-04 ± 1.52E-04 | 0.086 | -0.045 ± 0.013 | **7.81E-04** | -48.2 ± 13.9 | **5.44E-04** |

Legend: AAC = African Admixed, AFR = African, AJ = Ashkenazi Jewish, AMR = American Admixed, $AV_{ROH}$ = average length of runs of homozygosity, CAS = Central Asian, EAS = East Asian, EUR = European, $F_{ROH}$ = runs of homozygosity-based estimates for inbreeding coefficient, MDE = Middle-Eastern, $N_{ROH}$ = number of runs of homozygosity, SAS = South Asian, SE = standard error, $S_{ROH}$ = total length of runs of homozygosity. *p*-values indicate the statistical significance as following $p < 0.001$:***, $p < 0.01$:**, $p < 0.05$:*, $p \geq 0.05$: not significant

***Supplementary Table 9****: Linear regression for runs of homozygosity associated with Parkinson's disease at age of onset*

| Ancestry | $S_{ROH}$ | | | $AV_{ROH}$ | | | $N_{ROH}$ | | | $F_{ROH}$ | | |
|---|---|---|---|---|---|---|---|---|---|---|---|---|
| | **Cases (mean ± SE)** | **beta** | **p-value** | **Cases (mean ± SE)** | **beta** | **p-value** | **Cases (mean ± SE)** | **beta** | **p-value** | **Cases (mean ± SE)** | **beta** | **p-value** |
| **AAC** | 6330.5 ± 989.3 | 7.86E-06 | 0.793 | 1815.6 ± 84.6 | -7.91E-04 | **0.035*** | 2.1 ± 0.2 | -0.0131 | 0.939 | 0.0022 ± 0.00034 | 22.7 | 0.792 |
| **AFR** | 9820.3 ± 456.3 | 1.55E-05 | 0.426 | 2252.6 ± 28.4 | -2.55E-04 | 0.642 | 4 ± 0.1 | 0.0266 | 0.778 | 0.0034 ± 0.00016 | 44.8 | 0.425 |
| **AJ** | 22595 ± 816.2 | -4.37E-06 | 0.631 | 2879.5 ± 27.5 | 4.79E-04 | 0.082 | 7.3 ± 0.2 | -0.0549 | 0.281 | 0.0078 ± 0.00028 | -12.6 | 0.631 |
| **AMR** | 15162.8 ± 1491.6 | -5.51E-05 | **3.87E-4*** ** | 2302.8 ± 64.8 | -8.78E-04 | **0.019*** | 5.1 ± 0.3 | -0.235 | **0.003*** | 0.0053 ± 0.00052 | -159.0 | **3.73E-4*** ** |
| **CAS** | 13658.5 ± 2093.5 | -2.80E-05 | 0.554 | 1885 ± 51.6 | 2.30E-04 | 0.767 | 5.2 ± 0.6 | -0.0698 | 0.656 | 0.0047 ± 0.00073 | -80.9 | 0.554 |
| **EAS** | 5188.6 ± 201.3 | -2.14E-05 | 0.607 | 1735.7 ± 20.4 | 3.03E-04 | 0.406 | 2.5 ± 0.1 | -0.0382 | 0.716 | 0.0018 ± 7e-05 | -61.7 | 0.608 |
| **EUR** | 6391.1 ± 74.5 | -2.04E-05 | **0.027*** | 1907.7 ± 6.8 | -6.23E-05 | 0.569 | 3 ± 0 | -0.0499 | 0.094 | 0.0022 ± 2.6e-05 | -58.9 | **0.027*** |
| **MDE** | 26001.8 ± 2808 | -6.00E-06 | 0.609 | 2048.5 ± 50.3 | 7.68E-04 | 0.217 | 10.4 ± 1 | -0.0290 | 0.390 | 0.009 ± 0.00097 | -17.3 | 0.609 |
| **SAS** | 12647.7 ± 2021.5 | -7.17E-05 | 0.282 | 1995.5 ± 62.9 | 4.47E-04 | 0.801 | 5.3 ± 0.7 | -0.251 | 0.174 | 0.0044 ± 7e-04 | -207.0 | 0.282 |

Legend: AAC = African Admixed, AFR = African, AJ = Ashkenazi Jewish, AMR = American Admixed, $AV_{ROH}$ = average length of runs of homozygosity, CAS = Central Asian, EAS = East Asian, EUR = European, $F_{ROH}$ = runs of homozygosity-based estimates for inbreeding coefficient, MDE = Middle-Eastern, $N_{ROH}$ = number of runs of homozygosity, SAS = South Asian, SE = standard error, $S_{ROH}$ = total length of runs of homozygosity. p-values indicate the statistical significance as following p < 0.001:***, p < 0.01:**, p < 0.05:*, p ≥ 0.05: not significant

***Supplementary Table 10***: *Runs of homozygosity associated with Parkinson's disease at age of onset excluding Parkinson's disease recessive genes (PRKN, PINK1, and DJ-1)*

| Ancestry | $S_{ROH}$ | | | $AV_{ROH}$ | | | $N_{ROH}$ | | | $F_{ROH}$ | | |
|---|---|---|---|---|---|---|---|---|---|---|---|---|
| | **Cases (mean ± SE)** | **beta** | **p-value** | **Cases (mean ± SE)** | **beta** | **p-value** | **Cases (mean ± SE)** | **beta** | **p-value** | **Cases (mean ± SE)** | **beta** | **p-value** |
| **AAC** | 6330.5 ± 989.3 | 7.86E-06 | 0.793 | 1815.6 ± 84.6 | -7.91E-04 | **0.036*** | 2.1 ± 0.2 | -0.0131 | 0.939 | 0.0022 ± 0.00034 | 22.7 | 0.792 |
| **AFR** | 9814.6 ± 455.3 | 1.54E-05 | 0.430 | 2252.7 ± 28.4 | -2.54E-04 | 0.644 | 4 ± 0.1 | 0.0252 | 0.789 | 0.0034 ± 0.00016 | 44.6 | 0.429 |
| **AJ** | 22579.8 ± 815.6 | -4.41E-06 | 0.628 | 2878.5 ± 27.5 | 4.76E-04 | 0.084 | 7.3 ± 0.2 | -0.0549 | 0.280 | 0.0078 ± 0.00028 | -12.8 | 0.628 |
| **AMR** | 15143.2 ± 1486.5 | -5.54E-05 | **3.68E-4***** | 2302.8 ± 64.9 | -8.80E-04 | **0.019*** | 5.1 ± 0.3 | -0.237 | **0.003**** | 0.0052 ± 0.00051 | -160 | **3.69E-4***** |
| **CAS** | 13651.4 ± 2090.8 | -2.80E-05 | 0.554 | 1885.2 ± 51.6 | 2.30E-04 | 0.767 | 5.2 ± 0.6 | -0.0698 | 0.656 | 0.0047 ± 0.00072 | -80.9 | 0.554 |
| **EAS** | 5187.5 ± 201 | -2.08E-05 | 0.620 | 1735.7 ± 20.4 | 3.01E-04 | 0.409 | 2.5 ± 0.1 | -0.0354 | 0.737 | 0.0018 ± 7E-05 | -59.8 | 0.620 |
| **EUR** | 6389.6 ± 74.5 | -2.02E-05 | **0.029*** | 1907.8 ± 6.8 | -6.24E-05 | 0.568 | 3 ± 0 | -0.0490 | 0.101 | 0.0022 ± 2.6E-05 | -58.3 | **0.029*** |
| **MDE** | 25964.3 ± 2807.1 | -5.94E-06 | 0.612 | 2049.4 ± 50.4 | 7.72E-04 | 0.214 | 10.4 ± 1 | -0.0288 | 0.394 | 0.009 ± 0.00097 | -17.1 | 0.612 |
| **SAS** | 12647.6 ± 2021.5 | -7.16E-05 | 0.282 | 1995.5 ± 62.9 | 4.48E-04 | 0.801 | 5.3 ± 0.7 | -0.251 | 0.174 | 0.0044 ± 7E-04 | -207 | 0.252 |

Legend: AAC = African Admixed, AFR = African, AJ = Ashkenazi Jewish, AMR = American Admixed, $AV_{ROH}$ = average length of runs of homozygosity, CAS = Central Asian, EAS = East Asian, EUR = European, $F_{ROH}$ = runs of homozygosity-based estimates for inbreeding coefficient, MDE = Middle-Eastern, $N_{ROH}$ = number of runs of homozygosity, SAS = South Asian, SE = standard error, $S_{ROH}$ = total length of runs of homozygosity. p-values indicate the statistical significance as following $p < 0.001$:***, $p < 0.01$:**, $p < 0.05$:*, $p \geq 0.05$: not significant

***Supplementary Table 11***: *Runs of homozygosity in cases subsetted for different age of onset of Parkinson's disease versus controls*

| Ancestry | AAO | Number of cases (controls) | $S_{ROH}$ | | $AV_{ROH}$ | | $N_{ROH}$ | | $F_{ROH}$ | |
|---|---|---|---|---|---|---|---|---|---|---|
| | | | Beta (mean ± SE) | p-value | Beta (mean ± SE) | p-value | Beta (mean ± SE) | p-value | Beta (mean ± SE) | p-value |
| AAC | <35 | 3 (804) | 4.73E-06 ± 7.15E-05 | 0.947 | 9.94E-04 ± 3.82E-04 | **0.009**** | -0.12 ± 0.374 | 0.749 | 13.7 ± 206 | 0.947 |
| | 35-44 | 18 (804) | 2.89E-05 ± 1.71E-05 | 0.092 | 6.34E-04 ± 2.37E-04 | **0.007**** | 0.093 ± 0.077 | 0.231 | 83.3 ± 49.4 | 0.092 |
| | 45-54 | 34 (804) | -1.85E-06 ± 2.57E-05 | 0.943 | 2.66E-04 ± 1.47E-04 | 0.070 | -0.026 ± 0.096 | 0.790 | -5.4 ± 74.1 | 0.942 |
| | 55-65 | 36 (804) | -1.77E-06 ± 2.61E-05 | 0.946 | -1.3E-04 ± 1.74E-04 | 0.453 | -0.037 ± 0.099 | 0.709 | -5.21 ± 75.4 | 0.945 |
| | >65 | 58 (804) | 1.68E-05 ± 9.21E-06 | 0.069 | 2.97E-04 ± 1.32E-04 | **0.024*** | 0.082 ± 0.0491 | 0.095 | 48.4 ± 26.6 | 0.069 |
| AFR | <35 | 8 (1631) | 8.25E-06 ± 1.33E-05 | 0.534 | 2.05E-04 ± 4.22E-04 | 0.627 | 0.039 ± 0.055 | 0.479 | 23.8 ± 38.3 | 0.534 |
| | 35-44 | 13 (1631) | 3.86E-06 ± 2.91E-05 | 0.895 | 2.02E-04 ± 4.22E-04 | 0.632 | -0.023 ± 0.102 | 0.821 | 11.2 ± 83.9 | 0.894 |
| | 45-54 | 43 (1631) | -2.24E-06 ± 1.67E-05 | 0.893 | -3.63E-05 ± 2.08E-04 | 0.862 | 0.012 ± 0.042 | 0.780 | -6.43 ± 48.2 | 0.894 |
| | 55-65 | 50 (1631) | -1.65E-05 ± 2.02E-05 | 0.416 | 8.73E-05 ± 1.8E-04 | 0.627 | -0.037 ± 0.053 | 0.491 | -47.4 ± 58.3 | 0.416 |
| | >65 | 44 (1631) | -3.62E-05 ± 2.69E-05 | 0.179 | -1.14E-04 ± 1.93E-04 | 0.554 | -0.082 ± 0.069 | 0.234 | -104 ± 77.7 | 0.179 |
| AJ | <35 | 10 (458) | 7.87E-06 ± 1.45E-05 | 0.588 | 4.32E-04 ± 3.1E-04 | 0.163 | 0.019 ± 0.089 | 0.835 | 22.7 ± 41.9 | 0.587 |

| | | | | | | | | | | |
|---|---|---|---|---|---|---|---|---|---|---|
| | 35-44 | 73 (458) | 6.99E-06 ± 8.72E-06 | 0.423 | 3.17E-05 ± 2.09E-04 | 0.880 | 0.030 ± 0.041 | 0.461 | 20.2 ± 25.2 | 0.423 |
| | 45-54 | 151 (458) | 3.94E-06 ± 3.68E-06 | 0.284 | -3.13E-05 ± 1.38E-04 | 0.821 | 0.017 ± 0.022 | 0.421 | 11.4 ± 10.6 | 0.284 |
| | 55-65 | 258 (458) | -6.55E-06 ± 5.84E-06 | 0.263 | 4.63E-05 ± 9.36E-05 | 0.620 | -0.028 ± 0.023 | 0.237 | -18.9 ± 16.9 | 0.263 |
| | >65 | 310 (458) | 4.62E-06 ± 4E-06 | 0.248 | 1.26E-04 ± 9.95E-05 | 0.206 | 0.013 ± 0.020 | 0.527 | 13.3 ± 11.5 | 0.248 |
| **AMR** | <35 | 29 (139) | 2.74E-05 ± 1.65E-05 | 0.096 | 1.59E-04 ± 1.75E-04 | 0.363 | 0.177 ± 0.081 | **0.028*** | 79 ± 47.5 | 0.096 |
| | 35-44 | 62 (139) | 9.2E-06 ± 7.65E-06 | 0.229 | 3.34E-05 ± 1.45E-04 | 0.818 | 0.044 ± 0.037 | 0.230 | 26.5 ± 22.1 | 0.229 |
| | 45-54 | 69 (139) | 8.22E-06 ± 9.12E-06 | 0.367 | -3.2E-05 ± 1.23E-04 | 0.794 | 0.035 ± 0.042 | 0.400 | 23.7 ± 26.3 | 0.367 |
| | 55-65 | 53 (139) | 3.84E-06 ± 8.6E-06 | 0.655 | -1.25E-04 ± 1.66E-04 | 0.452 | 0.03 ± 0.04 | 0.453 | 11.1 ± 24.8 | 0.655 |
| | >65 | 41 (139) | 1.23E-05 ± 1.38E-05 | 0.371 | 2.17E-04 ± 1.6E-04 | 0.176 | 0.065 ± 0.058 | 0.269 | 35.6 ± 39.7 | 0.371 |
| **CAS** | <35 | 12 (267) | -2.12E-05 ± 4.26E-05 | 0.619 | 2.53E-04 ± 5.49E-04 | 0.645 | -0.070 ± 0.131 | 0.591 | -61.1 ± 123 | 0.619 |
| | 35-44 | 41 (267) | -3.09E-06 ± 7.69E-06 | 0.688 | -3.72E-04 ± 2.26E-04 | 0.099 | -0.010 ± 0.027 | 0.719 | -8.9 ± 22.2 | 0.688 |
| | 45-54 | 23 (267) | -1.38E-05 ± 2.06E-05 | 0.504 | 2.45E-04 ± 3.28E-04 | 0.456 | -0.046 ± 0.063 | 0.467 | -39.8 ± 59.6 | 0.504 |
| | 55-65 | 24 (267) | -1.34E-04 ± 6.25E-05 | **0.032*** | -3.93E-04 ± 3.58E-04 | 0.273 | -0.287 ± 0.128 | **0.024*** | -387 ± 180 | **0.032*** |
| | >65 | 10 (267) | -0.00446 ± 0.726 | 0.995 | -1.55E-04 ± 1.69E-04 | 0.361 | -11.1 ± 1760 | 0.995 | -12900 ± 2100000 | 0.995 |

| | | | | | | | | | | |
|---|---|---|---|---|---|---|---|---|---|---|
| | <35 | 59 (2302) | 2.74E-05 ± 1.84E-05 | 0.136 | 1.48E-05 ± 2.55E-04 | 0.954 | 0.067 ± 0.044 | 0.123 | 79.1 ± 53.1 | 0.136 |
| | 35-44 | 184 (2302) | -4.58E-06 ± 1.45E-05 | 0.753 | 5.47E-05 ± 1.75E-04 | 0.754 | -0.009 ± 0.037 | 0.807 | -13.2 ± 42 | 0.754 |
| | 45-54 | 237 (2302) | -4.18E-05 ± 2.51E-05 | 0.096 | -9.98E-05 ± 1.47E-04 | 0.497 | -0.086 ± 0.054 | 0.115 | -121 ± 72.6 | 0.096 |
| | 55-65 | 112 (2302) | -2.59E-05 ± 2.38E-05 | 0.276 | -2.73E-04 ± 1.66E-04 | 0.100 | -0.043 ± 0.050 | 0.383 | -74.9 ± 68.8 | 0.276 |
| **EAS** | >65 | 106 (2302) | -3.47E-06 ± 1.58E-05 | 0.826 | -5.62E-06 ± 2.15E-04 | 0.979 | -0.021 ± 0.041 | 0.613 | -9.98 ± 45.6 | 0.827 |
| | <35 | 244 (7505) | 8.4E-06 ± 4.3E-06 | 0.051 | -4.9E-05 ± 9.77E-05 | 0.616 | 0.027 ± 0.016 | 0.091 | 24.2 ± 12.4 | 0.051 |
| | 35-44 | 798 (7505) | -1.15E-05 ± 5.74E-06 | **0.044*** | -1.26E-04 ± 5.59E-05 | **0.025*** | -0.039 ± 0.018 | **0.027*** | -33.3 ± 16.6 | **0.045*** |
| | 45-54 | 1645 (7505) | -1.18E-05 ± 4.12E-06 | **0.004**** | -8.88E-05 ± 3.79E-05 | **0.019*** | -0.025 ± 0.011 | **0.024*** | -33.9 ± 11.9 | **0.004**** |
| | 55-65 | 2360 (7505) | -1.07E-05 ± 3.18E-06 | **7.49E-04***** | -5.89E-05 ± 3.21E-05 | 0.066 | -0.023 ± 0.009 | **0.011*** | -30.9 ± 9.18 | **7.5E-04***** |
| **EUR** | >65 | 2395 (7505) | -5.64E-06 ± 2.95E-06 | 0.056 | -1.57E-05 ± 3.74E-05 | 0.675 | -0.022 ± 0.010 | **0.031*** | -16.3 ± 8.52 | 0.056 |
| | <35 | 11 (20) | 6.51E-04 ± 2.08 | 1 | 0.01 ± 328 | 1 | 1.59 ± 5160 | 1 | 1880 ± 6010000 | 1 |
| | 35-44 | 20 (20) | 3.53E-05 ± 2.75E-05 | 0.2 | -3.42E-04 ± 7.88E-04 | 0.664 | 0.098 ± 0.077 | 0.202 | 102 ± 79.4 | 0.200 |
| | 45-54 | 40 (20) | 3.66E-05 ± 2.06E-05 | 0.076 | 1.84E-05 ± 4.12E-04 | 0.964 | 0.104 ± 0.060 | 0.082 | 106 ± 59.6 | 0.076 |
| **MDE** | 55-65 | 46 (20) | 2.88E-05 ± 2.2E-05 | 0.191 | 2.06E-04 ± 5.15E-04 | 0.689 | 0.088 ± 0.064 | 0.169 | 83.1 ± 63.5 | 0.191 |

| | | | | | | | | | | |
|---|---|---|---|---|---|---|---|---|---|---|
| | >65 | 42 (20) | 1.44E-05 ± 3.02E-05 | 0.632 | 6.13E-04 ± 4.62E-04 | 0.184 | 0.065 ± 0.092 | 0.484 | 41.7 ± 87.1 | 0.632 |
| | <35 | 4 (198) | -6.77E-06 ± 1.99E-05 | 0.734 | -5.83E-04 ± 7.13E-04 | 0.413 | -0.007 ± 0.048 | 0.892 | -19.5 ± 57.4 | 0.734 |
| | 35-44 | 18 (198) | -2.79E-05 ± 1.61E-05 | 0.084 | -1.15E-04 ± 3.28E-04 | 0.726 | -0.064 ± 0.038 | 0.090 | -80.4 ± 46.6 | 0.084 |
| | 45-54 | 11 (198) | -6.37E-06 ± 8.12E-06 | 0.433 | -3.03E-04 ± 3.73E-04 | 0.416 | -0.016 ± 0.024 | 0.499 | -18.4 ± 23.4 | 0.433 |
| | 55-65 | 12 (198) | -2.15E-05 ± 1.67E-05 | 0.199 | 2.89E-04 ± 4.04E-04 | 0.474 | -0.074 ± 0.055 | 0.182 | -62.1 ± 48.3 | 0.199 |
| **SAS** | >65 | 17 (198) | -3.23E-05 ± 2.29E-05 | 0.158 | -2.27E-04 ± 4.99E-04 | 0.649 | -0.093 ± 0.066 | 0.158 | -93.3 ± 66.1 | 0.158 |

Legend: AAC = African Admixed, AFR = African, AJ = Ashkenazi Jewish, AMR = American Admixed, $AV_{ROH}$ = average length of runs of homozygosity, CAS = Central Asian, EAS = East Asian, EUR = European, $F_{ROH}$ = runs of homozygosity-based estimates for inbreeding coefficient, MDE = Middle-Eastern, $N_{ROH}$ = number of runs of homozygosity, SAS = South Asian, SE = standard error, $S_{ROH}$ = total length of runs of homozygosity. p-values indicate the statistical significance as following $p < 0.001$:***, $p < 0.01$:**, $p < 0.05$:*, $p \geq 0.05$: not significant

***Supplementary Table 12:*** *T-test results for runs of homozygosity in cases subsetted for early-onset Parkinson's disease and late-onset Parkinson's disease versus controls*

| Ancestry | AAO | Number of cases (controls) | $S_{ROH}$ | | | $AV_{ROH}$ | | | $N_{ROH}$ | | | $F_{ROH}$ | | |
|---|---|---|---|---|---|---|---|---|---|---|---|---|---|---|
| | | | **Cases (mean ± SE)** | **Controls (mean ± SE)** | **p-value** | **Cases (mean ± SE)** | **Controls (mean ± SE)** | **p-value** | **Cases (mean ± SE)** | **Controls (mean ± SE)** | **p-value** | **Cases (mean ± SE)** | **Controls (mean ± SE)** | **p-value** |
| **AAC** | EOPD | 21 (804) | 8245.1 ± 3440.3 | 4373.8 ± 378.8 | 0.276 | 2240.1 ± 305.9 | 1580.9 ± 40.8 | **0.045*** | 2.6 ± 0.9 | 1.7 ± 0.1 | 0.332 | 0.003 ± 0.0012 | 0.002 ± 0.00013 | 0.276 |
| | LOPD | 128 (804) | 5091.8 ± 1457.4 | 4373.8 ± 378.8 | 0.634 | 1707.4 ± 115.3 | 1580.9 ± 40.8 | 0.302 | 1.8 ± 0.2 | 1.7 ± 0.1 | 0.933 | 0.002 ± 5E-04 | 0.002 ± 0.00013 | 0.634 |
| **AFR** | EOPD | 21 (1,631) | 25590.9 ± 12908.9 | 8158.4 ± 297.7 | 0.192 | 2510.2 ± 176 | 2118.8 ± 23.4 | **0.039*** | 7.3 ± 2.4 | 3.3 ± 0.1 | 0.109 | 0.009 ± 0.0045 | 0.003 ± 1E-04 | 0.192 |
| | LOPD | 137 (1,631) | 11624 ± 738.3 | 8158.4 ± 297.7 | **2.22E-05** | 2311.9 ± 70.1 | 2118.8 ± 23.4 | **9.82E-03**** | 4.7 ± 0.3 | 3.3 ± 0.1 | **1.82E-06***** | 0.004 ± 0.00026 | 0.003 ± 1E-04 | **2.21E-05***** |
| **AJ** | EOPD | 83 (458) | 20847.5 ± 1953.7 | 21501.1 ± 820 | 0.758 | 2854.1 ± 83.2 | 2826.3 ± 39.7 | 0.763 | 7 ± 0.5 | 7.2 ± 0.2 | 0.678 | 0.007 ± 0.00068 | 0.007 ± 0.00028 | 0.759 |
| | LOPD | 719 (458) | 22976.1 ± 1020.6 | 21501.1 ± 820 | 0.260 | 2901.1 ± 32.9 | 2826.3 ± 39.7 | 0.147 | 7.4 ± 0.2 | 7.2 ± 0.2 | 0.611 | 0.008 ± 0.00035 | 0.007 ± 0.00028 | 0.260 |
| **AMR** | EOPD | 91 (139) | 19577 ± 4274.6 | 13186.3 ± 1351 | 0.157 | 2388.5 ± 156.1 | 2483.4 ± 128.7 | 0.640 | 6 ± 0.7 | 4.6 ± 0.3 | 0.083 | 0.007 ± 0.0015 | 0.005 ± 0.00047 | 0.157 |
| | LOPD | 163 (139) | 13160.7 ± 1646.1 | 13186.3 ± 1351 | 0.990 | 2308.9 ± 91.3 | 2483.4 ± 128.7 | 0.270 | 4.8 ± 0.4 | 4.6 ± 0.3 | 0.743 | 0.005 ± 0.00057 | 0.005 ± 0.00047 | 0.991 |
| **CAS** | EOPD | 53 (267) | 9192.6 ± 2856.3 | 19719.4 ± 2506.5 | **6.33E-03** | 1567 ± 131.4 | 2087.3 ± 52.6 | **4.62E-04***** | 3.7 ± 0.8 | 6.8 ± 0.7 | **0.004**** | 0.003 ± 0.00099 | 0.007 ± 0.00087 | **0.006**** |
| | LOPD | 57 (267) | 5803.5 ± 898.6 | 19719.4 ± 2506.5 | **3.16E-07** | 1834.3 ± 92 | 2087.3 ± 52.6 | 0.019* | 2.8 ± 0.4 | 6.8 ± 0.7 | **4.49E-07***** | 0.002 ± 0.00031 | 0.007 ± 0.00087 | **3.16E-07***** |
| **EAS** | EOPD | 243 (2,302) | 5377.1 ± 518.9 | 7134.8 ± 197 | **1.69E-03** | 1777.1 ± 40.8 | 1854 ± 15 | 0.078 | 2.6 ± 0.2 | 3.4 ± 0.1 | **9.38E-04***** | 0.002 ± 0.00018 | 0.003 ± 6.8E-05 | **0.002**** |

| | | | | | | | | | | | | | | |
|---|---|---|---|---|---|---|---|---|---|---|---|---|---|---|
| | LOPD | 455 (2,302) | 4759.6 ± 284.7 | 7134.8 ± 197 | **1.24E-11** | 1699 ± 37.4 | 1854 ± 15 | **1.31E-04*** ** | 2.3 ± 0.1 | 3.4 ± 0.1 | **4.62E-12*** ** | 0.002 ± 9.9E-05 | 0.003 ± 6.8E-05 | **1.23E-11*** ** |
| **EUR** | EOPD | 1,042 (7,505) | 6727.8 ± 371.5 | 7053.2 ± 126.2 | 4.07E-01 | 1893.3 ± 23.3 | 1936.8 ± 8.9 | 0.081 | 3 ± 0.1 | 3.2 ± 0 | 0.202 | 0.002 ± 0.00013 | 0.002 ± 4.4E-05 | 0.407 |
| | LOPD | 6,400 (7,505) | 6440.6 ± 101.7 | 7053.2 ± 126.2 | **1.58E-04** | 1914.9 ± 9.3 | 1936.8 ± 8.9 | 0.088 | 3 ± 0 | 3.2 ± 0 | **0.002** ** | 0.002 ± 3.5E-05 | 0.002 ± 4.4E-05 | **1.58E-04*** ** |
| **MDE** | EOPD | 31 (20) | 54008 ± 11918.8 | 11143.3 ± 3046 | **1.39E-03** | 2251.8 ± 106.6 | 1814.9 ± 206.5 | 0.070 | 20.8 ± 4 | 4.6 ± 1 | **3.63E-04*** ** | 0.019 ± 0.0041 | 0.004 ± 0.0011 | **0.001** ** |
| | LOPD | 128 (20) | 23094.8 ± 3162 | 11143.3 ± 3046 | **8.18E-03** | 2007.9 ± 72.2 | 1814.9 ± 206.5 | 0.386 | 9.2 ± 1.1 | 4.6 ± 1 | **0.002** ** | 0.008 ± 0.0011 | 0.004 ± 0.0011 | **0.008** ** |
| **SAS** | EOPD | 22 (198) | 11189.9 ± 12058.8 | 34198.7 ± 4113.8 | **4.70E-06** | 1996.3 ± 57.8 | 2155 ± 66 | 0.074 | 5.3 ± 1.1 | 11.8 ± 1.3 | **2.2E-04*** ** | 0.004 ± 0.00089 | 0.0118 ± 0.0014 | **4.71E-06*** ** |
| | LOPD | 40 (198) | 15590.6 ± 29727.7 | 34198.7 ± 4113.8 | **3.57E-03** | 2160.3 ± 96.1 | 2155 ± 66 | 0.964 | 6.1 ± 1.5 | 11.8 ± 1.3 | **0.006** ** | 0.0054 ± 0.0016 | 0.0118 ± 0.0014 | **0.004** ** |

Legend: AAC = African Admixed, AFR = African, AJ = Ashkenazi Jewish, AMR = American Admixed, $AV_{ROH}$ = average length of runs of homozygosity, CAS = Central Asian, EAS = East Asian, EOPD = early-onset Parkinson's disease (< 45 years), EUR = European, $F_{ROH}$ = runs of homozygosity based estimates for inbreeding coefficient, LOPD = late-onset Parkinson's disease (≥ 45 years), MDE = Middle-Eastern, $N_{ROH}$ = number of runs of homozygosity, SAS = South Asian, SE = standard error, $S_{ROH}$ = total length of runs of homozygosity. p-values indicate the statistical significance as following p < 0.001:***, p < 0.01:**, p < 0.05:*, p ≥ 0.05: not significant

***Supplementary Table 13:*** *Runs of homozygosity in cases subsetted for early-onset and late-onset Parkinson's disease versus controls excluding Parkinson's disease recessive genes (PRKN, PINK1, and DJ-1)*

| **Ancestry** | **AAO** | **Number of cases (controls)** | **$S_{ROH}$** | | **$AV_{ROH}$** | | **$N_{ROH}$** | | **$F_{ROH}$** | |
|---|---|---|---|---|---|---|---|---|---|---|
| | | | **Beta (mean ± SE)** | **p-value** | **Beta (mean ± SE)** | **p-value** | **Beta (mean ± SE)** | **p-value** | **Beta (mean ± SE)** | **p-value** |

| | | | | | | | | | | |
|---|---|---|---|---|---|---|---|---|---|---|
| **AAC** | EOPD | 21 (804) | 2.66E-05 ± 1.63E-05 | 0.103 | 6.27E-04 ± 2.05E-04 | **0.005**** | 0.081 ± 0.073 | 0.077 | 76.7 ± 47 | 0.103 |
| | LOPD | 128 (804) | 1.26E-05 ± 8.02E-06 | 0.115 | 1.69E-04 ± 9.06E-05 | **0.005**** | 0.036 ± 0.043 | 0.077 | 36.5 ± 23.2 | 0.115 |
| **AFR** | EOPD | 21 (1631) | 1.54E-05 ± 7.46E-06 | **0.039*** | 3.45E-04 ± 2.71E-04 | **0.019*** | 0.060 ± 0.028 | 0.056 | 44.5 ± 21.5 | **0.039*** |
| | LOPD | 137 (1631) | -1.35E-05 ± 1.26E-05 | 0.287 | 1.23E-06 ± 1.14E-04 | **0.019*** | -0.024 ± 0.033 | 0.056 | -38.8 ± 36.4 | 0.287 |
| **AJ** | EOPD | 83 (458) | 6.71E-06 ± 8E-06 | 0.401 | 9.6E-05 ± 1.88E-04 | 0.336 | 0.026 ± 0.039 | 0.828 | 19.4 ± 23.1 | 0.401 |
| | LOPD | 719 (458) | 2.2E-06 ± 2.73E-06 | 0.420 | 8.03E-05 ± 7.22E-05 | 0.336 | 0.004 ± 0.014 | 0.828 | 6.36 ± 7.88 | 0.420 |
| **AMR** | EOPD | 91 (139) | 1.19E-05 ± 8.01E-06 | 0.138 | 5.73E-05 ± 1.21E-04 | 0.257 | 0.067 ± 0.041 | 0.257 | 34.3 ± 23.1 | 0.138 |
| | LOPD | 163 (139) | 3.29E-06 ± 6.45E-06 | 0.610 | -4.71E-05 ± 9.28E-05 | 0.257 | 0.022 ± 0.029 | 0.257 | 9.51 ± 18.6 | 0.610 |
| **CAS** | EOPD | 53 (267) | -4.73E-06 ± 7.69E-06 | 0.539 | -3.3E-04 ± 2.11E-04 | 0.196 | -0.016 ± 0.027 | 0.927 | -13.6 ± 22.2 | 0.539 |
| | LOPD | 57 (267) | -3.34E-05 ± 2.43E-05 | 0.170 | -2.9E-05 ± 2.45E-04 | 0.196 | -0.097 ± 0.062 | 0.927 | -96.3 ± 70.1 | 0.170 |
| **EAS** | EOPD | 243 (2302) | 4.32E-06 ± 1.3E-05 | 0.739 | 3E-05 ± 1.56E-04 | 0.430 | 0.015 ± 0.031 | 0.059 | 12.5 ± 37.4 | 0.738 |
| | LOPD | 455 (2302) | -2.16E-05 ± 1.21E-05 | 0.074 | -1.96E-04 ± 1.06E-04 | 0.430 | -0.050 ± 0.028 | 0.059 | -62.3 ± 34.8 | 0.074 |
| **EUR** | EOPD | 1042 (7505) | -1.71E-06 ± 3.56E-06 | 0.631 | -9.28E-05 ± 5.12E-05 | **0.003**** | -0.009 ± 0.013 | **1.89E-05***** | -4.93 ± 10.3 | 0.631 |
| | LOPD | 6400 (7505) | -7.96E-06 ± 2.01E-06 | **7.57E-05***** | -4.43E-05 ± 2.31E-05 | **0.003**** | -0.021 ± 0.006 | **1.89E-05***** | -23 ± 5.81 | **7.58E-05***** |

| | | | | | | | | | | |
|---|---|---|---|---|---|---|---|---|---|---|
| **MDE** | EOPD | 31 (20) | 3.46E-05 ± 2.6E-05 | 0.184 | -2.72E-04 ± 7.44E-04 | 0.133 | 0.096 ± 0.074 | **0.042*** | 99.7 ± 75.1 | 0.184 |
| | LOPD | 128 (20) | 3.33E-05 ± 1.69E-05 | **0.049*** | 2.86E-04 ± 3.03E-04 | 0.133 | 0.096 ± 0.047 | **0.042*** | 96.1 ± 48.8 | **0.049*** |
| **SAS** | EOPD | 22 (198) | -2.14E-05 ± 1.25E-05 | 0.087 | -1.84E-04 ± 3.06E-04 | 0.086 | -0.048 ± 0.030 | **7.81E-04***** | -61.7 ± 36 | 0.087 |
| | LOPD | 40 (198) | -1.44E-05 ± 7.55E-06 | 0.057 | -4.44E-05 ± 2.5E-04 | 0.086 | -0.041 ± 0.023 | **7.81E-04***** | -41.5 ± 21.8 | 0.057 |

Legend: AAC = African Admixed, AFR = African, AJ = Ashkenazi Jewish, AMR = American Admixed, $AV_{ROH}$ = average length of runs of homozygosity, CAS = Central Asian, EAS = East Asian, EOPD = early-onset Parkinson's disease (< 45 years), EUR = European, $F_{ROH}$ = runs of homozygosity based-estimates for inbreeding coefficient, LOPD = late-onset Parkinson's disease (≥ 45 years), MDE = Middle-Eastern, $N_{ROH}$ = number of runs of homozygosity, SAS = South Asian, SE = standard error, $S_{ROH}$ = total length of runs of homozygosity. p-values indicate the statistical significance as following p < 0.001:***, p < 0.01:**, p < 0.05:*, p ≥ 0.05: not significant

***Supplementary Table 14:*** *T-test results for runs of homozygosity in cases early-onset Parkinson's disease versus late-onset Parkinson's disease*

| **Ancestry** | **Number of EOPD (LOPD)** | **$S_{ROH}$** | | | **$AV_{ROH}$** | | | **$N_{ROH}$** | | | **$F_{ROH}$** | | |
|---|---|---|---|---|---|---|---|---|---|---|---|---|---|
| | | **EOPD (mean ± SE)** | **LOPD (mean ± SE)** | **p-value** | **EOPD (mean ± SE)** | **LOPD (mean ± SE)** | **p-value** | **EOPD (mean ± SE)** | **LOPD (mean ± SE)** | **p-value** | **EOPD (mean ± SE)** | **LOPD (mean ± SE)** | **p-value** |
| **AAC** | 21 (128) | 8245.1 ± 3440.3 | 5091.8 ± 1457.4 | 0.406 | 2240.1 ± 305.9 | 1707.4 ± 115.3 | 0.115 | 2.6 ± 0.9 | 1.8 ± 0.2 | 0.355 | 0.0029 ± 0.0012 | 0.002 ± 5E-04 | 0.406 |
| **AFR** | 21 (137) | 25590.9 ± 12908.9 | 11624 ± 738.3 | 0.293 | 2510.2 ± 176 | 2311.9 ± 70.1 | 0.304 | 7.3 ± 2.4 | 4.7 ± 0.3 | 0.300 | 0.0089 ± 0.0045 | 0.004 ± 3E-04 | 0.293 |
| **AJ** | 83 (719) | 20847.5 ± 1953.7 | 22976.1 ± 1020.6 | 0.336 | 2854.1 ± 83.2 | 2901.1 ± 32.9 | 0.600 | 7 ± 0.5 | 7.4 ± 0.2 | 0.495 | 0.0072 ± 7E-04 | 0.008 ± 4E-04 | 0.336 |
| **AMR** | 91 (163) | 19577 ± 4274.6 | 13160.7 ± 1646.1 | 0.164 | 2388.5 ± 156.1 | 2308.9 ± 91.3 | 0.661 | 6 ± 0.7 | 4.8 ± 0.4 | 0.144 | 0.0068 ± 0.0015 | 0.005 ± 6E-04 | 0.164 |
| **CAS** | 53 (57) | 9192.6 ± 2856.3 | 5803.5 ± 898.6 | 0.262 | 1567 ± 131.4 | 1834.3 ± 92 | 0.099 | 3.7 ± 0.8 | 2.8 ± 0.4 | 0.320 | 0.0032 ± 0.001 | 0.002 ± 3E-04 | 0.262 |
| **EAS** | 243 (455) | 5377.1 ± 518.9 | 4759.6 ± 284.7 | 0.297 | 1777.1 ± 40.8 | 1699 ± 37.4 | 0.159 | 2.6 ± 0.2 | 2.3 ± 0.1 | 0.205 | 0.0019 ± 2E-04 | 0.002 ± 1E-04 | 0.297 |
| **EUR** | 1,042 (6,400) | 6727.8 ± 371.5 | 6440.6 ± 101.7 | 0.456 | 1893.3 ± 23.3 | 1914.9 ± 9.3 | 0.389 | 3 ± 0.1 | 3 ± 0 | 0.925 | 0.0023 ± 1E-04 | 0.002 ± 0 | 0.456 |
| **MDE** | 31 (128) | 54008 ± 11918.8 | 23094.8 ± 3162 | **0.017*** | 2251.8 ± 106.6 | 2007.9 ± 72.2 | 0.063 | 20.8 ± 4 | 9.2 ± 1.1 | **0.008**** | 0.0187 ± 0.0041 | 0.008 ± 0.0011 | **0.017*** |
| **SAS** | 22 (40) | 11189.9 ± 2570.9 | 15590.6 ± 4700.4 | 0.415 | 1996.3 ± 57.8 | 2160.3 ± 96.1 | 0.149 | 5.3 ± 1.1 | 6.1 ± 1.5 | 0.672 | 0.0039 ± 9E-04 | 0.0054 ± 0.0016 | 0.415 |

Legend: AAC = African Admixed, AFR = African, AJ = Ashkenazi Jewish, AMR = American Admixed, $AV_{ROH}$ = average length of runs of homozygosity, CAS = Central Asian, EAS = East Asian, EOPD = early-onset Parkinson's disease (< 45 years), EUR = European, $F_{ROH}$ = runs of homozygosity-based estimates for inbreeding coefficient, LOPD = late-onset Parkinson's disease (≥ 45 years), MDE = Middle-Eastern, $N_{ROH}$ = number of runs of homozygosity, SAS = South Asian, SE = standard error, $S_{ROH}$ = total length of runs of homozygosity. *p*-values indicate the statistical significance as following $p < 0.001$:***, $p < 0.01$:**, $p < 0.05$:*, $p \geq 0.05$: not significant

***Supplementary Table 15:*** *Runs of homozygosity only in cases early-onset versus late-onset Parkinson's disease excluding Parkinson's disease recessive genes (PRKN, PINK1, and DJ-1)*

| Ancestry | Number of EOPD (LOPD) | $S_{ROH}$ | | $AV_{ROH}$ | | $N_{ROH}$ | | $F_{ROH}$ | |
|---|---|---|---|---|---|---|---|---|---|
| | | **Beta (mean ± SE)** | **p-value** | **Beta (mean ± SE)** | **p-value** | **Beta (mean ± SE)** | **p-value** | **Beta (mean ± SE)** | **p-value** |
| **AAC** | 21 (128) | -3.88E-05 ± 1.9E-05 | **0.041*** | -8.61E-04 ± 3.6E-04 | **0.017*** | -0.296 ± 0.138 | **0.032*** | -112 ± 54.7 | **0.041*** |
| **AFR** | 21 (137) | -4.31E-04 ± 2.42E-04 | 0.075 | 1.72E-03 ± 1.06E-03 | 0.105 | -0.557 ± 0.364 | 0.127 | -1240 ± 697 | 0.075 |
| **AJ** | 83 (719) | 5.95E-07 ± 4.97E-06 | 0.905 | 9.73E-06 ± 1.85E-04 | 0.958 | -0.003 ± 0.028 | 0.909 | 1.71 ± 14.3 | 0.905 |
| **AMR** | 91 (163) | -9.41E-06 ± 5.55E-06 | 0.090 | -2.22E-05 ± 1.42E-04 | 0.875 | -0.051 ± 0.028 | 0.073 | -27.1 ± 16 | 0.090 |
| **CAS** | 53 (57) | -2.25E-05 ± 2.54E-05 | 0.376 | 5.28E-04 ± 4.47E-04 | 0.237 | -0.075 ± 0.079 | 0.346 | -64.9 ± 73.2 | 0.376 |
| **EAS** | 243 (455) | -2.52E-05 ± 1.76E-05 | 0.151 | -1.58E-04 ± 1.39E-04 | 0.257 | -0.068 ± 0.045 | 0.129 | -72.8 ± 50.8 | 0.151 |
| **EUR** | 1042 (6400) | -8.32E-06 ± 4.37E-06 | 0.057 | -7.86E-06 ± 5.92E-05 | 0.894 | -0.016 ± 0.018 | 0.266 | -24 ± 12.6 | 0.057 |
| **MDE** | 31 (128) | 1.47E-06 ± 6.7E-06 | 0.826 | 3.19E-04 ± 5.1E-04 | 0.532 | 0.006 ± 0.021 | 0.764 | 4.25 ± 19.3 | 0.826 |
| **SAS** | 22 (40) | -7.97E-06 ± 4.15E-05 | 0.848 | -2E-04 ± 9.99E-04 | 0.841 | -0.052 ± 0.105 | 0.621 | -23 ± 120 | 0.848 |

Legend: AAC = African Admixed, AFR = African, AJ = Ashkenazi Jewish, AMR = American Admixed, $AV_{ROH}$ = average length of runs of homozygosity, CAS = Central Asian, EAS = East Asian, EOPD = early-onset Parkinson's disease (< 45 years), EUR = European, $F_{ROH}$ = runs of homozygosity-based estimates for inbreeding coefficient, LOPD = late-onset Parkinson's disease (≥ 45 years), MDE = Middle-Eastern, $N_{ROH}$ = number of runs of homozygosity, SAS = South Asian, SE= standard error, $S_{ROH}$ = total length of runs of homozygosity. p-values indicate the statistical significance as following p < 0.001:***, p < 0.01:**, p < 0.05:*, p ≥ 0.05: not significant

***Supplementary Table 16:*** *Average runs of homozygosity (2Mb to 10Mb) across ancestry groups*

| | | >2Mb | >3Mb | >4Mb | >5Mb | >6Mb | >7Mb | >8Mb | >9Mb | >10Mb |
|---|---|---|---|---|---|---|---|---|---|---|
| **AAC** | **Cases (mean ± SE)** | 1392.9 ± 114.6 | 1045.8 ± 141.9 | 882.9 ± 151.5 | 772.3 ± 174.7 | 663.7 ± 176.6 | 592.7 ± 181 | 553.1 ± 181.1 | 579.0 ± 188.5 | 548.2 ± 187.6 |
| | **Controls (mean ± SE)** | 1090.2 ± 155.4 | 634.9 ± 1797.3 | 515.4 ± 67.2 | 427.7 ±66.4 | 396.2 ± 68.8 | 350.2 ± 67.6 | 279.9 ± 65.3 | 244.5 ± 64 | 220.2 ± 64.3 |
| | **p-value** | **0.016 *** | **0.005 *** | **0.009 *** | **0.017 *** | 0.078 | 0.136 | 0.120 | 0.070 | 0.059 |
| **AFR** | **Cases (mean ± SE)** | 2052.5 ± 50.2 | 1494.8 ± 71 | 988.9 ± 72.4 | 672.1 ± 68.5 | 447 ± 61.3 | 272.3 ± 52.8 | 193 ± 47.1 | 168.7 ± 45.4 | 106.4 ± 37.8 |
| | **Controls (mean ± SE)** | 1676.2 ± 40 | 1197.8 ± 50.3 | 814.5 ± 50.5 | 560.2 ± 47.5 | 380.5 ± 44.2 | 286.8 ± 42.2 | 215.2 ± 39 | 172.9 ± 36.7 | 146.6 ± 35.2 |
| | **p-value** | 0.004 | 0.105 | 0.345 | 0.405 | 0.638 | 0.859 | 0.950 | 0.712 | 0.787 |
| **AJ** | **Cases (mean ± SE)** | 3610.5 ± 44.9 | 4262.4 ± 76 | 4239.9 ± 104.7 | 3766.3 ± 125.1 | 3085.9 ± 135.3 | 2445.4 ± 137.4 | 1924.5 ± 134 | 1541.7 ± 130.2 | 1167.9 ± 121.9 |
| | **Controls (mean ± SE)** | 3486.7 ± 64.4 | 4110.4 ± 105.4 | 3916.2 ± 148.5 | 3435.8 ± 177 | 2841 ± 196.2 | 2198.3 ± 197.2 | 1702.5 ± 191 | 1434.1 ± 183.4 | 1090.3 ± 169.2 |
| | **p-value** | 0.170 | 0.328 | 0.115 | 0.178 | 0.444 | 0.476 | 0.504 | 0.853 | 0.871 |
| **AMR** | **Cases (mean ± SE)** | 2480.7 ± 116.7 | 1990.1 ± 156.1 | 1762.2 ± 174.4 | 1600.8 ± 184.9 | 1443.9 ± 193.1 | 1421.9 ± 207.4 | 1314.6 ± 211.1 | 1287.3 ± 218.9 | 1214.8 ± 219.3 |
| | **Controls (mean ± SE)** | 2827.3 ± 304 | 2584.1 ± 355.4 | 2260.6 ± 370.4 | 2240.3 ± 383.5 | 2108.6 ± 401.4 | 1892.9 ± 405.6 | 1705.9 ± 401.9 | 1509.9 ± 401 | 1349 ± 398.8 |
| | **p-value** | 0.294 | 0.117 | 0.204 | 0.130 | 0.142 | 0.358 | 0.509 | 0.819 | 0.977 |
| **CAS** | **Cases (mean ± SE)** | 1720.6 ± 98.8 | 1251.4 ± 131.3 | 1060 ± 149.2 | 862.2 ± 152.7 | 819.8 ± 156.2 | 829 ± 172.1 | 644.5 ± 161.4 | 500.8 ± 151.9 | 446.7 ± 149.1 |
| | **Controls (mean ± SE)** | 1992.7 ± 95.6 | 1487.7 ± 140.3 | 1350.5 ± 158.1 | 1273.3 ± 170 | 1168 ± 182.4 | 1104.7 ± 186.5 | 974.6 ± 188.8 | 845.7 ± 184.2 | 801.6 ± 184.2 |
| | **p-value** | 0.811 | 0.360 | 0.157 | 0.301 | 0.287 | 0.127 | 0.331 | 0.585 | 0.831 |

| | | | | | | | | | | |
|---|---|---|---|---|---|---|---|---|---|---|
| **EAS** | **Cases (mean ± SE)** | 1313.9 ± 36.3 | 583.2 ± 37.2 | 292.8 ± 31.2 | 152.6 ± 25.3 | 88 ± 20.8 | 56.5 ± 18 | 37.1 ± 15.2 | 20.3 ± 11.7 | 14.5 ± 10.3 |
| | **Controls (mean ± SE)** | 1601.4 ± 29.5 | 853.6 ± 34.4 | 451.3 ± 30.6 | 206 ± 23.8 | 109.6 ± 19.1 | 59.1 ± 15 | 33.3 ± 12 | 14.5 ± 8.5 | 6.2 ± 6.2 |
| | **p-value** | 0.084 | 0.228 | 0.185 | 0.228 | 0.409 | 0.638 | 0.720 | 0.525 | 0.938 |
| **EUR** | **Cases (mean ± SE)** | 1529.8 ± 14.5 | 788.2 ± 16.6 | 447.2 ± 15.3 | 303 ± 14.2 | 228.7 ± 13.5 | 181.7 ± 12.8 | 153 ± 12.3 | 127.4 ± 11.6 | 107 ± 11.1 |
| | **Controls (mean ± SE)** | 1610.2 ± 18.7 | 856.2 ± 21.8 | 553.1 ± 21.4 | 420.2 ± 20.9 | 342.3 ± 20.6 | 292.9 ± 20.6 | 250.4 ± 20.1 | 223.8 ± 19.7 | 191.6 ± 19.2 |
| | **p-value** | **8.5E-5 ***** | **0.0046**** | **2.5E-5 ***** | **4.00E-6 ***** | **4.00E-6 ***** | **6.00E-6 ***** | **5.6E-5 ***** | **3.8E-5 ***** | **1.57E-4 ***** |
| **MDE** | **Cases (mean ± SE)** | 2045.1 ± 95.2 | 1836.8 ± 144.5 | 1685.2 ± 176.8 | 1346.6 ± 185 | 1060.5 ± 182.4 | 655.5 ± 159.5 | 530.1 ± 150.5 | 258.1 ± 115.1 | 211.7 ± 105.8 |
| | **Controls (mean ± SE)** | 1637.4 ± 360.9 | 1670.3 ± 672.3 | 2059.4 ± 1219.3 | 1845.7 ± 1217.6 | 1570.8 ± 1205.2 | 1570.8 ± 1205.2 | 1570.8 ± 1205.2 | 1152.5 ± 1152.5 | 1152.5 ± 1152.5 |
| | **p-value** | 0.165 | 0.219 | 0.825 | 0.875 | 0.730 | 0.348 | 0.252 | 0.124 | 0.114 |
| **SAS** | **Cases (mean ± SE)** | 1865.7 ± 136 | 1336.9 ± 180.1 | 939.2 ± 189.7 | 642.7 ± 179.2 | 475.2 ± 180.8 | 380.8 ± 171.7 | 254.6 ± 148.8 | 191.3 ± 136 | 108.9 ± 108.9 |
| | **Controls (mean ± SE)** | 2148.9 ± 115.4 | 1969 ± 177.5 | 2018.5 ± 209.1 | 2023.4 ± 240.2 | 1788.3 ± 250.1 | 1778.5 ± 264 | 1657 ± 272.3 | 1499.6 ± 272.5 | 1073.8 ± 252.4 |
| | **p-value** | 0.059 | **0.013 **** | **0.00047 ***** | **8.0E-5 ***** | **3.82E-4 ***** | **5.24E-4 ***** | **9.81E-4 ***** | **0.003 **** | **0.021 *** |

Legend: AAC = African Admixed, AFR = African, AJ = Ashkenazi Jewish, AMR = American Admixed, CAS = Central Asian, EAS = East Asian, EUR = European, Mb = megabases, MDE = Middle-Eastern, SAS = South Asian, SE = standard error. *p*-values indicate the statistical significance as following $p < 0.001$:***, $p < 0.01$:**, $p < 0.05$:*, $p \geq 0.05$: not significant. The Bonferroni threshold was adjusted by dividing 0.05 by the number of ranges to account for multiple comparisons.

***Supplementary Table 17:*** *Average runs of homozygosity (2Mb to 10Mb) for early-onset and late-onset Parkinson's disease across ancestry groups*

| **Ancestry** | **AAO** | **>2Mb** | | **>3Mb** | | **>4Mb** | | **>5Mb** | | **>6Mb** | | **>7Mb** | | **>8Mb** | | **>9Mb** | | **>10Mb** | |
|---|---|---|---|---|---|---|---|---|---|---|---|---|---|---|---|---|---|---|---|
| | | **EOPD** | **LOPD** | **EOPD** | **LOPD** | **EOPD** | **LOPD** | **EOPD** | **LOPD** | **EOPD** | **LOPD** | **EOPD** | **LOPD** | **EOPD** | **LOPD** | **EOPD** | **LOPD** | **EOPD** | **LOPD** |

| | | | | | | | | | | | | | | | | | | | |
|---|---|---|---|---|---|---|---|---|---|---|---|---|---|---|---|---|---|---|---|
| **AAC** | **Number of cases (controls)** | 21 (804) | 128 (804) | 21 (804) | 128 (804) | 21 (804) | 128 (804) | 21 (804) | 128 (804) | 21 (804) | 128 (804) | 21 (804) | 128 (804) | 21 (804) | 128 (804) | 21 (804) | 128 (804) | 21 (804) | 128 (804) |
| | **Cases (mean ± SE)** | 2000.4 ± 400.3 | 1349.6 ± 160 | 1561.9 ± 523.3 | 916 ± 184.3 | 1295.2 ± 531.2 | 659.7 ± 179.8 | 960.6 ± 533.6 | 474.5 ± 173.3 | 764 ± 530.7 | 382.9 ± 176 | 776.5 ± 538.2 | 275.7 ± 160.5 | 463.9 ± 463.9 | 275.7 ± 160.5 | 546.1 ± 546.1 | 281.5 ± 164.6 | 546.1 ± 546.1 | 219.9 ± 159.6 |
| | **Controls (mean ± SE)** | 1090.2 ± 55.4 | 1090.2 ± 55.4 | 634.9 ± 63.3 | 634.9 ± 63.3 | 515.4 ± 67.2 | 515.4 ± 67.2 | 427.7 ± 66.4 | 427.7 ± 66.4 | 396.2 ± 68.8 | 396.2 ± 68.8 | 350.2 ± 67.6 | 350.2 ± 67.6 | 279.9 ± 65.3 | 279.9 ± 65.3 | 244.5 ± 64 | 244.5 ± 64 | 220.2 ± 64.3 | 220.2 ± 64.3 |
| | **p-value** | **0.005 ** ** | 0.052 | **0.003 ** ** | **0.039 * ** | **0.017 * ** | **0.040 * ** | 0.063 | 0.082 | 0.115 | 0.191 | 0.094 | 0.385 | 0.175 | 0.296 | 0.109 | 0.235 | 0.116 | 0.356 |
| **AFR** | **Number of cases (controls)** | 21 (1631) | 137 (1631) | 21 (1631) | 137 (1631) | 21 (1631) | 137 (1631) | 21 (1631) | 137 (1631) | 21 (1631) | 137 (1631) | 21 (1631) | 137 (1631) | 21 (1631) | 137 (1631) | 21 (1631) | 137 (1631) | 21 (1631) | 137 (1631) |
| | **Cases (mean ± SE)** | 2455.3 ± 345.5 | 2251.8 ± 119.4 | 2776.9 ± 548.2 | 2046.7 ± 182.9 | 2181.5 ± 650.9 | 1288.2 ± 196 | 1594.2 ± 664.7 | 824.7 ± 178.2 | 1137.1 ± 645 | 399.1 ± 137.8 | 559.6 ± 559.6 | 163.2 ± 93.5 | 620.9 ± 620.9 | 0 ± 0 | 713.3 ± 713.3 | 0 ± 0 | 713.3 ± 713.3 | 0 ± 0 |
| | **Controls (mean ± SE)** | 1676.2 ± 40 | 1676.2 ± 40 | 1197.8 ± 50.3 | 1197.8 ± 50.3 | 814.5 ± 50.5 | 814.5 ± 50.5 | 560.2 ± 47.5 | 560.2 ± 47. | 380.5 ± 44.2 | 380.5 ± 44.2 | 286.8 ± 42.2 | 286.8 ± 42.2 | 215.2 ± 39 | 215.2 ± 39 | 172.9 ± 36.7 | 172.9 ± 36.7 | 146.6 ± 35.2 | 146.6 ± 35.2 |
| | **p-value** | 0.387 | 0.905 | 0.079 | 0.515 | 0.149 | 0.115 | 0.273 | 0.052 | 0.282 | 0.080 | 0.656 | 0.051 | 0.177 | 0.978 | 0.111 | 0.981 | 0.101 | 0.976 |
| **AJ** | **Number of cases (controls)** | 83 (458) | 719 (458) | 83 (458) | 719 (458) | 83 (458) | 719 (458) | 83 (458) | 719 (458) | 83 (458) | 719 (458) | 83 (458) | 719 (458) | 83 (458) | 719 (458) | 83 (458) | 719 (458) | 83 (458) | 719 (458) |

| | | | | | | | | | | | | | | | | | | | |
|---|---|---|---|---|---|---|---|---|---|---|---|---|---|---|---|---|---|---|---|
| | **Cases (mean ± SE)** | 3591.6 ± 137.1 | 3645.2 ± 52.8 | 4204.2 ± 250.9 | 4344.7 ± 88.8 | 4171.8 ± 373.4 | 4289.7 ± 122 | 3567.1 ± 446.1 | 3816.5 ± 146.8 | 2913.4 ± 462.5 | 3082.3 ± 161.1 | 1900.1 ± 440.6 | 2463.7 ± 164.2 | 1451.1 ± 414.7 | 1907 ± 159.8 | 1365.6 ± 410.4 | 1526.7 ± 155.7 | 1268.8 ± 403.7 | 1187.6 ± 147.7 |
| | **Controls (mean ± SE)** | 3486.7 ± 64.4 | 3486.7 ± 64.4 | 4110.4 ± 105.4 | 4110.4 ± 105.4 | 3916.2 ± 148.5 | 3916.2 ± 148.5 | 3435.8 ± 177 | 3435.8 ± 177 | 2841 ± 196.2 | 2841 ± 196.2 | 2198.3 ± 197.2 | 2198.3 ± 197.2 | 1702.5 ± 191 | 1702.5 ± 191 | 1434.1 ± 183.4 | 1434.1 ± 183.4 | 1090.3 ± 169.2 | 1090.3 ± 169.2 |
| | **p-value** | 0.757 | 0.132 | 0.748 | 0.162 | 0.512 | 0.112 | 0.547 | 0.175 | 0.786 | 0.651 | 0.895 | 0.637 | 0.792 | 0.778 | 0.343 | 0.795 | 0.321 | 0.843 |
| **AMR** | **Number of cases (controls)** | 91 (139) | 163 (139) | 91 (139) | 163 (139) | 91 (139) | 163 (139) | 91 (139) | 163 (139) | 91 (139) | 163 (139) | 91 (139) | 163 (139) | 91 (139) | 163 (139) | 91 (139) | 163 (139) | 91 (139) | 163 (139) |
| | **Cases (mean ± SE)** | 2640.3 ± 259 | 2566.6 ± 176.7 | 2222.5 ± 360.5 | 1887.7 ± 226.9 | 2177.7 ± 386.3 | 1553.6 ± 255.9 | 1906.8 ± 399.3 | 1451.2 ± 275 | 1930.5 ± 417 | 1285.7 ± 287.6 | 1611.7 ± 411.4 | 1379.8 ± 330 | 1390.3 ± 429 | 1371 ± 335.5 | 1462.2 ± 478.1 | 1263.3 ± 338.3 | 1538.3 ± 500.6 | 1207.3 ± 334.8 |
| | **Controls (mean ± SE)** | 2827.3 ± 304 | 2827.3 ± 304 | 2584.1 ± 355.4 | 2584.1 ± 355.4 | 2260.6 ± 370.4 | 2260.6 ± 370.4 | 2240.3 ± 383.5 | 2240.3 ± 383.5 | 2108.6 ± 401.4 | 2108.6 ± 401.4 | 1892.9 ± 405.6 | 1892.9 ± 405.6 | 1705.9 ± 401.9 | 1705.9 ± 401.9 | 1509.9 ± 401 | 1509.9 ± 401 | 1349 ± 398.8 | 1349 ± 398.8 |
| | **p-value** | 0.626 | 0.829 | 0.764 | 0.341 | 0.437 | 0.318 | 0.524 | 0.240 | 0.266 | 0.216 | 0.528 | 0.517 | 0.537 | 0.758 | 0.377 | 0.904 | 0.324 | 0.955 |
| **CAS** | **Number of cases (controls)** | 53 (267) | 57 (267) | 53 (267) | 57 (267) | 53 (267) | 57 (267) | 53 (267) | 57 (267) | 53 (267) | 57 (267) | 53 (267) | 57 (267) | 53 (267) | 57 (267) | 53 (267) | 57 (267) | 53 (267) | 57 (267) |
| | **Cases (mean ± SE)** | 1433.7 ± 237.9 | 1356.4 ± | 749.1 ± 264.6 | 704.8 ± 193.9 | 624.3 ± 273.5 | 240.6 ± 137.2 | 596.2 ± 290.5 | 92 ± 92 | 621.4 ± 301.9 | 0 ± 0 | 688.5 ± 338.7 | 0 ± 0 | 556.1 ± 317.7 | 0 ± 0 | 412.9 ± 289.5 | 0 ± 0 | 435.4 ± 304.9 | 0 ± 0 |

| | | | | | | | | | | | | | | | | | | | |
|---|---|---|---|---|---|---|---|---|---|---|---|---|---|---|---|---|---|---|---|
| | | | 186.3 | | | | | | | | | | | | | | | | |
| | **Controls (mean ± SE)** | 1992.7 ± 95.6 | 1992.7 ± 95.6 | 1487.7 ± 140.3 | 1487.7 ± 140.3 | 1350.5 ± 158.1 | 1350.5 ± 158.1 | 1273.3 ± 170 | 1273.3 ± 170 | 1168 ± 182.4 | 1168 ± 182.4 | 1104.7 ± 186.5 | 1104.7 ± 186.5 | 974.6 ± 188.8 | 974.6 ± 188.8 | 845.7 ± 184.2 | 845.7 ± 184.2 | 801.6 ± 184.2 | 801.6 ± 184.2 |
| | **p-value** | 0.396 | 0.335 | 0.214 | 0.733 | 0.522 | 0.261 | 0.648 | 0.304 | 0.854 | 0.990 | 0.833 | 0.986 | 0.610 | 0.988 | 0.800 | 0.990 | 0.693 | 0.986 |
| **EAS** | **Number of cases (controls)** | 243 (2302) | 455 (2302) | 243 (2302) | 455 (2302) | 243 (2302) | 455 (2302) | 243 (2302) | 455 (2302) | 243 (2302) | 455 (2302) | 243 (2302) | 455 (2302) | 243 (2302) | 455 (2302) | 243 (2302) | 455 (2302) | 243 (2302) | 455 (2302) |
| | **Cases (mean ± SE)** | 1212.8 ± 85.2 | 1249.1 ± 67.9 | 421.7 ± 79.8 | 581.4 ± 68.9 | 239.9 ± 68.9 | 283.9 ± 57.6 | 77.3 ± 45.7 | 172 ± 50.1 | 38 ± 38 | 113.4 ± 43.1 | 43.1 ± 43.1 | 70.7 ± 35.5 | 43.1 ± 43.1 | 21.5 ± 21.5 | 43.1 ± 43.1 | 21.5 ± 21.5 | 47 ± 47 | 0 ± 0 |
| | **Controls (mean ± SE)** | 1601.4 ± 29.5 | 1601.4 ± 29.5 | 853.6 ± 34.4 | 853.6 ± 34.4 | 451.3 ± 30.6 | 451.3 ± 30.6 | 206 ± 23.8 | 206 ± 23.8 | 109.6 ± 19.1 | 109.6 ± 19.1 | 59.1 ± 15 | 59.1 ± 15 | 33.3 ± 12 | 33.3 ± 12 | 14.5 ± 8.5 | 14.5 ± 8.5 | 6.2 ± 6.2 | 6.2 ± 6.2 |
| | **p-value** | 0.155 | **0.033 *** | 0.581 | 0.373 | 0.721 | 0.545 | 0.648 | 0.838 | 0.525 | 0.427 | 0.848 | 0.464 | 0.861 | 0.646 | 0.862 | 0.657 | 0.569 | 0.990 |
| **EUR** | **Number of cases (controls)** | 1042 (7505) | 6400 (7505) | 1042 (7505) | 6400 (7505) | 1042 (7505) | 6400 (7505) | 1042 (7505) | 6400 (7505) | 1042 (7505) | 6400 (7505) | 1042 (7505) | 6400 (7505) | 1042 (7505) | 6400 (7505) | 1042 (7505) | 6400 (7505) | 1042 (7505) | 6400 (7505) |
| | **Cases (mean ± SE)** | 1497 ± 50.1 | 1542.6 ± 19.9 | 815.1 ± 58.9 | 789.6 ± 22.6 | 500.7 ± 56.8 | 458.9 ± 21.1 | 359.2 ± 56.1 | 315.4 ± 19.6 | 296 ± 55 | 245.8 ± 18.8 | 266.5 ± 54.6 | 200.2 ± 18 | 266.1 ± 56.3 | 166.9 ± 17.5 | 254.3 ± 56.4 | 136.2 ± 16.4 | 223.5 ± 54.8 | 116.7 ± 15.8 |
| | **Controls (mean ± SE)** | 1610.2 ± 18.7 | 1610.2 ± 18.7 | 856.2 ± 21.8 | 856.2 ± 21.8 | 553.1 ± 21.4 | 553.1 ± 21.4 | 420.2 ± 20.9 | 420.2 ± 20.9 | 342.3 ± 20.6 | 342.3 ± 20.6 | 292.9 ± 20.6 | 292.9 ± 20.6 | 250.4 ± 20.1 | 250.4 ± 20.1 | 223.8 ± 19.7 | 223.8 ± 19.7 | 191.6 ± 19.2 | 191.6 ± 19.2 |

| | | | | | | | | | | | | | | | | | | | |
|---|---|---|---|---|---|---|---|---|---|---|---|---|---|---|---|---|---|---|---|
| | **p-value** | **0.04 *** | **0.010 *** | 0.064 | **0.042 *** | 0.336 | **0.001 **** | 0.329 | **4.08E-4 ***** | 0.807 | **3.73E-4 ***** | 0.879 | **4.35E-4*****| 0.380 | **0.001 **** | 0.156 | **3.68E-4 ***** | 0.122 | **0.002 **** |
| **MDE** | **Number of cases (controls)** | 214 (20) | 214 (20) | 214 (20) | 214 (20) | 214 (20) | 214 (20) | 214 (20) | 214 (20) | 214 (20) | 214 (20) | 214 (20) | 214 (20) | 214 (20) | 214 (20) | 214 (20) | 214(20) | 214(20) | 214 (20) |
| | **Cases (mean ± SE)** | 2498.4 ± 218 | 2003.7 ± 126.9 | 2762.6 ± 381.4 | 1732 ± 187.1 | 3057.4 ± 551.5 | 1618.9 ± 221.7 | 2582.3 ± 618 | 1273.5 ± 233 | 2052.3 ± 654.2 | 999.2 ± 228.4 | 1274.4 ± 610.4 | 664.7 ± 203.8 | 1305.7 ± 628.2 | 506.6 ± 187.9 | 790.8 ± 553 | 240 ± 137.5 | 790.8 ± 553 | 162.3 ± 114.4 |
| | **Controls (mean ± SE)** | 1637.4 ± 360.9 | 1637.4 ± 360.9 | 1670.3 ± 672.3 | 1670.3 ± 672.3 | 2059.4 ± 1219.3 | 2059.4 ± 1219.3 | 1845.7 ± 1217.6 | 1845.7 ± 1217.6 | 1570.8 ± 1205.2 | 1570.8 ± 1205.2 | 1570.8 ± 1205.2 | 1570.8 ± 1205.2 | 1570.8 ± 1205.2 | 1570.8 ± 1205.2 | 1152.5 ± 1152.5 | 1152.5 ± 1152.5 | 1152.5 ± 1152.5 | 1152.5 ± 1152.5 |
| | **p-value** | 0.893 | 0.212 | 0.705 | 0.420 | 0.726 | 0.944 | 0.566 | 0.986 | 0.687 | 0.840 | 0.722 | 0.406 | 0.722 | 0.322 | 0.804 | 0.213 | 0.804 | 0.203 |
| **SAS** | **Number of cases (controls)** | 22 (198) | 40 (198) | 22 (198) | 40 (198) | 22 (198) | 40 (198) | 22 (198) | 40 (198) | 22 (198) | 40 (198) | 22 (198) | 40 (198) | 22 (198) | 40 (198) | 22 (198) | 40 (198) | 22 (198) | 40 (198) |
| | **Cases (mean ± SE)** | 2063.5 ± 251.5 | 2168.8 ± 221.9 | 981.5 ± 353 | 1680.1 ± 319.6 | 424.5 ± 293.2 | 1101.5 ± 360.4 | 0 ± 0 | 838.2 ± 361.8 | 0 ± 0 | 858.5 ± 430.9 | 0 ± 0 | 736.6 ± 424.7 | 0 ± 0 | 543.9 ± 385.5 | 0 ± 0 | 559.7 ± 394.6 | 0 ± 0 | 318.7 ± 318.7 |
| | **Controls (mean ± SE)** | 2148.9 ± 115.4 | 2148.9 ± 115.4 | 1969 ± 177.5 | 1969 ± 177.5 | 2018.5 ± 209.1 | 2018.5 ± 209.1 | 2023.4 ± 240.2 | 2023.4 ± 240.2 | 1788.3 ± 250.1 | 1788.3 ± 250.1 | 1778.5 ± 264 | 1778.5 ± 264 | 1657 ± 272.3 | 1657 ± 272.3 | 1499.6 ± 272.5 | 1499.6 ± 272.5 | 1073.8 ± 252.4 | 1073.8 ± 252.4 |
| | **p-value** | 0.737 | 0.605 | 0.117 | 0.741 | **0.030 *** | 0.081 | 0.986 | **0.024 *** | 0.980 | 0.083 | 0.980 | 0.075 | 0.980 | 0.053 | 0.982 | 0.083 | 0.991 | 0.217 |

Legend: AAC = African Admixed, AFR = African, AJ = Ashkenazi Jewish, AMR = American Admixed, CAS = Central Asian, EAS = East Asian, EOPD = early-onset PD, EUR = European, LOPD = late-onset PD, Mb = megabases, MDE = Middle-Eastern, SAS = South Asian, SE = standard error. p-values indicate the statistical significance as following $p < 0.001$: ***, $p < 0.01$:**, $p < 0.05$:*, $p \geq 0.05$: not significant. The Bonferroni threshold was adjusted by dividing 0.05 by the number of ranges to account for multiple comparisons.

***Supplementary Table 18:*** *Homozygosity mapping results for runs of homozygosity enriched in individuals with consanguinity and early-onset Parkinson's disease.*

| ROH pools enriched in individuals with consanguinity | | | | | | | | | | |
|---|---|---|---|---|---|---|---|---|---|---|
| **Ancestry** | **Cases with consanguinity (Percentage)** | **Age at onset (mean ± SD)** | **Has family history (Percentage)** | **ROHs in cases only for individuals with consanguinity** | **Total ROH overlaps** | **Number of ROH enriched in cases** | **Number of ROH enriched in cases that pass Bonferroni** | **Total number of ROH overlapping PD genes** | **Number of ROH overlapping PD genes enriched in individuals with consanguinity** | **Number of ROHs overlapping PD genes that pass Bonferroni** |
| **AAC** | 6 (2.3) | 55 ± 24 | 16.7 | 3 | 405 | 122 | 51 | 45 | 18 | 9 |
| **AFR** | 13 (1.4) | 29 ± NA | 0 | 1 | 1,162 | 536 | 128 | 125 | 68 | 19 |
| **AJ** | 49 (4.9) | 64.5 ± 10 | 26.5 | 10 | 1,217 | 1,059 | 56 | 122 | 112 | 15 |
| **AMR** | 20 (5.5) | 42.9 ± 17 | 20 | 86 | 406 | 350 | 7 | 46 | 38 | 2 |
| **CAS** | 15 (5.7) | 38 ± 4.2 | 33.3 | 17 | 861 | 575 | 66 | 93 | 68 | 13 |
| **EAS** | 10 (0.7) | 42.6 ± 20 | 10 | 2 | 1047 | 412 | 118 | 100 | 38 | 14 |
| **EUR** | 75 (0.6) | 57.1 ± 16 | 33.3 | 1 | 1,648 | 1,490 | 851 | 145 | 130 | 93 |

| | | | | | | | | | | |
|---|---|---|---|---|---|---|---|---|---|---|
| **MDE** | 42 (19.6) | 46.4 ± 13 | 40.5 | 298 | 483 | 454 | 0 | 68 | 63 | 0 |
| **SAS** | 5 (4.3) | 51.3 ± 8.4 | 20 | 2 | 587 | 276 | 6 | 67 | 30 | 2 |
| **ROH pools enriched in EOPD cases** | | | | | | | | | | |
| **Ancestry** | **EOPD Cases (Percentage)** | **Age at onset (mean ± SD)** | **Has family history (Percentage)** | **ROHs in cases only for EOPD cases** | **Total ROH overlaps** | **Number of ROH enriched in EOPD cases** | **Number of ROH that pass Bonferroni** | **Total number of ROH overlapping PD genes** | **Number of ROH overlapping PD genes enriched in EOPD cases** | **Number of ROHs overlapping PD genes that pass Bonferroni** |
| **AAC** | 21 (8.2) | 37.6 ± 4.9 | 0.485 | 0 | 374 | 53 | 0 | 40 | 3 | 0 |
| **AFR** | 21 (2.3) | 35.7 ± 8.3 | 0.242 | 0 | 1,117 | 286 | 16 | 117 | 34 | 0 |
| **AJ** | 83 (8.3) | 39.1 ± 6.1 | 7.02 | 3 | 1,091 | 486 | 0 | 107 | 54 | 0 |
| **AMR** | 91 (25.1) | 35.7 ± 7.6 | 7.39 | 60 | 345 | 228 | 0 | 42 | 33 | 0 |
| **CAS** | 53 (20) | 35.9 ± 6.3 | 1.25 | 0 | 685 | 159 | 0 | 77 | 25 | 0 |
| **EAS** | 243 (15.8) | 37.2 ± 6.9 | 1.85 | 0 | 1034 | 276 | 0 | 25 | 2 | 0 |
| **EUR** | 1042 (8.9) | 37.5 ± 6.5 | 4.47 | 3 | 1,547 | 669 | 1 | 136 | 65 | 0 |
| **MDE** | 31 (14.5) | 35.2 ± 7.9 | 17.6 | 175 | 251 | 205 | 0 | 33 | 30 | 0 |
| **SAS** | 22 (18.8) | 37 ± 4.4 | 1.82 | 0 | 713 | 104 | 0 | 84 | 13 | 0 |

Legend: AAC = African Admixed, AFR = African, AJ = Ashkenazi Jewish, AMR = American Admixed, CAS = Central Asian, EAS = East Asian, EOPD = early-onset Parkinson's disease (< 45 years), EUR = European, LOPD = late-onset Parkinson's disease (≥ 45 years), MDE = Middle-Eastern, PD = Parkinson's disease, ROHs = runs of homozygosity, SAS = South Asian. The Bonferroni threshold was adjusted by dividing 0.05 by the number of enriched ROHs to account for multiple comparisons.

***Supplementary Table 19:*** *Annotated variants only present in Parkinson's disease cases prioritized from homozygosity mapping with additional annotation information*

| Ancestry | ROH Pool | Number of cases with ROH pool | Present in multiple cases | CHR:BP:REF:ALT | rsID | Gene | Variant type | Variant class | Variant Impact | Clinical significance | REVEL score | AlphaMissense class | Eve class |
|---|---|---|---|---|---|---|---|---|---|---|---|---|---|
| AMR | S129 | 4 | Yes | 4:130964119:G:T | rs78589926 | *TIGD1* | Splice donor variant & Non-coding transcript variant | SNV | High | Not reported | Not reported | Not reported | Not reported |
| MDE | S164 and S186 | 11 and 10 | Yes | 1:20649109:C:T | rs45539432&CM052340 | *PINK1* | Stop gained | SNV | High | Pathogenic | Not reported | Not reported | Not reported |

| | | | | | | | | | | | | |
|---|---|---|---|---|---|---|---|---|---|---|---|---|
| MDE | S419 | 8 | Yes | 14:66924264:A:G | rs41285470 | *GPHN* | Missense variant | SNV | Moderate | Not reported | Not reported | Not reported | Not reported |
| MDE | S419 | 8 | Yes | 14:60118341:C:T | rs773266496&COSV58303323 | *PCNX4* | Missense variant | SNV | Moderate | Not reported | 0.130 | Likely benign | Not reported |
| MDE | S419 | 8 | Yes | 14:67577308:C:T | rs369145807 | *PLEKHH1* | Missense variant | SNV | Moderate | Not reported | 0.068 | Not reported | Not reported |
| MDE | S419 | 8 | Yes | 14:69352130:G:A | rs144512254 | *GALNT16* | Missense variant | SNV | Moderate | Not reported | 0.062 | Likely benign | Not reported |
| MDE | S419 | 8 | Yes | 14:72940346:G:A | rs145062084&COSV105912372 | *DCAF4* | Missense variant | SNV | Moderate | Likely benign | 0.017 | Likely benign | Not reported |
| MDE | S419 | 8 | Yes | 14:105863242:T:C | rs2338627 | *IGHJ6* | Missense variant | SNV | Moderate | Not reported | Not reported | Not reported | Not reported |
| MDE | S419 | 8 | Yes | 14:105863243:A:C | rs2338626 | *IGHJ6* | Missense variant | SNV | Moderate | Not reported | Not reported | Not reported | Not reported |
| AAC | S397 | 2 | No | 6:42188686:C:T | rs137853903&CM045052&COSV57835221 | *GUCA1B* | Missense | SNV | Moderate | Benign & likely benign | 0.239 | Not reported | Ambiguous |

| | | | | | | | | | | | | | |
|---|---|---|---|---|---|---|---|---|---|---|---|---|---|
| | | | | | | | variant | | | | | | |
| AAC | S397 | 2 | No | 6:42745604:C:T | rs7742995 | *TBCC* | Missense variant | SNV | Moderate | Not reported | 0.042 | Not reported | Likely benign |
| AAC | S397 | 2 | No | 6:42978919:G:A | rs1420377361 | *PEX6* | Missense variant | SNV | Moderate | Not reported | 0.388 | Not reported | Not reported |
| AAC | S396 | 2 | No | 6:42188686:C:T | rs137853903&CM045052&COSV57835221 | *GUCA1B* | Missense variant | SNV | Moderate | Benign & likely benign | 0.239 | Not reported | ambiguous |
| AJ | S1665 | 3 | No | 13:23340893:C:A | rs142967124 | *SACS* | Missense variant | SNV | Moderate | Benign & likely benign | 0.122 | Benign | Likely benign |
| AJ | S1651 | 1 | No | 11:8985795:G:A | rs61756060&COSV58469013 | *NRIP3* | Missense variant | SNV | Moderate | Not reported | 0.219 | Not reported | Likely benign |
| AJ | S1650 | 3 | No | 11:551741:C:T | rs139348192 | *LRRC56* | Missense variant | SNV | Moderate | Benign | 0.101 | Pathogenic | Likely benign |
| AJ | S1623 | 3 | No | 9:2823786:C:T | rs775393848 | *PUM3* | Missense variant | SNV | Moderate | Not reported | 0.517 | Not reported | Likely benign |

| | | | | | | | | | | | | | |
|---|---|---|---|---|---|---|---|---|---|---|---|---|---|
| AJ | S1443 | 4 | No | 8:120255342:T:C | rs141272095 | *COL14A1* | Missense variant | SNV | Moderate | Not reported | 0.246 | Not reported | Likely benign |
| AMR | S82 | 5 | No | 9:84723600:C:A | rs139913267&COSV52853311&COSV52860967 | *NTRK2* | Missense variant | SNV | Moderate | Benign/likely benign | Not reported | Not reported | Not reported |
| MDE | S606 | 1 | No | 15:78766976:C:T | rs142824118&COSV66308197 | *ADAMTS7* | Missense variant | SNV | Moderate | Not reported | 0.035 | Not reported | Likely benign |
| SAS | S892 | 2 | No | 8:128916918:T:A | rs369143754 | *CCDC26* | Splice acceptor variant & Non-coding transcript variant | SNV | High | Not reported | Not reported | Not reported | Not reported |

Legend: AAC = African Admixed, AJ = Ashkenazi Jewish, ALT = alternative allele, AMR = American Admixed, CADD = Combined Annotation Dependent Depletion, Eve class = Evolutionary model of Variant Effect, GnomAD = Genome Aggregation Database, MDE = Middle Eastern, miRNA = micro non-coding RNA, REF = reference allele, REVEL = Rare Exome Variant Ensemble Learner, ROH = runs of homozygosity, SIFT = Sorting Intolerant From Tolerant, SAS = South Asian, SNV = single nucleotide variant

***Supplementary Table 20:*** *Runs of homozygosity pools enriched in individuals with consanguinity that pass Bonferroni correction*

| Ancestry | POOL | Total participants | Total cases | Total controls | CHR | Base position 1 | Base position 2 | ROH KB | NSNP | p-value | Proportion cases | Proportion controls | Bonferroni correction |
|---|---|---|---|---|---|---|---|---|---|---|---|---|---|
| **AAC** | S51 | 10 | 2 | 8 | 16 | 47412386 | 47986415 | 574.03 | 452 | 1.21E-07 | 3.33E-01 | 9.95E-03 | 9.95E-10 |
| | S60 | 9 | 2 | 7 | 4 | 33806951 | 34461178 | 654.228 | 852 | 2.10E-08 | 3.33E-01 | 8.71E-03 | 1.73E-10 |
| | S109 | 6 | 2 | 4 | 15 | 50538139 | 51386390 | 848.252 | 2004 | 3.50E-12 | 3.33E-01 | 4.98E-03 | 2.87E-14 |
| | S110 | 6 | 2 | 4 | 15 | 51387273 | 52073368 | 686.096 | 1125 | 3.50E-12 | 3.33E-01 | 4.98E-03 | 2.87E-14 |
| | S112 | 5 | 2 | 3 | 1 | 196205159 | 197607410 | 1402.252 | 1883 | 1.95E-14 | 3.33E-01 | 3.73E-03 | 1.60E-16 |
| | S121 | 5 | 2 | 3 | 4 | 95890460 | 96650758 | 760.299 | 1379 | 1.95E-14 | 3.33E-01 | 3.73E-03 | 1.60E-16 |
| | S122 | 5 | 2 | 3 | 4 | 96671601 | 97127636 | 456.036 | 1056 | 1.95E-14 | 3.33E-01 | 3.73E-03 | 1.60E-16 |
| | S175 | 4 | 2 | 2 | 7 | 26445327 | 27398076 | 952.75 | 2004 | 8.32E-18 | 3.33E-01 | 2.49E-03 | 6.82E-20 |
| | S206 | 4 | 2 | 2 | 16 | 49010331 | 49131107 | 120.777 | 345 | 8.32E-18 | 3.33E-01 | 2.49E-03 | 6.82E-20 |
| | S223 | 3 | 2 | 1 | 1 | 197960367 | 199306182 | 1345.816 | 2558 | 2.09E-23 | 3.33E-01 | 1.24E-03 | 1.71E-25 |
| | S279 | 3 | 2 | 1 | 8 | 15150815 | 15503777 | 352.963 | 1427 | 2.09E-23 | 3.33E-01 | 1.24E-03 | 1.71E-25 |
| | S288 | 3 | 2 | 1 | 9 | 79594520 | 81396355 | 1801.836 | 4538 | 2.09E-23 | 3.33E-01 | 1.24E-03 | 1.71E-25 |
| | S293 | 3 | 2 | 1 | 11 | 6685032 | 7879993 | 1194.962 | 3614 | 2.09E-23 | 3.33E-01 | 1.24E-03 | 1.71E-25 |

| | | | | | | | | | | | | | |
|---|---|---|---|---|---|---|---|---|---|---|---|---|---|
| | S295 | 3 | 2 | 1 | 11 | 30628734 | 32256825 | 1628.092 | 2981 | 2.09E-23 | 3.33E-01 | 1.24E-03 | 1.71E-25 |
| | S312 | 3 | 2 | 1 | 15 | 52714993 | 52954752 | 239.76 | 719 | 2.09E-23 | 3.33E-01 | 1.24E-03 | 1.71E-25 |
| | S313 | 3 | 2 | 1 | 15 | 53561105 | 53700150 | 139.046 | 351 | 2.09E-23 | 3.33E-01 | 1.24E-03 | 1.71E-25 |
| | S329 | 2 | 1 | 1 | 1 | 13621474 | 15419749 | 1798.276 | 4443 | 6.18E-05 | 1.67E-01 | 1.24E-03 | 5.06E-07 |
| | S339 | 2 | 1 | 1 | 1 | 218218586 | 219625675 | 1407.09 | 2829 | 6.18E-05 | 1.67E-01 | 1.24E-03 | 5.06E-07 |
| | S342 | 2 | 1 | 1 | 1 | 237088316 | 237669270 | 580.955 | 1465 | 6.18E-05 | 1.67E-01 | 1.24E-03 | 5.06E-07 |
| | S344 | 2 | 1 | 1 | 1 | 244196043 | 244910463 | 714.421 | 1634 | 6.18E-05 | 1.67E-01 | 1.24E-03 | 5.06E-07 |
| | S359 | 2 | 1 | 1 | 2 | 190557547 | 190695863 | 138.317 | 247 | 6.18E-05 | 1.67E-01 | 1.24E-03 | 5.06E-07 |
| | S366 | 2 | 1 | 1 | 3 | 43153774 | 44854369 | 1700.596 | 2783 | 6.18E-05 | 1.67E-01 | 1.24E-03 | 5.06E-07 |
| | S367 | 2 | 1 | 1 | 3 | 45134667 | 45475606 | 340.94 | 901 | 6.18E-05 | 1.67E-01 | 1.24E-03 | 5.06E-07 |
| | S369 | 2 | 1 | 1 | 3 | 131751329 | 132925541 | 1174.213 | 2814 | 6.18E-05 | 1.67E-01 | 1.24E-03 | 5.06E-07 |
| | S373 | 2 | 1 | 1 | 4 | 47409708 | 49019082 | 1609.375 | 2903 | 6.18E-05 | 1.67E-01 | 1.24E-03 | 5.06E-07 |
| | S378 | 2 | 1 | 1 | 4 | 86568541 | 88302712 | 1734.172 | 3821 | 6.18E-05 | 1.67E-01 | 1.24E-03 | 5.06E-07 |
| | S379 | 2 | 1 | 1 | 4 | 93835243 | 94715990 | 880.748 | 1974 | 6.18E-05 | 1.67E-01 | 1.24E-03 | 5.06E-07 |

| | | | | | | | | | | | | | |
|---|---|---|---|---|---|---|---|---|---|---|---|---|---|
| | S391 | 2 | 1 | 1 | 5 | 158105137 | 159510174 | 1405.038 | 2629 | 6.18E-05 | 1.67E-01 | 1.24E-03 | 5.06E-07 |
| | S393 | 2 | 1 | 1 | 6 | 32836793 | 35038899 | 2202.107 | 4929 | 6.18E-05 | 1.67E-01 | 1.24E-03 | 5.06E-07 |
| | S394 | 2 | 1 | 1 | 6 | 37203403 | 37594772 | 391.37 | 839 | 6.18E-05 | 1.67E-01 | 1.24E-03 | 5.06E-07 |
| | S395 | 2 | 1 | 1 | 6 | 38628081 | 38730063 | 101.983 | 196 | 6.18E-05 | 1.67E-01 | 1.24E-03 | 5.06E-07 |
| | S396 | 2 | 2 | 0 | 6 | 39932281 | 40995345 | 1063.065 | 2334 | 1.44E-34 | 3.33E-01 | 0.00E+00 | 1.18E-36 |
| | S397 | 2 | 2 | 0 | 6 | 42101237 | 42301027 | 199.791 | 468 | 1.44E-34 | 3.33E-01 | 0.00E+00 | 1.18E-36 |
| | S404 | 2 | 1 | 1 | 7 | 22118464 | 24040285 | 1921.822 | 4492 | 6.18E-05 | 1.67E-01 | 1.24E-03 | 5.06E-07 |
| | S408 | 2 | 1 | 1 | 7 | 134332988 | 134763420 | 430.433 | 1166 | 6.18E-05 | 1.67E-01 | 1.24E-03 | 5.06E-07 |
| | S409 | 2 | 2 | 0 | 7 | 136440705 | 137832195 | 1391.491 | 2691 | 1.44E-34 | 3.33E-01 | 0.00E+00 | 1.18E-36 |
| | S410 | 2 | 1 | 1 | 7 | 138410394 | 141012028 | 2601.635 | 5349 | 6.18E-05 | 1.67E-01 | 1.24E-03 | 5.06E-07 |
| | S411 | 2 | 1 | 1 | 7 | 141059113 | 146879533 | 5820.421 | 11327 | 6.18E-05 | 1.67E-01 | 1.24E-03 | 5.06E-07 |
| | S413 | 2 | 1 | 1 | 7 | 147802662 | 149669205 | 1866.544 | 4506 | 6.18E-05 | 1.67E-01 | 1.24E-03 | 5.06E-07 |
| | S414 | 2 | 1 | 1 | 7 | 156160369 | 157966851 | 1806.483 | 4507 | 6.18E-05 | 1.67E-01 | 1.24E-03 | 5.06E-07 |
| | S420 | 2 | 1 | 1 | 8 | 107635268 | 109528850 | 1893.583 | 4533 | 6.18E-05 | 1.67E-01 | 1.24E-03 | 5.06E-07 |

| | | | | | | | | | | | | | |
|---|---|---|---|---|---|---|---|---|---|---|---|---|---|
| | S445 | 2 | 1 | 1 | 12 | 11899749 | 12307869 | 408.121 | 898 | 6.18E-05 | 1.67E-01 | 1.24E-03 | 5.06E-07 |
| | S447 | 2 | 1 | 1 | 12 | 24786817 | 25522117 | 735.301 | 1988 | 6.18E-05 | 1.67E-01 | 1.24E-03 | 5.06E-07 |
| | S457 | 2 | 1 | 1 | 12 | 131246290 | 133220283 | 1973.994 | 4851 | 6.18E-05 | 1.67E-01 | 1.24E-03 | 5.06E-07 |
| | S465 | 2 | 1 | 1 | 14 | 53431888 | 56631839 | 3199.952 | 7120 | 6.18E-05 | 1.67E-01 | 1.24E-03 | 5.06E-07 |
| | S466 | 2 | 1 | 1 | 14 | 56967328 | 58646712 | 1679.385 | 3254 | 6.18E-05 | 1.67E-01 | 1.24E-03 | 5.06E-07 |
| | S469 | 2 | 1 | 1 | 15 | 73638536 | 75161712 | 1523.177 | 2644 | 6.18E-05 | 1.67E-01 | 1.24E-03 | 5.06E-07 |
| | S470 | 2 | 1 | 1 | 16 | 55154208 | 57060732 | 1906.525 | 4194 | 6.18E-05 | 1.67E-01 | 1.24E-03 | 5.06E-07 |
| | S475 | 2 | 1 | 1 | 17 | 56980770 | 58066902 | 1086.133 | 2315 | 6.18E-05 | 1.67E-01 | 1.24E-03 | 5.06E-07 |
| | S481 | 2 | 1 | 1 | 18 | 54886830 | 57310636 | 2423.807 | 4444 | 6.18E-05 | 1.67E-01 | 1.24E-03 | 5.06E-07 |
| | S488 | 2 | 1 | 1 | 21 | 18694867 | 19035647 | 340.781 | 1056 | 6.18E-05 | 1.67E-01 | 1.24E-03 | 5.06E-07 |
| **AFR** | S10 | 70 | 4 | 66 | 3 | 48728819 | 48913826 | 185.008 | 105 | 4.83E-05 | 3.08E-01 | 4.05E-02 | 9.02E-08 |
| | S41 | 50 | 4 | 46 | 6 | 28180332 | 28285755 | 105.424 | 192 | 4.80E-07 | 3.08E-01 | 2.82E-02 | 8.95E-10 |
| | S44 | 49 | 5 | 44 | 6 | 61660867 | 61817292 | 156.426 | 394 | 1.65E-11 | 3.85E-01 | 2.70E-02 | 3.08E-14 |
| | S64 | 36 | 4 | 32 | 6 | 29403395 | 29516457 | 113.063 | 270 | 9.50E-10 | 3.08E-01 | 1.96E-02 | 1.77E-12 |

| | | | | | | | | | | | | | |
|---|---|---|---|---|---|---|---|---|---|---|---|---|---|
| | S67 | 35 | 5 | 30 | 6 | 29636570 | 29764813 | 128.244 | 510 | 3.74E-16 | 3.85E-01 | 1.84E-02 | 6.97E-19 |
| | S70 | 34 | 3 | 31 | 3 | 84664823 | 84780882 | 116.06 | 286 | 1.27E-05 | 2.31E-01 | 1.90E-02 | 2.37E-08 |
| | S109 | 25 | 3 | 22 | 3 | 83530934 | 83675403 | 144.47 | 330 | 1.62E-07 | 2.31E-01 | 1.35E-02 | 3.02E-10 |
| | S139 | 22 | 4 | 18 | 6 | 31415915 | 31521867 | 105.953 | 129 | 7.62E-16 | 3.08E-01 | 1.10E-02 | 1.42E-18 |
| | S150 | 21 | 3 | 18 | 6 | 63185096 | 63285572 | 100.477 | 224 | 7.15E-09 | 2.31E-01 | 1.10E-02 | 1.33E-11 |
| | S179 | 19 | 3 | 16 | 6 | 68913394 | 69240716 | 327.323 | 639 | 9.26E-10 | 2.31E-01 | 9.81E-03 | 1.73E-12 |
| | S192 | 18 | 3 | 15 | 6 | 35588032 | 35711345 | 123.314 | 177 | 2.81E-10 | 2.31E-01 | 9.20E-03 | 5.25E-13 |
| | S212 | 17 | 3 | 14 | 3 | 43856298 | 44304154 | 447.857 | 697 | 7.45E-11 | 2.31E-01 | 8.58E-03 | 1.39E-13 |
| | S213 | 17 | 3 | 14 | 3 | 52502292 | 52986528 | 484.237 | 999 | 7.45E-11 | 2.31E-01 | 8.58E-03 | 1.39E-13 |
| | S266 | 15 | 4 | 11 | 6 | 48827282 | 49150144 | 322.863 | 802 | 4.06E-23 | 3.08E-01 | 6.74E-03 | 7.58E-26 |
| | S268 | 15 | 2 | 13 | 7 | 85243049 | 85497596 | 254.548 | 609 | 5.22E-05 | 1.54E-01 | 7.97E-03 | 9.74E-08 |
| | S280 | 14 | 2 | 12 | 2 | 57240622 | 57473421 | 232.8 | 688 | 2.55E-05 | 1.54E-01 | 7.36E-03 | 4.76E-08 |
| | S281 | 14 | 2 | 12 | 2 | 60731152 | 61071377 | 340.226 | 551 | 2.55E-05 | 1.54E-01 | 7.36E-03 | 4.76E-08 |
| | S296 | 14 | 2 | 12 | 6 | 74853271 | 75035474 | 182.204 | 280 | 2.55E-05 | 1.54E-01 | 7.36E-03 | 4.76E-08 |

| | | | | | | | | | | | | | |
|---|---|---|---|---|---|---|---|---|---|---|---|---|---|
| | S314 | 13 | 2 | 11 | 2 | 57715106 | 58316108 | 601.003 | 1307 | 1.12E-05 | 1.54E-01 | 6.74E-03 | 2.09E-08 |
| | S323 | 13 | 2 | 11 | 4 | 33645388 | 33917601 | 272.214 | 563 | 1.12E-05 | 1.54E-01 | 6.74E-03 | 2.09E-08 |
| | S324 | 13 | 2 | 11 | 4 | 34022455 | 34483644 | 461.19 | 794 | 1.12E-05 | 1.54E-01 | 6.74E-03 | 2.09E-08 |
| | S336 | 13 | 2 | 11 | 6 | 77675019 | 78007178 | 332.16 | 846 | 1.12E-05 | 1.54E-01 | 6.74E-03 | 2.09E-08 |
| | S337 | 13 | 2 | 11 | 7 | 86310360 | 86552968 | 242.609 | 570 | 1.12E-05 | 1.54E-01 | 6.74E-03 | 2.09E-08 |
| | S347 | 13 | 2 | 11 | 8 | 103876393 | 104330371 | 453.979 | 942 | 1.12E-05 | 1.54E-01 | 6.74E-03 | 2.09E-08 |
| | S349 | 13 | 2 | 11 | 11 | 30494135 | 30609819 | 115.685 | 221 | 1.12E-05 | 1.54E-01 | 6.74E-03 | 2.09E-08 |
| | S379 | 12 | 2 | 10 | 6 | 70033950 | 70185820 | 151.871 | 349 | 4.30E-06 | 1.54E-01 | 6.13E-03 | 8.02E-09 |
| | S381 | 12 | 2 | 10 | 6 | 73782476 | 73933339 | 150.864 | 347 | 4.30E-06 | 1.54E-01 | 6.13E-03 | 8.02E-09 |
| | S390 | 12 | 2 | 10 | 11 | 37510967 | 37618092 | 107.126 | 296 | 4.30E-06 | 1.54E-01 | 6.13E-03 | 8.02E-09 |
| | S433 | 11 | 2 | 9 | 6 | 33310149 | 33514091 | 203.943 | 296 | 1.39E-06 | 1.54E-01 | 5.52E-03 | 2.60E-09 |
| | S434 | 11 | 3 | 8 | 6 | 50663077 | 50992603 | 329.527 | 545 | 1.70E-16 | 2.31E-01 | 4.90E-03 | 3.16E-19 |
| | S435 | 11 | 3 | 8 | 6 | 51272039 | 51682129 | 410.091 | 822 | 1.70E-16 | 2.31E-01 | 4.90E-03 | 3.16E-19 |
| | S437 | 11 | 2 | 9 | 6 | 64268326 | 64533352 | 265.027 | 796 | 1.39E-06 | 1.54E-01 | 5.52E-03 | 2.60E-09 |

| | | | | | | | | | | | | | |
|---|---|---|---|---|---|---|---|---|---|---|---|---|---|
| | S439 | 11 | 3 | 8 | 6 | 67018676 | 67198121 | 179.446 | 629 | 1.70E-16 | 2.31E-01 | 4.90E-03 | 3.16E-19 |
| | S441 | 11 | 2 | 9 | 6 | 70720341 | 70955770 | 235.43 | 328 | 1.39E-06 | 1.54E-01 | 5.52E-03 | 2.60E-09 |
| | S442 | 11 | 2 | 9 | 6 | 71029738 | 71360658 | 330.921 | 824 | 1.39E-06 | 1.54E-01 | 5.52E-03 | 2.60E-09 |
| | S443 | 11 | 2 | 9 | 6 | 71437857 | 71584215 | 146.359 | 333 | 1.39E-06 | 1.54E-01 | 5.52E-03 | 2.60E-09 |
| | S444 | 11 | 2 | 9 | 6 | 71739873 | 72255955 | 516.083 | 927 | 1.39E-06 | 1.54E-01 | 5.52E-03 | 2.60E-09 |
| | S445 | 11 | 2 | 9 | 6 | 82145707 | 82257458 | 111.752 | 172 | 1.39E-06 | 1.54E-01 | 5.52E-03 | 2.60E-09 |
| | S448 | 11 | 2 | 9 | 6 | 145205964 | 146492402 | 1286.439 | 2362 | 1.39E-06 | 1.54E-01 | 5.52E-03 | 2.60E-09 |
| | S457 | 11 | 2 | 9 | 8 | 104471752 | 104867718 | 395.967 | 887 | 1.39E-06 | 1.54E-01 | 5.52E-03 | 2.60E-09 |
| | S458 | 11 | 2 | 9 | 8 | 108860914 | 109306803 | 445.89 | 1263 | 1.39E-06 | 1.54E-01 | 5.52E-03 | 2.60E-09 |
| | S459 | 11 | 2 | 9 | 8 | 118422426 | 118958393 | 535.968 | 1528 | 1.39E-06 | 1.54E-01 | 5.52E-03 | 2.60E-09 |
| | S460 | 11 | 2 | 9 | 8 | 119391387 | 119569200 | 177.814 | 453 | 1.39E-06 | 1.54E-01 | 5.52E-03 | 2.60E-09 |
| | S461 | 11 | 2 | 9 | 9 | 34421281 | 34540917 | 119.637 | 152 | 1.39E-06 | 1.54E-01 | 5.52E-03 | 2.60E-09 |
| | S475 | 11 | 2 | 9 | 15 | 50284963 | 50761937 | 476.975 | 1168 | 1.39E-06 | 1.54E-01 | 5.52E-03 | 2.60E-09 |
| | S476 | 11 | 2 | 9 | 15 | 50763691 | 52095388 | 1331.698 | 2725 | 1.39E-06 | 1.54E-01 | 5.52E-03 | 2.60E-09 |

| | | | | | | | | | | | | | |
|---|---|---|---|---|---|---|---|---|---|---|---|---|---|
| | S492 | 10 | 2 | 8 | 2 | 50664513 | 51076430 | 411.918 | 678 | 3.61E-07 | 1.54E-01 | 4.90E-03 | 6.73E-10 |
| | S493 | 10 | 2 | 8 | 2 | 51258955 | 51806510 | 547.556 | 1650 | 3.61E-07 | 1.54E-01 | 4.90E-03 | 6.73E-10 |
| | S500 | 10 | 3 | 7 | 3 | 34205539 | 35096242 | 890.704 | 1793 | 4.33E-18 | 2.31E-01 | 4.29E-03 | 8.08E-21 |
| | S502 | 10 | 3 | 7 | 3 | 35245121 | 35519479 | 274.359 | 554 | 4.33E-18 | 2.31E-01 | 4.29E-03 | 8.08E-21 |
| | S504 | 10 | 2 | 8 | 3 | 39501530 | 40162687 | 661.158 | 1720 | 3.61E-07 | 1.54E-01 | 4.90E-03 | 6.73E-10 |
| | S505 | 10 | 2 | 8 | 3 | 40476399 | 40644230 | 167.832 | 308 | 3.61E-07 | 1.54E-01 | 4.90E-03 | 6.73E-10 |
| | S506 | 10 | 3 | 7 | 3 | 45397767 | 45525262 | 127.496 | 246 | 4.33E-18 | 2.31E-01 | 4.29E-03 | 8.08E-21 |
| | S524 | 10 | 3 | 7 | 6 | 49560470 | 49682868 | 122.399 | 278 | 4.33E-18 | 2.31E-01 | 4.29E-03 | 8.08E-21 |
| | S525 | 10 | 3 | 7 | 6 | 51822479 | 51946991 | 124.513 | 253 | 4.33E-18 | 2.31E-01 | 4.29E-03 | 8.08E-21 |
| | S526 | 10 | 3 | 7 | 6 | 66378735 | 66621187 | 242.453 | 711 | 4.33E-18 | 2.31E-01 | 4.29E-03 | 8.08E-21 |
| | S527 | 10 | 3 | 7 | 6 | 66692713 | 66847728 | 155.016 | 413 | 4.33E-18 | 2.31E-01 | 4.29E-03 | 8.08E-21 |
| | S530 | 10 | 2 | 8 | 6 | 79811557 | 79982290 | 170.734 | 460 | 3.61E-07 | 1.54E-01 | 4.90E-03 | 6.73E-10 |
| | S531 | 10 | 2 | 8 | 6 | 80223177 | 80479678 | 256.502 | 728 | 3.61E-07 | 1.54E-01 | 4.90E-03 | 6.73E-10 |
| | S532 | 10 | 2 | 8 | 6 | 81168417 | 81390898 | 222.482 | 527 | 3.61E-07 | 1.54E-01 | 4.90E-03 | 6.73E-10 |

| | | | | | | | | | | | | | |
|---|---|---|---|---|---|---|---|---|---|---|---|---|---|
| | S543 | 10 | 2 | 8 | 8 | 117783209 | 118268462 | 485.254 | 1110 | 3.61E-07 | 1.54E-01 | 4.90E-03 | 6.73E-10 |
| | S573 | 9 | 2 | 7 | 1 | 176442466 | 176780654 | 338.189 | 464 | 6.97E-08 | 1.54E-01 | 4.29E-03 | 1.30E-10 |
| | S574 | 9 | 2 | 7 | 1 | 178525293 | 178959438 | 434.146 | 958 | 6.97E-08 | 1.54E-01 | 4.29E-03 | 1.30E-10 |
| | S586 | 9 | 3 | 6 | 3 | 57018060 | 58070339 | 1052.28 | 2016 | 4.92E-20 | 2.31E-01 | 3.68E-03 | 9.17E-23 |
| | S610 | 9 | 2 | 7 | 6 | 76973954 | 77110554 | 136.601 | 330 | 6.97E-08 | 1.54E-01 | 4.29E-03 | 1.30E-10 |
| | S613 | 9 | 2 | 7 | 6 | 82537954 | 82724948 | 186.995 | 411 | 6.97E-08 | 1.54E-01 | 4.29E-03 | 1.30E-10 |
| | S616 | 9 | 2 | 7 | 6 | 98490958 | 98745037 | 254.08 | 288 | 6.97E-08 | 1.54E-01 | 4.29E-03 | 1.30E-10 |
| | S632 | 9 | 2 | 7 | 8 | 105052808 | 105493352 | 440.545 | 791 | 6.97E-08 | 1.54E-01 | 4.29E-03 | 1.30E-10 |
| | S634 | 9 | 2 | 7 | 9 | 34768208 | 34889377 | 121.17 | 275 | 6.97E-08 | 1.54E-01 | 4.29E-03 | 1.30E-10 |
| | S646 | 9 | 2 | 7 | 11 | 105048665 | 105927769 | 879.105 | 1773 | 6.97E-08 | 1.54E-01 | 4.29E-03 | 1.30E-10 |
| | S647 | 9 | 2 | 7 | 11 | 106059457 | 106368624 | 309.168 | 580 | 6.97E-08 | 1.54E-01 | 4.29E-03 | 1.30E-10 |
| | S669 | 8 | 2 | 6 | 1 | 177287748 | 177464141 | 176.394 | 323 | 8.98E-09 | 1.54E-01 | 3.68E-03 | 1.67E-11 |
| | S683 | 8 | 3 | 5 | 3 | 22888766 | 24045642 | 1156.877 | 2574 | 1.84E-22 | 2.31E-01 | 3.07E-03 | 3.42E-25 |
| | S685 | 8 | 3 | 5 | 3 | 56812041 | 56955359 | 143.319 | 296 | 1.84E-22 | 2.31E-01 | 3.07E-03 | 3.42E-25 |

| | | | | | | | | | | | | | |
|---|---|---|---|---|---|---|---|---|---|---|---|---|---|
| | S740 | 8 | 2 | 6 | 8 | 115750076 | 115939770 | 189.695 | 319 | 8.98E-09 | 1.54E-01 | 3.68E-03 | 1.67E-11 |
| | S741 | 8 | 2 | 6 | 8 | 120534340 | 120829101 | 294.762 | 692 | 8.98E-09 | 1.54E-01 | 3.68E-03 | 1.67E-11 |
| | S746 | 8 | 2 | 6 | 10 | 55234687 | 55377027 | 142.341 | 289 | 8.98E-09 | 1.54E-01 | 3.68E-03 | 1.67E-11 |
| | S765 | 8 | 2 | 6 | 13 | 64055722 | 64494100 | 438.379 | 1104 | 8.98E-09 | 1.54E-01 | 3.68E-03 | 1.67E-11 |
| | S817 | 7 | 2 | 5 | 3 | 35998558 | 36224210 | 225.653 | 637 | 6.50E-10 | 1.54E-01 | 3.07E-03 | 1.21E-12 |
| | S818 | 7 | 3 | 4 | 3 | 56397760 | 56731411 | 333.652 | 737 | 1.40E-25 | 2.31E-01 | 2.45E-03 | 2.61E-28 |
| | S827 | 7 | 2 | 5 | 4 | 59934135 | 60574908 | 640.774 | 1766 | 6.50E-10 | 1.54E-01 | 3.07E-03 | 1.21E-12 |
| | S828 | 7 | 2 | 5 | 4 | 60784754 | 61001838 | 217.085 | 519 | 6.50E-10 | 1.54E-01 | 3.07E-03 | 1.21E-12 |
| | S857 | 7 | 2 | 5 | 6 | 38809086 | 39102251 | 293.166 | 685 | 6.50E-10 | 1.54E-01 | 3.07E-03 | 1.21E-12 |
| | S858 | 7 | 2 | 5 | 6 | 39883665 | 40116932 | 233.268 | 571 | 6.50E-10 | 1.54E-01 | 3.07E-03 | 1.21E-12 |
| | S859 | 7 | 2 | 5 | 6 | 40157348 | 41326358 | 1169.011 | 2675 | 6.50E-10 | 1.54E-01 | 3.07E-03 | 1.21E-12 |
| | S874 | 7 | 2 | 5 | 7 | 82744461 | 83107490 | 363.03 | 865 | 6.50E-10 | 1.54E-01 | 3.07E-03 | 1.21E-12 |
| | S875 | 7 | 2 | 5 | 7 | 83393186 | 83632522 | 239.337 | 716 | 6.50E-10 | 1.54E-01 | 3.07E-03 | 1.21E-12 |
| | S883 | 7 | 2 | 5 | 8 | 116588023 | 117399354 | 811.332 | 1861 | 6.50E-10 | 1.54E-01 | 3.07E-03 | 1.21E-12 |

| | | | | | | | | | | | | | |
|---|---|---|---|---|---|---|---|---|---|---|---|---|---|
| | S904 | 7 | 2 | 5 | 11 | 108629089 | 109973023 | 1343.935 | 2537 | 6.50E-10 | 1.54E-01 | 3.07E-03 | 1.21E-12 |
| | S905 | 7 | 2 | 5 | 11 | 110515513 | 110640876 | 125.364 | 301 | 6.50E-10 | 1.54E-01 | 3.07E-03 | 1.21E-12 |
| | S906 | 7 | 2 | 5 | 11 | 111296046 | 112587400 | 1291.355 | 2222 | 6.50E-10 | 1.54E-01 | 3.07E-03 | 1.21E-12 |
| | S935 | 6 | 2 | 4 | 1 | 168418335 | 168719795 | 301.461 | 852 | 1.98E-11 | 1.54E-01 | 2.45E-03 | 3.70E-14 |
| | S936 | 6 | 2 | 4 | 1 | 168930219 | 169202299 | 272.081 | 705 | 1.98E-11 | 1.54E-01 | 2.45E-03 | 3.70E-14 |
| | S937 | 6 | 2 | 4 | 1 | 169225917 | 169329424 | 103.508 | 244 | 1.98E-11 | 1.54E-01 | 2.45E-03 | 3.70E-14 |
| | S968 | 6 | 2 | 4 | 3 | 37228187 | 37466256 | 238.07 | 462 | 1.98E-11 | 1.54E-01 | 2.45E-03 | 3.70E-14 |
| | S969 | 6 | 2 | 4 | 3 | 38012187 | 38353299 | 341.113 | 537 | 1.98E-11 | 1.54E-01 | 2.45E-03 | 3.70E-14 |
| | S1011 | 6 | 2 | 4 | 6 | 36751302 | 36964044 | 212.743 | 534 | 1.98E-11 | 1.54E-01 | 2.45E-03 | 3.70E-14 |
| | S1040 | 6 | 2 | 4 | 8 | 105780711 | 106128237 | 347.527 | 850 | 1.98E-11 | 1.54E-01 | 2.45E-03 | 3.70E-14 |
| | S1041 | 6 | 2 | 4 | 8 | 107345781 | 107747646 | 401.866 | 1020 | 1.98E-11 | 1.54E-01 | 2.45E-03 | 3.70E-14 |
| | S1045 | 6 | 2 | 4 | 9 | 36138335 | 36282451 | 144.117 | 322 | 1.98E-11 | 1.54E-01 | 2.45E-03 | 3.70E-14 |
| | S1073 | 6 | 2 | 4 | 11 | 107126130 | 107965328 | 839.199 | 2034 | 1.98E-11 | 1.54E-01 | 2.45E-03 | 3.70E-14 |
| | S1128 | 5 | 2 | 3 | 1 | 58060710 | 58589588 | 528.879 | 1285 | 1.52E-13 | 1.54E-01 | 1.84E-03 | 2.84E-16 |

| | | | | | | | | | | | | | |
|---|---|---|---|---|---|---|---|---|---|---|---|---|---|
| | S1143 | 5 | 2 | 3 | 2 | 47646473 | 48759335 | 1112.863 | 2080 | 1.52E-13 | 1.54E-01 | 1.84E-03 | 2.84E-16 |
| | S1144 | 5 | 2 | 3 | 2 | 49056215 | 49415607 | 359.393 | 946 | 1.52E-13 | 1.54E-01 | 1.84E-03 | 2.84E-16 |
| | S1147 | 5 | 2 | 3 | 2 | 55118173 | 55435262 | 317.09 | 760 | 1.52E-13 | 1.54E-01 | 1.84E-03 | 2.84E-16 |
| | S1159 | 5 | 2 | 3 | 3 | 54998321 | 55586021 | 587.701 | 1521 | 1.52E-13 | 1.54E-01 | 1.84E-03 | 2.84E-16 |
| | S1199 | 5 | 2 | 3 | 6 | 37144230 | 37261415 | 117.186 | 226 | 1.52E-13 | 1.54E-01 | 1.84E-03 | 2.84E-16 |
| | S1200 | 5 | 2 | 3 | 6 | 37356336 | 38349910 | 993.575 | 2186 | 1.52E-13 | 1.54E-01 | 1.84E-03 | 2.84E-16 |
| | S1202 | 5 | 2 | 3 | 6 | 86802921 | 87341282 | 538.362 | 1289 | 1.52E-13 | 1.54E-01 | 1.84E-03 | 2.84E-16 |
| | S1236 | 5 | 2 | 3 | 8 | 96157210 | 97227476 | 1070.267 | 2080 | 1.52E-13 | 1.54E-01 | 1.84E-03 | 2.84E-16 |
| | S1257 | 5 | 2 | 3 | 11 | 40341946 | 40743942 | 401.997 | 1040 | 1.52E-13 | 1.54E-01 | 1.84E-03 | 2.84E-16 |
| | S1297 | 5 | 2 | 3 | 17 | 63596386 | 64036726 | 440.341 | 978 | 1.52E-13 | 1.54E-01 | 1.84E-03 | 2.84E-16 |
| | S1322 | 4 | 2 | 2 | 1 | 112485167 | 112789878 | 304.712 | 604 | 1.05E-16 | 1.54E-01 | 1.23E-03 | 1.96E-19 |
| | S1393 | 4 | 2 | 2 | 6 | 22038571 | 22214312 | 175.742 | 399 | 1.05E-16 | 1.54E-01 | 1.23E-03 | 1.96E-19 |
| | S1395 | 4 | 2 | 2 | 6 | 87947837 | 88082697 | 134.861 | 262 | 1.05E-16 | 1.54E-01 | 1.23E-03 | 1.96E-19 |
| | S1404 | 4 | 2 | 2 | 6 | 147236947 | 147857732 | 620.786 | 1324 | 1.05E-16 | 1.54E-01 | 1.23E-03 | 1.96E-19 |

| | | | | | | | | | | | | | |
|---|---|---|---|---|---|---|---|---|---|---|---|---|---|
| | S1434 | 4 | 2 | 2 | 8 | 101488186 | 101689246 | 201.061 | 412 | 1.05E-16 | 1.54E-01 | 1.23E-03 | 1.96E-19 |
| | S1446 | 4 | 2 | 2 | 9 | 79979163 | 80353847 | 374.685 | 846 | 1.05E-16 | 1.54E-01 | 1.23E-03 | 1.96E-19 |
| | S1447 | 4 | 2 | 2 | 9 | 84384006 | 84513082 | 129.077 | 340 | 1.05E-16 | 1.54E-01 | 1.23E-03 | 1.96E-19 |
| | S1466 | 4 | 2 | 2 | 11 | 25505018 | 26000335 | 495.318 | 1393 | 1.05E-16 | 1.54E-01 | 1.23E-03 | 1.96E-19 |
| | S1467 | 4 | 2 | 2 | 11 | 26543019 | 26749689 | 206.671 | 609 | 1.05E-16 | 1.54E-01 | 1.23E-03 | 1.96E-19 |
| | S1606 | 3 | 2 | 1 | 6 | 44569600 | 46097197 | 1527.598 | 3136 | 5.87E-22 | 1.54E-01 | 6.13E-04 | 1.10E-24 |
| | S1634 | 3 | 2 | 1 | 9 | 81631892 | 82533369 | 901.478 | 1237 | 5.87E-22 | 1.54E-01 | 6.13E-04 | 1.10E-24 |
| | S1635 | 3 | 2 | 1 | 9 | 82808145 | 83095464 | 287.32 | 554 | 5.87E-22 | 1.54E-01 | 6.13E-04 | 1.10E-24 |
| | S1657 | 3 | 2 | 1 | 11 | 24352804 | 24732818 | 380.015 | 1110 | 5.87E-22 | 1.54E-01 | 6.13E-04 | 1.10E-24 |
| | S1658 | 3 | 2 | 1 | 11 | 24749917 | 24895125 | 145.209 | 484 | 5.87E-22 | 1.54E-01 | 6.13E-04 | 1.10E-24 |
| | S1693 | 3 | 2 | 1 | 15 | 53478028 | 54002900 | 524.873 | 1648 | 5.87E-22 | 1.54E-01 | 6.13E-04 | 1.10E-24 |
| | S1756 | 2 | 2 | 0 | 4 | 37930085 | 39539792 | 1609.708 | 3733 | 2.00E-32 | 1.54E-01 | 0.00E+00 | 3.73E-35 |
| **AJ** | S2 | 51 | 15 | 36 | 6 | 61517308 | 61819485 | 302.178 | 809 | 1.73E-06 | 3.06E-01 | 7.86E-02 | 1.63E-09 |
| | S34 | 22 | 9 | 13 | 12 | 38519595 | 38765639 | 246.045 | 329 | 2.57E-06 | 1.84E-01 | 2.84E-02 | 2.43E-09 |

| | | | | | | | | | | | | | |
|---|---|---|---|---|---|---|---|---|---|---|---|---|---|
| | S62 | 17 | 8 | 9 | 13 | 88118485 | 88486560 | 368.076 | 639 | 1.01E-06 | 1.63E-01 | 1.97E-02 | 9.51E-10 |
| | S74 | 16 | 8 | 8 | 13 | 87912381 | 88019280 | 106.9 | 136 | 3.08E-07 | 1.63E-01 | 1.75E-02 | 2.90E-10 |
| | S88 | 15 | 8 | 7 | 13 | 89818363 | 90125024 | 306.662 | 848 | 8.01E-08 | 1.63E-01 | 1.53E-02 | 7.56E-11 |
| | S102 | 14 | 8 | 6 | 13 | 87051055 | 87541111 | 490.057 | 808 | 1.72E-08 | 1.63E-01 | 1.31E-02 | 1.62E-11 |
| | S124 | 13 | 7 | 6 | 13 | 86174511 | 86441080 | 266.57 | 416 | 6.16E-07 | 1.43E-01 | 1.31E-02 | 5.81E-10 |
| | S131 | 12 | 6 | 6 | 2 | 62598168 | 64070030 | 1471.863 | 1777 | 1.78E-05 | 1.22E-01 | 1.31E-02 | 1.68E-08 |
| | S151 | 12 | 7 | 5 | 12 | 40624560 | 40770009 | 145.45 | 277 | 1.29E-07 | 1.43E-01 | 1.09E-02 | 1.22E-10 |
| | S152 | 12 | 8 | 4 | 12 | 40788769 | 41954393 | 1165.625 | 2219 | 3.64E-10 | 1.63E-01 | 8.73E-03 | 3.43E-13 |
| | S153 | 12 | 8 | 4 | 12 | 42529792 | 42653120 | 123.329 | 241 | 3.64E-10 | 1.63E-01 | 8.73E-03 | 3.43E-13 |
| | S170 | 11 | 6 | 5 | 3 | 57039765 | 57148468 | 108.704 | 184 | 4.71E-06 | 1.22E-01 | 1.09E-02 | 4.44E-09 |
| | S210 | 11 | 7 | 4 | 20 | 24683099 | 25045238 | 362.14 | 652 | 2.03E-08 | 1.43E-01 | 8.73E-03 | 1.92E-11 |
| | S268 | 10 | 6 | 4 | 12 | 30971682 | 31113379 | 141.698 | 292 | 9.56E-07 | 1.22E-01 | 8.73E-03 | 9.03E-10 |
| | S269 | 10 | 7 | 3 | 12 | 43524729 | 43854597 | 329.869 | 511 | 2.21E-09 | 1.43E-01 | 6.55E-03 | 2.09E-12 |
| | S270 | 10 | 7 | 3 | 12 | 43922682 | 44327091 | 404.41 | 389 | 2.21E-09 | 1.43E-01 | 6.55E-03 | 2.09E-12 |

| | | | | | | | | | | | | | |
|---|---|---|---|---|---|---|---|---|---|---|---|---|---|
| | S271 | 10 | 7 | 3 | 12 | 44649310 | 45574415 | 925.106 | 963 | 2.21E-09 | 1.43E-01 | 6.55E-03 | 2.09E-12 |
| | S272 | 10 | 6 | 4 | 12 | 45610378 | 45913265 | 302.888 | 231 | 9.56E-07 | 1.22E-01 | 8.73E-03 | 9.03E-10 |
| | S297 | 10 | 7 | 3 | 20 | 23291185 | 23613694 | 322.51 | 537 | 2.21E-09 | 1.43E-01 | 6.55E-03 | 2.09E-12 |
| | S340 | 9 | 5 | 4 | 5 | 60276522 | 61263213 | 986.692 | 1263 | 3.59E-05 | 1.02E-01 | 8.73E-03 | 3.39E-08 |
| | S372 | 9 | 5 | 4 | 8 | 77682014 | 78472684 | 790.671 | 1224 | 3.59E-05 | 1.02E-01 | 8.73E-03 | 3.39E-08 |
| | S404 | 9 | 5 | 4 | 13 | 38618291 | 38770772 | 152.482 | 190 | 3.59E-05 | 1.02E-01 | 8.73E-03 | 3.39E-08 |
| | S410 | 9 | 5 | 4 | 16 | 58499171 | 58934725 | 435.555 | 942 | 3.59E-05 | 1.02E-01 | 8.73E-03 | 3.39E-08 |
| | S411 | 9 | 5 | 4 | 16 | 59418341 | 59854704 | 436.364 | 748 | 3.59E-05 | 1.02E-01 | 8.73E-03 | 3.39E-08 |
| | S418 | 9 | 7 | 2 | 20 | 20915880 | 22336148 | 1420.269 | 1464 | 1.47E-10 | 1.43E-01 | 4.37E-03 | 1.39E-13 |
| | S434 | 8 | 6 | 2 | 2 | 20934251 | 22009115 | 1074.865 | 1125 | 1.19E-08 | 1.22E-01 | 4.37E-03 | 1.12E-11 |
| | S435 | 8 | 6 | 2 | 2 | 26666708 | 28199122 | 1532.415 | 1444 | 1.19E-08 | 1.22E-01 | 4.37E-03 | 1.12E-11 |
| | S436 | 8 | 5 | 3 | 2 | 28465602 | 28567825 | 102.224 | 210 | 6.96E-06 | 1.02E-01 | 6.55E-03 | 6.57E-09 |
| | S502 | 8 | 5 | 3 | 7 | 85938095 | 87231016 | 1292.922 | 1935 | 6.96E-06 | 1.02E-01 | 6.55E-03 | 6.57E-09 |
| | S523 | 8 | 5 | 3 | 8 | 73072293 | 73226862 | 154.57 | 281 | 6.96E-06 | 1.02E-01 | 6.55E-03 | 6.57E-09 |

| | | | | | | | | | | | | | |
|---|---|---|---|---|---|---|---|---|---|---|---|---|---|
| | S555 | 8 | 5 | 3 | 12 | 30221394 | 30789240 | 567.847 | 1299 | 6.96E-06 | 1.02E-01 | 6.55E-03 | 6.57E-09 |
| | S570 | 8 | 5 | 3 | 13 | 77460429 | 78399696 | 939.268 | 1580 | 6.96E-06 | 1.02E-01 | 6.55E-03 | 6.57E-09 |
| | S589 | 8 | 5 | 3 | 16 | 55710216 | 57282328 | 1572.113 | 2370 | 6.96E-06 | 1.02E-01 | 6.55E-03 | 6.57E-09 |
| | S597 | 8 | 5 | 3 | 20 | 17272348 | 17556261 | 283.914 | 627 | 6.96E-06 | 1.02E-01 | 6.55E-03 | 6.57E-09 |
| | S598 | 8 | 6 | 2 | 20 | 17908406 | 18918138 | 1009.733 | 2049 | 1.19E-08 | 1.22E-01 | 4.37E-03 | 1.12E-11 |
| | S601 | 8 | 5 | 3 | 21 | 44532043 | 46078324 | 1546.282 | 3176 | 6.96E-06 | 1.02E-01 | 6.55E-03 | 6.57E-09 |
| | S627 | 7 | 5 | 2 | 2 | 22461063 | 23007989 | 546.927 | 668 | 8.44E-07 | 1.02E-01 | 4.37E-03 | 7.97E-10 |
| | S784 | 7 | 5 | 2 | 11 | 107182466 | 107691854 | 509.389 | 896 | 8.44E-07 | 1.02E-01 | 4.37E-03 | 7.97E-10 |
| | S785 | 7 | 5 | 2 | 11 | 107994281 | 108606766 | 612.486 | 691 | 8.44E-07 | 1.02E-01 | 4.37E-03 | 7.97E-10 |
| | S823 | 7 | 5 | 2 | 20 | 19128902 | 19634799 | 505.898 | 713 | 8.44E-07 | 1.02E-01 | 4.37E-03 | 7.97E-10 |
| | S825 | 7 | 5 | 2 | 20 | 38320055 | 39059954 | 739.9 | 953 | 8.44E-07 | 1.02E-01 | 4.37E-03 | 7.97E-10 |
| | S832 | 7 | 5 | 2 | 21 | 43194143 | 44092986 | 898.844 | 1041 | 8.44E-07 | 1.02E-01 | 4.37E-03 | 7.97E-10 |
| | S1080 | 6 | 5 | 1 | 20 | 12312034 | 12754957 | 442.924 | 812 | 5.08E-08 | 1.02E-01 | 2.18E-03 | 4.79E-11 |
| | S1081 | 6 | 5 | 1 | 20 | 14529989 | 15582464 | 1052.476 | 1783 | 5.08E-08 | 1.02E-01 | 2.18E-03 | 4.79E-11 |

| | | | | | | | | | | | | | |
|---|---|---|---|---|---|---|---|---|---|---|---|---|---|
| | S1106 | 5 | 4 | 1 | 1 | 204388918 | 205845567 | 1456.65 | 2230 | 4.46E-06 | 8.16E-02 | 2.18E-03 | 4.21E-09 |
| | S1107 | 5 | 4 | 1 | 1 | 212431624 | 212590188 | 158.565 | 340 | 4.46E-06 | 8.16E-02 | 2.18E-03 | 4.21E-09 |
| | S1116 | 5 | 4 | 1 | 2 | 64961311 | 65397338 | 436.028 | 785 | 4.46E-06 | 8.16E-02 | 2.18E-03 | 4.21E-09 |
| | S1296 | 5 | 4 | 1 | 16 | 73656643 | 74025872 | 369.23 | 432 | 4.46E-06 | 8.16E-02 | 2.18E-03 | 4.21E-09 |
| | S1325 | 5 | 4 | 1 | 21 | 39889466 | 39994119 | 104.654 | 264 | 4.46E-06 | 8.16E-02 | 2.18E-03 | 4.21E-09 |
| | S1341 | 4 | 4 | 0 | 1 | 159797454 | 159943263 | 145.81 | 263 | 1.23E-07 | 8.16E-02 | 0.00E+00 | 1.16E-10 |
| | S1443 | 4 | 4 | 0 | 8 | 125749215 | 127191815 | 1442.601 | 2246 | 1.23E-07 | 8.16E-02 | 0.00E+00 | 1.16E-10 |
| | S1623 | 3 | 3 | 0 | 9 | 1940954 | 3629808 | 1688.855 | 3403 | 1.48E-05 | 6.12E-02 | 0.00E+00 | 1.40E-08 |
| | S1650 | 3 | 3 | 0 | 11 | 2896280 | 3419246 | 522.967 | 955 | 1.48E-05 | 6.12E-02 | 0.00E+00 | 1.40E-08 |
| | S1651 | 3 | 3 | 0 | 11 | 4879183 | 5223561 | 344.379 | 857 | 1.48E-05 | 6.12E-02 | 0.00E+00 | 1.40E-08 |
| | S1665 | 3 | 3 | 0 | 13 | 22346187 | 23414536 | 1068.35 | 2389 | 1.48E-05 | 6.12E-02 | 0.00E+00 | 1.40E-08 |
| | S1666 | 3 | 3 | 0 | 13 | 73852645 | 74103309 | 250.665 | 391 | 1.48E-05 | 6.12E-02 | 0.00E+00 | 1.40E-08 |
| **AMR** | S77 | 5 | 4 | 1 | 8 | 42230547 | 43554958 | 1324.412 | 1012 | 8.34E-05 | 2.00E-01 | 7.19E-03 | 2.38E-07 |
| | S81 | 5 | 4 | 1 | 9 | 79266337 | 80842614 | 1576.278 | 2893 | 8.34E-05 | 2.00E-01 | 7.19E-03 | 2.38E-07 |

| | | | | | | | | | | | | | |
|---|---|---|---|---|---|---|---|---|---|---|---|---|---|
| | S82 | 5 | 5 | 0 | 9 | 82135547 | 84169832 | 2034.286 | 2754 | 1.13E-07 | 2.50E-01 | 0.00E+00 | 3.22E-10 |
| | S88 | 5 | 4 | 1 | 17 | 20558851 | 20936047 | 377.197 | 353 | 8.34E-05 | 2.00E-01 | 7.19E-03 | 2.38E-07 |
| | S121 | 4 | 4 | 0 | 10 | 24128513 | 25123571 | 995.059 | 1380 | 4.73E-06 | 2.00E-01 | 0.00E+00 | 1.35E-08 |
| | S122 | 4 | 4 | 0 | 10 | 25129805 | 25475212 | 345.408 | 485 | 4.73E-06 | 2.00E-01 | 0.00E+00 | 1.35E-08 |
| | S136 | 4 | 4 | 0 | 17 | 9973671 | 10259230 | 285.56 | 658 | 4.73E-06 | 2.00E-01 | 0.00E+00 | 1.35E-08 |
| **CAS** | S42 | 11 | 4 | 7 | 2 | 97020004 | 97717987 | 697.984 | 133 | 6.47E-05 | 2.67E-01 | 2.62E-02 | 1.13E-07 |
| | S48 | 11 | 4 | 7 | 8 | 98529140 | 99858154 | 1329.015 | 696 | 6.47E-05 | 2.67E-01 | 2.62E-02 | 1.13E-07 |
| | S68 | 9 | 4 | 5 | 7 | 118714624 | 118870761 | 156.138 | 156 | 5.09E-06 | 2.67E-01 | 1.87E-02 | 8.85E-09 |
| | S138 | 6 | 3 | 3 | 2 | 140629282 | 141846442 | 1217.161 | 2202 | 6.07E-05 | 2.00E-01 | 1.12E-02 | 1.05E-07 |
| | S139 | 6 | 3 | 3 | 2 | 141862174 | 141991767 | 129.594 | 186 | 6.07E-05 | 2.00E-01 | 1.12E-02 | 1.05E-07 |
| | S141 | 6 | 3 | 3 | 2 | 147415548 | 148475487 | 1059.94 | 1237 | 6.07E-05 | 2.00E-01 | 1.12E-02 | 1.05E-07 |
| | S176 | 6 | 3 | 3 | 7 | 43944655 | 44218344 | 273.69 | 375 | 6.07E-05 | 2.00E-01 | 1.12E-02 | 1.05E-07 |
| | S187 | 6 | 3 | 3 | 8 | 113104457 | 113490131 | 385.675 | 488 | 6.07E-05 | 2.00E-01 | 1.12E-02 | 1.05E-07 |
| | S199 | 6 | 3 | 3 | 11 | 97257693 | 97804305 | 546.613 | 649 | 6.07E-05 | 2.00E-01 | 1.12E-02 | 1.05E-07 |

| | | | | | | | | | | | | | |
|---|---|---|---|---|---|---|---|---|---|---|---|---|---|
| | S201 | 6 | 4 | 2 | 12 | 40662517 | 42677591 | 2015.075 | 3661 | 4.94E-09 | 2.67E-01 | 7.49E-03 | 8.60E-12 |
| | S207 | 6 | 3 | 3 | 17 | 18258430 | 18872587 | 614.158 | 523 | 6.07E-05 | 2.00E-01 | 1.12E-02 | 1.05E-07 |
| | S232 | 5 | 3 | 2 | 2 | 103942665 | 104621405 | 678.741 | 837 | 7.06E-06 | 2.00E-01 | 7.49E-03 | 1.23E-08 |
| | S234 | 5 | 3 | 2 | 2 | 112340863 | 113461230 | 1120.368 | 1486 | 7.06E-06 | 2.00E-01 | 7.49E-03 | 1.23E-08 |
| | S235 | 5 | 3 | 2 | 2 | 143505975 | 143954195 | 448.221 | 606 | 7.06E-06 | 2.00E-01 | 7.49E-03 | 1.23E-08 |
| | S236 | 5 | 3 | 2 | 2 | 143969076 | 144543747 | 574.672 | 387 | 7.06E-06 | 2.00E-01 | 7.49E-03 | 1.23E-08 |
| | S237 | 5 | 3 | 2 | 2 | 148806575 | 149304921 | 498.347 | 580 | 7.06E-06 | 2.00E-01 | 7.49E-03 | 1.23E-08 |
| | S239 | 5 | 3 | 2 | 2 | 151144438 | 152252685 | 1108.248 | 1397 | 7.06E-06 | 2.00E-01 | 7.49E-03 | 1.23E-08 |
| | S240 | 5 | 3 | 2 | 2 | 152328192 | 153142511 | 814.32 | 1480 | 7.06E-06 | 2.00E-01 | 7.49E-03 | 1.23E-08 |
| | S241 | 5 | 4 | 1 | 2 | 158883384 | 160573098 | 1689.715 | 2874 | 7.89E-11 | 2.67E-01 | 3.75E-03 | 1.37E-13 |
| | S276 | 5 | 3 | 2 | 4 | 132016378 | 132804596 | 788.219 | 1458 | 7.06E-06 | 2.00E-01 | 7.49E-03 | 1.23E-08 |
| | S277 | 5 | 3 | 2 | 4 | 141052916 | 141188498 | 135.583 | 165 | 7.06E-06 | 2.00E-01 | 7.49E-03 | 1.23E-08 |
| | S278 | 5 | 3 | 2 | 4 | 141239910 | 141557767 | 317.858 | 423 | 7.06E-06 | 2.00E-01 | 7.49E-03 | 1.23E-08 |
| | S308 | 5 | 3 | 2 | 7 | 47666099 | 48697848 | 1031.75 | 1762 | 7.06E-06 | 2.00E-01 | 7.49E-03 | 1.23E-08 |

| | | | | | | | | | | | | | |
|---|---|---|---|---|---|---|---|---|---|---|---|---|---|
| | S311 | 5 | 3 | 2 | 7 | 132498641 | 134063722 | 1565.082 | 1708 | 7.06E-06 | 2.00E-01 | 7.49E-03 | 1.23E-08 |
| | S341 | 5 | 3 | 2 | 12 | 44863086 | 47069014 | 2205.929 | 2919 | 7.06E-06 | 2.00E-01 | 7.49E-03 | 1.23E-08 |
| | S353 | 5 | 3 | 2 | 17 | 14867684 | 17192330 | 2324.647 | 2715 | 7.06E-06 | 2.00E-01 | 7.49E-03 | 1.23E-08 |
| | S354 | 5 | 3 | 2 | 17 | 18960666 | 19329720 | 369.055 | 178 | 7.06E-06 | 2.00E-01 | 7.49E-03 | 1.23E-08 |
| | S382 | 4 | 3 | 1 | 2 | 23540706 | 24466643 | 925.938 | 1132 | 2.86E-07 | 2.00E-01 | 3.75E-03 | 4.97E-10 |
| | S384 | 4 | 3 | 1 | 2 | 29386339 | 30267999 | 881.661 | 1290 | 2.86E-07 | 2.00E-01 | 3.75E-03 | 4.97E-10 |
| | S385 | 4 | 3 | 1 | 2 | 49772441 | 49972265 | 199.825 | 297 | 2.86E-07 | 2.00E-01 | 3.75E-03 | 4.97E-10 |
| | S386 | 4 | 3 | 1 | 2 | 51866901 | 52025961 | 159.061 | 234 | 2.86E-07 | 2.00E-01 | 3.75E-03 | 4.97E-10 |
| | S390 | 4 | 3 | 1 | 2 | 99497959 | 100604995 | 1107.037 | 1328 | 2.86E-07 | 2.00E-01 | 3.75E-03 | 4.97E-10 |
| | S391 | 4 | 3 | 1 | 2 | 100722823 | 101159757 | 436.935 | 722 | 2.86E-07 | 2.00E-01 | 3.75E-03 | 4.97E-10 |
| | S394 | 4 | 3 | 1 | 2 | 154281988 | 154575357 | 293.37 | 477 | 2.86E-07 | 2.00E-01 | 3.75E-03 | 4.97E-10 |
| | S395 | 4 | 3 | 1 | 2 | 154595043 | 155643731 | 1048.689 | 1387 | 2.86E-07 | 2.00E-01 | 3.75E-03 | 4.97E-10 |
| | S396 | 4 | 3 | 1 | 2 | 155825013 | 156877938 | 1052.926 | 1015 | 2.86E-07 | 2.00E-01 | 3.75E-03 | 4.97E-10 |
| | S397 | 4 | 3 | 1 | 2 | 162599870 | 162754693 | 154.824 | 202 | 2.86E-07 | 2.00E-01 | 3.75E-03 | 4.97E-10 |

| | | | | | | | | | | | | | |
|---|---|---|---|---|---|---|---|---|---|---|---|---|---|
| | S398 | 4 | 3 | 1 | 2 | 165453151 | 167634399 | 2181.249 | 3154 | 2.86E-07 | 2.00E-01 | 3.75E-03 | 4.97E-10 |
| | S399 | 4 | 3 | 1 | 2 | 167644704 | 167844661 | 199.958 | 360 | 2.86E-07 | 2.00E-01 | 3.75E-03 | 4.97E-10 |
| | S452 | 4 | 3 | 1 | 6 | 33994939 | 35614884 | 1619.946 | 1978 | 2.86E-07 | 2.00E-01 | 3.75E-03 | 4.97E-10 |
| | S457 | 4 | 4 | 0 | 6 | 110411111 | 111140162 | 729.052 | 1070 | 1.62E-13 | 2.67E-01 | 0.00E+00 | 2.83E-16 |
| | S458 | 4 | 3 | 1 | 6 | 114136451 | 114259142 | 122.692 | 110 | 2.86E-07 | 2.00E-01 | 3.75E-03 | 4.97E-10 |
| | S459 | 4 | 3 | 1 | 6 | 114278343 | 116101118 | 1822.776 | 2629 | 2.86E-07 | 2.00E-01 | 3.75E-03 | 4.97E-10 |
| | S460 | 4 | 3 | 1 | 6 | 116501286 | 117285199 | 783.914 | 1107 | 2.86E-07 | 2.00E-01 | 3.75E-03 | 4.97E-10 |
| | S484 | 4 | 4 | 0 | 8 | 96251190 | 97440997 | 1189.808 | 1302 | 1.62E-13 | 2.67E-01 | 0.00E+00 | 2.83E-16 |
| | S486 | 4 | 3 | 1 | 8 | 98086716 | 98247413 | 160.698 | 268 | 2.86E-07 | 2.00E-01 | 3.75E-03 | 4.97E-10 |
| | S511 | 4 | 3 | 1 | 11 | 75194690 | 76847851 | 1653.162 | 2032 | 2.86E-07 | 2.00E-01 | 3.75E-03 | 4.97E-10 |
| | S512 | 4 | 3 | 1 | 11 | 84667999 | 86999400 | 2331.402 | 3319 | 2.86E-07 | 2.00E-01 | 3.75E-03 | 4.97E-10 |
| | S513 | 4 | 3 | 1 | 11 | 87008557 | 87247736 | 239.18 | 542 | 2.86E-07 | 2.00E-01 | 3.75E-03 | 4.97E-10 |
| | S514 | 4 | 3 | 1 | 11 | 87248673 | 87691952 | 443.28 | 779 | 2.86E-07 | 2.00E-01 | 3.75E-03 | 4.97E-10 |
| | S518 | 4 | 3 | 1 | 11 | 102339006 | 104987712 | 2648.707 | 4346 | 2.86E-07 | 2.00E-01 | 3.75E-03 | 4.97E-10 |

| | | | | | | | | | | | | | |
|---|---|---|---|---|---|---|---|---|---|---|---|---|---|
| | S651 | 3 | 3 | 0 | 6 | 36648823 | 39157799 | 2508.977 | 4342 | 1.42E-09 | 2.00E-01 | 0.00E+00 | 2.47E-12 |
| | S652 | 3 | 3 | 0 | 6 | 39766113 | 40372145 | 606.033 | 888 | 1.42E-09 | 2.00E-01 | 0.00E+00 | 2.47E-12 |
| | S822 | 2 | 2 | 0 | 1 | 210614853 | 210878283 | 263.431 | 521 | 1.05E-05 | 1.33E-01 | 0.00E+00 | 1.82E-08 |
| | S825 | 2 | 2 | 0 | 2 | 10879840 | 12560515 | 1680.676 | 2478 | 1.05E-05 | 1.33E-01 | 0.00E+00 | 1.82E-08 |
| | S826 | 2 | 2 | 0 | 2 | 12951890 | 15863284 | 2911.395 | 4475 | 1.05E-05 | 1.33E-01 | 0.00E+00 | 1.82E-08 |
| | S827 | 2 | 2 | 0 | 2 | 16368968 | 17996554 | 1627.587 | 2063 | 1.05E-05 | 1.33E-01 | 0.00E+00 | 1.82E-08 |
| | S828 | 2 | 2 | 0 | 2 | 18171735 | 20375251 | 2203.517 | 3441 | 1.05E-05 | 1.33E-01 | 0.00E+00 | 1.82E-08 |
| | S840 | 2 | 2 | 0 | 2 | 223312391 | 225222611 | 1910.221 | 2874 | 1.05E-05 | 1.33E-01 | 0.00E+00 | 1.82E-08 |
| | S841 | 2 | 2 | 0 | 2 | 225434130 | 227933798 | 2499.669 | 3670 | 1.05E-05 | 1.33E-01 | 0.00E+00 | 1.82E-08 |
| | S842 | 2 | 2 | 0 | 2 | 227938269 | 229819388 | 1881.12 | 2709 | 1.05E-05 | 1.33E-01 | 0.00E+00 | 1.82E-08 |
| | S901 | 2 | 2 | 0 | 6 | 23750907 | 23913300 | 162.394 | 392 | 1.05E-05 | 1.33E-01 | 0.00E+00 | 1.82E-08 |
| | S903 | 2 | 2 | 0 | 6 | 125419743 | 127356617 | 1936.875 | 1330 | 1.05E-05 | 1.33E-01 | 0.00E+00 | 1.82E-08 |
| | S983 | 2 | 2 | 0 | 15 | 55999329 | 57329488 | 1330.16 | 1791 | 1.05E-05 | 1.33E-01 | 0.00E+00 | 1.82E-08 |
| | S984 | 2 | 2 | 0 | 15 | 58538767 | 60687613 | 2148.847 | 2852 | 1.05E-05 | 1.33E-01 | 0.00E+00 | 1.82E-08 |

| | | | | | | | | | | | | | |
|---|---|---|---|---|---|---|---|---|---|---|---|---|---|
| | S1009 | 2 | 2 | 0 | 19 | 3561221 | 3931179 | 369.959 | 586 | 1.05E-05 | 1.33E-01 | 0.00E+00 | 1.82E-08 |
| | S140 | 71 | 3 | 68 | 10 | 73002790 | 73383033 | 380.244 | 306 | 5.62E-05 | 3.00E-01 | 2.95E-02 | 1.36E-07 |
| | S427 | 22 | 2 | 20 | 6 | 145191755 | 146344288 | 1152.534 | 1464 | 4.52E-06 | 2.00E-01 | 8.69E-03 | 1.10E-08 |
| | S438 | 21 | 2 | 19 | 1 | 102554496 | 103131712 | 577.217 | 564 | 2.51E-06 | 2.00E-01 | 8.25E-03 | 6.09E-09 |
| | S459 | 21 | 2 | 19 | 11 | 85770648 | 86122418 | 351.771 | 384 | 2.51E-06 | 2.00E-01 | 8.25E-03 | 6.09E-09 |
| | S461 | 21 | 2 | 19 | 14 | 40984076 | 41132848 | 148.773 | 364 | 2.51E-06 | 2.00E-01 | 8.25E-03 | 6.09E-09 |
| | S466 | 20 | 2 | 18 | 1 | 103437627 | 103799249 | 361.623 | 109 | 1.32E-06 | 2.00E-01 | 7.82E-03 | 3.19E-09 |
| **EAS** | S488 | 20 | 2 | 18 | 10 | 74084639 | 74226087 | 141.449 | 149 | 1.32E-06 | 2.00E-01 | 7.82E-03 | 3.19E-09 |
| | S509 | 19 | 2 | 17 | 6 | 50667413 | 51009564 | 342.152 | 363 | 6.46E-07 | 2.00E-01 | 7.38E-03 | 1.57E-09 |
| | S510 | 19 | 2 | 17 | 6 | 55218009 | 55320140 | 102.132 | 146 | 6.46E-07 | 2.00E-01 | 7.38E-03 | 1.57E-09 |
| | S511 | 19 | 2 | 17 | 6 | 55330098 | 55541872 | 211.775 | 365 | 6.46E-07 | 2.00E-01 | 7.38E-03 | 1.57E-09 |
| | S515 | 19 | 2 | 17 | 8 | 77709107 | 78018344 | 309.238 | 479 | 6.46E-07 | 2.00E-01 | 7.38E-03 | 1.57E-09 |
| | S540 | 18 | 2 | 16 | 6 | 86358359 | 86643756 | 285.398 | 492 | 2.93E-07 | 2.00E-01 | 6.95E-03 | 7.11E-10 |
| | S579 | 17 | 3 | 14 | 12 | 85074606 | 85243787 | 169.182 | 150 | 2.24E-19 | 3.00E-01 | 6.08E-03 | 5.43E-22 |

| | | | | | | | | | | | | | |
|---|---|---|---|---|---|---|---|---|---|---|---|---|---|
| | S590 | 16 | 2 | 14 | 5 | 88220002 | 88640561 | 420.56 | 369 | 4.51E-08 | 2.00E-01 | 6.08E-03 | 1.09E-10 |
| | S612 | 16 | 2 | 14 | 14 | 41867372 | 42243063 | 375.692 | 533 | 4.51E-08 | 2.00E-01 | 6.08E-03 | 1.09E-10 |
| | S622 | 15 | 2 | 13 | 1 | 92380023 | 93057526 | 677.504 | 462 | 1.47E-08 | 2.00E-01 | 5.65E-03 | 3.57E-11 |
| | S648 | 15 | 3 | 12 | 8 | 82515228 | 82813602 | 298.375 | 451 | 7.10E-22 | 3.00E-01 | 5.21E-03 | 1.72E-24 |
| | S654 | 15 | 2 | 13 | 11 | 82784557 | 82985843 | 201.287 | 221 | 1.47E-08 | 2.00E-01 | 5.65E-03 | 3.57E-11 |
| | S656 | 15 | 2 | 13 | 12 | 83343594 | 83722633 | 379.04 | 687 | 1.47E-08 | 2.00E-01 | 5.65E-03 | 3.57E-11 |
| | S657 | 15 | 2 | 13 | 12 | 83777529 | 84602145 | 824.617 | 1421 | 1.47E-08 | 2.00E-01 | 5.65E-03 | 3.57E-11 |
| | S670 | 14 | 2 | 12 | 1 | 27697089 | 28078183 | 381.095 | 287 | 4.10E-09 | 2.00E-01 | 5.21E-03 | 9.95E-12 |
| | S672 | 14 | 2 | 12 | 2 | 139938532 | 140069194 | 130.663 | 134 | 4.10E-09 | 2.00E-01 | 5.21E-03 | 9.95E-12 |
| | S689 | 14 | 2 | 12 | 6 | 55850178 | 55985642 | 135.465 | 133 | 4.10E-09 | 2.00E-01 | 5.21E-03 | 9.95E-12 |
| | S695 | 14 | 2 | 12 | 8 | 86333471 | 86477333 | 143.863 | 115 | 4.10E-09 | 2.00E-01 | 5.21E-03 | 9.95E-12 |
| | S707 | 14 | 2 | 12 | 17 | 28002165 | 28888684 | 886.52 | 399 | 4.10E-09 | 2.00E-01 | 5.21E-03 | 9.95E-12 |
| | S715 | 13 | 3 | 10 | 2 | 146610620 | 147097731 | 487.112 | 535 | 3.88E-25 | 3.00E-01 | 4.34E-03 | 9.41E-28 |
| | S742 | 13 | 2 | 11 | 8 | 83886231 | 84059832 | 173.602 | 217 | 9.41E-10 | 2.00E-01 | 4.78E-03 | 2.28E-12 |

| | | | | | | | | | | | | | |
|---|---|---|---|---|---|---|---|---|---|---|---|---|---|
| | S744 | 13 | 2 | 11 | 8 | 87259956 | 87449608 | 189.653 | 285 | 9.41E-10 | 2.00E-01 | 4.78E-03 | 2.28E-12 |
| | S753 | 13 | 3 | 10 | 12 | 81497836 | 81620912 | 123.077 | 146 | 3.88E-25 | 3.00E-01 | 4.34E-03 | 9.41E-28 |
| | S758 | 13 | 2 | 11 | 14 | 42863867 | 43859856 | 995.99 | 1748 | 9.41E-10 | 2.00E-01 | 4.78E-03 | 2.28E-12 |
| | S803 | 12 | 2 | 10 | 11 | 104564908 | 104890662 | 325.755 | 326 | 1.70E-10 | 2.00E-01 | 4.34E-03 | 4.12E-13 |
| | S804 | 12 | 3 | 9 | 12 | 80907722 | 81017645 | 109.924 | 182 | 3.56E-27 | 3.00E-01 | 3.91E-03 | 8.63E-30 |
| | S864 | 11 | 2 | 9 | 8 | 86633621 | 86799281 | 165.661 | 225 | 2.25E-11 | 2.00E-01 | 3.91E-03 | 5.45E-14 |
| | S878 | 11 | 2 | 9 | 11 | 105086766 | 105227380 | 140.615 | 128 | 2.25E-11 | 2.00E-01 | 3.91E-03 | 5.45E-14 |
| | S882 | 11 | 2 | 9 | 13 | 66334267 | 66451173 | 116.907 | 149 | 2.25E-11 | 2.00E-01 | 3.91E-03 | 5.45E-14 |
| | S904 | 10 | 2 | 8 | 2 | 24027952 | 24305626 | 277.675 | 293 | 1.99E-12 | 2.00E-01 | 3.48E-03 | 4.84E-15 |
| | S979 | 10 | 2 | 8 | 13 | 66950964 | 67151966 | 201.003 | 174 | 1.99E-12 | 2.00E-01 | 3.48E-03 | 4.84E-15 |
| | S994 | 9 | 2 | 7 | 1 | 34934701 | 36260026 | 1325.326 | 1061 | 1.04E-13 | 2.00E-01 | 3.04E-03 | 2.52E-16 |
| | S996 | 9 | 2 | 7 | 1 | 96138507 | 96403485 | 264.979 | 216 | 1.04E-13 | 2.00E-01 | 3.04E-03 | 2.52E-16 |
| | S1003 | 9 | 2 | 7 | 2 | 24428996 | 25047564 | 618.569 | 702 | 1.04E-13 | 2.00E-01 | 3.04E-03 | 2.52E-16 |
| | S1058 | 9 | 2 | 7 | 7 | 110829559 | 111176421 | 346.863 | 288 | 1.04E-13 | 2.00E-01 | 3.04E-03 | 2.52E-16 |

| | | | | | | | | | | | | | |
|---|---|---|---|---|---|---|---|---|---|---|---|---|---|
| | S1059 | 9 | 2 | 7 | 7 | 114634276 | 114902722 | 268.447 | 217 | 1.04E-13 | 2.00E-01 | 3.04E-03 | 2.52E-16 |
| | S1079 | 9 | 2 | 7 | 11 | 84504716 | 84644243 | 139.528 | 110 | 1.04E-13 | 2.00E-01 | 3.04E-03 | 2.52E-16 |
| | S1092 | 9 | 2 | 7 | 17 | 30226621 | 31418963 | 1192.343 | 776 | 1.04E-13 | 2.00E-01 | 3.04E-03 | 2.52E-16 |
| | S1100 | 8 | 2 | 6 | 1 | 96703314 | 96878909 | 175.596 | 197 | 2.60E-15 | 2.00E-01 | 2.61E-03 | 6.32E-18 |
| | S1104 | 8 | 2 | 6 | 1 | 246691322 | 246868937 | 177.616 | 273 | 2.60E-15 | 2.00E-01 | 2.61E-03 | 6.32E-18 |
| | S1105 | 8 | 2 | 6 | 2 | 22737657 | 22861991 | 124.335 | 173 | 2.60E-15 | 2.00E-01 | 2.61E-03 | 6.32E-18 |
| | S1149 | 8 | 2 | 6 | 6 | 142585268 | 142688704 | 103.437 | 132 | 2.60E-15 | 2.00E-01 | 2.61E-03 | 6.32E-18 |
| | S1156 | 8 | 2 | 6 | 7 | 145227251 | 145448233 | 220.983 | 222 | 2.60E-15 | 2.00E-01 | 2.61E-03 | 6.32E-18 |
| | S1157 | 8 | 2 | 6 | 7 | 145683698 | 146174758 | 491.061 | 477 | 2.60E-15 | 2.00E-01 | 2.61E-03 | 6.32E-18 |
| | S1179 | 8 | 2 | 6 | 11 | 106622331 | 107246772 | 624.442 | 922 | 2.60E-15 | 2.00E-01 | 2.61E-03 | 6.32E-18 |
| | S1215 | 7 | 2 | 5 | 1 | 247268027 | 247387029 | 119.003 | 106 | 2.30E-17 | 2.00E-01 | 2.17E-03 | 5.57E-20 |
| | S1216 | 7 | 2 | 5 | 2 | 22891705 | 23008130 | 116.426 | 133 | 2.30E-17 | 2.00E-01 | 2.17E-03 | 5.57E-20 |
| | S1242 | 7 | 2 | 5 | 4 | 60634189 | 60811323 | 177.135 | 322 | 2.30E-17 | 2.00E-01 | 2.17E-03 | 5.57E-20 |
| | S1288 | 7 | 2 | 5 | 6 | 143218743 | 143325646 | 106.904 | 172 | 2.30E-17 | 2.00E-01 | 2.17E-03 | 5.57E-20 |

| | | | | | | | | | | | | | |
|---|---|---|---|---|---|---|---|---|---|---|---|---|---|
| | S1291 | 7 | 2 | 5 | 7 | 112627005 | 112886665 | 259.661 | 143 | 2.30E-17 | 2.00E-01 | 2.17E-03 | 5.57E-20 |
| | S1318 | 7 | 2 | 5 | 12 | 78309047 | 78650119 | 341.073 | 477 | 2.30E-17 | 2.00E-01 | 2.17E-03 | 5.57E-20 |
| | S1319 | 7 | 2 | 5 | 12 | 78902944 | 79293191 | 390.248 | 366 | 2.30E-17 | 2.00E-01 | 2.17E-03 | 5.57E-20 |
| | S1351 | 6 | 2 | 4 | 1 | 97658971 | 98253450 | 594.48 | 428 | 4.23E-20 | 2.00E-01 | 1.74E-03 | 1.03E-22 |
| | S1461 | 6 | 2 | 4 | 21 | 22990735 | 23242777 | 252.043 | 458 | 4.23E-20 | 2.00E-01 | 1.74E-03 | 1.03E-22 |
| | S1462 | 6 | 2 | 4 | 21 | 23279959 | 23728393 | 448.435 | 645 | 4.23E-20 | 2.00E-01 | 1.74E-03 | 1.03E-22 |
| | S1463 | 6 | 2 | 4 | 21 | 23945679 | 24103230 | 157.552 | 248 | 4.23E-20 | 2.00E-01 | 1.74E-03 | 1.03E-22 |
| | S1466 | 5 | 2 | 3 | 1 | 33459576 | 33579832 | 120.257 | 147 | 6.39E-24 | 2.00E-01 | 1.30E-03 | 1.55E-26 |
| | S1533 | 5 | 2 | 3 | 5 | 14069612 | 14594268 | 524.657 | 447 | 6.39E-24 | 2.00E-01 | 1.30E-03 | 1.55E-26 |
| | S1552 | 5 | 2 | 3 | 7 | 26981837 | 27382217 | 400.381 | 382 | 6.39E-24 | 2.00E-01 | 1.30E-03 | 1.55E-26 |
| | S1563 | 5 | 2 | 3 | 7 | 111793692 | 112144480 | 350.789 | 322 | 6.39E-24 | 2.00E-01 | 1.30E-03 | 1.55E-26 |
| | S1583 | 5 | 2 | 3 | 10 | 82192350 | 82870159 | 677.81 | 760 | 6.39E-24 | 2.00E-01 | 1.30E-03 | 1.55E-26 |
| | S1708 | 4 | 2 | 2 | 5 | 81803826 | 82590479 | 786.654 | 565 | 1.22E-29 | 2.00E-01 | 8.69E-04 | 2.95E-32 |
| | S1763 | 4 | 2 | 2 | 11 | 80489125 | 80942904 | 453.78 | 635 | 1.22E-29 | 2.00E-01 | 8.69E-04 | 2.95E-32 |

| | | | | | | | | | | | | | |
|---|---|---|---|---|---|---|---|---|---|---|---|---|---|
| | S1812 | 3 | 1 | 2 | 1 | 99813047 | 101218347 | 1405.301 | 1252 | 1.81E-05 | 1.00E-01 | 8.69E-04 | 4.39E-08 |
| | S1813 | 3 | 1 | 2 | 1 | 106456826 | 107053821 | 596.996 | 603 | 1.81E-05 | 1.00E-01 | 8.69E-04 | 4.39E-08 |
| | S1814 | 3 | 1 | 2 | 1 | 116879836 | 117138990 | 259.155 | 271 | 1.81E-05 | 1.00E-01 | 8.69E-04 | 4.39E-08 |
| | S1816 | 3 | 1 | 2 | 1 | 159972666 | 160170862 | 198.197 | 224 | 1.81E-05 | 1.00E-01 | 8.69E-04 | 4.39E-08 |
| | S1818 | 3 | 1 | 2 | 1 | 171080339 | 171247758 | 167.42 | 242 | 1.81E-05 | 1.00E-01 | 8.69E-04 | 4.39E-08 |
| | S1823 | 3 | 2 | 1 | 1 | 242657535 | 244144327 | 1486.793 | 1452 | 3.70E-39 | 2.00E-01 | 4.34E-04 | 8.99E-42 |
| | S1824 | 3 | 1 | 2 | 2 | 18373851 | 20413320 | 2039.47 | 2497 | 1.81E-05 | 1.00E-01 | 8.69E-04 | 4.39E-08 |
| | S1825 | 3 | 2 | 1 | 2 | 30337825 | 30769204 | 431.38 | 784 | 3.70E-39 | 2.00E-01 | 4.34E-04 | 8.99E-42 |
| | S1826 | 3 | 1 | 2 | 2 | 34634477 | 35711093 | 1076.617 | 1873 | 1.81E-05 | 1.00E-01 | 8.69E-04 | 4.39E-08 |
| | S1833 | 3 | 1 | 2 | 2 | 118270876 | 118746725 | 475.85 | 470 | 1.81E-05 | 1.00E-01 | 8.69E-04 | 4.39E-08 |
| | S1844 | 3 | 1 | 2 | 3 | 67031572 | 67365561 | 333.99 | 554 | 1.81E-05 | 1.00E-01 | 8.69E-04 | 4.39E-08 |
| | S1850 | 3 | 1 | 2 | 3 | 125266261 | 126925703 | 1659.443 | 2150 | 1.81E-05 | 1.00E-01 | 8.69E-04 | 4.39E-08 |
| | S1862 | 3 | 1 | 2 | 5 | 1857754 | 2111239 | 253.486 | 424 | 1.81E-05 | 1.00E-01 | 8.69E-04 | 4.39E-08 |
| | S1866 | 3 | 1 | 2 | 5 | 34752123 | 34893413 | 141.291 | 161 | 1.81E-05 | 1.00E-01 | 8.69E-04 | 4.39E-08 |

| | | | | | | | | | | | | | |
|---|---|---|---|---|---|---|---|---|---|---|---|---|---|
| | S1869 | 3 | 1 | 2 | 5 | 38128221 | 38310106 | 181.886 | 226 | 1.81E-05 | 1.00E-01 | 8.69E-04 | 4.39E-08 |
| | S1877 | 3 | 1 | 2 | 6 | 9218232 | 9988810 | 770.579 | 735 | 1.81E-05 | 1.00E-01 | 8.69E-04 | 4.39E-08 |
| | S1883 | 3 | 1 | 2 | 6 | 46070087 | 46367432 | 297.346 | 256 | 1.81E-05 | 1.00E-01 | 8.69E-04 | 4.39E-08 |
| | S1890 | 3 | 1 | 2 | 6 | 162333574 | 163245816 | 912.243 | 1217 | 1.81E-05 | 1.00E-01 | 8.69E-04 | 4.39E-08 |
| | S1900 | 3 | 1 | 2 | 7 | 156271913 | 157849514 | 1577.602 | 2437 | 1.81E-05 | 1.00E-01 | 8.69E-04 | 4.39E-08 |
| | S1902 | 3 | 1 | 2 | 8 | 19507372 | 19634163 | 126.792 | 171 | 1.81E-05 | 1.00E-01 | 8.69E-04 | 4.39E-08 |
| | S1908 | 3 | 1 | 2 | 8 | 133775165 | 134958276 | 1183.112 | 1395 | 1.81E-05 | 1.00E-01 | 8.69E-04 | 4.39E-08 |
| | S1909 | 3 | 1 | 2 | 8 | 135282086 | 135729689 | 447.604 | 409 | 1.81E-05 | 1.00E-01 | 8.69E-04 | 4.39E-08 |
| | S1910 | 3 | 1 | 2 | 8 | 142435107 | 143232207 | 797.101 | 1087 | 1.81E-05 | 1.00E-01 | 8.69E-04 | 4.39E-08 |
| | S1911 | 3 | 1 | 2 | 9 | 23185204 | 24689761 | 1504.558 | 1391 | 1.81E-05 | 1.00E-01 | 8.69E-04 | 4.39E-08 |
| | S1918 | 3 | 1 | 2 | 9 | 127861807 | 128395939 | 534.133 | 584 | 1.81E-05 | 1.00E-01 | 8.69E-04 | 4.39E-08 |
| | S1927 | 3 | 1 | 2 | 11 | 112348647 | 112831159 | 482.513 | 528 | 1.81E-05 | 1.00E-01 | 8.69E-04 | 4.39E-08 |
| | S1950 | 3 | 1 | 2 | 17 | 19986129 | 20229581 | 243.453 | 275 | 1.81E-05 | 1.00E-01 | 8.69E-04 | 4.39E-08 |
| | S1952 | 3 | 1 | 2 | 18 | 29163891 | 29419557 | 255.667 | 230 | 1.81E-05 | 1.00E-01 | 8.69E-04 | 4.39E-08 |

| | | | | | | | | | | | | | |
|---|---|---|---|---|---|---|---|---|---|---|---|---|---|
| | S1959 | 2 | 1 | 1 | 1 | 29373186 | 29802640 | 429.455 | 627 | 1.18E-07 | 1.00E-01 | 4.34E-04 | 2.86E-10 |
| | S1967 | 2 | 1 | 1 | 1 | 229709434 | 230647630 | 938.197 | 1143 | 1.18E-07 | 1.00E-01 | 4.34E-04 | 2.86E-10 |
| | S1968 | 2 | 1 | 1 | 1 | 244488897 | 244753806 | 264.91 | 210 | 1.18E-07 | 1.00E-01 | 4.34E-04 | 2.86E-10 |
| | S1969 | 2 | 1 | 1 | 2 | 322579 | 1778691 | 1456.113 | 2035 | 1.18E-07 | 1.00E-01 | 4.34E-04 | 2.86E-10 |
| | S1985 | 2 | 1 | 1 | 3 | 171456424 | 171854880 | 398.457 | 416 | 1.18E-07 | 1.00E-01 | 4.34E-04 | 2.86E-10 |
| | S1989 | 2 | 1 | 1 | 4 | 94971560 | 95242732 | 271.173 | 261 | 1.18E-07 | 1.00E-01 | 4.34E-04 | 2.86E-10 |
| | S1995 | 2 | 1 | 1 | 5 | 6293401 | 8055735 | 1762.335 | 2205 | 1.18E-07 | 1.00E-01 | 4.34E-04 | 2.86E-10 |
| | S1998 | 2 | 1 | 1 | 5 | 32869565 | 33719331 | 849.767 | 996 | 1.18E-07 | 1.00E-01 | 4.34E-04 | 2.86E-10 |
| | S2004 | 2 | 1 | 1 | 6 | 1846822 | 3048454 | 1201.633 | 1609 | 1.18E-07 | 1.00E-01 | 4.34E-04 | 2.86E-10 |
| | S2007 | 2 | 1 | 1 | 6 | 138110216 | 138837055 | 726.84 | 793 | 1.18E-07 | 1.00E-01 | 4.34E-04 | 2.86E-10 |
| | S2013 | 2 | 1 | 1 | 7 | 68164301 | 68347681 | 183.381 | 292 | 1.18E-07 | 1.00E-01 | 4.34E-04 | 2.86E-10 |
| | S2021 | 2 | 1 | 1 | 8 | 1478333 | 1749240 | 270.908 | 474 | 1.18E-07 | 1.00E-01 | 4.34E-04 | 2.86E-10 |
| | S2027 | 2 | 1 | 1 | 9 | 20527697 | 21940018 | 1412.322 | 1990 | 1.18E-07 | 1.00E-01 | 4.34E-04 | 2.86E-10 |
| | S2028 | 2 | 1 | 1 | 9 | 85747002 | 86611354 | 864.353 | 699 | 1.18E-07 | 1.00E-01 | 4.34E-04 | 2.86E-10 |

| | | | | | | | | | | | | | |
|---|---|---|---|---|---|---|---|---|---|---|---|---|---|
| | S2034 | 2 | 1 | 1 | 10 | 54527929 | 55092722 | 564.794 | 779 | 1.18E-07 | 1.00E-01 | 4.34E-04 | 2.86E-10 |
| | S2036 | 2 | 1 | 1 | 11 | 111289904 | 111998826 | 708.923 | 489 | 1.18E-07 | 1.00E-01 | 4.34E-04 | 2.86E-10 |
| | S2047 | 2 | 1 | 1 | 14 | 71134362 | 71755168 | 620.807 | 483 | 1.18E-07 | 1.00E-01 | 4.34E-04 | 2.86E-10 |
| | S2063 | 2 | 1 | 1 | 19 | 33673118 | 34338451 | 665.334 | 647 | 1.18E-07 | 1.00E-01 | 4.34E-04 | 2.86E-10 |
| | S2065 | 2 | 2 | 0 | 19 | 45042304 | 46610987 | 1568.684 | 1941 | 3.74E-58 | 2.00E-01 | 0.00E+00 | 9.07E-61 |
| | S2066 | 2 | 2 | 0 | 21 | 16643809 | 17950673 | 1306.865 | 1830 | 3.74E-58 | 2.00E-01 | 0.00E+00 | 9.07E-61 |
| | S2068 | 2 | 1 | 1 | 21 | 36541167 | 36645075 | 103.909 | 121 | 1.18E-07 | 1.00E-01 | 4.34E-04 | 2.86E-10 |
| **EUR** | S568 | 107 | 7 | 100 | 3 | 163220382 | 163432481 | 212.1 | 401 | 8.67E-08 | 9.33E-02 | 1.33E-02 | 5.82E-11 |
| | S598 | 103 | 7 | 96 | 3 | 162726490 | 162941917 | 215.428 | 241 | 3.94E-08 | 9.33E-02 | 1.28E-02 | 2.64E-11 |
| | S717 | 86 | 7 | 79 | 4 | 127010622 | 127332691 | 322.07 | 463 | 6.03E-10 | 9.33E-02 | 1.05E-02 | 4.04E-13 |
| | S718 | 86 | 7 | 79 | 4 | 127342147 | 127470436 | 128.29 | 122 | 6.03E-10 | 9.33E-02 | 1.05E-02 | 4.04E-13 |
| | S732 | 83 | 6 | 77 | 1 | 45483005 | 46104019 | 621.015 | 973 | 1.82E-07 | 8.00E-02 | 1.03E-02 | 1.22E-10 |
| | S735 | 82 | 6 | 76 | 4 | 150376254 | 150861537 | 485.284 | 611 | 1.44E-07 | 8.00E-02 | 1.01E-02 | 9.67E-11 |
| | S788 | 75 | 8 | 67 | 3 | 136228302 | 136957860 | 729.559 | 999 | 2.31E-15 | 1.07E-01 | 8.93E-03 | 1.55E-18 |

| | | | | | | | | | | | | | |
|---|---|---|---|---|---|---|---|---|---|---|---|---|---|
| | S796 | 75 | 7 | 68 | 10 | 103044198 | 103193308 | 149.111 | 260 | 1.47E-11 | 9.33E-02 | 9.06E-03 | 9.86E-15 |
| | S841 | 70 | 10 | 60 | 16 | 47075338 | 47946642 | 871.305 | 378 | 1.20E-26 | 1.33E-01 | 7.99E-03 | 8.02E-30 |
| | S843 | 69 | 7 | 62 | 7 | 118321110 | 118510547 | 189.438 | 249 | 1.18E-12 | 9.33E-02 | 8.26E-03 | 7.90E-16 |
| | S881 | 64 | 6 | 58 | 1 | 92419450 | 93018831 | 599.382 | 859 | 6.73E-10 | 8.00E-02 | 7.73E-03 | 4.51E-13 |
| | S900 | 63 | 5 | 58 | 11 | 66581032 | 66906236 | 325.205 | 227 | 7.23E-07 | 6.67E-02 | 7.73E-03 | 4.85E-10 |
| | S933 | 61 | 8 | 53 | 5 | 60612700 | 61170442 | 557.743 | 801 | 3.33E-19 | 1.07E-01 | 7.06E-03 | 2.23E-22 |
| | S953 | 59 | 5 | 54 | 3 | 161193918 | 161461944 | 268.027 | 332 | 2.32E-07 | 6.67E-02 | 7.20E-03 | 1.56E-10 |
| | S960 | 59 | 5 | 54 | 5 | 88208666 | 88354818 | 146.153 | 160 | 2.32E-07 | 6.67E-02 | 7.20E-03 | 1.56E-10 |
| | S970 | 58 | 5 | 53 | 3 | 160982708 | 161090052 | 107.345 | 163 | 1.71E-07 | 6.67E-02 | 7.06E-03 | 1.15E-10 |
| | S1036 | 55 | 6 | 49 | 5 | 101986791 | 102198676 | 211.886 | 499 | 1.23E-11 | 8.00E-02 | 6.53E-03 | 8.28E-15 |
| | S1057 | 54 | 6 | 48 | 5 | 102250985 | 102534752 | 283.768 | 580 | 7.30E-12 | 8.00E-02 | 6.40E-03 | 4.89E-15 |
| | S1077 | 53 | 5 | 48 | 5 | 41674753 | 42265491 | 590.739 | 748 | 3.08E-08 | 6.67E-02 | 6.40E-03 | 2.07E-11 |
| | S1089 | 53 | 5 | 48 | 11 | 28030407 | 28216895 | 186.489 | 180 | 3.08E-08 | 6.67E-02 | 6.40E-03 | 2.07E-11 |
| | S1097 | 52 | 5 | 47 | 2 | 99249653 | 99525207 | 275.555 | 417 | 2.10E-08 | 6.67E-02 | 6.26E-03 | 1.41E-11 |

| | | | | | | | | | | | | | |
|---|---|---|---|---|---|---|---|---|---|---|---|---|---|
| | S1105 | 52 | 5 | 47 | 5 | 42427878 | 42569483 | 141.606 | 200 | 2.10E-08 | 6.67E-02 | 6.26E-03 | 1.41E-11 |
| | S1106 | 52 | 5 | 47 | 5 | 42670202 | 42814715 | 144.514 | 240 | 2.10E-08 | 6.67E-02 | 6.26E-03 | 1.41E-11 |
| | S1117 | 51 | 5 | 46 | 2 | 98943776 | 99126956 | 183.181 | 227 | 1.42E-08 | 6.67E-02 | 6.13E-03 | 9.49E-12 |
| | S1135 | 50 | 5 | 45 | 3 | 160436998 | 160601723 | 164.726 | 176 | 9.37E-09 | 6.67E-02 | 6.00E-03 | 6.28E-12 |
| | S1154 | 49 | 5 | 44 | 5 | 51763217 | 51878223 | 115.007 | 187 | 6.10E-09 | 6.67E-02 | 5.86E-03 | 4.09E-12 |
| | S1161 | 49 | 4 | 45 | 11 | 57753377 | 57897864 | 144.488 | 150 | 1.27E-05 | 5.33E-02 | 6.00E-03 | 8.49E-09 |
| | S1178 | 48 | 8 | 40 | 5 | 85531640 | 85871407 | 339.768 | 701 | 8.93E-25 | 1.07E-01 | 5.33E-03 | 5.99E-28 |
| | S1195 | 47 | 4 | 43 | 2 | 57393323 | 57618261 | 224.939 | 632 | 7.24E-06 | 5.33E-02 | 5.73E-03 | 4.85E-09 |
| | S1200 | 47 | 5 | 42 | 3 | 158089171 | 158580216 | 491.046 | 1021 | 2.45E-09 | 6.67E-02 | 5.60E-03 | 1.64E-12 |
| | S1237 | 46 | 7 | 39 | 15 | 72670982 | 72858511 | 187.53 | 280 | 1.68E-19 | 9.33E-02 | 5.20E-03 | 1.13E-22 |
| | S1269 | 44 | 5 | 39 | 4 | 115601444 | 115708695 | 107.252 | 145 | 5.33E-10 | 6.67E-02 | 5.20E-03 | 3.58E-13 |
| | S1283 | 44 | 6 | 38 | 9 | 99536384 | 99873516 | 337.133 | 210 | 1.02E-14 | 8.00E-02 | 5.06E-03 | 6.85E-18 |
| | S1297 | 43 | 5 | 38 | 5 | 144360132 | 144654828 | 294.697 | 415 | 3.06E-10 | 6.67E-02 | 5.06E-03 | 2.05E-13 |
| | S1298 | 43 | 5 | 38 | 5 | 144825397 | 145199225 | 373.829 | 732 | 3.06E-10 | 6.67E-02 | 5.06E-03 | 2.05E-13 |

| | | | | | | | | | | | | | |
|---|---|---|---|---|---|---|---|---|---|---|---|---|---|
| | S1327 | 42 | 5 | 37 | 5 | 99075682 | 99206934 | 131.253 | 240 | 1.71E-10 | 6.67E-02 | 4.93E-03 | 1.15E-13 |
| | S1330 | 42 | 5 | 37 | 6 | 35138422 | 35604002 | 465.581 | 599 | 1.71E-10 | 6.67E-02 | 4.93E-03 | 1.15E-13 |
| | S1342 | 42 | 7 | 35 | 15 | 82531989 | 83265360 | 733.372 | 1071 | 1.87E-21 | 9.33E-02 | 4.66E-03 | 1.26E-24 |
| | S1354 | 41 | 6 | 35 | 5 | 99997761 | 100510473 | 512.713 | 658 | 7.63E-16 | 8.00E-02 | 4.66E-03 | 5.12E-19 |
| | S1360 | 41 | 5 | 36 | 7 | 86122679 | 86256197 | 133.519 | 248 | 9.30E-11 | 6.67E-02 | 4.80E-03 | 6.24E-14 |
| | S1391 | 40 | 4 | 36 | 4 | 131500825 | 131711689 | 210.865 | 431 | 6.63E-07 | 5.33E-02 | 4.80E-03 | 4.44E-10 |
| | S1395 | 40 | 4 | 36 | 5 | 101043619 | 101234176 | 190.558 | 276 | 6.63E-07 | 5.33E-02 | 4.80E-03 | 4.44E-10 |
| | S1421 | 39 | 5 | 34 | 5 | 100635457 | 100804169 | 168.713 | 239 | 2.50E-11 | 6.67E-02 | 4.53E-03 | 1.68E-14 |
| | S1422 | 39 | 4 | 35 | 5 | 100875291 | 100980866 | 105.576 | 117 | 4.39E-07 | 5.33E-02 | 4.66E-03 | 2.95E-10 |
| | S1442 | 39 | 4 | 35 | 16 | 70449053 | 70618451 | 169.399 | 169 | 4.39E-07 | 5.33E-02 | 4.66E-03 | 2.95E-10 |
| | S1448 | 38 | 4 | 34 | 2 | 57049020 | 57152377 | 103.358 | 229 | 2.85E-07 | 5.33E-02 | 4.53E-03 | 1.91E-10 |
| | S1455 | 38 | 5 | 33 | 4 | 116822583 | 116933293 | 110.711 | 225 | 1.23E-11 | 6.67E-02 | 4.40E-03 | 8.28E-15 |
| | S1458 | 38 | 4 | 34 | 4 | 131025609 | 131262485 | 236.877 | 382 | 2.85E-07 | 5.33E-02 | 4.53E-03 | 1.91E-10 |
| | S1491 | 37 | 6 | 31 | 5 | 98403473 | 98608014 | 204.542 | 405 | 1.25E-17 | 8.00E-02 | 4.13E-03 | 8.39E-21 |

| | | | | | | | | | | | | | |
|---|---|---|---|---|---|---|---|---|---|---|---|---|---|
| | S1506 | 37 | 6 | 31 | 10 | 105550827 | 105729875 | 179.049 | 317 | 1.25E-17 | 8.00E-02 | 4.13E-03 | 8.39E-21 |
| | S1512 | 37 | 4 | 33 | 13 | 88983387 | 89158806 | 175.42 | 172 | 1.81E-07 | 5.33E-02 | 4.40E-03 | 1.21E-10 |
| | S1528 | 36 | 4 | 32 | 5 | 40748166 | 40896378 | 148.213 | 299 | 1.12E-07 | 5.33E-02 | 4.26E-03 | 7.50E-11 |
| | S1535 | 36 | 5 | 31 | 5 | 152961976 | 153068546 | 106.571 | 183 | 2.67E-12 | 6.67E-02 | 4.13E-03 | 1.79E-15 |
| | S1559 | 36 | 4 | 32 | 14 | 64147430 | 64414763 | 267.334 | 444 | 1.12E-07 | 5.33E-02 | 4.26E-03 | 7.50E-11 |
| | S1578 | 35 | 4 | 31 | 5 | 40351325 | 40562747 | 211.423 | 424 | 6.73E-08 | 5.33E-02 | 4.13E-03 | 4.51E-11 |
| | S1599 | 35 | 5 | 30 | 11 | 105245498 | 105399022 | 153.525 | 214 | 1.16E-12 | 6.67E-02 | 4.00E-03 | 7.78E-16 |
| | S1615 | 34 | 4 | 30 | 5 | 40107100 | 40297252 | 190.153 | 515 | 3.93E-08 | 5.33E-02 | 4.00E-03 | 2.64E-11 |
| | S1626 | 34 | 6 | 28 | 7 | 126199359 | 126319192 | 119.834 | 218 | 3.04E-19 | 8.00E-02 | 3.73E-03 | 2.04E-22 |
| | S1639 | 34 | 5 | 29 | 11 | 104830020 | 104930680 | 100.661 | 107 | 4.81E-13 | 6.67E-02 | 3.86E-03 | 3.23E-16 |
| | S1648 | 34 | 4 | 30 | 14 | 45875744 | 46200129 | 324.386 | 640 | 3.93E-08 | 5.33E-02 | 4.00E-03 | 2.64E-11 |
| | S1649 | 34 | 5 | 29 | 14 | 64443730 | 64598978 | 155.249 | 224 | 4.81E-13 | 6.67E-02 | 3.86E-03 | 3.23E-16 |
| | S1650 | 34 | 4 | 30 | 15 | 50918936 | 51207300 | 288.365 | 506 | 3.93E-08 | 5.33E-02 | 4.00E-03 | 2.64E-11 |
| | S1651 | 34 | 8 | 26 | 15 | 64144763 | 64757158 | 612.396 | 768 | 1.58E-35 | 1.07E-01 | 3.46E-03 | 1.06E-38 |

| | | | | | | | | | | | | | |
|---|---|---|---|---|---|---|---|---|---|---|---|---|---|
| | S1659 | 33 | 4 | 29 | 1 | 75049336 | 75362848 | 313.513 | 512 | 2.22E-08 | 5.33E-02 | 3.86E-03 | 1.49E-11 |
| | S1660 | 33 | 6 | 27 | 1 | 102752294 | 103347535 | 595.242 | 1608 | 7.60E-20 | 8.00E-02 | 3.60E-03 | 5.10E-23 |
| | S1661 | 33 | 6 | 27 | 1 | 103347850 | 103497405 | 149.556 | 182 | 7.60E-20 | 8.00E-02 | 3.60E-03 | 5.10E-23 |
| | S1693 | 33 | 5 | 28 | 9 | 124806523 | 125988959 | 1182.437 | 1276 | 1.89E-13 | 6.67E-02 | 3.73E-03 | 1.27E-16 |
| | S1752 | 32 | 5 | 27 | 9 | 30173292 | 30422187 | 248.896 | 402 | 7.02E-14 | 6.67E-02 | 3.60E-03 | 4.71E-17 |
| | S1763 | 32 | 4 | 28 | 14 | 39253478 | 39418472 | 164.995 | 339 | 1.21E-08 | 5.33E-02 | 3.73E-03 | 8.15E-12 |
| | S1764 | 32 | 4 | 28 | 14 | 39532707 | 39719604 | 186.898 | 400 | 1.21E-08 | 5.33E-02 | 3.73E-03 | 8.15E-12 |
| | S1786 | 31 | 6 | 25 | 1 | 96479319 | 96599318 | 120 | 208 | 3.62E-21 | 8.00E-02 | 3.33E-03 | 2.43E-24 |
| | S1789 | 31 | 4 | 27 | 2 | 54392335 | 54538546 | 146.212 | 292 | 6.38E-09 | 5.33E-02 | 3.60E-03 | 4.28E-12 |
| | S1798 | 31 | 6 | 25 | 3 | 137793741 | 137902489 | 108.749 | 199 | 3.62E-21 | 8.00E-02 | 3.33E-03 | 2.43E-24 |
| | S1809 | 31 | 4 | 27 | 5 | 86884740 | 87001969 | 117.23 | 209 | 6.38E-09 | 5.33E-02 | 3.60E-03 | 4.28E-12 |
| | S1831 | 31 | 4 | 27 | 12 | 40266815 | 40421822 | 155.008 | 387 | 6.38E-09 | 5.33E-02 | 3.60E-03 | 4.28E-12 |
| | S1848 | 31 | 5 | 26 | 22 | 31219664 | 31428273 | 208.61 | 293 | 2.45E-14 | 6.67E-02 | 3.46E-03 | 1.64E-17 |
| | S1854 | 30 | 6 | 24 | 1 | 96628173 | 96819837 | 191.665 | 419 | 6.79E-22 | 8.00E-02 | 3.20E-03 | 4.55E-25 |

| | | | | | | | | | | | | | |
|---|---|---|---|---|---|---|---|---|---|---|---|---|---|
| | S1870 | 30 | 4 | 26 | 3 | 34870090 | 34978458 | 108.369 | 265 | 3.21E-09 | 5.33E-02 | 3.46E-03 | 2.16E-12 |
| | S1872 | 30 | 5 | 25 | 3 | 159760961 | 159879492 | 118.532 | 192 | 7.94E-15 | 6.67E-02 | 3.33E-03 | 5.32E-18 |
| | S1886 | 30 | 6 | 24 | 5 | 119032102 | 119285329 | 253.228 | 323 | 6.79E-22 | 8.00E-02 | 3.20E-03 | 4.55E-25 |
| | S1914 | 30 | 7 | 23 | 16 | 29991755 | 30108956 | 117.202 | 121 | 1.98E-30 | 9.33E-02 | 3.06E-03 | 1.33E-33 |
| | S1937 | 29 | 4 | 25 | 2 | 103834512 | 104099228 | 264.717 | 440 | 1.54E-09 | 5.33E-02 | 3.33E-03 | 1.04E-12 |
| | S1959 | 29 | 4 | 25 | 4 | 147148704 | 147327594 | 178.891 | 191 | 1.54E-09 | 5.33E-02 | 3.33E-03 | 1.04E-12 |
| | S1968 | 29 | 3 | 26 | 6 | 68961693 | 69223908 | 262.216 | 558 | 3.18E-05 | 4.00E-02 | 3.46E-03 | 2.13E-08 |
| | S1971 | 29 | 4 | 25 | 7 | 87944332 | 88048209 | 103.878 | 103 | 1.54E-09 | 5.33E-02 | 3.33E-03 | 1.04E-12 |
| | S1981 | 29 | 8 | 21 | 10 | 99630143 | 99870954 | 240.812 | 336 | 7.04E-42 | 1.07E-01 | 2.80E-03 | 4.72E-45 |
| | S1984 | 29 | 4 | 25 | 12 | 41120483 | 41284161 | 163.679 | 295 | 1.54E-09 | 5.33E-02 | 3.33E-03 | 1.04E-12 |
| | S2016 | 28 | 3 | 25 | 4 | 65440570 | 65654198 | 213.629 | 528 | 2.12E-05 | 4.00E-02 | 3.33E-03 | 1.42E-08 |
| | S2028 | 28 | 7 | 21 | 5 | 118190909 | 118378925 | 188.017 | 373 | 1.13E-32 | 9.33E-02 | 2.80E-03 | 7.59E-36 |
| | S2031 | 28 | 7 | 21 | 5 | 127800776 | 127991179 | 190.404 | 259 | 1.13E-32 | 9.33E-02 | 2.80E-03 | 7.59E-36 |
| | S2043 | 28 | 3 | 25 | 8 | 66251491 | 66547961 | 296.471 | 450 | 2.12E-05 | 4.00E-02 | 3.33E-03 | 1.42E-08 |

| | | | | | | | | | | | | | |
|---|---|---|---|---|---|---|---|---|---|---|---|---|---|
| | S2056 | 28 | 5 | 23 | 13 | 83363264 | 83515286 | 152.023 | 263 | 6.58E-16 | 6.67E-02 | 3.06E-03 | 4.42E-19 |
| | S2068 | 27 | 3 | 24 | 1 | 152969457 | 153087421 | 117.965 | 209 | 1.37E-05 | 4.00E-02 | 3.20E-03 | 9.16E-09 |
| | S2070 | 27 | 3 | 24 | 2 | 25710129 | 25841729 | 131.601 | 171 | 1.37E-05 | 4.00E-02 | 3.20E-03 | 9.16E-09 |
| | S2073 | 27 | 3 | 24 | 2 | 80726281 | 80951884 | 225.604 | 300 | 1.37E-05 | 4.00E-02 | 3.20E-03 | 9.16E-09 |
| | S2081 | 27 | 4 | 23 | 3 | 35625565 | 35765955 | 140.391 | 222 | 3.03E-10 | 5.33E-02 | 3.06E-03 | 2.03E-13 |
| | S2083 | 27 | 3 | 24 | 3 | 153991255 | 154128535 | 137.281 | 322 | 1.37E-05 | 4.00E-02 | 3.20E-03 | 9.16E-09 |
| | S2084 | 27 | 3 | 24 | 3 | 154707839 | 154856915 | 149.077 | 161 | 1.37E-05 | 4.00E-02 | 3.20E-03 | 9.16E-09 |
| | S2090 | 27 | 3 | 24 | 4 | 120037024 | 120425543 | 388.52 | 687 | 1.37E-05 | 4.00E-02 | 3.20E-03 | 9.16E-09 |
| | S2102 | 27 | 3 | 24 | 6 | 68477133 | 68580739 | 103.607 | 337 | 1.37E-05 | 4.00E-02 | 3.20E-03 | 9.16E-09 |
| | S2112 | 27 | 4 | 23 | 9 | 70159361 | 70457170 | 297.81 | 615 | 3.03E-10 | 5.33E-02 | 3.06E-03 | 2.03E-13 |
| | S2114 | 27 | 7 | 20 | 10 | 99295965 | 99494770 | 198.806 | 310 | 6.43E-34 | 9.33E-02 | 2.66E-03 | 4.31E-37 |
| | S2140 | 26 | 4 | 22 | 2 | 73009827 | 73130477 | 120.651 | 163 | 1.22E-10 | 5.33E-02 | 2.93E-03 | 8.21E-14 |
| | S2141 | 26 | 4 | 22 | 2 | 73136702 | 73261579 | 124.878 | 232 | 1.22E-10 | 5.33E-02 | 2.93E-03 | 8.21E-14 |
| | S2142 | 26 | 3 | 23 | 2 | 73267667 | 73717963 | 450.297 | 529 | 8.53E-06 | 4.00E-02 | 3.06E-03 | 5.72E-09 |

| | | | | | | | | | | | | | |
|---|---|---|---|---|---|---|---|---|---|---|---|---|---|
| | S2151 | 26 | 3 | 23 | 3 | 101504877 | 101748891 | 244.015 | 361 | 8.53E-06 | 4.00E-02 | 3.06E-03 | 5.72E-09 |
| | S2153 | 26 | 7 | 19 | 3 | 138301308 | 138406810 | 105.503 | 131 | 2.93E-35 | 9.33E-02 | 2.53E-03 | 1.97E-38 |
| | S2154 | 26 | 7 | 19 | 3 | 138417207 | 138607871 | 190.665 | 202 | 2.93E-35 | 9.33E-02 | 2.53E-03 | 1.97E-38 |
| | S2156 | 26 | 3 | 23 | 3 | 154867643 | 154995835 | 128.193 | 149 | 8.53E-06 | 4.00E-02 | 3.06E-03 | 5.72E-09 |
| | S2160 | 26 | 5 | 21 | 4 | 105103424 | 105382725 | 279.302 | 449 | 3.73E-17 | 6.67E-02 | 2.80E-03 | 2.50E-20 |
| | S2162 | 26 | 5 | 21 | 4 | 105586876 | 105871223 | 284.348 | 445 | 3.73E-17 | 6.67E-02 | 2.80E-03 | 2.50E-20 |
| | S2165 | 26 | 3 | 23 | 4 | 146090945 | 146242957 | 152.013 | 202 | 8.53E-06 | 4.00E-02 | 3.06E-03 | 5.72E-09 |
| | S2168 | 26 | 4 | 22 | 4 | 158052381 | 158170954 | 118.574 | 136 | 1.22E-10 | 5.33E-02 | 2.93E-03 | 8.21E-14 |
| | S2179 | 26 | 5 | 21 | 7 | 89508645 | 89645391 | 136.747 | 146 | 3.73E-17 | 6.67E-02 | 2.80E-03 | 2.50E-20 |
| | S2181 | 26 | 6 | 20 | 7 | 117312687 | 117559590 | 246.904 | 274 | 2.33E-25 | 8.00E-02 | 2.66E-03 | 1.56E-28 |
| | S2193 | 26 | 4 | 22 | 12 | 43309248 | 43445404 | 136.157 | 247 | 1.22E-10 | 5.33E-02 | 2.93E-03 | 8.21E-14 |
| | S2213 | 25 | 3 | 22 | 1 | 77614023 | 78049412 | 435.39 | 719 | 5.13E-06 | 4.00E-02 | 2.93E-03 | 3.44E-09 |
| | S2224 | 25 | 3 | 22 | 2 | 109483602 | 109664198 | 180.597 | 338 | 5.13E-06 | 4.00E-02 | 2.93E-03 | 3.44E-09 |
| | S2237 | 25 | 4 | 21 | 3 | 120945746 | 121049185 | 103.44 | 253 | 4.60E-11 | 5.33E-02 | 2.80E-03 | 3.09E-14 |

| | | | | | | | | | | | | | |
|---|---|---|---|---|---|---|---|---|---|---|---|---|---|
| | S2257 | 25 | 4 | 21 | 5 | 91501022 | 91648736 | 147.715 | 186 | 4.60E-11 | 5.33E-02 | 2.80E-03 | 3.09E-14 |
| | S2273 | 25 | 5 | 20 | 7 | 110843621 | 111219777 | 376.157 | 380 | 7.48E-18 | 6.67E-02 | 2.66E-03 | 5.02E-21 |
| | S2302 | 25 | 3 | 22 | 14 | 46742130 | 46908844 | 166.715 | 229 | 5.13E-06 | 4.00E-02 | 2.93E-03 | 3.44E-09 |
| | S2305 | 25 | 7 | 18 | 15 | 84946006 | 85497641 | 551.636 | 973 | 1.05E-36 | 9.33E-02 | 2.40E-03 | 7.01E-40 |
| | S2308 | 25 | 3 | 22 | 20 | 37063522 | 37263747 | 200.226 | 282 | 5.13E-06 | 4.00E-02 | 2.93E-03 | 3.44E-09 |
| | S2331 | 24 | 3 | 21 | 4 | 66844732 | 67033166 | 188.435 | 370 | 2.96E-06 | 4.00E-02 | 2.80E-03 | 1.99E-09 |
| | S2332 | 24 | 3 | 21 | 4 | 67192689 | 67325161 | 132.473 | 298 | 2.96E-06 | 4.00E-02 | 2.80E-03 | 1.99E-09 |
| | S2337 | 24 | 5 | 19 | 4 | 106121378 | 106377212 | 255.835 | 360 | 1.31E-18 | 6.67E-02 | 2.53E-03 | 8.80E-22 |
| | S2342 | 24 | 4 | 20 | 4 | 159954848 | 160105947 | 151.1 | 267 | 1.59E-11 | 5.33E-02 | 2.66E-03 | 1.07E-14 |
| | S2345 | 24 | 4 | 20 | 5 | 64176099 | 64439914 | 263.816 | 341 | 1.59E-11 | 5.33E-02 | 2.66E-03 | 1.07E-14 |
| | S2361 | 24 | 4 | 20 | 7 | 88659195 | 88881919 | 222.725 | 402 | 1.59E-11 | 5.33E-02 | 2.66E-03 | 1.07E-14 |
| | S2386 | 24 | 4 | 20 | 11 | 106098603 | 106381264 | 282.662 | 399 | 1.59E-11 | 5.33E-02 | 2.66E-03 | 1.07E-14 |
| | S2391 | 24 | 3 | 21 | 12 | 123582733 | 123773596 | 190.864 | 255 | 2.96E-06 | 4.00E-02 | 2.80E-03 | 1.99E-09 |
| | S2411 | 23 | 3 | 20 | 1 | 79194804 | 79428138 | 233.335 | 513 | 1.63E-06 | 4.00E-02 | 2.66E-03 | 1.09E-09 |

| | | | | | | | | | | | | | |
|---|---|---|---|---|---|---|---|---|---|---|---|---|---|
| | S2413 | 23 | 3 | 20 | 1 | 153240271 | 153357702 | 117.432 | 229 | 1.63E-06 | 4.00E-02 | 2.66E-03 | 1.09E-09 |
| | S2439 | 23 | 5 | 18 | 3 | 152349004 | 152562959 | 213.956 | 343 | 1.98E-19 | 6.67E-02 | 2.40E-03 | 1.33E-22 |
| | S2444 | 23 | 5 | 18 | 3 | 180804790 | 181161985 | 357.196 | 444 | 1.98E-19 | 6.67E-02 | 2.40E-03 | 1.33E-22 |
| | S2448 | 23 | 5 | 18 | 4 | 106595388 | 106696952 | 101.565 | 188 | 1.98E-19 | 6.67E-02 | 2.40E-03 | 1.33E-22 |
| | S2449 | 23 | 4 | 19 | 4 | 107712350 | 107830649 | 118.3 | 159 | 5.04E-12 | 5.33E-02 | 2.53E-03 | 3.38E-15 |
| | S2459 | 23 | 3 | 20 | 5 | 63407384 | 63589089 | 181.706 | 462 | 1.63E-06 | 4.00E-02 | 2.66E-03 | 1.09E-09 |
| | S2468 | 23 | 4 | 19 | 6 | 71358447 | 71667230 | 308.784 | 582 | 5.04E-12 | 5.33E-02 | 2.53E-03 | 3.38E-15 |
| | S2478 | 23 | 5 | 18 | 7 | 90001843 | 91005210 | 1003.368 | 2238 | 1.98E-19 | 6.67E-02 | 2.40E-03 | 1.33E-22 |
| | S2479 | 23 | 3 | 20 | 7 | 102769649 | 103149134 | 379.486 | 426 | 1.63E-06 | 4.00E-02 | 2.66E-03 | 1.09E-09 |
| | S2487 | 23 | 5 | 18 | 9 | 29388763 | 29511848 | 123.086 | 338 | 1.98E-19 | 6.67E-02 | 2.40E-03 | 1.33E-22 |
| | S2489 | 23 | 5 | 18 | 9 | 96519987 | 96842892 | 322.906 | 188 | 1.98E-19 | 6.67E-02 | 2.40E-03 | 1.33E-22 |
| | S2499 | 23 | 3 | 20 | 11 | 59339302 | 59855905 | 516.604 | 577 | 1.63E-06 | 4.00E-02 | 2.66E-03 | 1.09E-09 |
| | S2501 | 23 | 5 | 18 | 12 | 18153827 | 18355318 | 201.492 | 412 | 1.98E-19 | 6.67E-02 | 2.40E-03 | 1.33E-22 |
| | S2503 | 23 | 4 | 19 | 12 | 41615371 | 41820036 | 204.666 | 523 | 5.04E-12 | 5.33E-02 | 2.53E-03 | 3.38E-15 |

| | | | | | | | | | | | | | |
|---|---|---|---|---|---|---|---|---|---|---|---|---|---|
| | S2521 | 23 | 4 | 19 | 18 | 22650558 | 22800302 | 149.745 | 228 | 5.04E-12 | 5.33E-02 | 2.53E-03 | 3.38E-15 |
| | S2522 | 23 | 4 | 19 | 18 | 23494486 | 23725402 | 230.917 | 239 | 5.04E-12 | 5.33E-02 | 2.53E-03 | 3.38E-15 |
| | S2533 | 22 | 4 | 18 | 1 | 118756334 | 119181912 | 425.579 | 881 | 1.44E-12 | 5.33E-02 | 2.40E-03 | 9.63E-16 |
| | S2534 | 22 | 4 | 18 | 1 | 155061442 | 155464151 | 402.71 | 432 | 1.44E-12 | 5.33E-02 | 2.40E-03 | 9.63E-16 |
| | S2551 | 22 | 4 | 18 | 3 | 36064241 | 36313019 | 248.779 | 441 | 1.44E-12 | 5.33E-02 | 2.40E-03 | 9.63E-16 |
| | S2556 | 22 | 7 | 15 | 3 | 139405330 | 139577110 | 171.781 | 309 | 7.71E-42 | 9.33E-02 | 2.00E-03 | 5.17E-45 |
| | S2560 | 22 | 4 | 18 | 4 | 53010350 | 53351440 | 341.091 | 570 | 1.44E-12 | 5.33E-02 | 2.40E-03 | 9.63E-16 |
| | S2578 | 22 | 4 | 18 | 5 | 94036860 | 94209819 | 172.96 | 153 | 1.44E-12 | 5.33E-02 | 2.40E-03 | 9.63E-16 |
| | S2579 | 22 | 6 | 16 | 5 | 104033089 | 104163108 | 130.02 | 291 | 4.44E-30 | 8.00E-02 | 2.13E-03 | 2.98E-33 |
| | S2580 | 22 | 6 | 16 | 5 | 114916898 | 115105269 | 188.372 | 346 | 4.44E-30 | 8.00E-02 | 2.13E-03 | 2.98E-33 |
| | S2582 | 22 | 4 | 18 | 6 | 71744101 | 71857261 | 113.161 | 310 | 1.44E-12 | 5.33E-02 | 2.40E-03 | 9.63E-16 |
| | S2593 | 22 | 5 | 17 | 9 | 94129286 | 94646689 | 517.404 | 1087 | 2.52E-20 | 6.67E-02 | 2.27E-03 | 1.69E-23 |
| | S2611 | 22 | 3 | 19 | 13 | 53482298 | 53615104 | 132.807 | 243 | 8.51E-07 | 4.00E-02 | 2.53E-03 | 5.71E-10 |
| | S2623 | 22 | 3 | 19 | 20 | 24730591 | 25039695 | 309.105 | 639 | 8.51E-07 | 4.00E-02 | 2.53E-03 | 5.71E-10 |

| | | | | | | | | | | | | | |
|---|---|---|---|---|---|---|---|---|---|---|---|---|---|
| | S2628 | 21 | 6 | 15 | 1 | 94921350 | 95089873 | 168.524 | 438 | 1.54E-31 | 8.00E-02 | 2.00E-03 | 1.03E-34 |
| | S2630 | 21 | 3 | 18 | 1 | 186052504 | 186179685 | 127.182 | 189 | 4.18E-07 | 4.00E-02 | 2.40E-03 | 2.80E-10 |
| | S2643 | 21 | 4 | 17 | 2 | 166340163 | 166498891 | 158.729 | 302 | 3.64E-13 | 5.33E-02 | 2.27E-03 | 2.44E-16 |
| | S2650 | 21 | 4 | 17 | 3 | 120551674 | 120659251 | 107.578 | 219 | 3.64E-13 | 5.33E-02 | 2.27E-03 | 2.44E-16 |
| | S2654 | 21 | 4 | 17 | 3 | 155360686 | 155879249 | 518.564 | 815 | 3.64E-13 | 5.33E-02 | 2.27E-03 | 2.44E-16 |
| | S2663 | 21 | 3 | 18 | 4 | 104009571 | 104303404 | 293.834 | 406 | 4.18E-07 | 4.00E-02 | 2.40E-03 | 2.80E-10 |
| | S2674 | 21 | 5 | 16 | 5 | 62230173 | 62704338 | 474.166 | 731 | 2.64E-21 | 6.67E-02 | 2.13E-03 | 1.77E-24 |
| | S2675 | 21 | 4 | 17 | 5 | 64771729 | 65093506 | 321.778 | 439 | 3.64E-13 | 5.33E-02 | 2.27E-03 | 2.44E-16 |
| | S2679 | 21 | 4 | 17 | 5 | 92788688 | 92914109 | 125.422 | 162 | 3.64E-13 | 5.33E-02 | 2.27E-03 | 2.44E-16 |
| | S2683 | 21 | 3 | 18 | 6 | 66695187 | 66845333 | 150.147 | 444 | 4.18E-07 | 4.00E-02 | 2.40E-03 | 2.80E-10 |
| | S2698 | 21 | 3 | 18 | 7 | 53846990 | 54420876 | 573.887 | 1172 | 4.18E-07 | 4.00E-02 | 2.40E-03 | 2.80E-10 |
| | S2699 | 21 | 3 | 18 | 7 | 54442337 | 54556182 | 113.846 | 263 | 4.18E-07 | 4.00E-02 | 2.40E-03 | 2.80E-10 |
| | S2726 | 21 | 4 | 17 | 11 | 104173698 | 104319001 | 145.304 | 274 | 3.64E-13 | 5.33E-02 | 2.27E-03 | 2.44E-16 |
| | S2728 | 21 | 4 | 17 | 12 | 113172684 | 113337273 | 164.59 | 205 | 3.64E-13 | 5.33E-02 | 2.27E-03 | 2.44E-16 |

| | | | | | | | | | | | | | |
|---|---|---|---|---|---|---|---|---|---|---|---|---|---|
| | S2730 | 21 | 3 | 18 | 13 | 51851960 | 52727001 | 875.042 | 797 | 4.18E-07 | 4.00E-02 | 2.40E-03 | 2.80E-10 |
| | S2733 | 21 | 3 | 18 | 14 | 48258630 | 48514048 | 255.419 | 395 | 4.18E-07 | 4.00E-02 | 2.40E-03 | 2.80E-10 |
| | S2750 | 20 | 5 | 15 | 1 | 69325182 | 69577300 | 252.119 | 475 | 2.20E-22 | 6.67E-02 | 2.00E-03 | 1.48E-25 |
| | S2757 | 20 | 5 | 15 | 1 | 179090815 | 179502350 | 411.536 | 1109 | 2.20E-22 | 6.67E-02 | 2.00E-03 | 1.48E-25 |
| | S2769 | 20 | 3 | 17 | 2 | 74088950 | 74200888 | 111.939 | 162 | 1.91E-07 | 4.00E-02 | 2.27E-03 | 1.28E-10 |
| | S2770 | 20 | 3 | 17 | 2 | 74206825 | 74492875 | 286.051 | 245 | 1.91E-07 | 4.00E-02 | 2.27E-03 | 1.28E-10 |
| | S2776 | 20 | 3 | 17 | 2 | 109156091 | 109333040 | 176.95 | 287 | 1.91E-07 | 4.00E-02 | 2.27E-03 | 1.28E-10 |
| | S2786 | 20 | 3 | 17 | 3 | 156019099 | 156180200 | 161.102 | 217 | 1.91E-07 | 4.00E-02 | 2.27E-03 | 1.28E-10 |
| | S2792 | 20 | 3 | 17 | 4 | 63117518 | 63239728 | 122.211 | 359 | 1.91E-07 | 4.00E-02 | 2.27E-03 | 1.28E-10 |
| | S2797 | 20 | 3 | 17 | 4 | 103076526 | 103461086 | 384.561 | 517 | 1.91E-07 | 4.00E-02 | 2.27E-03 | 1.28E-10 |
| | S2798 | 20 | 5 | 15 | 4 | 108215568 | 108628351 | 412.784 | 690 | 2.20E-22 | 6.67E-02 | 2.00E-03 | 1.48E-25 |
| | S2814 | 20 | 6 | 14 | 5 | 120723494 | 121037190 | 313.697 | 610 | 3.81E-33 | 8.00E-02 | 1.87E-03 | 2.56E-36 |
| | S2834 | 20 | 6 | 14 | 7 | 116675151 | 116807803 | 132.653 | 141 | 3.81E-33 | 8.00E-02 | 1.87E-03 | 2.56E-36 |
| | S2841 | 20 | 5 | 15 | 9 | 97417559 | 97519530 | 101.972 | 116 | 2.20E-22 | 6.67E-02 | 2.00E-03 | 1.48E-25 |

| | | | | | | | | | | | | | |
|---|---|---|---|---|---|---|---|---|---|---|---|---|---|
| | S2844 | 20 | 4 | 16 | 10 | 87024733 | 87139423 | 114.691 | 198 | 8.03E-14 | 5.33E-02 | 2.13E-03 | 5.38E-17 |
| | S2854 | 20 | 5 | 15 | 11 | 81943048 | 82276048 | 333.001 | 754 | 2.20E-22 | 6.67E-02 | 2.00E-03 | 1.48E-25 |
| | S2855 | 20 | 3 | 17 | 11 | 83322955 | 83767909 | 444.955 | 674 | 1.91E-07 | 4.00E-02 | 2.27E-03 | 1.28E-10 |
| | S2878 | 20 | 5 | 15 | 20 | 38504150 | 39239740 | 735.591 | 1173 | 2.20E-22 | 6.67E-02 | 2.00E-03 | 1.48E-25 |
| | S2888 | 19 | 3 | 16 | 1 | 186881247 | 186999579 | 118.333 | 305 | 8.07E-08 | 4.00E-02 | 2.13E-03 | 5.41E-11 |
| | S2892 | 19 | 3 | 16 | 2 | 40045179 | 40199527 | 154.349 | 418 | 8.07E-08 | 4.00E-02 | 2.13E-03 | 5.41E-11 |
| | S2894 | 19 | 3 | 16 | 2 | 51800133 | 51960015 | 159.883 | 341 | 8.07E-08 | 4.00E-02 | 2.13E-03 | 5.41E-11 |
| | S2895 | 19 | 5 | 14 | 2 | 84608215 | 84912339 | 304.125 | 406 | 1.42E-23 | 6.67E-02 | 1.87E-03 | 9.51E-27 |
| | S2904 | 19 | 5 | 14 | 3 | 146215990 | 146366672 | 150.683 | 232 | 1.42E-23 | 6.67E-02 | 1.87E-03 | 9.51E-27 |
| | S2907 | 19 | 3 | 16 | 4 | 72020257 | 72135818 | 115.562 | 107 | 8.07E-08 | 4.00E-02 | 2.13E-03 | 5.41E-11 |
| | S2908 | 19 | 3 | 16 | 4 | 72989406 | 73124906 | 135.501 | 112 | 8.07E-08 | 4.00E-02 | 2.13E-03 | 5.41E-11 |
| | S2909 | 19 | 3 | 16 | 4 | 79023203 | 79273570 | 250.368 | 351 | 8.07E-08 | 4.00E-02 | 2.13E-03 | 5.41E-11 |
| | S2910 | 19 | 3 | 16 | 4 | 79275415 | 79408348 | 132.934 | 156 | 8.07E-08 | 4.00E-02 | 2.13E-03 | 5.41E-11 |
| | S2911 | 19 | 3 | 16 | 4 | 79640216 | 79753497 | 113.282 | 263 | 8.07E-08 | 4.00E-02 | 2.13E-03 | 5.41E-11 |

| | | | | | | | | | | | | | |
|---|---|---|---|---|---|---|---|---|---|---|---|---|---|
| | S2912 | 19 | 3 | 16 | 4 | 79896824 | 80058292 | 161.469 | 295 | 8.07E-08 | 4.00E-02 | 2.13E-03 | 5.41E-11 |
| | S2915 | 19 | 3 | 16 | 4 | 102299244 | 102467856 | 168.613 | 321 | 8.07E-08 | 4.00E-02 | 2.13E-03 | 5.41E-11 |
| | S2919 | 19 | 3 | 16 | 4 | 120862858 | 121050058 | 187.201 | 405 | 8.07E-08 | 4.00E-02 | 2.13E-03 | 5.41E-11 |
| | S2920 | 19 | 4 | 15 | 4 | 122203821 | 122337115 | 133.295 | 153 | 1.51E-14 | 5.33E-02 | 2.00E-03 | 1.02E-17 |
| | S2921 | 19 | 4 | 15 | 4 | 122362008 | 122684588 | 322.581 | 557 | 1.51E-14 | 5.33E-02 | 2.00E-03 | 1.02E-17 |
| | S2928 | 19 | 4 | 15 | 4 | 160668218 | 160879898 | 211.681 | 521 | 1.51E-14 | 5.33E-02 | 2.00E-03 | 1.02E-17 |
| | S2938 | 19 | 5 | 14 | 5 | 94465772 | 94860050 | 394.279 | 592 | 1.42E-23 | 6.67E-02 | 1.87E-03 | 9.51E-27 |
| | S2941 | 19 | 6 | 13 | 5 | 103842705 | 103979003 | 136.299 | 270 | 6.41E-35 | 8.00E-02 | 1.73E-03 | 4.30E-38 |
| | S2960 | 19 | 4 | 15 | 7 | 113297046 | 113698834 | 401.789 | 580 | 1.51E-14 | 5.33E-02 | 2.00E-03 | 1.02E-17 |
| | S2965 | 19 | 3 | 16 | 8 | 74532571 | 74663290 | 130.72 | 218 | 8.07E-08 | 4.00E-02 | 2.13E-03 | 5.41E-11 |
| | S2969 | 19 | 5 | 14 | 9 | 103206390 | 103481476 | 275.087 | 457 | 1.42E-23 | 6.67E-02 | 1.87E-03 | 9.51E-27 |
| | S2975 | 19 | 4 | 15 | 10 | 87393376 | 87709196 | 315.821 | 433 | 1.51E-14 | 5.33E-02 | 2.00E-03 | 1.02E-17 |
| | S2979 | 19 | 6 | 13 | 10 | 110444589 | 110589446 | 144.858 | 235 | 6.41E-35 | 8.00E-02 | 1.73E-03 | 4.30E-38 |
| | S2983 | 19 | 4 | 15 | 11 | 63987039 | 64108085 | 121.047 | 177 | 1.51E-14 | 5.33E-02 | 2.00E-03 | 1.02E-17 |

| | | | | | | | | | | | | | |
|---|---|---|---|---|---|---|---|---|---|---|---|---|---|
| | S2995 | 19 | 5 | 14 | 13 | 90027560 | 90164304 | 136.745 | 433 | 1.42E-23 | 6.67E-02 | 1.87E-03 | 9.51E-27 |
| | S2999 | 19 | 4 | 15 | 14 | 43208865 | 43440539 | 231.675 | 548 | 1.51E-14 | 5.33E-02 | 2.00E-03 | 1.02E-17 |
| | S3011 | 19 | 3 | 16 | 20 | 21570020 | 22519928 | 949.909 | 1213 | 8.07E-08 | 4.00E-02 | 2.13E-03 | 5.41E-11 |
| | S3015 | 18 | 4 | 14 | 1 | 88424691 | 89054547 | 629.857 | 1044 | 2.37E-15 | 5.33E-02 | 1.87E-03 | 1.59E-18 |
| | S3021 | 18 | 3 | 15 | 1 | 153455093 | 153583772 | 128.68 | 206 | 3.10E-08 | 4.00E-02 | 2.00E-03 | 2.08E-11 |
| | S3041 | 18 | 3 | 15 | 3 | 37699926 | 37905377 | 205.452 | 277 | 3.10E-08 | 4.00E-02 | 2.00E-03 | 2.08E-11 |
| | S3050 | 18 | 3 | 15 | 4 | 77791086 | 77960750 | 169.665 | 226 | 3.10E-08 | 4.00E-02 | 2.00E-03 | 2.08E-11 |
| | S3053 | 18 | 4 | 14 | 4 | 101802162 | 102103230 | 301.069 | 444 | 2.37E-15 | 5.33E-02 | 1.87E-03 | 1.59E-18 |
| | S3058 | 18 | 4 | 14 | 4 | 123892694 | 124489142 | 596.449 | 1049 | 2.37E-15 | 5.33E-02 | 1.87E-03 | 1.59E-18 |
| | S3059 | 18 | 4 | 14 | 4 | 129270221 | 129410251 | 140.031 | 267 | 2.37E-15 | 5.33E-02 | 1.87E-03 | 1.59E-18 |
| | S3065 | 18 | 3 | 15 | 5 | 36470689 | 36595701 | 125.013 | 266 | 3.10E-08 | 4.00E-02 | 2.00E-03 | 2.08E-11 |
| | S3068 | 18 | 6 | 12 | 5 | 113485440 | 113767881 | 282.442 | 766 | 6.85E-37 | 8.00E-02 | 1.60E-03 | 4.59E-40 |
| | S3081 | 18 | 5 | 13 | 7 | 116386431 | 116522675 | 136.245 | 251 | 6.74E-25 | 6.67E-02 | 1.73E-03 | 4.52E-28 |
| | S3086 | 18 | 3 | 15 | 8 | 93417861 | 93603344 | 185.484 | 223 | 3.10E-08 | 4.00E-02 | 2.00E-03 | 2.08E-11 |

| | | | | | | | | | | | | | |
|---|---|---|---|---|---|---|---|---|---|---|---|---|---|
| | S3094 | 18 | 5 | 13 | 9 | 97854739 | 98082725 | 227.987 | 352 | 6.74E-25 | 6.67E-02 | 1.73E-03 | 4.52E-28 |
| | S3104 | 18 | 3 | 15 | 11 | 43031345 | 43254070 | 222.726 | 652 | 3.10E-08 | 4.00E-02 | 2.00E-03 | 2.08E-11 |
| | S3110 | 18 | 3 | 15 | 12 | 49348706 | 49543609 | 194.904 | 237 | 3.10E-08 | 4.00E-02 | 2.00E-03 | 2.08E-11 |
| | S3113 | 18 | 3 | 15 | 13 | 51524383 | 51626868 | 102.486 | 100 | 3.10E-08 | 4.00E-02 | 2.00E-03 | 2.08E-11 |
| | S3115 | 18 | 3 | 15 | 13 | 80375230 | 80586775 | 211.546 | 357 | 3.10E-08 | 4.00E-02 | 2.00E-03 | 2.08E-11 |
| | S3117 | 18 | 3 | 15 | 13 | 85579280 | 85738495 | 159.216 | 343 | 3.10E-08 | 4.00E-02 | 2.00E-03 | 2.08E-11 |
| | S3121 | 18 | 5 | 13 | 16 | 14382126 | 14861506 | 479.381 | 301 | 6.74E-25 | 6.67E-02 | 1.73E-03 | 4.52E-28 |
| | S3122 | 18 | 5 | 13 | 16 | 15304078 | 15554459 | 250.382 | 259 | 6.74E-25 | 6.67E-02 | 1.73E-03 | 4.52E-28 |
| | S3138 | 17 | 7 | 10 | 1 | 98266257 | 98474702 | 208.446 | 423 | 2.08E-54 | 9.33E-02 | 1.33E-03 | 1.39E-57 |
| | S3144 | 17 | 3 | 14 | 2 | 19889650 | 20178444 | 288.795 | 564 | 1.06E-08 | 4.00E-02 | 1.87E-03 | 7.13E-12 |
| | S3146 | 17 | 4 | 13 | 2 | 35888056 | 36141118 | 253.063 | 612 | 3.00E-16 | 5.33E-02 | 1.73E-03 | 2.01E-19 |
| | S3152 | 17 | 3 | 14 | 2 | 106142889 | 106337831 | 194.943 | 335 | 1.06E-08 | 4.00E-02 | 1.87E-03 | 7.13E-12 |
| | S3153 | 17 | 3 | 14 | 2 | 106520001 | 106943651 | 423.651 | 925 | 1.06E-08 | 4.00E-02 | 1.87E-03 | 7.13E-12 |
| | S3163 | 17 | 3 | 14 | 2 | 181324823 | 181446867 | 122.045 | 189 | 1.06E-08 | 4.00E-02 | 1.87E-03 | 7.13E-12 |

| | | | | | | | | | | | | | |
|---|---|---|---|---|---|---|---|---|---|---|---|---|---|
| | S3166 | 17 | 4 | 13 | 3 | 36817092 | 37404889 | 587.798 | 667 | 3.00E-16 | 5.33E-02 | 1.73E-03 | 2.01E-19 |
| | S3167 | 17 | 4 | 13 | 3 | 37418996 | 37569129 | 150.134 | 288 | 3.00E-16 | 5.33E-02 | 1.73E-03 | 2.01E-19 |
| | S3168 | 17 | 4 | 13 | 3 | 39378730 | 39561173 | 182.444 | 286 | 3.00E-16 | 5.33E-02 | 1.73E-03 | 2.01E-19 |
| | S3169 | 17 | 5 | 12 | 3 | 39932537 | 40150764 | 218.228 | 330 | 2.24E-26 | 6.67E-02 | 1.60E-03 | 1.50E-29 |
| | S3174 | 17 | 3 | 14 | 3 | 111175609 | 111535059 | 359.451 | 599 | 1.06E-08 | 4.00E-02 | 1.87E-03 | 7.13E-12 |
| | S3179 | 17 | 3 | 14 | 3 | 130583509 | 130789857 | 206.349 | 228 | 1.06E-08 | 4.00E-02 | 1.87E-03 | 7.13E-12 |
| | S3183 | 17 | 4 | 13 | 4 | 62816824 | 62929036 | 112.213 | 188 | 3.00E-16 | 5.33E-02 | 1.73E-03 | 2.01E-19 |
| | S3200 | 17 | 3 | 14 | 4 | 156695073 | 156890211 | 195.139 | 309 | 1.06E-08 | 4.00E-02 | 1.87E-03 | 7.13E-12 |
| | S3204 | 17 | 4 | 13 | 5 | 58044929 | 58254966 | 210.038 | 569 | 3.00E-16 | 5.33E-02 | 1.73E-03 | 2.01E-19 |
| | S3219 | 17 | 4 | 13 | 7 | 52956080 | 53094947 | 138.868 | 372 | 3.00E-16 | 5.33E-02 | 1.73E-03 | 2.01E-19 |
| | S3222 | 17 | 4 | 13 | 7 | 103569959 | 103718024 | 148.066 | 267 | 3.00E-16 | 5.33E-02 | 1.73E-03 | 2.01E-19 |
| | S3228 | 17 | 4 | 13 | 9 | 73871927 | 74111519 | 239.593 | 384 | 3.00E-16 | 5.33E-02 | 1.73E-03 | 2.01E-19 |
| | S3230 | 17 | 5 | 12 | 9 | 111073708 | 111928034 | 854.327 | 1828 | 2.24E-26 | 6.67E-02 | 1.60E-03 | 1.50E-29 |
| | S3257 | 17 | 5 | 12 | 13 | 83745085 | 83895499 | 150.415 | 113 | 2.24E-26 | 6.67E-02 | 1.60E-03 | 1.50E-29 |

| | | | | | | | | | | | | | |
|---|---|---|---|---|---|---|---|---|---|---|---|---|---|
| | S3262 | 17 | 3 | 14 | 13 | 85163328 | 85279952 | 116.625 | 264 | 1.06E-08 | 4.00E-02 | 1.87E-03 | 7.13E-12 |
| | S3277 | 16 | 6 | 10 | 1 | 99883099 | 100255291 | 372.193 | 758 | 1.43E-41 | 8.00E-02 | 1.33E-03 | 9.61E-45 |
| | S3279 | 16 | 5 | 11 | 1 | 100361534 | 100560437 | 198.904 | 273 | 4.88E-28 | 6.67E-02 | 1.47E-03 | 3.27E-31 |
| | S3282 | 16 | 3 | 13 | 1 | 153809535 | 154158721 | 349.187 | 478 | 3.20E-09 | 4.00E-02 | 1.73E-03 | 2.15E-12 |
| | S3283 | 16 | 3 | 13 | 1 | 154576516 | 154683482 | 106.967 | 179 | 3.20E-09 | 4.00E-02 | 1.73E-03 | 2.15E-12 |
| | S3299 | 16 | 3 | 13 | 2 | 71490543 | 71667696 | 177.154 | 332 | 3.20E-09 | 4.00E-02 | 1.73E-03 | 2.15E-12 |
| | S3306 | 16 | 3 | 13 | 2 | 181537230 | 181924539 | 387.31 | 427 | 3.20E-09 | 4.00E-02 | 1.73E-03 | 2.15E-12 |
| | S3307 | 16 | 3 | 13 | 2 | 181996698 | 182300974 | 304.277 | 490 | 3.20E-09 | 4.00E-02 | 1.73E-03 | 2.15E-12 |
| | S3321 | 16 | 3 | 13 | 3 | 110369920 | 110581278 | 211.359 | 356 | 3.20E-09 | 4.00E-02 | 1.73E-03 | 2.15E-12 |
| | S3322 | 16 | 3 | 13 | 3 | 110802487 | 110952118 | 149.632 | 139 | 3.20E-09 | 4.00E-02 | 1.73E-03 | 2.15E-12 |
| | S3330 | 16 | 3 | 13 | 4 | 75905987 | 76402625 | 496.639 | 974 | 3.20E-09 | 4.00E-02 | 1.73E-03 | 2.15E-12 |
| | S3334 | 16 | 4 | 12 | 4 | 123205795 | 123317378 | 111.584 | 154 | 2.93E-17 | 5.33E-02 | 1.60E-03 | 1.96E-20 |
| | S3335 | 16 | 4 | 12 | 4 | 124746308 | 124943442 | 197.135 | 485 | 2.93E-17 | 5.33E-02 | 1.60E-03 | 1.96E-20 |
| | S3337 | 16 | 4 | 12 | 4 | 156031242 | 156229420 | 198.179 | 458 | 2.93E-17 | 5.33E-02 | 1.60E-03 | 1.96E-20 |

| | | | | | | | | | | | | | |
|---|---|---|---|---|---|---|---|---|---|---|---|---|---|
| | S3353 | 16 | 4 | 12 | 7 | 103755459 | 103893018 | 137.56 | 337 | 2.93E-17 | 5.33E-02 | 1.60E-03 | 1.96E-20 |
| | S3354 | 16 | 4 | 12 | 7 | 103951375 | 104082720 | 131.346 | 226 | 2.93E-17 | 5.33E-02 | 1.60E-03 | 1.96E-20 |
| | S3359 | 16 | 4 | 12 | 7 | 113760374 | 113960428 | 200.055 | 234 | 2.93E-17 | 5.33E-02 | 1.60E-03 | 1.96E-20 |
| | S3368 | 16 | 4 | 12 | 9 | 95446162 | 95554049 | 107.888 | 157 | 2.93E-17 | 5.33E-02 | 1.60E-03 | 1.96E-20 |
| | S3370 | 16 | 6 | 10 | 9 | 108879247 | 109071512 | 192.266 | 421 | 1.43E-41 | 8.00E-02 | 1.33E-03 | 9.61E-45 |
| | S3380 | 16 | 6 | 10 | 10 | 100833866 | 100999999 | 166.134 | 247 | 1.43E-41 | 8.00E-02 | 1.33E-03 | 9.61E-45 |
| | S3389 | 16 | 5 | 11 | 12 | 31628444 | 31745841 | 117.398 | 202 | 4.88E-28 | 6.67E-02 | 1.47E-03 | 3.27E-31 |
| | S3390 | 16 | 3 | 13 | 12 | 48390156 | 48566275 | 176.12 | 460 | 3.20E-09 | 4.00E-02 | 1.73E-03 | 2.15E-12 |
| | S3413 | 16 | 4 | 12 | 20 | 37614458 | 37932599 | 318.142 | 470 | 2.93E-17 | 5.33E-02 | 1.60E-03 | 1.96E-20 |
| | S3419 | 15 | 5 | 10 | 1 | 90760037 | 91024576 | 264.54 | 417 | 6.39E-30 | 6.67E-02 | 1.33E-03 | 4.28E-33 |
| | S3423 | 15 | 5 | 10 | 1 | 111863560 | 111992930 | 129.371 | 296 | 6.39E-30 | 6.67E-02 | 1.33E-03 | 4.28E-33 |
| | S3432 | 15 | 3 | 12 | 1 | 242651579 | 243624192 | 972.614 | 1472 | 8.22E-10 | 4.00E-02 | 1.60E-03 | 5.52E-13 |
| | S3438 | 15 | 3 | 12 | 2 | 40399996 | 40841566 | 441.571 | 770 | 8.22E-10 | 4.00E-02 | 1.60E-03 | 5.52E-13 |
| | S3443 | 15 | 4 | 11 | 2 | 76427435 | 76694126 | 266.692 | 646 | 2.10E-18 | 5.33E-02 | 1.47E-03 | 1.41E-21 |

| | | | | | | | | | | | | | |
|---|---|---|---|---|---|---|---|---|---|---|---|---|---|
| | S3444 | 15 | 4 | 11 | 2 | 85290783 | 85447453 | 156.671 | 305 | 2.10E-18 | 5.33E-02 | 1.47E-03 | 1.41E-21 |
| | S3445 | 15 | 3 | 12 | 2 | 85838133 | 86096216 | 258.084 | 412 | 8.22E-10 | 4.00E-02 | 1.60E-03 | 5.52E-13 |
| | S3446 | 15 | 3 | 12 | 2 | 86332262 | 86438385 | 106.124 | 154 | 8.22E-10 | 4.00E-02 | 1.60E-03 | 5.52E-13 |
| | S3454 | 15 | 3 | 12 | 3 | 22533607 | 23728406 | 1194.8 | 2487 | 8.22E-10 | 4.00E-02 | 1.60E-03 | 5.52E-13 |
| | S3457 | 15 | 4 | 11 | 3 | 40194610 | 40669866 | 475.257 | 501 | 2.10E-18 | 5.33E-02 | 1.47E-03 | 1.41E-21 |
| | S3462 | 15 | 6 | 9 | 3 | 134919546 | 135037148 | 117.603 | 244 | 2.24E-44 | 8.00E-02 | 1.20E-03 | 1.50E-47 |
| | S3465 | 15 | 5 | 10 | 3 | 140255544 | 140393506 | 137.963 | 213 | 6.39E-30 | 6.67E-02 | 1.33E-03 | 4.28E-33 |
| | S3467 | 15 | 5 | 10 | 3 | 141877041 | 142051807 | 174.767 | 361 | 6.39E-30 | 6.67E-02 | 1.33E-03 | 4.28E-33 |
| | S3468 | 15 | 5 | 10 | 3 | 144193170 | 144800114 | 606.945 | 887 | 6.39E-30 | 6.67E-02 | 1.33E-03 | 4.28E-33 |
| | S3469 | 15 | 5 | 10 | 3 | 144937030 | 145178697 | 241.668 | 394 | 6.39E-30 | 6.67E-02 | 1.33E-03 | 4.28E-33 |
| | S3470 | 15 | 5 | 10 | 3 | 178620339 | 178843865 | 223.527 | 499 | 6.39E-30 | 6.67E-02 | 1.33E-03 | 4.28E-33 |
| | S3475 | 15 | 5 | 10 | 4 | 99414429 | 99721371 | 306.943 | 521 | 6.39E-30 | 6.67E-02 | 1.33E-03 | 4.28E-33 |
| | S3476 | 15 | 5 | 10 | 4 | 100084122 | 100231322 | 147.201 | 310 | 6.39E-30 | 6.67E-02 | 1.33E-03 | 4.28E-33 |
| | S3477 | 15 | 4 | 11 | 4 | 101398129 | 101602384 | 204.256 | 283 | 2.10E-18 | 5.33E-02 | 1.47E-03 | 1.41E-21 |

| | | | | | | | | | | | | | |
|---|---|---|---|---|---|---|---|---|---|---|---|---|---|
| | S3478 | 15 | 4 | 11 | 4 | 101623076 | 101762340 | 139.265 | 325 | 2.10E-18 | 5.33E-02 | 1.47E-03 | 1.41E-21 |
| | S3486 | 15 | 5 | 10 | 5 | 107971620 | 108258657 | 287.038 | 597 | 6.39E-30 | 6.67E-02 | 1.33E-03 | 4.28E-33 |
| | S3487 | 15 | 5 | 10 | 5 | 108261691 | 109123256 | 861.566 | 1775 | 6.39E-30 | 6.67E-02 | 1.33E-03 | 4.28E-33 |
| | S3488 | 15 | 6 | 9 | 5 | 109553994 | 109897792 | 343.799 | 793 | 2.24E-44 | 8.00E-02 | 1.20E-03 | 1.50E-47 |
| | S3489 | 15 | 5 | 10 | 5 | 109899863 | 110016297 | 116.435 | 170 | 6.39E-30 | 6.67E-02 | 1.33E-03 | 4.28E-33 |
| | S3490 | 15 | 6 | 9 | 5 | 110727539 | 111055212 | 327.674 | 428 | 2.24E-44 | 8.00E-02 | 1.20E-03 | 1.50E-47 |
| | S3492 | 15 | 5 | 10 | 5 | 112201871 | 112637844 | 435.974 | 906 | 6.39E-30 | 6.67E-02 | 1.33E-03 | 4.28E-33 |
| | S3495 | 15 | 5 | 10 | 5 | 116669616 | 116786594 | 116.979 | 218 | 6.39E-30 | 6.67E-02 | 1.33E-03 | 4.28E-33 |
| | S3509 | 15 | 4 | 11 | 7 | 52724462 | 52880113 | 155.652 | 431 | 2.10E-18 | 5.33E-02 | 1.47E-03 | 1.41E-21 |
| | S3512 | 15 | 4 | 11 | 7 | 68438460 | 68558144 | 119.685 | 260 | 2.10E-18 | 5.33E-02 | 1.47E-03 | 1.41E-21 |
| | S3522 | 15 | 4 | 11 | 9 | 33187899 | 33318590 | 130.692 | 178 | 2.10E-18 | 5.33E-02 | 1.47E-03 | 1.41E-21 |
| | S3527 | 15 | 5 | 10 | 9 | 102590939 | 102797485 | 206.547 | 595 | 6.39E-30 | 6.67E-02 | 1.33E-03 | 4.28E-33 |
| | S3528 | 15 | 5 | 10 | 9 | 109282483 | 109419417 | 136.935 | 232 | 6.39E-30 | 6.67E-02 | 1.33E-03 | 4.28E-33 |
| | S3533 | 15 | 7 | 8 | 10 | 106395543 | 106607970 | 212.428 | 325 | 8.87E-62 | 9.33E-02 | 1.07E-03 | 5.95E-65 |

| | | | | | | | | | | | | | |
|---|---|---|---|---|---|---|---|---|---|---|---|---|---|
| | S3534 | 15 | 4 | 11 | 10 | 114971070 | 115134555 | 163.486 | 176 | 2.10E-18 | 5.33E-02 | 1.47E-03 | 1.41E-21 |
| | S3535 | 15 | 5 | 10 | 11 | 26153726 | 26653514 | 499.789 | 1020 | 6.39E-30 | 6.67E-02 | 1.33E-03 | 4.28E-33 |
| | S3536 | 15 | 3 | 12 | 11 | 40853581 | 41188057 | 334.477 | 567 | 8.22E-10 | 4.00E-02 | 1.60E-03 | 5.52E-13 |
| | S3537 | 15 | 3 | 12 | 11 | 41393487 | 41701745 | 308.259 | 602 | 8.22E-10 | 4.00E-02 | 1.60E-03 | 5.52E-13 |
| | S3546 | 15 | 3 | 12 | 12 | 30775783 | 30919706 | 143.924 | 324 | 8.22E-10 | 4.00E-02 | 1.60E-03 | 5.52E-13 |
| | S3548 | 15 | 4 | 11 | 12 | 46545731 | 46736257 | 190.527 | 246 | 2.10E-18 | 5.33E-02 | 1.47E-03 | 1.41E-21 |
| | S3554 | 15 | 3 | 12 | 13 | 94082159 | 94199614 | 117.456 | 227 | 8.22E-10 | 4.00E-02 | 1.60E-03 | 5.52E-13 |
| | S3555 | 15 | 4 | 11 | 14 | 48886762 | 49111687 | 224.926 | 370 | 2.10E-18 | 5.33E-02 | 1.47E-03 | 1.41E-21 |
| | S3556 | 15 | 4 | 11 | 14 | 49374173 | 49725908 | 351.736 | 439 | 2.10E-18 | 5.33E-02 | 1.47E-03 | 1.41E-21 |
| | S3559 | 15 | 3 | 12 | 14 | 61958980 | 62176938 | 217.959 | 487 | 8.22E-10 | 4.00E-02 | 1.60E-03 | 5.52E-13 |
| | S3560 | 15 | 3 | 12 | 14 | 71183965 | 71624711 | 440.747 | 540 | 8.22E-10 | 4.00E-02 | 1.60E-03 | 5.52E-13 |
| | S3565 | 15 | 4 | 11 | 15 | 45143019 | 45253280 | 110.262 | 160 | 2.10E-18 | 5.33E-02 | 1.47E-03 | 1.41E-21 |
| | S3566 | 15 | 3 | 12 | 16 | 20406378 | 20593795 | 187.418 | 189 | 8.22E-10 | 4.00E-02 | 1.60E-03 | 5.52E-13 |
| | S3567 | 15 | 9 | 6 | 16 | 49626115 | 49842961 | 216.847 | 421 | 1.95E-105 | 1.20E-01 | 7.99E-04 | 1.31E-108 |

| | | | | | | | | | | | | | |
|---|---|---|---|---|---|---|---|---|---|---|---|---|---|
| | S3568 | 15 | 9 | 6 | 16 | 49920928 | 50053087 | 132.16 | 221 | 1.95E-105 | 1.20E-01 | 7.99E-04 | 1.31E-108 |
| | S3569 | 15 | 3 | 12 | 16 | 74896352 | 75123124 | 226.773 | 403 | 8.22E-10 | 4.00E-02 | 1.60E-03 | 5.52E-13 |
| | S3581 | 15 | 3 | 12 | 20 | 40669156 | 40811614 | 142.459 | 372 | 8.22E-10 | 4.00E-02 | 1.60E-03 | 5.52E-13 |
| | S3586 | 14 | 5 | 9 | 1 | 108368891 | 108651391 | 282.501 | 295 | 4.51E-32 | 6.67E-02 | 1.20E-03 | 3.02E-35 |
| | S3587 | 14 | 4 | 10 | 1 | 112474673 | 112644156 | 169.484 | 324 | 1.04E-19 | 5.33E-02 | 1.33E-03 | 6.95E-23 |
| | S3588 | 14 | 4 | 10 | 1 | 112683940 | 112868110 | 184.171 | 314 | 1.04E-19 | 5.33E-02 | 1.33E-03 | 6.95E-23 |
| | S3589 | 14 | 4 | 10 | 1 | 112985572 | 113194234 | 208.663 | 310 | 1.04E-19 | 5.33E-02 | 1.33E-03 | 6.95E-23 |
| | S3590 | 14 | 5 | 9 | 1 | 113271452 | 113492013 | 220.562 | 406 | 4.51E-32 | 6.67E-02 | 1.20E-03 | 3.02E-35 |
| | S3591 | 14 | 4 | 10 | 1 | 117024431 | 117558213 | 533.783 | 1108 | 1.04E-19 | 5.33E-02 | 1.33E-03 | 6.95E-23 |
| | S3604 | 14 | 3 | 11 | 2 | 105640761 | 105903778 | 263.018 | 565 | 1.74E-10 | 4.00E-02 | 1.47E-03 | 1.17E-13 |
| | S3621 | 14 | 5 | 9 | 3 | 43884819 | 44031978 | 147.16 | 223 | 4.51E-32 | 6.67E-02 | 1.20E-03 | 3.02E-35 |
| | S3623 | 14 | 5 | 9 | 3 | 142328162 | 142820107 | 491.946 | 605 | 4.51E-32 | 6.67E-02 | 1.20E-03 | 3.02E-35 |
| | S3631 | 14 | 3 | 11 | 4 | 80439027 | 80572955 | 133.929 | 177 | 1.74E-10 | 4.00E-02 | 1.47E-03 | 1.17E-13 |
| | S3633 | 14 | 5 | 9 | 4 | 100327069 | 100689879 | 362.811 | 781 | 4.51E-32 | 6.67E-02 | 1.20E-03 | 3.02E-35 |

| | | | | | | | | | | | | | |
|---|---|---|---|---|---|---|---|---|---|---|---|---|---|
| | S3635 | 14 | 4 | 10 | 4 | 111629692 | 112326279 | 696.588 | 839 | 1.04E-19 | 5.33E-02 | 1.33E-03 | 6.95E-23 |
| | S3637 | 14 | 5 | 9 | 4 | 152491242 | 152626885 | 135.644 | 121 | 4.51E-32 | 6.67E-02 | 1.20E-03 | 3.02E-35 |
| | S3640 | 14 | 3 | 11 | 4 | 161328894 | 162324602 | 995.709 | 2003 | 1.74E-10 | 4.00E-02 | 1.47E-03 | 1.17E-13 |
| | S3642 | 14 | 4 | 10 | 5 | 55224050 | 55413868 | 189.819 | 243 | 1.04E-19 | 5.33E-02 | 1.33E-03 | 6.95E-23 |
| | S3643 | 14 | 4 | 10 | 5 | 55593749 | 56123279 | 529.531 | 779 | 1.04E-19 | 5.33E-02 | 1.33E-03 | 6.95E-23 |
| | S3646 | 14 | 5 | 9 | 5 | 65783584 | 66215943 | 432.36 | 568 | 4.51E-32 | 6.67E-02 | 1.20E-03 | 3.02E-35 |
| | S3647 | 14 | 5 | 9 | 5 | 97176979 | 97453455 | 276.477 | 473 | 4.51E-32 | 6.67E-02 | 1.20E-03 | 3.02E-35 |
| | S3651 | 14 | 3 | 11 | 5 | 160974709 | 161401577 | 426.869 | 908 | 1.74E-10 | 4.00E-02 | 1.47E-03 | 1.17E-13 |
| | S3652 | 14 | 3 | 11 | 5 | 161703565 | 161870295 | 166.731 | 347 | 1.74E-10 | 4.00E-02 | 1.47E-03 | 1.17E-13 |
| | S3661 | 14 | 3 | 11 | 6 | 131646664 | 131801369 | 154.706 | 177 | 1.74E-10 | 4.00E-02 | 1.47E-03 | 1.17E-13 |
| | S3663 | 14 | 5 | 9 | 6 | 162515945 | 163183260 | 667.316 | 1299 | 4.51E-32 | 6.67E-02 | 1.20E-03 | 3.02E-35 |
| | S3668 | 14 | 5 | 9 | 7 | 109100534 | 109589327 | 488.794 | 873 | 4.51E-32 | 6.67E-02 | 1.20E-03 | 3.02E-35 |
| | S3674 | 14 | 3 | 11 | 8 | 95577650 | 95684951 | 107.302 | 259 | 1.74E-10 | 4.00E-02 | 1.47E-03 | 1.17E-13 |
| | S3676 | 14 | 3 | 11 | 8 | 96300258 | 96435196 | 134.939 | 233 | 1.74E-10 | 4.00E-02 | 1.47E-03 | 1.17E-13 |

| | | | | | | | | | | | | | |
|---|---|---|---|---|---|---|---|---|---|---|---|---|---|
| | S3679 | 14 | 5 | 9 | 9 | 104299284 | 104783622 | 484.339 | 850 | 4.51E-32 | 6.67E-02 | 1.20E-03 | 3.02E-35 |
| | S3682 | 14 | 4 | 10 | 9 | 110007352 | 110219729 | 212.378 | 462 | 1.04E-19 | 5.33E-02 | 1.33E-03 | 6.95E-23 |
| | S3683 | 14 | 4 | 10 | 9 | 112215418 | 112505048 | 289.631 | 508 | 1.04E-19 | 5.33E-02 | 1.33E-03 | 6.95E-23 |
| | S3698 | 14 | 3 | 11 | 12 | 29224621 | 29496222 | 271.602 | 524 | 1.74E-10 | 4.00E-02 | 1.47E-03 | 1.17E-13 |
| | S3700 | 14 | 3 | 11 | 12 | 46848016 | 47082605 | 234.59 | 425 | 1.74E-10 | 4.00E-02 | 1.47E-03 | 1.17E-13 |
| | S3704 | 14 | 3 | 11 | 12 | 102210457 | 102502099 | 291.643 | 321 | 1.74E-10 | 4.00E-02 | 1.47E-03 | 1.17E-13 |
| | S3708 | 14 | 3 | 11 | 13 | 76566952 | 76809889 | 242.938 | 452 | 1.74E-10 | 4.00E-02 | 1.47E-03 | 1.17E-13 |
| | S3709 | 14 | 3 | 11 | 13 | 92656308 | 93204655 | 548.348 | 916 | 1.74E-10 | 4.00E-02 | 1.47E-03 | 1.17E-13 |
| | S3719 | 14 | 4 | 10 | 15 | 46794051 | 47655539 | 861.489 | 1360 | 1.04E-19 | 5.33E-02 | 1.33E-03 | 6.95E-23 |
| | S3723 | 14 | 6 | 8 | 16 | 61062499 | 61737667 | 675.169 | 828 | 1.39E-47 | 8.00E-02 | 1.07E-03 | 9.32E-51 |
| | S3736 | 13 | 4 | 9 | 1 | 158051970 | 158405408 | 353.439 | 790 | 3.22E-21 | 5.33E-02 | 1.20E-03 | 2.16E-24 |
| | S3737 | 13 | 4 | 9 | 1 | 158410638 | 158668470 | 257.833 | 575 | 3.22E-21 | 5.33E-02 | 1.20E-03 | 2.16E-24 |
| | S3738 | 13 | 3 | 10 | 1 | 158790636 | 158936329 | 145.694 | 359 | 2.92E-11 | 4.00E-02 | 1.33E-03 | 1.96E-14 |
| | S3739 | 13 | 3 | 10 | 1 | 176025894 | 176966604 | 940.711 | 1736 | 2.92E-11 | 4.00E-02 | 1.33E-03 | 1.96E-14 |

| | | | | | | | | | | | | | |
|---|---|---|---|---|---|---|---|---|---|---|---|---|---|
| | S3751 | 13 | 4 | 9 | 2 | 49830251 | 50273223 | 442.973 | 964 | 3.22E-21 | 5.33E-02 | 1.20E-03 | 2.16E-24 |
| | S3755 | 13 | 5 | 8 | 2 | 77994546 | 78329816 | 335.271 | 635 | 1.49E-34 | 6.67E-02 | 1.07E-03 | 9.99E-38 |
| | S3756 | 13 | 5 | 8 | 2 | 78578705 | 78792492 | 213.788 | 355 | 1.49E-34 | 6.67E-02 | 1.07E-03 | 9.99E-38 |
| | S3770 | 13 | 5 | 8 | 3 | 43247708 | 43806958 | 559.251 | 705 | 1.49E-34 | 6.67E-02 | 1.07E-03 | 9.99E-38 |
| | S3773 | 13 | 4 | 9 | 4 | 15021890 | 15336242 | 314.353 | 556 | 3.22E-21 | 5.33E-02 | 1.20E-03 | 2.16E-24 |
| | S3776 | 13 | 3 | 10 | 4 | 77249630 | 77372145 | 122.516 | 304 | 2.92E-11 | 4.00E-02 | 1.33E-03 | 1.96E-14 |
| | S3781 | 13 | 3 | 10 | 4 | 125345151 | 125533756 | 188.606 | 339 | 2.92E-11 | 4.00E-02 | 1.33E-03 | 1.96E-14 |
| | S3784 | 13 | 4 | 9 | 4 | 154900767 | 155009051 | 108.285 | 155 | 3.22E-21 | 5.33E-02 | 1.20E-03 | 2.16E-24 |
| | S3790 | 13 | 5 | 8 | 5 | 56391208 | 56606118 | 214.911 | 466 | 1.49E-34 | 6.67E-02 | 1.07E-03 | 9.99E-38 |
| | S3807 | 13 | 4 | 9 | 6 | 142816051 | 143009259 | 193.209 | 349 | 3.22E-21 | 5.33E-02 | 1.20E-03 | 2.16E-24 |
| | S3808 | 13 | 4 | 9 | 7 | 23067749 | 23926540 | 858.792 | 1480 | 3.22E-21 | 5.33E-02 | 1.20E-03 | 2.16E-24 |
| | S3826 | 13 | 4 | 9 | 9 | 32939659 | 33090322 | 150.664 | 293 | 3.22E-21 | 5.33E-02 | 1.20E-03 | 2.16E-24 |
| | S3827 | 13 | 4 | 9 | 9 | 83354952 | 84054511 | 699.56 | 1341 | 3.22E-21 | 5.33E-02 | 1.20E-03 | 2.16E-24 |
| | S3828 | 13 | 4 | 9 | 9 | 105079259 | 105519419 | 440.161 | 629 | 3.22E-21 | 5.33E-02 | 1.20E-03 | 2.16E-24 |

| | | | | | | | | | | | | | |
|---|---|---|---|---|---|---|---|---|---|---|---|---|---|
| | S3831 | 13 | 4 | 9 | 9 | 109603390 | 109981608 | 378.219 | 868 | 3.22E-21 | 5.33E-02 | 1.20E-03 | 2.16E-24 |
| | S3832 | 13 | 5 | 8 | 9 | 122808071 | 123055769 | 247.699 | 245 | 1.49E-34 | 6.67E-02 | 1.07E-03 | 9.99E-38 |
| | S3841 | 13 | 3 | 10 | 10 | 88700053 | 88986603 | 286.551 | 480 | 2.92E-11 | 4.00E-02 | 1.33E-03 | 1.96E-14 |
| | S3848 | 13 | 3 | 10 | 12 | 22168408 | 22587409 | 419.002 | 576 | 2.92E-11 | 4.00E-02 | 1.33E-03 | 1.96E-14 |
| | S3873 | 13 | 5 | 8 | 14 | 50513086 | 50640439 | 127.354 | 178 | 1.49E-34 | 6.67E-02 | 1.07E-03 | 9.99E-38 |
| | S3876 | 13 | 4 | 9 | 14 | 68661235 | 68850260 | 189.026 | 474 | 3.22E-21 | 5.33E-02 | 1.20E-03 | 2.16E-24 |
| | S3877 | 13 | 3 | 10 | 14 | 69353697 | 69669707 | 316.011 | 421 | 2.92E-11 | 4.00E-02 | 1.33E-03 | 1.96E-14 |
| | S3891 | 13 | 3 | 10 | 15 | 78420357 | 78662037 | 241.681 | 400 | 2.92E-11 | 4.00E-02 | 1.33E-03 | 1.96E-14 |
| | S3894 | 13 | 3 | 10 | 16 | 64747261 | 65197000 | 449.74 | 962 | 2.92E-11 | 4.00E-02 | 1.33E-03 | 1.96E-14 |
| | S3897 | 13 | 3 | 10 | 16 | 74092772 | 74280381 | 187.61 | 358 | 2.92E-11 | 4.00E-02 | 1.33E-03 | 1.96E-14 |
| | S3899 | 13 | 4 | 9 | 16 | 75488802 | 75615181 | 126.38 | 215 | 3.22E-21 | 5.33E-02 | 1.20E-03 | 2.16E-24 |
| | S3901 | 13 | 3 | 10 | 16 | 76012030 | 76207515 | 195.486 | 355 | 2.92E-11 | 4.00E-02 | 1.33E-03 | 1.96E-14 |
| | S3905 | 13 | 3 | 10 | 18 | 49658476 | 50557654 | 899.179 | 1740 | 2.92E-11 | 4.00E-02 | 1.33E-03 | 1.96E-14 |
| | S3908 | 13 | 4 | 9 | 20 | 44962225 | 46208137 | 1245.913 | 2007 | 3.22E-21 | 5.33E-02 | 1.20E-03 | 2.16E-24 |

| | | | | | | | | | | | | | |
|---|---|---|---|---|---|---|---|---|---|---|---|---|---|
| | S3915 | 12 | 3 | 9 | 1 | 105160481 | 105349960 | 189.48 | 426 | 3.64E-12 | 4.00E-02 | 1.20E-03 | 2.44E-15 |
| | S3918 | 12 | 3 | 9 | 1 | 169116349 | 169504265 | 387.917 | 826 | 3.64E-12 | 4.00E-02 | 1.20E-03 | 2.44E-15 |
| | S3919 | 12 | 3 | 9 | 1 | 169552079 | 169679328 | 127.25 | 317 | 3.64E-12 | 4.00E-02 | 1.20E-03 | 2.44E-15 |
| | S3926 | 12 | 3 | 9 | 1 | 210074471 | 210244031 | 169.561 | 249 | 3.64E-12 | 4.00E-02 | 1.20E-03 | 2.44E-15 |
| | S3944 | 12 | 3 | 9 | 2 | 180600610 | 180769937 | 169.328 | 370 | 3.64E-12 | 4.00E-02 | 1.20E-03 | 2.44E-15 |
| | S3951 | 12 | 4 | 8 | 3 | 42818666 | 43057981 | 239.316 | 321 | 5.64E-23 | 5.33E-02 | 1.07E-03 | 3.78E-26 |
| | S3956 | 12 | 5 | 7 | 3 | 143529605 | 143642198 | 112.594 | 236 | 1.90E-37 | 6.67E-02 | 9.33E-04 | 1.28E-40 |
| | S3960 | 12 | 4 | 8 | 4 | 12137081 | 13072922 | 935.842 | 1989 | 5.64E-23 | 5.33E-02 | 1.07E-03 | 3.78E-26 |
| | S3964 | 12 | 4 | 8 | 4 | 55690554 | 56662840 | 972.287 | 1845 | 5.64E-23 | 5.33E-02 | 1.07E-03 | 3.78E-26 |
| | S3977 | 12 | 5 | 7 | 5 | 66336449 | 66615465 | 279.017 | 559 | 1.90E-37 | 6.67E-02 | 9.33E-04 | 1.28E-40 |
| | S3979 | 12 | 3 | 9 | 5 | 72186060 | 73204954 | 1018.895 | 1358 | 3.64E-12 | 4.00E-02 | 1.20E-03 | 2.44E-15 |
| | S3980 | 12 | 3 | 9 | 5 | 74945323 | 76297886 | 1352.564 | 2231 | 3.64E-12 | 4.00E-02 | 1.20E-03 | 2.44E-15 |
| | S3981 | 12 | 5 | 7 | 5 | 96626713 | 96745995 | 119.283 | 236 | 1.90E-37 | 6.67E-02 | 9.33E-04 | 1.28E-40 |
| | S3990 | 12 | 3 | 9 | 6 | 132608199 | 133163967 | 555.769 | 978 | 3.64E-12 | 4.00E-02 | 1.20E-03 | 2.44E-15 |

| | | | | | | | | | | | | | |
|---|---|---|---|---|---|---|---|---|---|---|---|---|---|
| | S3991 | 12 | 4 | 8 | 7 | 24108825 | 24212310 | 103.486 | 213 | 5.64E-23 | 5.33E-02 | 1.07E-03 | 3.78E-26 |
| | S4002 | 12 | 3 | 9 | 7 | 80201263 | 80436361 | 235.099 | 398 | 3.64E-12 | 4.00E-02 | 1.20E-03 | 2.44E-15 |
| | S4003 | 12 | 3 | 9 | 7 | 80531052 | 80701284 | 170.233 | 242 | 3.64E-12 | 4.00E-02 | 1.20E-03 | 2.44E-15 |
| | S4012 | 12 | 3 | 9 | 7 | 94921248 | 95451030 | 529.783 | 728 | 3.64E-12 | 4.00E-02 | 1.20E-03 | 2.44E-15 |
| | S4018 | 12 | 3 | 9 | 8 | 95194325 | 95374265 | 179.941 | 374 | 3.64E-12 | 4.00E-02 | 1.20E-03 | 2.44E-15 |
| | S4026 | 12 | 3 | 9 | 9 | 84627121 | 84802872 | 175.752 | 283 | 3.64E-12 | 4.00E-02 | 1.20E-03 | 2.44E-15 |
| | S4027 | 12 | 3 | 9 | 9 | 85095932 | 85201941 | 106.01 | 151 | 3.64E-12 | 4.00E-02 | 1.20E-03 | 2.44E-15 |
| | S4031 | 12 | 3 | 9 | 9 | 122024841 | 122181435 | 156.595 | 224 | 3.64E-12 | 4.00E-02 | 1.20E-03 | 2.44E-15 |
| | S4032 | 12 | 3 | 9 | 9 | 122225314 | 122376416 | 151.103 | 157 | 3.64E-12 | 4.00E-02 | 1.20E-03 | 2.44E-15 |
| | S4034 | 12 | 4 | 8 | 9 | 123984155 | 124102218 | 118.064 | 250 | 5.64E-23 | 5.33E-02 | 1.07E-03 | 3.78E-26 |
| | S4038 | 12 | 3 | 9 | 10 | 89225030 | 89505741 | 280.712 | 458 | 3.64E-12 | 4.00E-02 | 1.20E-03 | 2.44E-15 |
| | S4039 | 12 | 3 | 9 | 10 | 89521788 | 89865786 | 343.999 | 686 | 3.64E-12 | 4.00E-02 | 1.20E-03 | 2.44E-15 |
| | S4043 | 12 | 7 | 5 | 10 | 97987740 | 98253487 | 265.748 | 335 | 1.97E-77 | 9.33E-02 | 6.66E-04 | 1.32E-80 |
| | S4049 | 12 | 3 | 9 | 11 | 101006738 | 101288537 | 281.8 | 601 | 3.64E-12 | 4.00E-02 | 1.20E-03 | 2.44E-15 |

| | | | | | | | | | | | | | |
|---|---|---|---|---|---|---|---|---|---|---|---|---|---|
| | S4051 | 12 | 4 | 8 | 11 | 102163463 | 102359398 | 195.936 | 243 | 5.64E-23 | 5.33E-02 | 1.07E-03 | 3.78E-26 |
| | S4053 | 12 | 4 | 8 | 11 | 103042416 | 103185625 | 143.21 | 221 | 5.64E-23 | 5.33E-02 | 1.07E-03 | 3.78E-26 |
| | S4056 | 12 | 3 | 9 | 12 | 21026265 | 21527675 | 501.411 | 1106 | 3.64E-12 | 4.00E-02 | 1.20E-03 | 2.44E-15 |
| | S4057 | 12 | 3 | 9 | 12 | 21594374 | 21963053 | 368.68 | 364 | 3.64E-12 | 4.00E-02 | 1.20E-03 | 2.44E-15 |
| | S4066 | 12 | 3 | 9 | 12 | 105549022 | 105811720 | 262.699 | 511 | 3.64E-12 | 4.00E-02 | 1.20E-03 | 2.44E-15 |
| | S4071 | 12 | 4 | 8 | 14 | 51245994 | 51597625 | 351.632 | 780 | 5.64E-23 | 5.33E-02 | 1.07E-03 | 3.78E-26 |
| | S4079 | 12 | 4 | 8 | 15 | 55357863 | 55537491 | 179.629 | 378 | 5.64E-23 | 5.33E-02 | 1.07E-03 | 3.78E-26 |
| | S4082 | 12 | 3 | 9 | 15 | 79283727 | 79560175 | 276.449 | 574 | 3.64E-12 | 4.00E-02 | 1.20E-03 | 2.44E-15 |
| | S4083 | 12 | 4 | 8 | 15 | 80221491 | 80334134 | 112.644 | 262 | 5.64E-23 | 5.33E-02 | 1.07E-03 | 3.78E-26 |
| | S4084 | 12 | 6 | 6 | 16 | 50982680 | 51286761 | 304.082 | 587 | 1.35E-55 | 8.00E-02 | 7.99E-04 | 9.04E-59 |
| | S4086 | 12 | 3 | 9 | 16 | 76398926 | 76558842 | 159.917 | 368 | 3.64E-12 | 4.00E-02 | 1.20E-03 | 2.44E-15 |
| | S4089 | 12 | 3 | 9 | 18 | 35402493 | 36084761 | 682.269 | 1161 | 3.64E-12 | 4.00E-02 | 1.20E-03 | 2.44E-15 |
| | S4097 | 12 | 3 | 9 | 20 | 48588786 | 48882173 | 293.388 | 571 | 3.64E-12 | 4.00E-02 | 1.20E-03 | 2.44E-15 |
| | S4100 | 11 | 5 | 6 | 1 | 7698700 | 9019095 | 1320.396 | 2489 | 7.27E-41 | 6.67E-02 | 7.99E-04 | 4.87E-44 |

| | | | | | | | | | | | | | |
|---|---|---|---|---|---|---|---|---|---|---|---|---|---|
| | S4103 | 11 | 2 | 9 | 1 | 36142826 | 36392544 | 249.719 | 285 | 2.23E-05 | 2.67E-02 | 1.20E-03 | 1.49E-08 |
| | S4107 | 11 | 2 | 9 | 1 | 84756600 | 84925457 | 168.858 | 438 | 2.23E-05 | 2.67E-02 | 1.20E-03 | 1.49E-08 |
| | S4108 | 11 | 4 | 7 | 1 | 107493034 | 107984430 | 491.397 | 1164 | 4.75E-25 | 5.33E-02 | 9.33E-04 | 3.19E-28 |
| | S4111 | 11 | 3 | 8 | 1 | 115493253 | 115870126 | 376.874 | 780 | 3.11E-13 | 4.00E-02 | 1.07E-03 | 2.09E-16 |
| | S4113 | 11 | 3 | 8 | 1 | 159146030 | 159758862 | 612.833 | 1450 | 3.11E-13 | 4.00E-02 | 1.07E-03 | 2.09E-16 |
| | S4115 | 11 | 3 | 8 | 1 | 160672214 | 160905887 | 233.674 | 426 | 3.11E-13 | 4.00E-02 | 1.07E-03 | 2.09E-16 |
| | S4116 | 11 | 3 | 8 | 1 | 160950381 | 161259053 | 308.673 | 581 | 3.11E-13 | 4.00E-02 | 1.07E-03 | 2.09E-16 |
| | S4122 | 11 | 2 | 9 | 1 | 184020326 | 184627672 | 607.347 | 1163 | 2.23E-05 | 2.67E-02 | 1.20E-03 | 1.49E-08 |
| | S4138 | 11 | 3 | 8 | 2 | 105262911 | 105434566 | 171.656 | 376 | 3.11E-13 | 4.00E-02 | 1.07E-03 | 2.09E-16 |
| | S4150 | 11 | 2 | 9 | 2 | 228215401 | 228465495 | 250.095 | 483 | 2.23E-05 | 2.67E-02 | 1.20E-03 | 1.49E-08 |
| | S4153 | 11 | 2 | 9 | 3 | 16764179 | 17118586 | 354.408 | 641 | 2.23E-05 | 2.67E-02 | 1.20E-03 | 1.49E-08 |
| | S4154 | 11 | 2 | 9 | 3 | 17148999 | 17960084 | 811.086 | 857 | 2.23E-05 | 2.67E-02 | 1.20E-03 | 1.49E-08 |
| | S4157 | 11 | 4 | 7 | 3 | 42513672 | 42670027 | 156.356 | 240 | 4.75E-25 | 5.33E-02 | 9.33E-04 | 3.19E-28 |
| | S4158 | 11 | 3 | 8 | 3 | 64560783 | 64893582 | 332.8 | 627 | 3.11E-13 | 4.00E-02 | 1.07E-03 | 2.09E-16 |

| | | | | | | | | | | | | | |
|---|---|---|---|---|---|---|---|---|---|---|---|---|---|
| | S4159 | 11 | 3 | 8 | 3 | 64900811 | 65746810 | 846 | 1902 | 3.11E-13 | 4.00E-02 | 1.07E-03 | 2.09E-16 |
| | S4160 | 11 | 3 | 8 | 3 | 76723424 | 76954294 | 230.871 | 487 | 3.11E-13 | 4.00E-02 | 1.07E-03 | 2.09E-16 |
| | S4162 | 11 | 4 | 7 | 3 | 131972365 | 132109860 | 137.496 | 317 | 4.75E-25 | 5.33E-02 | 9.33E-04 | 3.19E-28 |
| | S4169 | 11 | 4 | 7 | 4 | 13295980 | 13526740 | 230.761 | 236 | 4.75E-25 | 5.33E-02 | 9.33E-04 | 3.19E-28 |
| | S4171 | 11 | 2 | 9 | 4 | 22600521 | 22829316 | 228.796 | 337 | 2.23E-05 | 2.67E-02 | 1.20E-03 | 1.49E-08 |
| | S4172 | 11 | 2 | 9 | 4 | 22877503 | 23357754 | 480.252 | 580 | 2.23E-05 | 2.67E-02 | 1.20E-03 | 1.49E-08 |
| | S4173 | 11 | 2 | 9 | 4 | 23364823 | 23565257 | 200.435 | 397 | 2.23E-05 | 2.67E-02 | 1.20E-03 | 1.49E-08 |
| | S4181 | 11 | 2 | 9 | 4 | 153673034 | 153896765 | 223.732 | 388 | 2.23E-05 | 2.67E-02 | 1.20E-03 | 1.49E-08 |
| | S4190 | 11 | 2 | 9 | 5 | 32866393 | 33474832 | 608.44 | 1190 | 2.23E-05 | 2.67E-02 | 1.20E-03 | 1.49E-08 |
| | S4196 | 11 | 5 | 6 | 5 | 66974494 | 67142346 | 167.853 | 211 | 7.27E-41 | 6.67E-02 | 7.99E-04 | 4.87E-44 |
| | S4197 | 11 | 5 | 6 | 5 | 67187297 | 67339845 | 152.549 | 238 | 7.27E-41 | 6.67E-02 | 7.99E-04 | 4.87E-44 |
| | S4201 | 11 | 5 | 6 | 5 | 68012384 | 68371535 | 359.152 | 647 | 7.27E-41 | 6.67E-02 | 7.99E-04 | 4.87E-44 |
| | S4204 | 11 | 5 | 6 | 5 | 126163276 | 126450912 | 287.637 | 615 | 7.27E-41 | 6.67E-02 | 7.99E-04 | 4.87E-44 |
| | S4207 | 11 | 3 | 8 | 6 | 135386348 | 136047840 | 661.493 | 812 | 3.11E-13 | 4.00E-02 | 1.07E-03 | 2.09E-16 |

| | | | | | | | | | | | | | |
|---|---|---|---|---|---|---|---|---|---|---|---|---|---|
| | S4208 | 11 | 4 | 7 | 6 | 147016387 | 147497968 | 481.582 | 474 | 4.75E-25 | 5.33E-02 | 9.33E-04 | 3.19E-28 |
| | S4210 | 11 | 4 | 7 | 6 | 157997246 | 158853069 | 855.824 | 1826 | 4.75E-25 | 5.33E-02 | 9.33E-04 | 3.19E-28 |
| | S4211 | 11 | 3 | 8 | 6 | 161444200 | 161686497 | 242.298 | 542 | 3.11E-13 | 4.00E-02 | 1.07E-03 | 2.09E-16 |
| | S4212 | 11 | 6 | 5 | 6 | 163582685 | 163903689 | 321.005 | 630 | 1.08E-60 | 8.00E-02 | 6.66E-04 | 7.23E-64 |
| | S4213 | 11 | 2 | 9 | 7 | 5297549 | 5842100 | 544.552 | 1163 | 2.23E-05 | 2.67E-02 | 1.20E-03 | 1.49E-08 |
| | S4214 | 11 | 2 | 9 | 7 | 5864236 | 6838803 | 974.568 | 1654 | 2.23E-05 | 2.67E-02 | 1.20E-03 | 1.49E-08 |
| | S4215 | 11 | 3 | 8 | 7 | 20089076 | 20225757 | 136.682 | 267 | 3.11E-13 | 4.00E-02 | 1.07E-03 | 2.09E-16 |
| | S4228 | 11 | 2 | 9 | 7 | 79287574 | 79815754 | 528.181 | 811 | 2.23E-05 | 2.67E-02 | 1.20E-03 | 1.49E-08 |
| | S4229 | 11 | 3 | 8 | 7 | 81340584 | 81772828 | 432.245 | 510 | 3.11E-13 | 4.00E-02 | 1.07E-03 | 2.09E-16 |
| | S4230 | 11 | 4 | 7 | 7 | 81774653 | 82384018 | 609.366 | 1198 | 4.75E-25 | 5.33E-02 | 9.33E-04 | 3.19E-28 |
| | S4232 | 11 | 4 | 7 | 7 | 94426694 | 94684311 | 257.618 | 360 | 4.75E-25 | 5.33E-02 | 9.33E-04 | 3.19E-28 |
| | S4233 | 11 | 3 | 8 | 7 | 105403992 | 105509980 | 105.989 | 170 | 3.11E-13 | 4.00E-02 | 1.07E-03 | 2.09E-16 |
| | S4234 | 11 | 4 | 7 | 7 | 108000008 | 108139685 | 139.678 | 164 | 4.75E-25 | 5.33E-02 | 9.33E-04 | 3.19E-28 |
| | S4235 | 11 | 2 | 9 | 7 | 129246980 | 130071005 | 824.026 | 1441 | 2.23E-05 | 2.67E-02 | 1.20E-03 | 1.49E-08 |

| | | | | | | | | | | | | | |
|---|---|---|---|---|---|---|---|---|---|---|---|---|---|
| | S4244 | 11 | 3 | 8 | 8 | 71833712 | 71955377 | 121.666 | 288 | 3.11E-13 | 4.00E-02 | 1.07E-03 | 2.09E-16 |
| | S4252 | 11 | 3 | 8 | 8 | 129064176 | 129340981 | 276.806 | 463 | 3.11E-13 | 4.00E-02 | 1.07E-03 | 2.09E-16 |
| | S4254 | 11 | 2 | 9 | 8 | 136664978 | 137613088 | 948.111 | 1700 | 2.23E-05 | 2.67E-02 | 1.20E-03 | 1.49E-08 |
| | S4262 | 11 | 2 | 9 | 9 | 85399570 | 85977842 | 578.273 | 742 | 2.23E-05 | 2.67E-02 | 1.20E-03 | 1.49E-08 |
| | S4264 | 11 | 5 | 6 | 9 | 107582729 | 107726008 | 143.28 | 316 | 7.27E-41 | 6.67E-02 | 7.99E-04 | 4.87E-44 |
| | S4265 | 11 | 2 | 9 | 9 | 116335288 | 116722340 | 387.053 | 738 | 2.23E-05 | 2.67E-02 | 1.20E-03 | 1.49E-08 |
| | S4273 | 11 | 2 | 9 | 10 | 82737604 | 83312126 | 574.523 | 1181 | 2.23E-05 | 2.67E-02 | 1.20E-03 | 1.49E-08 |
| | S4277 | 11 | 3 | 8 | 11 | 22857299 | 23056677 | 199.379 | 462 | 3.11E-13 | 4.00E-02 | 1.07E-03 | 2.09E-16 |
| | S4278 | 11 | 3 | 8 | 11 | 24064194 | 24418784 | 354.591 | 770 | 3.11E-13 | 4.00E-02 | 1.07E-03 | 2.09E-16 |
| | S4279 | 11 | 3 | 8 | 11 | 24421480 | 24735779 | 314.3 | 721 | 3.11E-13 | 4.00E-02 | 1.07E-03 | 2.09E-16 |
| | S4285 | 11 | 3 | 8 | 11 | 100884590 | 101002222 | 117.633 | 322 | 3.11E-13 | 4.00E-02 | 1.07E-03 | 2.09E-16 |
| | S4289 | 11 | 2 | 9 | 12 | 69603502 | 70418763 | 815.262 | 1946 | 2.23E-05 | 2.67E-02 | 1.20E-03 | 1.49E-08 |
| | S4295 | 11 | 2 | 9 | 12 | 108933605 | 109202226 | 268.622 | 416 | 2.23E-05 | 2.67E-02 | 1.20E-03 | 1.49E-08 |
| | S4296 | 11 | 2 | 9 | 13 | 47410436 | 47555398 | 144.963 | 255 | 2.23E-05 | 2.67E-02 | 1.20E-03 | 1.49E-08 |

| | | | | | | | | | | | | | |
|---|---|---|---|---|---|---|---|---|---|---|---|---|---|
| | S4299 | 11 | 3 | 8 | 13 | 75974766 | 76279883 | 305.118 | 759 | 3.11E-13 | 4.00E-02 | 1.07E-03 | 2.09E-16 |
| | S4304 | 11 | 3 | 8 | 13 | 94442131 | 95046259 | 604.129 | 1140 | 3.11E-13 | 4.00E-02 | 1.07E-03 | 2.09E-16 |
| | S4306 | 11 | 4 | 7 | 13 | 102103344 | 102299561 | 196.218 | 410 | 4.75E-25 | 5.33E-02 | 9.33E-04 | 3.19E-28 |
| | S4311 | 11 | 2 | 9 | 14 | 56317271 | 56437219 | 119.949 | 288 | 2.23E-05 | 2.67E-02 | 1.20E-03 | 1.49E-08 |
| | S4320 | 11 | 4 | 7 | 15 | 52973737 | 53244988 | 271.252 | 561 | 4.75E-25 | 5.33E-02 | 9.33E-04 | 3.19E-28 |
| | S4323 | 11 | 5 | 6 | 15 | 65502862 | 66126696 | 623.835 | 933 | 7.27E-41 | 6.67E-02 | 7.99E-04 | 4.87E-44 |
| | S4324 | 11 | 5 | 6 | 16 | 51998601 | 52452407 | 453.807 | 820 | 7.27E-41 | 6.67E-02 | 7.99E-04 | 4.87E-44 |
| | S4325 | 11 | 5 | 6 | 16 | 58288385 | 58461459 | 173.075 | 271 | 7.27E-41 | 6.67E-02 | 7.99E-04 | 4.87E-44 |
| | S4329 | 11 | 2 | 9 | 16 | 73544935 | 73865221 | 320.287 | 584 | 2.23E-05 | 2.67E-02 | 1.20E-03 | 1.49E-08 |
| | S4330 | 11 | 3 | 8 | 16 | 77098072 | 77389530 | 291.459 | 885 | 3.11E-13 | 4.00E-02 | 1.07E-03 | 2.09E-16 |
| | S4331 | 11 | 3 | 8 | 16 | 77457800 | 77709032 | 251.233 | 769 | 3.11E-13 | 4.00E-02 | 1.07E-03 | 2.09E-16 |
| | S4333 | 11 | 3 | 8 | 18 | 27519587 | 27668626 | 149.04 | 262 | 3.11E-13 | 4.00E-02 | 1.07E-03 | 2.09E-16 |
| | S4335 | 11 | 2 | 9 | 18 | 57290500 | 57670960 | 380.461 | 725 | 2.23E-05 | 2.67E-02 | 1.20E-03 | 1.49E-08 |
| | S4340 | 11 | 2 | 9 | 20 | 49781513 | 50058789 | 277.277 | 544 | 2.23E-05 | 2.67E-02 | 1.20E-03 | 1.49E-08 |

| | | | | | | | | | | | | | |
|---|---|---|---|---|---|---|---|---|---|---|---|---|---|
| | S4344 | 11 | 3 | 8 | 22 | 34151045 | 34597404 | 446.36 | 994 | 3.11E-13 | 4.00E-02 | 1.07E-03 | 2.09E-16 |
| | S4345 | 11 | 3 | 8 | 22 | 34749533 | 34895284 | 145.752 | 318 | 3.11E-13 | 4.00E-02 | 1.07E-03 | 2.09E-16 |
| | S4349 | 10 | 2 | 8 | 1 | 23015700 | 23814521 | 798.822 | 1199 | 7.49E-06 | 2.67E-02 | 1.07E-03 | 5.02E-09 |
| | S4356 | 10 | 4 | 6 | 1 | 114799108 | 115297896 | 498.789 | 1189 | 1.54E-27 | 5.33E-02 | 7.99E-04 | 1.04E-30 |
| | S4359 | 10 | 4 | 6 | 1 | 157345386 | 157481515 | 136.13 | 314 | 1.54E-27 | 5.33E-02 | 7.99E-04 | 1.04E-30 |
| | S4360 | 10 | 3 | 7 | 1 | 159957347 | 160496566 | 539.22 | 945 | 1.64E-14 | 4.00E-02 | 9.33E-04 | 1.10E-17 |
| | S4361 | 10 | 3 | 7 | 1 | 162470743 | 162600585 | 129.843 | 222 | 1.64E-14 | 4.00E-02 | 9.33E-04 | 1.10E-17 |
| | S4362 | 10 | 3 | 7 | 1 | 162998128 | 163530994 | 532.867 | 1437 | 1.64E-14 | 4.00E-02 | 9.33E-04 | 1.10E-17 |
| | S4363 | 10 | 3 | 7 | 1 | 163539720 | 164268365 | 728.646 | 1443 | 1.64E-14 | 4.00E-02 | 9.33E-04 | 1.10E-17 |
| | S4364 | 10 | 3 | 7 | 1 | 168333311 | 168455826 | 122.516 | 237 | 1.64E-14 | 4.00E-02 | 9.33E-04 | 1.10E-17 |
| | S4371 | 10 | 4 | 6 | 2 | 79508574 | 79696007 | 187.434 | 345 | 1.54E-27 | 5.33E-02 | 7.99E-04 | 1.04E-30 |
| | S4386 | 10 | 2 | 8 | 3 | 1409603 | 1842964 | 433.362 | 1346 | 7.49E-06 | 2.67E-02 | 1.07E-03 | 5.02E-09 |
| | S4387 | 10 | 3 | 7 | 3 | 2393584 | 2709509 | 315.926 | 730 | 1.64E-14 | 4.00E-02 | 9.33E-04 | 1.10E-17 |
| | S4389 | 10 | 4 | 6 | 3 | 60800620 | 61950254 | 1149.635 | 2275 | 1.54E-27 | 5.33E-02 | 7.99E-04 | 1.04E-30 |

| | | | | | | | | | | | | | |
|---|---|---|---|---|---|---|---|---|---|---|---|---|---|
| | S4391 | 10 | 3 | 7 | 3 | 69855721 | 70036135 | 180.415 | 175 | 1.64E-14 | 4.00E-02 | 9.33E-04 | 1.10E-17 |
| | S4394 | 10 | 3 | 7 | 3 | 112764530 | 113086154 | 321.625 | 542 | 1.64E-14 | 4.00E-02 | 9.33E-04 | 1.10E-17 |
| | S4395 | 10 | 3 | 7 | 3 | 123430486 | 123695355 | 264.87 | 309 | 1.64E-14 | 4.00E-02 | 9.33E-04 | 1.10E-17 |
| | S4396 | 10 | 3 | 7 | 3 | 123876585 | 124550093 | 673.509 | 958 | 1.64E-14 | 4.00E-02 | 9.33E-04 | 1.10E-17 |
| | S4397 | 10 | 3 | 7 | 3 | 124556912 | 124967889 | 410.978 | 915 | 1.64E-14 | 4.00E-02 | 9.33E-04 | 1.10E-17 |
| | S4399 | 10 | 3 | 7 | 3 | 150713302 | 150872381 | 159.08 | 177 | 1.64E-14 | 4.00E-02 | 9.33E-04 | 1.10E-17 |
| | S4401 | 10 | 3 | 7 | 4 | 13835317 | 14009972 | 174.656 | 350 | 1.64E-14 | 4.00E-02 | 9.33E-04 | 1.10E-17 |
| | S4412 | 10 | 4 | 6 | 5 | 122198652 | 122478049 | 279.398 | 285 | 1.54E-27 | 5.33E-02 | 7.99E-04 | 1.04E-30 |
| | S4413 | 10 | 5 | 5 | 5 | 124279867 | 124566139 | 286.273 | 510 | 5.77E-45 | 6.67E-02 | 6.66E-04 | 3.87E-48 |
| | S4414 | 10 | 4 | 6 | 5 | 125630120 | 125804876 | 174.757 | 396 | 1.54E-27 | 5.33E-02 | 7.99E-04 | 1.04E-30 |
| | S4417 | 10 | 3 | 7 | 5 | 134230104 | 135130379 | 900.276 | 1001 | 1.64E-14 | 4.00E-02 | 9.33E-04 | 1.10E-17 |
| | S4420 | 10 | 4 | 6 | 6 | 39932543 | 40144923 | 212.381 | 376 | 1.54E-27 | 5.33E-02 | 7.99E-04 | 1.04E-30 |
| | S4426 | 10 | 2 | 8 | 6 | 99841307 | 100029426 | 188.12 | 309 | 7.49E-06 | 2.67E-02 | 1.07E-03 | 5.02E-09 |
| | S4429 | 10 | 4 | 6 | 6 | 158887949 | 159197271 | 309.323 | 481 | 1.54E-27 | 5.33E-02 | 7.99E-04 | 1.04E-30 |

| | | | | | | | | | | | | | |
|---|---|---|---|---|---|---|---|---|---|---|---|---|---|
| | S4431 | 10 | 3 | 7 | 7 | 1224511 | 2672318 | 1447.808 | 2986 | 1.64E-14 | 4.00E-02 | 9.33E-04 | 1.10E-17 |
| | S4432 | 10 | 3 | 7 | 7 | 10356343 | 10592588 | 236.246 | 644 | 1.64E-14 | 4.00E-02 | 9.33E-04 | 1.10E-17 |
| | S4433 | 10 | 2 | 8 | 7 | 21078350 | 21675136 | 596.787 | 1551 | 7.49E-06 | 2.67E-02 | 1.07E-03 | 5.02E-09 |
| | S4434 | 10 | 3 | 7 | 7 | 22240125 | 22384820 | 144.696 | 284 | 1.64E-14 | 4.00E-02 | 9.33E-04 | 1.10E-17 |
| | S4442 | 10 | 3 | 7 | 7 | 128941276 | 129198928 | 257.653 | 414 | 1.64E-14 | 4.00E-02 | 9.33E-04 | 1.10E-17 |
| | S4456 | 10 | 2 | 8 | 8 | 122658757 | 122769894 | 111.138 | 158 | 7.49E-06 | 2.67E-02 | 1.07E-03 | 5.02E-09 |
| | S4457 | 10 | 3 | 7 | 8 | 127792869 | 128160234 | 367.366 | 697 | 1.64E-14 | 4.00E-02 | 9.33E-04 | 1.10E-17 |
| | S4459 | 10 | 2 | 8 | 8 | 135948293 | 136260338 | 312.046 | 596 | 7.49E-06 | 2.67E-02 | 1.07E-03 | 5.02E-09 |
| | S4461 | 10 | 2 | 8 | 9 | 10963411 | 11083979 | 120.569 | 352 | 7.49E-06 | 2.67E-02 | 1.07E-03 | 5.02E-09 |
| | S4465 | 10 | 5 | 5 | 9 | 74714451 | 74845691 | 131.241 | 189 | 5.77E-45 | 6.67E-02 | 6.66E-04 | 3.87E-48 |
| | S4467 | 10 | 2 | 8 | 9 | 86521274 | 86833583 | 312.31 | 492 | 7.49E-06 | 2.67E-02 | 1.07E-03 | 5.02E-09 |
| | S4469 | 10 | 3 | 7 | 9 | 98217063 | 98402965 | 185.903 | 417 | 1.64E-14 | 4.00E-02 | 9.33E-04 | 1.10E-17 |
| | S4479 | 10 | 2 | 8 | 10 | 119296873 | 119821778 | 524.906 | 965 | 7.49E-06 | 2.67E-02 | 1.07E-03 | 5.02E-09 |
| | S4480 | 10 | 2 | 8 | 11 | 4427419 | 5075101 | 647.683 | 1628 | 7.49E-06 | 2.67E-02 | 1.07E-03 | 5.02E-09 |

| | | | | | | | | | | | | | |
|---|---|---|---|---|---|---|---|---|---|---|---|---|---|
| | S4484 | 10 | 3 | 7 | 11 | 22312503 | 22625424 | 312.922 | 576 | 1.64E-14 | 4.00E-02 | 9.33E-04 | 1.10E-17 |
| | S4485 | 10 | 2 | 8 | 11 | 23395219 | 23499332 | 104.114 | 282 | 7.49E-06 | 2.67E-02 | 1.07E-03 | 5.02E-09 |
| | S4489 | 10 | 2 | 8 | 11 | 92580342 | 92872820 | 292.479 | 405 | 7.49E-06 | 2.67E-02 | 1.07E-03 | 5.02E-09 |
| | S4490 | 10 | 2 | 8 | 11 | 94744147 | 94890796 | 146.65 | 108 | 7.49E-06 | 2.67E-02 | 1.07E-03 | 5.02E-09 |
| | S4494 | 10 | 3 | 7 | 12 | 23086330 | 23494197 | 407.868 | 504 | 1.64E-14 | 4.00E-02 | 9.33E-04 | 1.10E-17 |
| | S4495 | 10 | 3 | 7 | 12 | 23826293 | 24187936 | 361.644 | 723 | 1.64E-14 | 4.00E-02 | 9.33E-04 | 1.10E-17 |
| | S4496 | 10 | 3 | 7 | 12 | 24203498 | 24393854 | 190.357 | 399 | 1.64E-14 | 4.00E-02 | 9.33E-04 | 1.10E-17 |
| | S4499 | 10 | 2 | 8 | 12 | 64210920 | 64467462 | 256.543 | 353 | 7.49E-06 | 2.67E-02 | 1.07E-03 | 5.02E-09 |
| | S4506 | 10 | 2 | 8 | 12 | 96432833 | 96763449 | 330.617 | 699 | 7.49E-06 | 2.67E-02 | 1.07E-03 | 5.02E-09 |
| | S4513 | 10 | 4 | 6 | 12 | 114833879 | 115016132 | 182.254 | 360 | 1.54E-27 | 5.33E-02 | 7.99E-04 | 1.04E-30 |
| | S4514 | 10 | 2 | 8 | 12 | 125196264 | 125345986 | 149.723 | 276 | 7.49E-06 | 2.67E-02 | 1.07E-03 | 5.02E-09 |
| | S4516 | 10 | 4 | 6 | 13 | 90667118 | 90802500 | 135.383 | 252 | 1.54E-27 | 5.33E-02 | 7.99E-04 | 1.04E-30 |
| | S4519 | 10 | 3 | 7 | 13 | 101094089 | 101653039 | 558.951 | 1137 | 1.64E-14 | 4.00E-02 | 9.33E-04 | 1.10E-17 |
| | S4520 | 10 | 2 | 8 | 14 | 37036574 | 37593915 | 557.342 | 589 | 7.49E-06 | 2.67E-02 | 1.07E-03 | 5.02E-09 |

| | | | | | | | | | | | | | |
|---|---|---|---|---|---|---|---|---|---|---|---|---|---|
| | S4521 | 10 | 3 | 7 | 14 | 69909652 | 70187041 | 277.39 | 686 | 1.64E-14 | 4.00E-02 | 9.33E-04 | 1.10E-17 |
| | S4528 | 10 | 5 | 5 | 15 | 66746574 | 66932154 | 185.581 | 352 | 5.77E-45 | 6.67E-02 | 6.66E-04 | 3.87E-48 |
| | S4529 | 10 | 5 | 5 | 15 | 66932872 | 67118464 | 185.593 | 385 | 5.77E-45 | 6.67E-02 | 6.66E-04 | 3.87E-48 |
| | S4535 | 10 | 3 | 7 | 16 | 24668787 | 25063831 | 395.045 | 671 | 1.64E-14 | 4.00E-02 | 9.33E-04 | 1.10E-17 |
| | S4537 | 10 | 5 | 5 | 16 | 26937027 | 27162303 | 225.277 | 568 | 5.77E-45 | 6.67E-02 | 6.66E-04 | 3.87E-48 |
| | S4538 | 10 | 4 | 6 | 16 | 53134146 | 53342939 | 208.794 | 264 | 1.54E-27 | 5.33E-02 | 7.99E-04 | 1.04E-30 |
| | S4539 | 10 | 4 | 6 | 16 | 56338927 | 56929410 | 590.484 | 1140 | 1.54E-27 | 5.33E-02 | 7.99E-04 | 1.04E-30 |
| | S4540 | 10 | 5 | 5 | 16 | 57412095 | 57855908 | 443.814 | 847 | 5.77E-45 | 6.67E-02 | 6.66E-04 | 3.87E-48 |
| | S4549 | 10 | 2 | 8 | 18 | 51453244 | 51588653 | 135.41 | 199 | 7.49E-06 | 2.67E-02 | 1.07E-03 | 5.02E-09 |
| | S4552 | 10 | 4 | 6 | 19 | 31082574 | 31630862 | 548.289 | 778 | 1.54E-27 | 5.33E-02 | 7.99E-04 | 1.04E-30 |
| | S4553 | 10 | 5 | 5 | 19 | 31928119 | 32439163 | 511.045 | 854 | 5.77E-45 | 6.67E-02 | 6.66E-04 | 3.87E-48 |
| | S4557 | 10 | 2 | 8 | 19 | 39128280 | 39241322 | 113.043 | 182 | 7.49E-06 | 2.67E-02 | 1.07E-03 | 5.02E-09 |
| | S4562 | 9 | 5 | 4 | 1 | 18808720 | 19714095 | 905.376 | 2184 | 5.66E-50 | 6.67E-02 | 5.33E-04 | 3.79E-53 |
| | S4563 | 9 | 3 | 6 | 1 | 21530637 | 22019906 | 489.27 | 1353 | 4.50E-16 | 4.00E-02 | 7.99E-04 | 3.02E-19 |

| | | | | | | | | | | | | | |
|---|---|---|---|---|---|---|---|---|---|---|---|---|---|
| | S4566 | 9 | 2 | 7 | 1 | 36598595 | 36701114 | 102.52 | 159 | 1.99E-06 | 2.67E-02 | 9.33E-04 | 1.33E-09 |
| | S4567 | 9 | 2 | 7 | 1 | 43659475 | 43877501 | 218.027 | 356 | 1.99E-06 | 2.67E-02 | 9.33E-04 | 1.33E-09 |
| | S4568 | 9 | 2 | 7 | 1 | 57144256 | 57659636 | 515.381 | 1034 | 1.99E-06 | 2.67E-02 | 9.33E-04 | 1.33E-09 |
| | S4569 | 9 | 5 | 4 | 1 | 66482157 | 67301843 | 819.687 | 1798 | 5.66E-50 | 6.67E-02 | 5.33E-04 | 3.79E-53 |
| | S4571 | 9 | 4 | 5 | 1 | 110486865 | 110661096 | 174.232 | 365 | 1.41E-30 | 5.33E-02 | 6.66E-04 | 9.47E-34 |
| | S4572 | 9 | 3 | 6 | 1 | 167228524 | 167749038 | 520.515 | 1088 | 4.50E-16 | 4.00E-02 | 7.99E-04 | 3.02E-19 |
| | S4574 | 9 | 3 | 6 | 1 | 199645702 | 200047874 | 402.173 | 907 | 4.50E-16 | 4.00E-02 | 7.99E-04 | 3.02E-19 |
| | S4580 | 9 | 2 | 7 | 2 | 44183401 | 44309494 | 126.094 | 201 | 1.99E-06 | 2.67E-02 | 9.33E-04 | 1.33E-09 |
| | S4592 | 9 | 3 | 6 | 3 | 14667411 | 15272505 | 605.095 | 977 | 4.50E-16 | 4.00E-02 | 7.99E-04 | 3.02E-19 |
| | S4593 | 9 | 3 | 6 | 3 | 15290398 | 16112722 | 822.325 | 1437 | 4.50E-16 | 4.00E-02 | 7.99E-04 | 3.02E-19 |
| | S4596 | 9 | 2 | 7 | 3 | 24314143 | 24504776 | 190.634 | 293 | 1.99E-06 | 2.67E-02 | 9.33E-04 | 1.33E-09 |
| | S4602 | 9 | 3 | 6 | 3 | 70398296 | 71000213 | 601.918 | 830 | 4.50E-16 | 4.00E-02 | 7.99E-04 | 3.02E-19 |
| | S4603 | 9 | 3 | 6 | 3 | 104575557 | 104676465 | 100.909 | 183 | 4.50E-16 | 4.00E-02 | 7.99E-04 | 3.02E-19 |
| | S4605 | 9 | 2 | 7 | 3 | 104845312 | 105113360 | 268.049 | 632 | 1.99E-06 | 2.67E-02 | 9.33E-04 | 1.33E-09 |

| | | | | | | | | | | | | | |
|---|---|---|---|---|---|---|---|---|---|---|---|---|---|
| | S4606 | 9 | 2 | 7 | 3 | 105856566 | 106105366 | 248.801 | 388 | 1.99E-06 | 2.67E-02 | 9.33E-04 | 1.33E-09 |
| | S4607 | 9 | 2 | 7 | 3 | 106200948 | 106406190 | 205.243 | 335 | 1.99E-06 | 2.67E-02 | 9.33E-04 | 1.33E-09 |
| | S4610 | 9 | 3 | 6 | 3 | 113967463 | 114235308 | 267.846 | 377 | 4.50E-16 | 4.00E-02 | 7.99E-04 | 3.02E-19 |
| | S4611 | 9 | 3 | 6 | 3 | 114747253 | 114850556 | 103.304 | 121 | 4.50E-16 | 4.00E-02 | 7.99E-04 | 3.02E-19 |
| | S4612 | 9 | 3 | 6 | 3 | 114927416 | 115210665 | 283.25 | 232 | 4.50E-16 | 4.00E-02 | 7.99E-04 | 3.02E-19 |
| | S4614 | 9 | 3 | 6 | 3 | 126167480 | 126377383 | 209.904 | 466 | 4.50E-16 | 4.00E-02 | 7.99E-04 | 3.02E-19 |
| | S4615 | 9 | 3 | 6 | 3 | 149949639 | 150244227 | 294.589 | 539 | 4.50E-16 | 4.00E-02 | 7.99E-04 | 3.02E-19 |
| | S4620 | 9 | 2 | 7 | 4 | 20897300 | 21145173 | 247.874 | 534 | 1.99E-06 | 2.67E-02 | 9.33E-04 | 1.33E-09 |
| | S4622 | 9 | 3 | 6 | 4 | 54850038 | 55016727 | 166.69 | 278 | 4.50E-16 | 4.00E-02 | 7.99E-04 | 3.02E-19 |
| | S4624 | 9 | 2 | 7 | 4 | 163334277 | 163496093 | 161.817 | 304 | 1.99E-06 | 2.67E-02 | 9.33E-04 | 1.33E-09 |
| | S4628 | 9 | 2 | 7 | 5 | 76921710 | 77378485 | 456.776 | 833 | 1.99E-06 | 2.67E-02 | 9.33E-04 | 1.33E-09 |
| | S4630 | 9 | 3 | 6 | 5 | 79805294 | 80260350 | 455.057 | 940 | 4.50E-16 | 4.00E-02 | 7.99E-04 | 3.02E-19 |
| | S4632 | 9 | 4 | 5 | 5 | 124578128 | 125507383 | 929.256 | 1900 | 1.41E-30 | 5.33E-02 | 6.66E-04 | 9.47E-34 |
| | S4634 | 9 | 2 | 7 | 5 | 159708201 | 159855895 | 147.695 | 227 | 1.99E-06 | 2.67E-02 | 9.33E-04 | 1.33E-09 |

| | | | | | | | | | | | | | |
|---|---|---|---|---|---|---|---|---|---|---|---|---|---|
| | S4635 | 9 | 3 | 6 | 5 | 159869312 | 160150806 | 281.495 | 376 | 4.50E-16 | 4.00E-02 | 7.99E-04 | 3.02E-19 |
| | S4636 | 9 | 3 | 6 | 5 | 163697822 | 164049107 | 351.286 | 728 | 4.50E-16 | 4.00E-02 | 7.99E-04 | 3.02E-19 |
| | S4638 | 9 | 2 | 7 | 6 | 21818985 | 22058588 | 239.604 | 480 | 1.99E-06 | 2.67E-02 | 9.33E-04 | 1.33E-09 |
| | S4639 | 9 | 2 | 7 | 6 | 22960022 | 23228668 | 268.647 | 486 | 1.99E-06 | 2.67E-02 | 9.33E-04 | 1.33E-09 |
| | S4642 | 9 | 3 | 6 | 6 | 40904167 | 41338238 | 434.072 | 655 | 4.50E-16 | 4.00E-02 | 7.99E-04 | 3.02E-19 |
| | S4645 | 9 | 4 | 5 | 6 | 147718119 | 147995687 | 277.569 | 642 | 1.41E-30 | 5.33E-02 | 6.66E-04 | 9.47E-34 |
| | S4647 | 9 | 3 | 6 | 6 | 151376381 | 151902397 | 526.017 | 960 | 4.50E-16 | 4.00E-02 | 7.99E-04 | 3.02E-19 |
| | S4648 | 9 | 5 | 4 | 6 | 164602133 | 165511640 | 909.508 | 2158 | 5.66E-50 | 6.67E-02 | 5.33E-04 | 3.79E-53 |
| | S4649 | 9 | 3 | 6 | 7 | 4637402 | 4831264 | 193.863 | 299 | 4.50E-16 | 4.00E-02 | 7.99E-04 | 3.02E-19 |
| | S4650 | 9 | 3 | 6 | 7 | 4847612 | 5101392 | 253.781 | 665 | 4.50E-16 | 4.00E-02 | 7.99E-04 | 3.02E-19 |
| | S4651 | 9 | 4 | 5 | 7 | 14149680 | 14346907 | 197.228 | 412 | 1.41E-30 | 5.33E-02 | 6.66E-04 | 9.47E-34 |
| | S4652 | 9 | 2 | 7 | 7 | 19611482 | 20058372 | 446.891 | 994 | 1.99E-06 | 2.67E-02 | 9.33E-04 | 1.33E-09 |
| | S4654 | 9 | 2 | 7 | 7 | 32418271 | 33234842 | 816.572 | 1790 | 1.99E-06 | 2.67E-02 | 9.33E-04 | 1.33E-09 |
| | S4655 | 9 | 2 | 7 | 7 | 33332922 | 33507007 | 174.086 | 350 | 1.99E-06 | 2.67E-02 | 9.33E-04 | 1.33E-09 |

| | | | | | | | | | | | | | |
|---|---|---|---|---|---|---|---|---|---|---|---|---|---|
| | S4661 | 9 | 4 | 5 | 7 | 128381802 | 128733088 | 351.287 | 828 | 1.41E-30 | 5.33E-02 | 6.66E-04 | 9.47E-34 |
| | S4672 | 9 | 2 | 7 | 8 | 127389769 | 127519182 | 129.414 | 314 | 1.99E-06 | 2.67E-02 | 9.33E-04 | 1.33E-09 |
| | S4674 | 9 | 2 | 7 | 8 | 127557457 | 127788701 | 231.245 | 431 | 1.99E-06 | 2.67E-02 | 9.33E-04 | 1.33E-09 |
| | S4679 | 9 | 4 | 5 | 9 | 75254932 | 75435447 | 180.516 | 385 | 1.41E-30 | 5.33E-02 | 6.66E-04 | 9.47E-34 |
| | S4682 | 9 | 2 | 7 | 10 | 8740581 | 9163012 | 422.432 | 804 | 1.99E-06 | 2.67E-02 | 9.33E-04 | 1.33E-09 |
| | S4683 | 9 | 2 | 7 | 10 | 9646289 | 9907872 | 261.584 | 459 | 1.99E-06 | 2.67E-02 | 9.33E-04 | 1.33E-09 |
| | S4685 | 9 | 2 | 7 | 10 | 59823437 | 60007089 | 183.653 | 328 | 1.99E-06 | 2.67E-02 | 9.33E-04 | 1.33E-09 |
| | S4686 | 9 | 4 | 5 | 10 | 97140410 | 97577159 | 436.75 | 704 | 1.41E-30 | 5.33E-02 | 6.66E-04 | 9.47E-34 |
| | S4687 | 9 | 4 | 5 | 10 | 97608158 | 97730376 | 122.219 | 182 | 1.41E-30 | 5.33E-02 | 6.66E-04 | 9.47E-34 |
| | S4688 | 9 | 5 | 4 | 10 | 97732914 | 97873937 | 141.024 | 167 | 5.66E-50 | 6.67E-02 | 5.33E-04 | 3.79E-53 |
| | S4689 | 9 | 2 | 7 | 10 | 121705316 | 122133359 | 428.044 | 1000 | 1.99E-06 | 2.67E-02 | 9.33E-04 | 1.33E-09 |
| | S4694 | 9 | 2 | 7 | 11 | 79503121 | 79696782 | 193.662 | 359 | 1.99E-06 | 2.67E-02 | 9.33E-04 | 1.33E-09 |
| | S4696 | 9 | 3 | 6 | 11 | 99742967 | 99844216 | 101.25 | 286 | 4.50E-16 | 4.00E-02 | 7.99E-04 | 3.02E-19 |
| | S4699 | 9 | 3 | 6 | 12 | 16365732 | 16583317 | 217.586 | 300 | 4.50E-16 | 4.00E-02 | 7.99E-04 | 3.02E-19 |

| | | | | | | | | | | | | | |
|---|---|---|---|---|---|---|---|---|---|---|---|---|---|
| | S4709 | 9 | 4 | 5 | 12 | 114394173 | 114547018 | 152.846 | 289 | 1.41E-30 | 5.33E-02 | 6.66E-04 | 9.47E-34 |
| | S4710 | 9 | 4 | 5 | 12 | 115867416 | 116351437 | 484.022 | 688 | 1.41E-30 | 5.33E-02 | 6.66E-04 | 9.47E-34 |
| | S4711 | 9 | 4 | 5 | 12 | 116365752 | 116486938 | 121.187 | 266 | 1.41E-30 | 5.33E-02 | 6.66E-04 | 9.47E-34 |
| | S4717 | 9 | 2 | 7 | 13 | 44669217 | 44779412 | 110.196 | 152 | 1.99E-06 | 2.67E-02 | 9.33E-04 | 1.33E-09 |
| | S4725 | 9 | 3 | 6 | 14 | 91226541 | 91747623 | 521.083 | 977 | 4.50E-16 | 4.00E-02 | 7.99E-04 | 3.02E-19 |
| | S4727 | 9 | 3 | 6 | 14 | 94809152 | 95311752 | 502.601 | 1121 | 4.50E-16 | 4.00E-02 | 7.99E-04 | 3.02E-19 |
| | S4730 | 9 | 5 | 4 | 16 | 16854795 | 17110288 | 255.494 | 454 | 5.66E-50 | 6.67E-02 | 5.33E-04 | 3.79E-53 |
| | S4731 | 9 | 3 | 6 | 16 | 19279092 | 19384618 | 105.527 | 226 | 4.50E-16 | 4.00E-02 | 7.99E-04 | 3.02E-19 |
| | S4732 | 9 | 5 | 4 | 16 | 27223625 | 27840555 | 616.931 | 818 | 5.66E-50 | 6.67E-02 | 5.33E-04 | 3.79E-53 |
| | S4733 | 9 | 4 | 5 | 16 | 52757088 | 52945399 | 188.312 | 335 | 1.41E-30 | 5.33E-02 | 6.66E-04 | 9.47E-34 |
| | S4734 | 9 | 3 | 6 | 16 | 55188902 | 55673342 | 484.441 | 858 | 4.50E-16 | 4.00E-02 | 7.99E-04 | 3.02E-19 |
| | S4736 | 9 | 2 | 7 | 16 | 78094370 | 78606465 | 512.096 | 1753 | 1.99E-06 | 2.67E-02 | 9.33E-04 | 1.33E-09 |
| | S4738 | 9 | 3 | 6 | 18 | 26520059 | 26782785 | 262.727 | 518 | 4.50E-16 | 4.00E-02 | 7.99E-04 | 3.02E-19 |
| | S4746 | 9 | 3 | 6 | 20 | 18485702 | 18914007 | 428.306 | 920 | 4.50E-16 | 4.00E-02 | 7.99E-04 | 3.02E-19 |

| | | | | | | | | | | | | | |
|---|---|---|---|---|---|---|---|---|---|---|---|---|---|
| | S4749 | 9 | 3 | 6 | 20 | 24078358 | 24280743 | 202.386 | 345 | 4.50E-16 | 4.00E-02 | 7.99E-04 | 3.02E-19 |
| | S4750 | 9 | 3 | 6 | 21 | 33354925 | 33852066 | 497.142 | 667 | 4.50E-16 | 4.00E-02 | 7.99E-04 | 3.02E-19 |
| | S4751 | 9 | 3 | 6 | 22 | 33213881 | 33356446 | 142.566 | 228 | 4.50E-16 | 4.00E-02 | 7.99E-04 | 3.02E-19 |
| | S4753 | 9 | 2 | 7 | 22 | 47270455 | 47878603 | 608.149 | 1339 | 1.99E-06 | 2.67E-02 | 9.33E-04 | 1.33E-09 |
| | S4754 | 8 | 4 | 4 | 1 | 6601672 | 7187275 | 585.604 | 1273 | 2.26E-34 | 5.33E-02 | 5.33E-04 | 1.52E-37 |
| | S4757 | 8 | 2 | 6 | 1 | 38820417 | 38954990 | 134.574 | 313 | 3.81E-07 | 2.67E-02 | 7.99E-04 | 2.56E-10 |
| | S4758 | 8 | 2 | 6 | 1 | 38973972 | 39811650 | 837.679 | 1602 | 3.81E-07 | 2.67E-02 | 7.99E-04 | 2.56E-10 |
| | S4761 | 8 | 4 | 4 | 1 | 109905062 | 110381992 | 476.931 | 786 | 2.26E-34 | 5.33E-02 | 5.33E-04 | 1.52E-37 |
| | S4763 | 8 | 3 | 5 | 1 | 200181610 | 200447162 | 265.553 | 441 | 5.06E-18 | 4.00E-02 | 6.66E-04 | 3.40E-21 |
| | S4765 | 8 | 2 | 6 | 1 | 207118477 | 207969549 | 851.073 | 1522 | 3.81E-07 | 2.67E-02 | 7.99E-04 | 2.56E-10 |
| | S4769 | 8 | 2 | 6 | 2 | 1797292 | 2003713 | 206.422 | 402 | 3.81E-07 | 2.67E-02 | 7.99E-04 | 2.56E-10 |
| | S4771 | 8 | 2 | 6 | 2 | 2247281 | 2532844 | 285.564 | 536 | 3.81E-07 | 2.67E-02 | 7.99E-04 | 2.56E-10 |
| | S4778 | 8 | 2 | 6 | 3 | 30227026 | 30682944 | 455.919 | 925 | 3.81E-07 | 2.67E-02 | 7.99E-04 | 2.56E-10 |
| | S4780 | 8 | 2 | 6 | 3 | 54541513 | 55315794 | 774.282 | 1544 | 3.81E-07 | 2.67E-02 | 7.99E-04 | 2.56E-10 |

| | | | | | | | | | | | | | |
|---|---|---|---|---|---|---|---|---|---|---|---|---|---|
| | S4781 | 8 | 3 | 5 | 3 | 60424973 | 60591036 | 166.064 | 485 | 5.06E-18 | 4.00E-02 | 6.66E-04 | 3.40E-21 |
| | S4782 | 8 | 3 | 5 | 3 | 66660387 | 66856351 | 195.965 | 440 | 5.06E-18 | 4.00E-02 | 6.66E-04 | 3.40E-21 |
| | S4783 | 8 | 3 | 5 | 3 | 67924310 | 68164445 | 240.136 | 395 | 5.06E-18 | 4.00E-02 | 6.66E-04 | 3.40E-21 |
| | S4784 | 8 | 3 | 5 | 3 | 68285725 | 69387118 | 1101.394 | 1919 | 5.06E-18 | 4.00E-02 | 6.66E-04 | 3.40E-21 |
| | S4785 | 8 | 2 | 6 | 3 | 75762886 | 76444458 | 681.573 | 975 | 3.81E-07 | 2.67E-02 | 7.99E-04 | 2.56E-10 |
| | S4786 | 8 | 4 | 4 | 3 | 175327907 | 175534037 | 206.131 | 406 | 2.26E-34 | 5.33E-02 | 5.33E-04 | 1.52E-37 |
| | S4789 | 8 | 2 | 6 | 4 | 82172483 | 82695162 | 522.68 | 947 | 3.81E-07 | 2.67E-02 | 7.99E-04 | 2.56E-10 |
| | S4797 | 8 | 2 | 6 | 5 | 76506020 | 76662097 | 156.078 | 309 | 3.81E-07 | 2.67E-02 | 7.99E-04 | 2.56E-10 |
| | S4800 | 8 | 4 | 4 | 5 | 145936404 | 146357478 | 421.075 | 574 | 2.26E-34 | 5.33E-02 | 5.33E-04 | 1.52E-37 |
| | S4803 | 8 | 3 | 5 | 6 | 6226151 | 6593462 | 367.312 | 748 | 5.06E-18 | 4.00E-02 | 6.66E-04 | 3.40E-21 |
| | S4804 | 8 | 4 | 4 | 6 | 7006326 | 7232156 | 225.831 | 390 | 2.26E-34 | 5.33E-02 | 5.33E-04 | 1.52E-37 |
| | S4805 | 8 | 4 | 4 | 6 | 7569093 | 7761495 | 192.403 | 414 | 2.26E-34 | 5.33E-02 | 5.33E-04 | 1.52E-37 |
| | S4806 | 8 | 4 | 4 | 6 | 7772466 | 8376017 | 603.552 | 1269 | 2.26E-34 | 5.33E-02 | 5.33E-04 | 1.52E-37 |
| | S4807 | 8 | 4 | 4 | 6 | 8378389 | 8887404 | 509.016 | 1051 | 2.26E-34 | 5.33E-02 | 5.33E-04 | 1.52E-37 |

| | | | | | | | | | | | | | |
|---|---|---|---|---|---|---|---|---|---|---|---|---|---|
| | S4817 | 8 | 4 | 4 | 6 | 137283083 | 138644122 | 1361.04 | 2503 | 2.26E-34 | 5.33E-02 | 5.33E-04 | 1.52E-37 |
| | S4827 | 8 | 2 | 6 | 8 | 23837494 | 25028994 | 1191.501 | 1710 | 3.81E-07 | 2.67E-02 | 7.99E-04 | 2.56E-10 |
| | S4829 | 8 | 2 | 6 | 8 | 68212014 | 68314186 | 102.173 | 181 | 3.81E-07 | 2.67E-02 | 7.99E-04 | 2.56E-10 |
| | S4830 | 8 | 2 | 6 | 8 | 68372232 | 68973448 | 601.217 | 1041 | 3.81E-07 | 2.67E-02 | 7.99E-04 | 2.56E-10 |
| | S4831 | 8 | 2 | 6 | 8 | 123409792 | 123597973 | 188.182 | 466 | 3.81E-07 | 2.67E-02 | 7.99E-04 | 2.56E-10 |
| | S4832 | 8 | 2 | 6 | 8 | 123681170 | 123853016 | 171.847 | 276 | 3.81E-07 | 2.67E-02 | 7.99E-04 | 2.56E-10 |
| | S4833 | 8 | 2 | 6 | 8 | 123900286 | 125259706 | 1359.421 | 2329 | 3.81E-07 | 2.67E-02 | 7.99E-04 | 2.56E-10 |
| | S4834 | 8 | 2 | 6 | 8 | 134563496 | 134814337 | 250.842 | 536 | 3.81E-07 | 2.67E-02 | 7.99E-04 | 2.56E-10 |
| | S4835 | 8 | 2 | 6 | 9 | 4740545 | 4883257 | 142.713 | 317 | 3.81E-07 | 2.67E-02 | 7.99E-04 | 2.56E-10 |
| | S4842 | 8 | 2 | 6 | 9 | 78349772 | 78998022 | 648.251 | 1141 | 3.81E-07 | 2.67E-02 | 7.99E-04 | 2.56E-10 |
| | S4848 | 8 | 2 | 6 | 9 | 113648961 | 113817969 | 169.009 | 345 | 3.81E-07 | 2.67E-02 | 7.99E-04 | 2.56E-10 |
| | S4851 | 8 | 2 | 6 | 9 | 126850851 | 127066219 | 215.369 | 293 | 3.81E-07 | 2.67E-02 | 7.99E-04 | 2.56E-10 |
| | S4856 | 8 | 3 | 5 | 10 | 76388111 | 76900800 | 512.69 | 703 | 5.06E-18 | 4.00E-02 | 6.66E-04 | 3.40E-21 |
| | S4858 | 8 | 2 | 6 | 10 | 116756707 | 116971850 | 215.144 | 227 | 3.81E-07 | 2.67E-02 | 7.99E-04 | 2.56E-10 |

| | | | | | | | | | | | | | |
|---|---|---|---|---|---|---|---|---|---|---|---|---|---|
| | S4861 | 8 | 3 | 5 | 11 | 34628852 | 35482083 | 853.232 | 1728 | 5.06E-18 | 4.00E-02 | 6.66E-04 | 3.40E-21 |
| | S4862 | 8 | 3 | 5 | 11 | 61463423 | 61865710 | 402.288 | 458 | 5.06E-18 | 4.00E-02 | 6.66E-04 | 3.40E-21 |
| | S4865 | 8 | 2 | 6 | 11 | 95072383 | 95174814 | 102.432 | 195 | 3.81E-07 | 2.67E-02 | 7.99E-04 | 2.56E-10 |
| | S4868 | 8 | 2 | 6 | 12 | 11231190 | 11542930 | 311.741 | 460 | 3.81E-07 | 2.67E-02 | 7.99E-04 | 2.56E-10 |
| | S4870 | 8 | 2 | 6 | 12 | 24912795 | 25543144 | 630.35 | 1474 | 3.81E-07 | 2.67E-02 | 7.99E-04 | 2.56E-10 |
| | S4873 | 8 | 4 | 4 | 12 | 116519565 | 116631678 | 112.114 | 225 | 2.26E-34 | 5.33E-02 | 5.33E-04 | 1.52E-37 |
| | S4876 | 8 | 2 | 6 | 13 | 44887231 | 46175106 | 1287.876 | 1862 | 3.81E-07 | 2.67E-02 | 7.99E-04 | 2.56E-10 |
| | S4888 | 8 | 3 | 5 | 15 | 23960958 | 25248109 | 1287.152 | 1985 | 5.06E-18 | 4.00E-02 | 6.66E-04 | 3.40E-21 |
| | S4890 | 8 | 4 | 4 | 15 | 61048372 | 61374329 | 325.958 | 531 | 2.26E-34 | 5.33E-02 | 5.33E-04 | 1.52E-37 |
| | S4891 | 8 | 4 | 4 | 15 | 61540658 | 61835798 | 295.141 | 568 | 2.26E-34 | 5.33E-02 | 5.33E-04 | 1.52E-37 |
| | S4892 | 8 | 5 | 3 | 15 | 62261912 | 62400579 | 138.668 | 302 | 3.12E-56 | 6.67E-02 | 4.00E-04 | 2.09E-59 |
| | S4895 | 8 | 5 | 3 | 15 | 68589185 | 68724294 | 135.11 | 215 | 3.12E-56 | 6.67E-02 | 4.00E-04 | 2.09E-59 |
| | S4896 | 8 | 2 | 6 | 15 | 97302448 | 97534348 | 231.901 | 455 | 3.81E-07 | 2.67E-02 | 7.99E-04 | 2.56E-10 |
| | S4898 | 8 | 3 | 5 | 16 | 12209094 | 12515873 | 306.78 | 617 | 5.06E-18 | 4.00E-02 | 6.66E-04 | 3.40E-21 |

| | | | | | | | | | | | | | |
|---|---|---|---|---|---|---|---|---|---|---|---|---|---|
| | S4900 | 8 | 4 | 4 | 16 | 62922806 | 63074628 | 151.823 | 250 | 2.26E-34 | 5.33E-02 | 5.33E-04 | 1.52E-37 |
| | S4901 | 8 | 2 | 6 | 16 | 80276142 | 81134969 | 858.828 | 1923 | 3.81E-07 | 2.67E-02 | 7.99E-04 | 2.56E-10 |
| | S4907 | 8 | 3 | 5 | 18 | 13572355 | 14132941 | 560.587 | 1012 | 5.06E-18 | 4.00E-02 | 6.66E-04 | 3.40E-21 |
| | S4909 | 8 | 2 | 6 | 18 | 25631759 | 26459190 | 827.432 | 1303 | 3.81E-07 | 2.67E-02 | 7.99E-04 | 2.56E-10 |
| | S4911 | 8 | 2 | 6 | 18 | 57854558 | 58115947 | 261.39 | 509 | 3.81E-07 | 2.67E-02 | 7.99E-04 | 2.56E-10 |
| | S4912 | 8 | 2 | 6 | 18 | 58117197 | 59191972 | 1074.776 | 2069 | 3.81E-07 | 2.67E-02 | 7.99E-04 | 2.56E-10 |
| | S4913 | 8 | 3 | 5 | 18 | 60044465 | 60425987 | 381.523 | 538 | 5.06E-18 | 4.00E-02 | 6.66E-04 | 3.40E-21 |
| | S4915 | 8 | 3 | 5 | 19 | 4976401 | 5245619 | 269.219 | 664 | 5.06E-18 | 4.00E-02 | 6.66E-04 | 3.40E-21 |
| | S4916 | 8 | 3 | 5 | 19 | 32874928 | 32982129 | 107.202 | 302 | 5.06E-18 | 4.00E-02 | 6.66E-04 | 3.40E-21 |
| | S4923 | 8 | 3 | 5 | 20 | 46952464 | 47087472 | 135.009 | 243 | 5.06E-18 | 4.00E-02 | 6.66E-04 | 3.40E-21 |
| | S4925 | 8 | 2 | 6 | 20 | 52062666 | 53169564 | 1106.899 | 1851 | 3.81E-07 | 2.67E-02 | 7.99E-04 | 2.56E-10 |
| | S4929 | 8 | 2 | 6 | 21 | 43836504 | 44092475 | 255.972 | 396 | 3.81E-07 | 2.67E-02 | 7.99E-04 | 2.56E-10 |
| | S4934 | 8 | 2 | 6 | 22 | 39362248 | 39570681 | 208.434 | 266 | 3.81E-07 | 2.67E-02 | 7.99E-04 | 2.56E-10 |
| | S4936 | 7 | 3 | 4 | 1 | 9989294 | 10944390 | 955.097 | 1856 | 1.59E-20 | 4.00E-02 | 5.33E-04 | 1.07E-23 |

| | | | | | | | | | | | | | |
|---|---|---|---|---|---|---|---|---|---|---|---|---|---|
| | S4940 | 7 | 2 | 5 | 1 | 30133716 | 30483565 | 349.85 | 797 | 4.60E-08 | 2.67E-02 | 6.66E-04 | 3.09E-11 |
| | S4948 | 7 | 2 | 5 | 1 | 238558740 | 238747202 | 188.463 | 384 | 4.60E-08 | 2.67E-02 | 6.66E-04 | 3.09E-11 |
| | S4953 | 7 | 2 | 5 | 2 | 191627083 | 191733298 | 106.216 | 156 | 4.60E-08 | 2.67E-02 | 6.66E-04 | 3.09E-11 |
| | S4958 | 7 | 2 | 5 | 3 | 3516970 | 3658272 | 141.303 | 321 | 4.60E-08 | 2.67E-02 | 6.66E-04 | 3.09E-11 |
| | S4964 | 7 | 3 | 4 | 3 | 183467293 | 183579399 | 112.107 | 193 | 1.59E-20 | 4.00E-02 | 5.33E-04 | 1.07E-23 |
| | S4965 | 7 | 3 | 4 | 3 | 187063125 | 187580707 | 517.583 | 999 | 1.59E-20 | 4.00E-02 | 5.33E-04 | 1.07E-23 |
| | S4974 | 7 | 3 | 4 | 5 | 80420045 | 81166695 | 746.651 | 1526 | 1.59E-20 | 4.00E-02 | 5.33E-04 | 1.07E-23 |
| | S4975 | 7 | 3 | 4 | 5 | 81307822 | 81629911 | 322.09 | 500 | 1.59E-20 | 4.00E-02 | 5.33E-04 | 1.07E-23 |
| | S4978 | 7 | 2 | 5 | 5 | 164379548 | 164974265 | 594.718 | 822 | 4.60E-08 | 2.67E-02 | 6.66E-04 | 3.09E-11 |
| | S4979 | 7 | 2 | 5 | 5 | 165424979 | 165526110 | 101.132 | 186 | 4.60E-08 | 2.67E-02 | 6.66E-04 | 3.09E-11 |
| | S4984 | 7 | 4 | 3 | 6 | 8991328 | 9190384 | 199.057 | 319 | 3.00E-39 | 5.33E-02 | 4.00E-04 | 2.01E-42 |
| | S4991 | 7 | 3 | 4 | 6 | 138972226 | 139095603 | 123.378 | 234 | 1.59E-20 | 4.00E-02 | 5.33E-04 | 1.07E-23 |
| | S4992 | 7 | 2 | 5 | 6 | 148891314 | 149289626 | 398.313 | 761 | 4.60E-08 | 2.67E-02 | 6.66E-04 | 3.09E-11 |
| | S4994 | 7 | 2 | 5 | 6 | 155873417 | 156554841 | 681.425 | 1356 | 4.60E-08 | 2.67E-02 | 6.66E-04 | 3.09E-11 |

| | | | | | | | | | | | | | |
|---|---|---|---|---|---|---|---|---|---|---|---|---|---|
| | S4996 | 7 | 2 | 5 | 7 | 7096062 | 7401521 | 305.46 | 784 | 4.60E-08 | 2.67E-02 | 6.66E-04 | 3.09E-11 |
| | S4998 | 7 | 3 | 4 | 7 | 8213875 | 8715936 | 502.062 | 977 | 1.59E-20 | 4.00E-02 | 5.33E-04 | 1.07E-23 |
| | S5000 | 7 | 3 | 4 | 7 | 15103127 | 15582769 | 479.643 | 1019 | 1.59E-20 | 4.00E-02 | 5.33E-04 | 1.07E-23 |
| | S5001 | 7 | 3 | 4 | 7 | 18110576 | 18352248 | 241.673 | 442 | 1.59E-20 | 4.00E-02 | 5.33E-04 | 1.07E-23 |
| | S5004 | 7 | 3 | 4 | 7 | 93394948 | 94055956 | 661.009 | 1003 | 1.59E-20 | 4.00E-02 | 5.33E-04 | 1.07E-23 |
| | S5007 | 7 | 2 | 5 | 7 | 157593810 | 157740587 | 146.778 | 308 | 4.60E-08 | 2.67E-02 | 6.66E-04 | 3.09E-11 |
| | S5009 | 7 | 2 | 5 | 8 | 23635862 | 23768832 | 132.971 | 298 | 4.60E-08 | 2.67E-02 | 6.66E-04 | 3.09E-11 |
| | S5021 | 7 | 2 | 5 | 10 | 77417925 | 77865495 | 447.571 | 826 | 4.60E-08 | 2.67E-02 | 6.66E-04 | 3.09E-11 |
| | S5022 | 7 | 2 | 5 | 10 | 117437399 | 117731241 | 293.843 | 555 | 4.60E-08 | 2.67E-02 | 6.66E-04 | 3.09E-11 |
| | S5029 | 7 | 3 | 4 | 11 | 114388318 | 115822692 | 1434.375 | 2373 | 1.59E-20 | 4.00E-02 | 5.33E-04 | 1.07E-23 |
| | S5030 | 7 | 2 | 5 | 11 | 118955052 | 119125195 | 170.144 | 260 | 4.60E-08 | 2.67E-02 | 6.66E-04 | 3.09E-11 |
| | S5031 | 7 | 4 | 3 | 12 | 1010556 | 1804326 | 793.771 | 1395 | 3.00E-39 | 5.33E-02 | 4.00E-04 | 2.01E-42 |
| | S5033 | 7 | 3 | 4 | 12 | 20006480 | 20374978 | 368.499 | 705 | 1.59E-20 | 4.00E-02 | 5.33E-04 | 1.07E-23 |
| | S5035 | 7 | 2 | 5 | 12 | 53531587 | 53710439 | 178.853 | 279 | 4.60E-08 | 2.67E-02 | 6.66E-04 | 3.09E-11 |

| | | | | | | | | | | | | | |
|---|---|---|---|---|---|---|---|---|---|---|---|---|---|
| | S5040 | 7 | 2 | 5 | 12 | 118024123 | 118462612 | 438.49 | 711 | 4.60E-08 | 2.67E-02 | 6.66E-04 | 3.09E-11 |
| | S5044 | 7 | 2 | 5 | 13 | 74723411 | 75013512 | 290.102 | 404 | 4.60E-08 | 2.67E-02 | 6.66E-04 | 3.09E-11 |
| | S5052 | 7 | 4 | 3 | 15 | 69823443 | 69935918 | 112.476 | 250 | 3.00E-39 | 5.33E-02 | 4.00E-04 | 2.01E-42 |
| | S5055 | 7 | 2 | 5 | 16 | 12774758 | 12919187 | 144.43 | 318 | 4.60E-08 | 2.67E-02 | 6.66E-04 | 3.09E-11 |
| | S5056 | 7 | 3 | 4 | 16 | 13079905 | 13245014 | 165.11 | 342 | 1.59E-20 | 4.00E-02 | 5.33E-04 | 1.07E-23 |
| | S5061 | 7 | 3 | 4 | 18 | 4375900 | 4683756 | 307.857 | 638 | 1.59E-20 | 4.00E-02 | 5.33E-04 | 1.07E-23 |
| | S5062 | 7 | 3 | 4 | 18 | 12484728 | 13458064 | 973.337 | 1836 | 1.59E-20 | 4.00E-02 | 5.33E-04 | 1.07E-23 |
| | S5064 | 7 | 2 | 5 | 18 | 59698392 | 59802901 | 104.51 | 205 | 4.60E-08 | 2.67E-02 | 6.66E-04 | 3.09E-11 |
| | S5066 | 7 | 2 | 5 | 18 | 61178794 | 61339044 | 160.251 | 251 | 4.60E-08 | 2.67E-02 | 6.66E-04 | 3.09E-11 |
| | S5072 | 7 | 3 | 4 | 20 | 47972893 | 48518558 | 545.666 | 959 | 1.59E-20 | 4.00E-02 | 5.33E-04 | 1.07E-23 |
| | S5073 | 7 | 3 | 4 | 21 | 34342724 | 34746398 | 403.675 | 563 | 1.59E-20 | 4.00E-02 | 5.33E-04 | 1.07E-23 |
| | S5081 | 7 | 3 | 4 | 22 | 39179687 | 39289038 | 109.352 | 221 | 1.59E-20 | 4.00E-02 | 5.33E-04 | 1.07E-23 |
| | S5085 | 6 | 2 | 4 | 1 | 11805747 | 12134732 | 328.986 | 715 | 2.77E-09 | 2.67E-02 | 5.33E-04 | 1.86E-12 |
| | S5086 | 6 | 2 | 4 | 1 | 12240622 | 13373715 | 1133.094 | 1014 | 2.77E-09 | 2.67E-02 | 5.33E-04 | 1.86E-12 |

| | | | | | | | | | | | | | |
|---|---|---|---|---|---|---|---|---|---|---|---|---|---|
| | S5087 | 6 | 3 | 3 | 1 | 20565333 | 20744926 | 179.594 | 501 | 7.44E-24 | 4.00E-02 | 4.00E-04 | 4.99E-27 |
| | S5089 | 6 | 2 | 4 | 1 | 203167962 | 203362106 | 194.145 | 438 | 2.77E-09 | 2.67E-02 | 5.33E-04 | 1.86E-12 |
| | S5090 | 6 | 3 | 3 | 1 | 208038505 | 208455658 | 417.154 | 892 | 7.44E-24 | 4.00E-02 | 4.00E-04 | 4.99E-27 |
| | S5095 | 6 | 2 | 4 | 1 | 239925699 | 240061959 | 136.261 | 440 | 2.77E-09 | 2.67E-02 | 5.33E-04 | 1.86E-12 |
| | S5104 | 6 | 2 | 4 | 3 | 29133845 | 29670933 | 537.089 | 1291 | 2.77E-09 | 2.67E-02 | 5.33E-04 | 1.86E-12 |
| | S5108 | 6 | 2 | 4 | 3 | 186011964 | 186228961 | 216.998 | 259 | 2.77E-09 | 2.67E-02 | 5.33E-04 | 1.86E-12 |
| | S5109 | 6 | 2 | 4 | 3 | 192442281 | 192621852 | 179.572 | 260 | 2.77E-09 | 2.67E-02 | 5.33E-04 | 1.86E-12 |
| | S5110 | 6 | 2 | 4 | 3 | 194876514 | 195509651 | 633.138 | 1198 | 2.77E-09 | 2.67E-02 | 5.33E-04 | 1.86E-12 |
| | S5114 | 6 | 2 | 4 | 4 | 24487818 | 24733937 | 246.12 | 403 | 2.77E-09 | 2.67E-02 | 5.33E-04 | 1.86E-12 |
| | S5122 | 6 | 2 | 4 | 5 | 147296856 | 147460390 | 163.535 | 189 | 2.77E-09 | 2.67E-02 | 5.33E-04 | 1.86E-12 |
| | S5126 | 6 | 3 | 3 | 6 | 37207819 | 37357639 | 149.821 | 209 | 7.44E-24 | 4.00E-02 | 4.00E-04 | 4.99E-27 |
| | S5128 | 6 | 2 | 4 | 6 | 41527553 | 42320118 | 792.566 | 1519 | 2.77E-09 | 2.67E-02 | 5.33E-04 | 1.86E-12 |
| | S5131 | 6 | 2 | 4 | 6 | 150147387 | 150278470 | 131.084 | 322 | 2.77E-09 | 2.67E-02 | 5.33E-04 | 1.86E-12 |
| | S5133 | 6 | 2 | 4 | 6 | 155037515 | 155419151 | 381.637 | 754 | 2.77E-09 | 2.67E-02 | 5.33E-04 | 1.86E-12 |

| | | | | | | | | | | | | | |
|---|---|---|---|---|---|---|---|---|---|---|---|---|---|
| | S5135 | 6 | 3 | 3 | 7 | 17116005 | 17969582 | 853.578 | 1743 | 7.44E-24 | 4.00E-02 | 4.00E-04 | 4.99E-27 |
| | S5141 | 6 | 2 | 4 | 8 | 2353774 | 2510250 | 156.477 | 192 | 2.77E-09 | 2.67E-02 | 5.33E-04 | 1.86E-12 |
| | S5142 | 6 | 2 | 4 | 8 | 5548502 | 6129625 | 581.124 | 2472 | 2.77E-09 | 2.67E-02 | 5.33E-04 | 1.86E-12 |
| | S5143 | 6 | 2 | 4 | 8 | 20747941 | 21034004 | 286.064 | 658 | 2.77E-09 | 2.67E-02 | 5.33E-04 | 1.86E-12 |
| | S5151 | 6 | 3 | 3 | 9 | 76695330 | 76829672 | 134.343 | 261 | 7.44E-24 | 4.00E-02 | 4.00E-04 | 4.99E-27 |
| | S5152 | 6 | 2 | 4 | 9 | 89411806 | 89629605 | 217.8 | 538 | 2.77E-09 | 2.67E-02 | 5.33E-04 | 1.86E-12 |
| | S5153 | 6 | 2 | 4 | 10 | 4567117 | 4918672 | 351.556 | 815 | 2.77E-09 | 2.67E-02 | 5.33E-04 | 1.86E-12 |
| | S5156 | 6 | 2 | 4 | 10 | 6193032 | 7133489 | 940.458 | 2209 | 2.77E-09 | 2.67E-02 | 5.33E-04 | 1.86E-12 |
| | S5158 | 6 | 3 | 3 | 10 | 10270906 | 10487471 | 216.566 | 283 | 7.44E-24 | 4.00E-02 | 4.00E-04 | 4.99E-27 |
| | S5159 | 6 | 3 | 3 | 10 | 10635037 | 11241862 | 606.826 | 1129 | 7.44E-24 | 4.00E-02 | 4.00E-04 | 4.99E-27 |
| | S5160 | 6 | 2 | 4 | 10 | 48972420 | 49404701 | 432.282 | 894 | 2.77E-09 | 2.67E-02 | 5.33E-04 | 1.86E-12 |
| | S5165 | 6 | 2 | 4 | 10 | 120198681 | 120609072 | 410.392 | 701 | 2.77E-09 | 2.67E-02 | 5.33E-04 | 1.86E-12 |
| | S5178 | 6 | 2 | 4 | 12 | 3742080 | 4349866 | 607.787 | 1159 | 2.77E-09 | 2.67E-02 | 5.33E-04 | 1.86E-12 |
| | S5179 | 6 | 2 | 4 | 12 | 5296681 | 5611049 | 314.369 | 567 | 2.77E-09 | 2.67E-02 | 5.33E-04 | 1.86E-12 |

| | | | | | | | | | | | | | |
|---|---|---|---|---|---|---|---|---|---|---|---|---|---|
| | S5180 | 6 | 2 | 4 | 12 | 52250686 | 52549360 | 298.675 | 731 | 2.77E-09 | 2.67E-02 | 5.33E-04 | 1.86E-12 |
| | S5187 | 6 | 2 | 4 | 13 | 108842523 | 109533802 | 691.28 | 1574 | 2.77E-09 | 2.67E-02 | 5.33E-04 | 1.86E-12 |
| | S5199 | 6 | 3 | 3 | 15 | 59266157 | 59502327 | 236.171 | 445 | 7.44E-24 | 4.00E-02 | 4.00E-04 | 4.99E-27 |
| | S5200 | 6 | 4 | 2 | 15 | 70116822 | 70363632 | 246.811 | 470 | 9.56E-46 | 5.33E-02 | 2.66E-04 | 6.41E-49 |
| | S5201 | 6 | 2 | 4 | 15 | 89916435 | 90692159 | 775.725 | 1371 | 2.77E-09 | 2.67E-02 | 5.33E-04 | 1.86E-12 |
| | S5211 | 6 | 3 | 3 | 18 | 1787728 | 1916395 | 128.668 | 181 | 7.44E-24 | 4.00E-02 | 4.00E-04 | 4.99E-27 |
| | S5212 | 6 | 3 | 3 | 18 | 2902404 | 3365773 | 463.37 | 970 | 7.44E-24 | 4.00E-02 | 4.00E-04 | 4.99E-27 |
| | S5213 | 6 | 2 | 4 | 18 | 5334544 | 5559267 | 224.724 | 345 | 2.77E-09 | 2.67E-02 | 5.33E-04 | 1.86E-12 |
| | S5216 | 6 | 3 | 3 | 18 | 73454806 | 73835678 | 380.873 | 886 | 7.44E-24 | 4.00E-02 | 4.00E-04 | 4.99E-27 |
| | S5219 | 6 | 2 | 4 | 20 | 16073849 | 16744125 | 670.277 | 1575 | 2.77E-09 | 2.67E-02 | 5.33E-04 | 1.86E-12 |
| | S5221 | 6 | 3 | 3 | 21 | 31365198 | 31772197 | 407 | 822 | 7.44E-24 | 4.00E-02 | 4.00E-04 | 4.99E-27 |
| | S5227 | 5 | 2 | 3 | 1 | 60670085 | 61092333 | 422.249 | 953 | 5.52E-11 | 2.67E-02 | 4.00E-04 | 3.70E-14 |
| | S5235 | 5 | 2 | 3 | 2 | 173630158 | 173740905 | 110.748 | 167 | 5.52E-11 | 2.67E-02 | 4.00E-04 | 3.70E-14 |
| | S5243 | 5 | 3 | 2 | 3 | 173711383 | 174527931 | 816.549 | 1291 | 1.64E-28 | 4.00E-02 | 2.66E-04 | 1.10E-31 |

| | | | | | | | | | | | | | |
|---|---|---|---|---|---|---|---|---|---|---|---|---|---|
| | S5244 | 5 | 4 | 1 | 3 | 182675426 | 182843221 | 167.796 | 275 | 7.78E-55 | 5.33E-02 | 1.33E-04 | 5.22E-58 |
| | S5248 | 5 | 2 | 3 | 4 | 25861086 | 25967309 | 106.224 | 202 | 5.52E-11 | 2.67E-02 | 4.00E-04 | 3.70E-14 |
| | S5261 | 5 | 2 | 3 | 5 | 170740401 | 171616668 | 876.268 | 1284 | 5.52E-11 | 2.67E-02 | 4.00E-04 | 3.70E-14 |
| | S5262 | 5 | 2 | 3 | 6 | 12258537 | 12400414 | 141.878 | 276 | 5.52E-11 | 2.67E-02 | 4.00E-04 | 3.70E-14 |
| | S5263 | 5 | 2 | 3 | 6 | 12530603 | 13356097 | 825.495 | 1230 | 5.52E-11 | 2.67E-02 | 4.00E-04 | 3.70E-14 |
| | S5264 | 5 | 2 | 3 | 6 | 13467716 | 13961763 | 494.048 | 761 | 5.52E-11 | 2.67E-02 | 4.00E-04 | 3.70E-14 |
| | S5265 | 5 | 2 | 3 | 6 | 13965926 | 14133342 | 167.417 | 294 | 5.52E-11 | 2.67E-02 | 4.00E-04 | 3.70E-14 |
| | S5269 | 5 | 3 | 2 | 6 | 105198282 | 105396602 | 198.321 | 372 | 1.64E-28 | 4.00E-02 | 2.66E-04 | 1.10E-31 |
| | S5270 | 5 | 2 | 3 | 6 | 105617855 | 105869500 | 251.646 | 451 | 5.52E-11 | 2.67E-02 | 4.00E-04 | 3.70E-14 |
| | S5271 | 5 | 2 | 3 | 6 | 105880938 | 106358088 | 477.151 | 817 | 5.52E-11 | 2.67E-02 | 4.00E-04 | 3.70E-14 |
| | S5287 | 5 | 2 | 3 | 8 | 21757132 | 22574864 | 817.733 | 1395 | 5.52E-11 | 2.67E-02 | 4.00E-04 | 3.70E-14 |
| | S5297 | 5 | 2 | 3 | 8 | 141012783 | 141419373 | 406.591 | 761 | 5.52E-11 | 2.67E-02 | 4.00E-04 | 3.70E-14 |
| | S5298 | 5 | 2 | 3 | 8 | 141528693 | 142423309 | 894.617 | 1876 | 5.52E-11 | 2.67E-02 | 4.00E-04 | 3.70E-14 |
| | S5300 | 5 | 2 | 3 | 9 | 3086025 | 3570204 | 484.18 | 833 | 5.52E-11 | 2.67E-02 | 4.00E-04 | 3.70E-14 |

| | | | | | | | | | | | | | |
|---|---|---|---|---|---|---|---|---|---|---|---|---|---|
| | S5307 | 5 | 2 | 3 | 10 | 52393841 | 52921401 | 527.561 | 854 | 5.52E-11 | 2.67E-02 | 4.00E-04 | 3.70E-14 |
| | S5308 | 5 | 2 | 3 | 10 | 131566072 | 133466191 | 1900.12 | 3836 | 5.52E-11 | 2.67E-02 | 4.00E-04 | 3.70E-14 |
| | S5310 | 5 | 2 | 3 | 11 | 116608867 | 117059966 | 451.1 | 1025 | 5.52E-11 | 2.67E-02 | 4.00E-04 | 3.70E-14 |
| | S5314 | 5 | 3 | 2 | 12 | 12104304 | 12873451 | 769.148 | 1465 | 1.64E-28 | 4.00E-02 | 2.66E-04 | 1.10E-31 |
| | S5316 | 5 | 3 | 2 | 12 | 117599701 | 117727676 | 127.976 | 313 | 1.64E-28 | 4.00E-02 | 2.66E-04 | 1.10E-31 |
| | S5329 | 5 | 2 | 3 | 15 | 29253412 | 29454723 | 201.312 | 387 | 5.52E-11 | 2.67E-02 | 4.00E-04 | 3.70E-14 |
| | S5330 | 5 | 3 | 2 | 15 | 88871064 | 89515313 | 644.25 | 1064 | 1.64E-28 | 4.00E-02 | 2.66E-04 | 1.10E-31 |
| | S5331 | 5 | 2 | 3 | 15 | 92223230 | 92363732 | 140.503 | 280 | 5.52E-11 | 2.67E-02 | 4.00E-04 | 3.70E-14 |
| | S5332 | 5 | 3 | 2 | 15 | 92454402 | 94026616 | 1572.215 | 3733 | 1.64E-28 | 4.00E-02 | 2.66E-04 | 1.10E-31 |
| | S5337 | 5 | 2 | 3 | 17 | 1754605 | 2901672 | 1147.068 | 2088 | 5.52E-11 | 2.67E-02 | 4.00E-04 | 3.70E-14 |
| | S5343 | 5 | 2 | 3 | 18 | 2362860 | 2601771 | 238.912 | 486 | 5.52E-11 | 2.67E-02 | 4.00E-04 | 3.70E-14 |
| | S5344 | 5 | 2 | 3 | 18 | 71387001 | 71716158 | 329.158 | 842 | 5.52E-11 | 2.67E-02 | 4.00E-04 | 3.70E-14 |
| | S5345 | 5 | 2 | 3 | 18 | 71872556 | 72350248 | 477.693 | 931 | 5.52E-11 | 2.67E-02 | 4.00E-04 | 3.70E-14 |
| | S5350 | 5 | 3 | 2 | 21 | 31192067 | 31296802 | 104.736 | 126 | 1.64E-28 | 4.00E-02 | 2.66E-04 | 1.10E-31 |

| | | | | | | | | | | | | | |
|---|---|---|---|---|---|---|---|---|---|---|---|---|---|
| | S5357 | 4 | 2 | 2 | 1 | 59765623 | 60409875 | 644.253 | 1126 | 1.59E-13 | 2.67E-02 | 2.66E-04 | 1.07E-16 |
| | S5365 | 4 | 4 | 0 | 3 | 188777145 | 190116061 | 1338.917 | 2412 | 1.85E-68 | 5.33E-02 | 0.00E+00 | 1.24E-71 |
| | S5366 | 4 | 2 | 2 | 3 | 194040829 | 194550048 | 509.22 | 1031 | 1.59E-13 | 2.67E-02 | 2.66E-04 | 1.07E-16 |
| | S5376 | 4 | 2 | 2 | 5 | 148836339 | 149160025 | 323.687 | 574 | 1.59E-13 | 2.67E-02 | 2.66E-04 | 1.07E-16 |
| | S5378 | 4 | 2 | 2 | 5 | 169417174 | 169904066 | 486.893 | 994 | 1.59E-13 | 2.67E-02 | 2.66E-04 | 1.07E-16 |
| | S5379 | 4 | 2 | 2 | 5 | 170202868 | 170388480 | 185.613 | 368 | 1.59E-13 | 2.67E-02 | 2.66E-04 | 1.07E-16 |
| | S5403 | 4 | 2 | 2 | 12 | 6806740 | 7895125 | 1088.386 | 1691 | 1.59E-13 | 2.67E-02 | 2.66E-04 | 1.07E-16 |
| | S5404 | 4 | 2 | 2 | 12 | 127659470 | 128324588 | 665.119 | 1324 | 1.59E-13 | 2.67E-02 | 2.66E-04 | 1.07E-16 |
| | S5405 | 4 | 2 | 2 | 12 | 129151166 | 129726850 | 575.685 | 1358 | 1.59E-13 | 2.67E-02 | 2.66E-04 | 1.07E-16 |
| | S5413 | 4 | 2 | 2 | 14 | 100888741 | 101169312 | 280.572 | 611 | 1.59E-13 | 2.67E-02 | 2.66E-04 | 1.07E-16 |
| | S5414 | 4 | 2 | 2 | 15 | 27219111 | 28712165 | 1493.055 | 1484 | 1.59E-13 | 2.67E-02 | 2.66E-04 | 1.07E-16 |
| | S5437 | 4 | 2 | 2 | 21 | 27367234 | 27903745 | 536.512 | 1027 | 1.59E-13 | 2.67E-02 | 2.66E-04 | 1.07E-16 |
| | S5438 | 4 | 2 | 2 | 21 | 41666784 | 41940272 | 273.489 | 800 | 1.59E-13 | 2.67E-02 | 2.66E-04 | 1.07E-16 |
| | S5439 | 3 | 2 | 1 | 1 | 3731690 | 3850023 | 118.334 | 373 | 9.63E-18 | 2.67E-02 | 1.33E-04 | 6.46E-21 |

| | | | | | | | | | | | | | |
|---|---|---|---|---|---|---|---|---|---|---|---|---|---|
| | S5445 | 3 | 2 | 1 | 3 | 73382679 | 73984585 | 601.907 | 1130 | 9.63E-18 | 2.67E-02 | 1.33E-04 | 6.46E-21 |
| | S5454 | 3 | 2 | 1 | 8 | 4213245 | 4457827 | 244.583 | 1167 | 9.63E-18 | 2.67E-02 | 1.33E-04 | 6.46E-21 |
| | S5456 | 3 | 2 | 1 | 9 | 135776385 | 136047043 | 270.659 | 503 | 9.63E-18 | 2.67E-02 | 1.33E-04 | 6.46E-21 |
| | S5478 | 3 | 2 | 1 | 18 | 7460197 | 8787859 | 1327.663 | 2290 | 9.63E-18 | 2.67E-02 | 1.33E-04 | 6.46E-21 |
| **SAS** | S432 | 7 | 3 | 4 | 8 | 66043202 | 66722849 | 679.648 | 184 | 9.21E-06 | 3.75E-01 | 2.02E-02 | 3.34E-08 |
| | S541 | 6 | 3 | 3 | 7 | 55649184 | 57334218 | 1685.035 | 315 | 1.16E-06 | 3.75E-01 | 1.52E-02 | 4.22E-09 |
| | S595 | 6 | 3 | 3 | 16 | 71247813 | 72152410 | 904.598 | 250 | 1.16E-06 | 3.75E-01 | 1.52E-02 | 4.22E-09 |
| | S620 | 5 | 3 | 2 | 1 | 4467712 | 4644895 | 177.184 | 118 | 6.54E-08 | 3.75E-01 | 1.01E-02 | 2.37E-10 |
| | S887 | 2 | 2 | 0 | 4 | 62816976 | 65403313 | 2586.338 | 791 | 1.68E-07 | 2.50E-01 | 0.00E+00 | 6.11E-10 |
| | S892 | 2 | 2 | 0 | 8 | 128064327 | 129470564 | 1406.238 | 522 | 1.68E-07 | 2.50E-01 | 0.00E+00 | 6.11E-10 |

Legend: AAC = African Admixed, AFR = African, AJ = Ashkenazi Jewish, AMR = American Admixed, CAS = Central Asian, CHR = chromosome, EAS = East Asian, EUR = European, KB = kilobase, NSNP = number of single nucleotide polymorphisms, ROH = runs of homozygosity, SAS = South Asian, SNP = single nucleotide polymorphism. The Bonferroni threshold was adjusted by dividing 0.05 by the number of enriched ROHs to account for multiple comparisons.

***Supplementary Table 21:*** *Runs of homozygosity pools enriched in individuals with consanguinity that pass Bonferroni correction intersecting with known Parkinson's disease, dystonia, and atypical parkinsonism gene regions and risk loci*

| Ancestry | ROH POOL | Total participants | Total cases | Total controls | CHR | Base position 1 | Base position 2 | ROH KB | NSNP | PD, parkinsonism, or dystonia gene | p-value | Proportion cases | Proportion controls | Bonferroni correction |
|---|---|---|---|---|---|---|---|---|---|---|---|---|---|---|
| AAC | S109 | 6 | 2 | 4 | 15 | 50538139 | 51386390 | 848.252 | 2004 | *USP8* | 3.50E-12 | 3.33E-01 | 4.98E-03 | 1.95E-13 |
| | S232 | 3 | 1 | 2 | 2 | 102642764 | 104499042 | 1856.279 | 3085 | *MAP4K4* | 1.27E-03 | 1.67E-01 | 2.49E-03 | 7.05E-05 |
| | S318 | 3 | 1 | 2 | 16 | 17746297 | 18219727 | 473.431 | 870 | *SYT17* | 1.27E-03 | 1.67E-01 | 2.49E-03 | 7.05E-05 |
| | S320 | 3 | 1 | 2 | 17 | 44919781 | 46788132 | 1868.352 | 2462 | *UBTF FAM171A2 MAP3K14 CRHR1 MAPT-AS1 KANSL1 MAPT NSF WNT3* | 1.27E-03 | 1.67E-01 | 2.49E-03 | 7.05E-05 |
| | S393 | 2 | 1 | 1 | 6 | 32836793 | 35038899 | 2202.107 | 4929 | *SLC44A4 HLA-DRB5 ¬†AHR* | 6.18E-05 | 1.67E-01 | 1.24E-03 | 3.43E-06 |
| | S404 | 2 | 1 | 1 | 7 | 22118464 | 24040285 | 1921.822 | 4492 | *GPNMB KLHL7 NUPL2* | 6.18E-05 | 1.67E-01 | 1.24E-03 | 3.43E-06 |
| | S420 | 2 | 1 | 1 | 8 | 107635268 | 109528850 | 1893.583 | 4533 | *SYBU* | 6.18E-05 | 1.67E-01 | 1.24E-03 | 3.43E-06 |
| | S457 | 2 | 1 | 1 | 12 | 131246290 | 133220283 | 1973.994 | 4851 | *FBRSL1* | 6.18E-05 | 1.67E-01 | 1.24E-03 | 3.43E-06 |

| | | | | | | | | | | | | | | |
|---|---|---|---|---|---|---|---|---|---|---|---|---|---|---|
| | S465 | 2 | 1 | 1 | 14 | 53431888 | 56631839 | 3199.952 | 7120 | *GCH1* | 6.18E-05 | 1.67E-01 | 1.24E-03 | 3.43E-06 |
| **AFR** | S139 | 22 | 4 | 18 | 6 | 31415915 | 31521867 | 105.953 | 129 | *SLC44A4* | 7.62E-16 | 3.08E-01 | 1.10E-02 | 1.12E-17 |
| | S1731 | 2 | 1 | 1 | 1 | 154314139 | 154678568 | 364.43 | 762 | *PMVK GBAP1 ASH1L KRTCAP2* | 1.10E-04 | 7.69E-02 | 6.13E-04 | 1.62E-06 |
| | S1748 | 2 | 1 | 1 | 3 | 29269509 | 29926787 | 657.279 | 1803 | *LINC00693* | 1.10E-04 | 7.69E-02 | 6.13E-04 | 1.62E-06 |
| | S1779 | 2 | 1 | 1 | 6 | 162191075 | 163885184 | 1694.11 | 4348 | *PARK2* | 1.10E-04 | 7.69E-02 | 6.13E-04 | 1.62E-06 |
| | S213 | 17 | 3 | 14 | 3 | 52502292 | 52986528 | 484.237 | 999 | *ITIH3 ITIH4* | 7.45E-11 | 2.31E-01 | 8.58E-03 | 1.10E-12 |
| | S41 | 50 | 4 | 46 | 6 | 28180332 | 28285755 | 105.424 | 192 | *LOC100131289 RSL24D1P1* | 4.80E-07 | 3.08E-01 | 2.82E-02 | 7.06E-09 |
| | S433 | 11 | 2 | 9 | 6 | 33310149 | 33514091 | 203.943 | 296 | *HLA-DRB5* | 1.39E-06 | 1.54E-01 | 5.52E-03 | 2.05E-08 |
| | S441 | 11 | 2 | 9 | 6 | 70720341 | 70955770 | 235.43 | 328 | *RIMS1* | 1.39E-06 | 1.54E-01 | 5.52E-03 | 2.05E-08 |
| | S442 | 11 | 2 | 9 | 6 | 71029738 | 71360658 | 330.921 | 824 | *RIMS1* | 1.39E-06 | 1.54E-01 | 5.52E-03 | 2.05E-08 |
| | S443 | 11 | 2 | 9 | 6 | 71437857 | 71584215 | 146.359 | 333 | *RIMS1* | 1.39E-06 | 1.54E-01 | 5.52E-03 | 2.05E-08 |
| | S444 | 11 | 2 | 9 | 6 | 71739873 | 72255955 | 516.083 | 927 | *RIMS1* | 1.39E-06 | 1.54E-01 | 5.52E-03 | 2.05E-08 |
| | S448 | 11 | 2 | 9 | 6 | 145205964 | 146492402 | 1286.439 | 2362 | *RAB32* | 1.39E-06 | 1.54E-01 | 5.52E-03 | 2.05E-08 |
| | S458 | 11 | 2 | 9 | 8 | 108860914 | 109306803 | 445.89 | 1263 | *SYBU* | 1.39E-06 | 1.54E-01 | 5.52E-03 | 2.05E-08 |

| | | | | | | | | | | | | | | |
|---|---|---|---|---|---|---|---|---|---|---|---|---|---|---|
| | S461 | 11 | 2 | 9 | 9 | 34421281 | 34540917 | 119.637 | 152 | *UBAP2* | 1.39E-06 | 1.54E-01 | 5.52E-03 | 2.05E-08 |
| | S475 | 11 | 2 | 9 | 15 | 50284963 | 50761937 | 476.975 | 1168 | *USP8* | 1.39E-06 | 1.54E-01 | 5.52E-03 | 2.05E-08 |
| | S476 | 11 | 2 | 9 | 15 | 50763691 | 52095388 | 1331.698 | 2725 | *USP8* | 1.39E-06 | 1.54E-01 | 5.52E-03 | 2.05E-08 |
| | S634 | 9 | 2 | 7 | 9 | 34768208 | 34889377 | 121.17 | 275 | *UBAP2* | 6.97E-08 | 1.54E-01 | 4.29E-03 | 1.02E-09 |
| | S64 | 36 | 4 | 32 | 6 | 29403395 | 29516457 | 113.063 | 270 | *TRIM40* | 9.50E-10 | 3.08E-01 | 1.96E-02 | 1.40E-11 |
| | S67 | 35 | 5 | 30 | 6 | 29636570 | 29764813 | 128.244 | 510 | *TRIM40* | 3.74E-16 | 3.85E-01 | 1.84E-02 | 5.50E-18 |
| **AJ** | S1039 | 6 | 4 | 2 | 13 | 95543241 | 96945540 | 1402.3 | 1766 | *MBNL2* | 4.93E-05 | 8.16E-02 | 4.36E-04 | 4.41E-07 |
| | S1106 | 5 | 4 | 1 | 1 | 204388918 | 205845567 | 1456.65 | 2230 | *RAB29 SLC41A1* | 4.46E-06 | 8.16E-02 | 2.18E-04 | 3.98E-08 |
| | S151 | 12 | 7 | 5 | 12 | 40624560 | 40770009 | 145.45 | 277 | *LRRK2 CNTN1 PDZRN4* | 1.29E-07 | 1.43E-01 | 1.09E-03 | 1.15E-09 |
| | S152 | 12 | 8 | 4 | 12 | 40788769 | 41954393 | 1165.625 | 2219 | *LRRK2 CNTN1 PDZRN4 GXYLT1* | 3.64E-10 | 1.63E-01 | 8.73E-04 | 3.25E-12 |
| | S153 | 12 | 8 | 4 | 12 | 42529792 | 42653120 | 123.329 | 241 | *GXYLT1* | 3.64E-10 | 1.63E-01 | 8.73E-04 | 3.25E-12 |
| | S1536 | 4 | 3 | 1 | 20 | 909079 | 3069655 | 2160.577 | 3718 | *DDRGK1 PSMF1* | 3.30E-04 | 6.12E-02 | 2.18E-04 | 2.95E-06 |
| | S194 | 11 | 5 | 6 | 12 | 46228486 | 46901666 | 673.181 | 802 | *SCAF11 SLC38A1* | 3.91E-04 | 1.02E-01 | 1.31E-03 | 3.50E-06 |

| | | | | | | | | | | | | | | |
|---|---|---|---|---|---|---|---|---|---|---|---|---|---|---|
| | S24 | 27 | 9 | 18 | 12 | 33129292 | 34190817 | 1061.526 | 1700 | *FGD4 PKP2 ALG10* | 8.04E-05 | 1.84E-01 | 3.93E-03 | 7.18E-07 |
| | S271 | 10 | 7 | 3 | 12 | 44649310 | 45574415 | 925.106 | 963 | *SCAF11 SLC38A1* | 2.21E-09 | 1.43E-01 | 6.54E-04 | 1.97E-11 |
| | S272 | 10 | 6 | 4 | 12 | 45610378 | 45913265 | 302.888 | 231 | *SCAF11 SLC38A1* | 9.56E-07 | 1.22E-01 | 8.73E-04 | 8.54E-09 |
| | S340 | 9 | 5 | 4 | 5 | 60276522 | 61263213 | 986.692 | 1263 | *ELOVL7* | 3.59E-05 | 1.02E-01 | 8.73E-04 | 3.21E-07 |
| | S410 | 9 | 5 | 4 | 16 | 58499171 | 58934725 | 435.555 | 942 | *CNOT1* | 3.59E-05 | 1.02E-01 | 8.73E-04 | 3.21E-07 |
| | S411 | 9 | 5 | 4 | 16 | 59418341 | 59854704 | 436.364 | 748 | *CNOT1* | 3.59E-05 | 1.02E-01 | 8.73E-04 | 3.21E-07 |
| | S800 | 7 | 4 | 3 | 13 | 97321467 | 97927891 | 606.425 | 869 | *MBNL2* | 2.76E-04 | 8.16E-02 | 6.54E-04 | 2.46E-06 |
| | S832 | 7 | 5 | 2 | 21 | 43194143 | 44092986 | 898.844 | 1041 | *PDXK* | 8.44E-07 | 1.02E-01 | 4.36E-04 | 7.53E-09 |
| **AMR** | S200 | 3 | 3 | 0 | 7 | 70645942 | 73741147 | 3095.206 | 4119 | *GALNT17* | 1.91E-04 | 1.50E-01 | 0.00E+00 | 5.02E-06 |
| | S211 | 3 | 3 | 0 | 10 | 15180551 | 15923173 | 742.623 | 1062 | *ITGA8 FAM171A1* | 1.91E-04 | 1.50E-01 | 0.00E+00 | 5.02E-06 |
| **CAS** | S201 | 6 | 4 | 2 | 12 | 40662517 | 42677591 | 2015.075 | 3661 | *LRRK2 CNTN1 PDZRN4 GXYLT1* | 4.94E-09 | 2.67E-01 | 7.49E-03 | 7.27E-11 |
| | S341 | 5 | 3 | 2 | 12 | 44863086 | 47069014 | 2205.929 | 2919 | *SCAF11 SLC38A1* | 7.06E-06 | 2.00E-01 | 7.49E-03 | 1.04E-07 |
| | S353 | 5 | 3 | 2 | 17 | 14867684 | 17192330 | 2324.647 | 2715 | *PIGL* | 7.06E-06 | 2.00E-01 | 7.49E-03 | 1.04E-07 |

| | | | | | | | | | | | | | | |
|---|---|---|---|---|---|---|---|---|---|---|---|---|---|---|
| | S391 | 4 | 3 | 1 | 2 | 100722823 | 101159757 | 436.935 | 722 | *MAP4K4* | 2.86E-07 | 2.00E-01 | 3.75E-03 | 4.20E-09 |
| | S398 | 4 | 3 | 1 | 2 | 165453151 | 167634399 | 2181.249 | 3154 | *STK39* | 2.86E-07 | 2.00E-01 | 3.75E-03 | 4.20E-09 |
| | S399 | 4 | 3 | 1 | 2 | 167644704 | 167844661 | 199.958 | 360 | *STK39* | 2.86E-07 | 2.00E-01 | 3.75E-03 | 4.20E-09 |
| | S457 | 4 | 4 | 0 | 6 | 110411111 | 111140162 | 729.052 | 1070 | *FYN* | 1.62E-13 | 2.67E-01 | 0.00E+00 | 2.39E-15 |
| | S512 | 4 | 3 | 1 | 11 | 84667999 | 86999400 | 2331.402 | 3319 | *DLG2* | 2.86E-07 | 2.00E-01 | 3.75E-03 | 4.20E-09 |
| | S608 | 3 | 2 | 1 | 3 | 151124172 | 153121159 | 1996.988 | 2899 | *MED12L* | 5.26E-04 | 1.33E-01 | 3.75E-03 | 7.74E-06 |
| | S618 | 3 | 2 | 1 | 4 | 14928795 | 15879079 | 950.285 | 1654 | *BST1 BST1 FAM200B CD38* | 5.26E-04 | 1.33E-01 | 3.75E-03 | 7.74E-06 |
| | S633 | 3 | 2 | 1 | 4 | 168662491 | 168844939 | 182.449 | 307 | *CLCN3* | 5.26E-04 | 1.33E-01 | 3.75E-03 | 7.74E-06 |
| | S827 | 2 | 2 | 0 | 2 | 16368968 | 17996554 | 1627.587 | 2063 | *KCNS3* | 1.05E-05 | 1.33E-01 | 0.00E+00 | 1.54E-07 |
| | S828 | 2 | 2 | 0 | 2 | 18171735 | 20375251 | 2203.517 | 3441 | *KCNS3* | 1.05E-05 | 1.33E-01 | 0.00E+00 | 1.54E-07 |
| **EAS** | S1079 | 9 | 2 | 7 | 11 | 84504716 | 84644243 | 139.528 | 110 | *DLG2* | 1.04E-13 | 2.00E-01 | 3.04E-03 | 2.73E-15 |
| | S1520 | 5 | 1 | 4 | 4 | 76422974 | 76561775 | 138.802 | 180 | *SCARB2 FAM47E FAM47E-STBD1* | 1.10E-03 | 1.00E-01 | 1.74E-03 | 2.90E-05 |
| | S1594 | 5 | 1 | 4 | 12 | 46976766 | 47096091 | 119.326 | 236 | *SCAF11* | 1.10E-03 | 1.00E-01 | 1.74E-03 | 2.90E-05 |

| | | | | | | | | | | | | | | |
|---|---|---|---|---|---|---|---|---|---|---|---|---|---|---|
| | S1688 | 4 | 1 | 3 | 3 | 178535521 | 178850775 | 315.255 | 446 | *AC076966.1* | 2.32E-04 | 1.00E-01 | 1.30E-03 | 6.12E-06 |
| | S1689 | 4 | 1 | 3 | 3 | 179312326 | 179604007 | 291.682 | 327 | *AC076966.1* | 2.32E-04 | 1.00E-01 | 1.30E-03 | 6.12E-06 |
| | S1784 | 4 | 1 | 3 | 13 | 96654734 | 97783122 | 1128.389 | 937 | *MBNL2* | 2.32E-04 | 1.00E-01 | 1.30E-03 | 6.12E-06 |
| | S1818 | 3 | 1 | 2 | 1 | 171080339 | 171247758 | 167.42 | 242 | *VAMP4* | 1.81E-05 | 1.00E-01 | 8.69E-04 | 4.75E-07 |
| | S1824 | 3 | 1 | 2 | 2 | 18373851 | 20413320 | 2039.47 | 2497 | *KCNS3* | 1.81E-05 | 1.00E-01 | 8.69E-04 | 4.75E-07 |
| | S1890 | 3 | 1 | 2 | 6 | 162333574 | 163245816 | 912.243 | 1217 | *PARK2* | 1.81E-05 | 1.00E-01 | 8.69E-04 | 4.75E-07 |
| | S2068 | 2 | 1 | 1 | 21 | 36541167 | 36645075 | 103.909 | 121 | *DYRK1A* | 1.18E-07 | 1.00E-01 | 4.34E-04 | 3.10E-09 |
| | S293 | 34 | 2 | 32 | 22 | 41017765 | 41949177 | 931.413 | 990 | *RBX1* | 3.68E-04 | 2.00E-01 | 1.39E-02 | 9.69E-06 |
| | S427 | 22 | 2 | 20 | 6 | 145191755 | 146344288 | 1152.534 | 1464 | *RAB32* | 4.52E-06 | 2.00E-01 | 8.69E-03 | 1.19E-07 |
| | S622 | 15 | 2 | 13 | 1 | 92380023 | 93057526 | 677.504 | 462 | *MTF2* | 1.47E-08 | 2.00E-01 | 5.65E-03 | 3.87E-10 |
| | S654 | 15 | 2 | 13 | 11 | 82784557 | 82985843 | 201.287 | 221 | *DLG2* | 1.47E-08 | 2.00E-01 | 5.65E-03 | 3.87E-10 |
| **EUR** | S1036 | 55 | 6 | 49 | 5 | 101986791 | 102198676 | 211.886 | 499 | *PAM* | 1.23E-11 | 8.00E-02 | 6.53E-03 | 9.50E-14 |
| | S1057 | 54 | 6 | 48 | 5 | 102250985 | 102534752 | 283.768 | 580 | *PAM* | 7.30E-12 | 8.00E-02 | 6.40E-03 | 5.61E-14 |
| | S1135 | 50 | 5 | 45 | 3 | 160436998 | 160601723 | 164.726 | 176 | *SPTSSB* | 9.37E-09 | 6.67E-02 | 6.00E-03 | 7.21E-11 |

| | | | | | | | | | | | | | | |
|---|---|---|---|---|---|---|---|---|---|---|---|---|---|---|
| | S1650 | 34 | 4 | 30 | 15 | 50918936 | 51207300 | 288.365 | 506 | *USP8* | 3.93E-08 | 5.33E-02 | 4.00E-03 | 3.02E-10 |
| | S1699 | 33 | 3 | 30 | 12 | 123032370 | 123283558 | 251.189 | 232 | *HIP1R* | 1.28E-04 | 4.00E-02 | 4.00E-03 | 9.82E-07 |
| | S1756 | 32 | 3 | 29 | 12 | 40435763 | 40587993 | 152.231 | 390 | *LRRK2 CNTN1 PDZRN4* | 9.31E-05 | 4.00E-02 | 3.86E-03 | 7.16E-07 |
| | S1758 | 32 | 3 | 29 | 12 | 122319709 | 122626238 | 306.53 | 401 | *HIP1R OGFOD2* | 9.31E-05 | 4.00E-02 | 3.86E-03 | 7.16E-07 |
| | S1831 | 31 | 4 | 27 | 12 | 40266815 | 40421822 | 155.008 | 387 | *LRRK2 CNTN1* | 6.38E-09 | 5.33E-02 | 3.60E-03 | 4.91E-11 |
| | S1914 | 30 | 7 | 23 | 16 | 29991755 | 30108956 | 117.202 | 121 | *SETD1A* | 1.98E-30 | 9.33E-02 | 3.06E-03 | 1.52E-32 |
| | S1984 | 29 | 4 | 25 | 12 | 41120483 | 41284161 | 163.679 | 295 | *LRRK2 CNTN1 PDZRN4 GXYLT1* | 1.54E-09 | 5.33E-02 | 3.33E-03 | 1.19E-11 |
| | S2068 | 27 | 3 | 24 | 1 | 152969457 | 153087421 | 117.965 | 209 | *HRNR* | 1.37E-05 | 4.00E-02 | 3.20E-03 | 1.05E-07 |
| | S2391 | 24 | 3 | 21 | 12 | 123582733 | 123773596 | 190.864 | 255 | *HIP1R* | 2.96E-06 | 4.00E-02 | 2.80E-03 | 2.28E-08 |
| | S2439 | 23 | 5 | 18 | 3 | 152349004 | 152562959 | 213.956 | 343 | *MED12L* | 1.98E-19 | 6.67E-02 | 2.40E-03 | 1.52E-21 |
| | S2468 | 23 | 4 | 19 | 6 | 71358447 | 71667230 | 308.784 | 582 | *RIMS1* | 5.04E-12 | 5.33E-02 | 2.53E-03 | 3.88E-14 |
| | S2503 | 23 | 4 | 19 | 12 | 41615371 | 41820036 | 204.666 | 523 | *PDZRN4 GXYLT1* | 5.04E-12 | 5.33E-02 | 2.53E-03 | 3.88E-14 |
| | S2534 | 22 | 4 | 18 | 1 | 155061442 | 155464151 | 402.71 | 432 | *GBAP1 ASH1L PMVK KRTCAP2* | 1.44E-12 | 5.33E-02 | 2.40E-03 | 1.10E-14 |

| | | | | | | | | | | | | | | |
|---|---|---|---|---|---|---|---|---|---|---|---|---|---|---|
| | | | | | | | | | | *GBA GBA1 SEMA4A* | | | | |
| | S2582 | 22 | 4 | 18 | 6 | 71744101 | 71857261 | 113.161 | 310 | *RIMS1* | 1.44E-12 | 5.33E-02 | 2.40E-03 | 1.10E-14 |
| | S2770 | 20 | 3 | 17 | 2 | 74206825 | 74492875 | 286.051 | 245 | *DCTN1* | 1.91E-07 | 4.00E-02 | 2.27E-03 | 1.47E-09 |
| | S2855 | 20 | 3 | 17 | 11 | 83322955 | 83767909 | 444.955 | 674 | *DLG2* | 1.91E-07 | 4.00E-02 | 2.27E-03 | 1.47E-09 |
| | S2941 | 19 | 6 | 13 | 5 | 103842705 | 103979003 | 136.299 | 270 | *PAM* | 6.41E-35 | 8.00E-02 | 1.73E-03 | 4.93E-37 |
| | S3282 | 16 | 3 | 13 | 1 | 153809535 | 154158721 | 349.187 | 478 | *PMVK* | 3.20E-09 | 4.00E-02 | 1.73E-03 | 2.46E-11 |
| | S3283 | 16 | 3 | 13 | 1 | 154576516 | 154683482 | 106.967 | 179 | *PMVK KRTCAP2 GBAP1 ASH1L PMVK* | 3.20E-09 | 4.00E-02 | 1.73E-03 | 2.46E-11 |
| | S3330 | 16 | 3 | 13 | 4 | 75905987 | 76402625 | 496.639 | 974 | *SCARB2 FAM47E FAM47E-STBD1* | 3.20E-09 | 4.00E-02 | 1.73E-03 | 2.46E-11 |
| | S3389 | 16 | 5 | 11 | 12 | 31628444 | 31745841 | 117.398 | 202 | *FGD4* | 4.88E-28 | 6.67E-02 | 1.47E-03 | 3.75E-30 |
| | S3470 | 15 | 5 | 10 | 3 | 178620339 | 178843865 | 223.527 | 499 | *AC076966.1* | 6.39E-30 | 6.67E-02 | 1.33E-03 | 4.91E-32 |
| | S3522 | 15 | 4 | 11 | 9 | 33187899 | 33318590 | 130.692 | 178 | *UBAP2* | 2.10E-18 | 5.33E-02 | 1.47E-03 | 1.62E-20 |
| | S3548 | 15 | 4 | 11 | 12 | 46545731 | 46736257 | 190.527 | 246 | *SCAF11* | 2.10E-18 | 5.33E-02 | 1.47E-03 | 1.62E-20 |
| | S3565 | 15 | 4 | 11 | 15 | 45143019 | 45253280 | 110.262 | 160 | *SPG11* | 2.10E-18 | 5.33E-02 | 1.47E-03 | 1.62E-20 |

| | | | | | | | | | | | | | | |
|---|---|---|---|---|---|---|---|---|---|---|---|---|---|---|
| | S3567 | 15 | 9 | 6 | 16 | 49626115 | 49842961 | 216.847 | 421 | *NOD2* | 1.95E-105 | 1.20E-01 | 7.99E-04 | 1.50E-107 |
| | S3568 | 15 | 9 | 6 | 16 | 49920928 | 50053087 | 132.16 | 221 | *NOD2* | 1.95E-105 | 1.20E-01 | 7.99E-04 | 1.50E-107 |
| | S3569 | 15 | 3 | 12 | 16 | 74896352 | 75123124 | 226.773 | 403 | *FA2H* | 8.22E-10 | 4.00E-02 | 1.60E-03 | 6.33E-12 |
| | S3627 | 14 | 2 | 12 | 4 | 2295975 | 2404772 | 108.798 | 288 | *ADD1* | 2.34E-04 | 2.67E-02 | 1.60E-03 | 1.80E-06 |
| | S3635 | 14 | 4 | 10 | 4 | 111629692 | 112326279 | 696.588 | 839 | *ANK2* | 1.04E-19 | 5.33E-02 | 1.33E-03 | 7.97E-22 |
| | S3663 | 14 | 5 | 9 | 6 | 162515945 | 163183260 | 667.316 | 1299 | *PARK2* | 4.51E-32 | 6.67E-02 | 1.20E-03 | 3.47E-34 |
| | S3700 | 14 | 3 | 11 | 12 | 46848016 | 47082605 | 234.59 | 425 | *SCAF11* | 1.74E-10 | 4.00E-02 | 1.47E-03 | 1.34E-12 |
| | S3726 | 14 | 2 | 12 | 17 | 62828828 | 63866085 | 1037.258 | 1077 | *BRIP1* | 2.34E-04 | 2.67E-02 | 1.60E-03 | 1.80E-06 |
| | S3730 | 14 | 2 | 12 | 18 | 45670414 | 45809520 | 139.107 | 300 | *SYT4* | 2.34E-04 | 2.67E-02 | 1.60E-03 | 1.80E-06 |
| | S3773 | 13 | 4 | 9 | 4 | 15021890 | 15336242 | 314.353 | 556 | *BST1* | 3.22E-21 | 5.33E-02 | 1.20E-03 | 2.48E-23 |
| | S3776 | 13 | 3 | 10 | 4 | 77249630 | 77372145 | 122.516 | 304 | *FAM47E-STBD1* | 2.92E-11 | 4.00E-02 | 1.33E-03 | 2.24E-13 |
| | S3808 | 13 | 4 | 9 | 7 | 23067749 | 23926540 | 858.792 | 1480 | *GPNMB KLHL7 NUPL2* | 3.22E-21 | 5.33E-02 | 1.20E-03 | 2.48E-23 |
| | S3826 | 13 | 4 | 9 | 9 | 32939659 | 33090322 | 150.664 | 293 | *UBAP2* | 3.22E-21 | 5.33E-02 | 1.20E-03 | 2.48E-23 |
| | S3880 | 13 | 2 | 11 | 14 | 75008572 | 75131153 | 122.582 | 109 | *RPS6KL1* | 1.20E-04 | 2.67E-02 | 1.47E-03 | 9.23E-07 |

| | | | | | | | | | | | | | | |
|---|---|---|---|---|---|---|---|---|---|---|---|---|---|---|
| | S3897 | 13 | 3 | 10 | 16 | 74092772 | 74280381 | 187.61 | 358 | *FA2H* | 2.92E-11 | 4.00E-02 | 1.33E-03 | 2.24E-13 |
| | S3905 | 13 | 3 | 10 | 18 | 49658476 | 50557654 | 899.179 | 1740 | *MEX3C* | 2.92E-11 | 4.00E-02 | 1.33E-03 | 2.24E-13 |
| | S3980 | 12 | 3 | 9 | 5 | 74945323 | 76297886 | 1352.564 | 2231 | *SV2C* | 3.64E-12 | 4.00E-02 | 1.20E-03 | 2.80E-14 |
| | S3990 | 12 | 3 | 9 | 6 | 132608199 | 133163967 | 555.769 | 978 | *RPS12 HMGB1P13* | 3.64E-12 | 4.00E-02 | 1.20E-03 | 2.80E-14 |
| | S3991 | 12 | 4 | 8 | 7 | 24108825 | 24212310 | 103.486 | 213 | *GPNMB* | 5.64E-23 | 5.33E-02 | 1.07E-03 | 4.34E-25 |
| | S4072 | 12 | 2 | 10 | 14 | 55104131 | 55449449 | 345.319 | 570 | *GCH1* | 5.54E-05 | 2.67E-02 | 1.33E-03 | 4.26E-07 |
| | S4084 | 12 | 6 | 6 | 16 | 50982680 | 51286761 | 304.082 | 587 | *NOD2* | 1.35E-55 | 8.00E-02 | 7.99E-04 | 1.04E-57 |
| | S4100 | 11 | 5 | 6 | 1 | 7698700 | 9019095 | 1320.396 | 2489 | *DJ-1* | 7.27E-41 | 6.67E-02 | 7.99E-04 | 5.59E-43 |
| | S4115 | 11 | 3 | 8 | 1 | 160672214 | 160905887 | 233.674 | 426 | *FCGR2A* | 3.11E-13 | 4.00E-02 | 1.07E-03 | 2.40E-15 |
| | S4116 | 11 | 3 | 8 | 1 | 160950381 | 161259053 | 308.673 | 581 | *FCGR2A* | 3.11E-13 | 4.00E-02 | 1.07E-03 | 2.40E-15 |
| | S4154 | 11 | 2 | 9 | 3 | 17148999 | 17960084 | 811.086 | 857 | *SATB1 TBC1D5* | 2.23E-05 | 2.67E-02 | 1.20E-03 | 1.71E-07 |
| | S4208 | 11 | 4 | 7 | 6 | 147016387 | 147497968 | 481.582 | 474 | *RAB32* | 4.75E-25 | 5.33E-02 | 9.33E-04 | 3.65E-27 |
| | S4211 | 11 | 3 | 8 | 6 | 161444200 | 161686497 | 242.298 | 542 | *PARK2* | 3.11E-13 | 4.00E-02 | 1.07E-03 | 2.40E-15 |
| | S4252 | 11 | 3 | 8 | 8 | 129064176 | 129340981 | 276.806 | 463 | *FAM49B* | 3.11E-13 | 4.00E-02 | 1.07E-03 | 2.40E-15 |

| | | | | | | | | | | | | | | |
|---|---|---|---|---|---|---|---|---|---|---|---|---|---|---|
| | S4324 | 11 | 5 | 6 | 16 | 51998601 | 52452407 | 453.807 | 820 | *CASC16 CHD9 TOX3 PHBP2* | 7.27E-41 | 6.67E-02 | 7.99E-04 | 5.59E-43 |
| | S4325 | 11 | 5 | 6 | 16 | 58288385 | 58461459 | 173.075 | 271 | *CNOT1* | 7.27E-41 | 6.67E-02 | 7.99E-04 | 5.59E-43 |
| | S4361 | 10 | 3 | 7 | 1 | 162470743 | 162600585 | 129.843 | 222 | *FCGR2A* | 1.64E-14 | 4.00E-02 | 9.33E-04 | 1.26E-16 |
| | S4395 | 10 | 3 | 7 | 3 | 123430486 | 123695355 | 264.87 | 309 | *KPNA1* | 1.64E-14 | 4.00E-02 | 9.33E-04 | 1.26E-16 |
| | S4399 | 10 | 3 | 7 | 3 | 150713302 | 150872381 | 159.08 | 177 | *MED12L* | 1.64E-14 | 4.00E-02 | 9.33E-04 | 1.26E-16 |
| | S4413 | 10 | 5 | 5 | 5 | 124279867 | 124566139 | 286.273 | 510 | *ZNF608* | 5.77E-45 | 6.67E-02 | 6.66E-04 | 4.44E-47 |
| | S4414 | 10 | 4 | 6 | 5 | 125630120 | 125804876 | 174.757 | 396 | *ZNF608* | 1.54E-27 | 5.33E-02 | 7.99E-04 | 1.19E-29 |
| | S4417 | 10 | 3 | 7 | 5 | 134230104 | 135130379 | 900.276 | 1001 | *C5orf24 AC006077.1* | 1.64E-14 | 4.00E-02 | 9.33E-04 | 1.26E-16 |
| | S4434 | 10 | 3 | 7 | 7 | 22240125 | 22384820 | 144.696 | 284 | *GPNMB* | 1.64E-14 | 4.00E-02 | 9.33E-04 | 1.26E-16 |
| | S444 | 129 | 6 | 123 | 14 | 66782901 | 67094191 | 311.291 | 340 | *TMEM229B* | 1.51E-04 | 8.00E-02 | 1.64E-02 | 1.16E-06 |
| | S4479 | 10 | 2 | 8 | 10 | 119296873 | 119821778 | 524.906 | 965 | *BAG3 INPP5F* | 7.49E-06 | 2.67E-02 | 1.07E-03 | 5.76E-08 |
| | S4520 | 10 | 2 | 8 | 14 | 37036574 | 37593915 | 557.342 | 589 | *MIPOL1* | 7.49E-06 | 2.67E-02 | 1.07E-03 | 5.76E-08 |
| | S4538 | 10 | 4 | 6 | 16 | 53134146 | 53342939 | 208.794 | 264 | *CASC16 CHD9 PHBP2* | 1.54E-27 | 5.33E-02 | 7.99E-04 | 1.19E-29 |
| | S4540 | 10 | 5 | 5 | 16 | 57412095 | 57855908 | 443.814 | 847 | *CNOT1* | 5.77E-45 | 6.67E-02 | 6.66E-04 | 4.44E-47 |

| | | | | | | | | | | | | | | |
|---|---|---|---|---|---|---|---|---|---|---|---|---|---|---|
| | S4549 | 10 | 2 | 8 | 18 | 51453244 | 51588653 | 135.41 | 199 | *MEX3C SMAD4* | 7.49E-06 | 2.67E-02 | 1.07E-03 | 5.76E-08 |
| | S4632 | 9 | 4 | 5 | 5 | 124578128 | 125507383 | 929.256 | 1900 | *ZNF608* | 1.41E-30 | 5.33E-02 | 6.66E-04 | 1.09E-32 |
| | S4731 | 9 | 3 | 6 | 16 | 19279092 | 19384618 | 105.527 | 226 | *SYT17* | 4.50E-16 | 4.00E-02 | 7.99E-04 | 3.46E-18 |
| | S4733 | 9 | 4 | 5 | 16 | 52757088 | 52945399 | 188.312 | 335 | *CASC16 CHD9 PHBP2* | 1.41E-30 | 5.33E-02 | 6.66E-04 | 1.09E-32 |
| | S4797 | 8 | 2 | 6 | 5 | 76506020 | 76662097 | 156.078 | 309 | *SV2C* | 3.81E-07 | 2.67E-02 | 7.99E-04 | 2.93E-09 |
| | S4890 | 8 | 4 | 4 | 15 | 61048372 | 61374329 | 325.958 | 531 | *VPS13C lincRNA* | 2.26E-34 | 5.33E-02 | 5.33E-04 | 1.74E-36 |
| | S4891 | 8 | 4 | 4 | 15 | 61540658 | 61835798 | 295.141 | 568 | *VPS13C lincRNA VPS13C* | 2.26E-34 | 5.33E-02 | 5.33E-04 | 1.74E-36 |
| | S4892 | 8 | 5 | 3 | 15 | 62261912 | 62400579 | 138.668 | 302 | *VPS13C* | 3.12E-56 | 6.67E-02 | 4.00E-04 | 2.40E-58 |
| | S4929 | 8 | 2 | 6 | 21 | 43836504 | 44092475 | 255.972 | 396 | *PDXK* | 3.81E-07 | 2.67E-02 | 7.99E-04 | 2.93E-09 |
| | S4964 | 7 | 3 | 4 | 3 | 183467293 | 183579399 | 112.107 | 193 | *MCCC1* | 1.59E-20 | 4.00E-02 | 5.33E-04 | 1.23E-22 |
| | S5009 | 7 | 2 | 5 | 8 | 23635862 | 23768832 | 132.971 | 298 | *BIN3* | 4.60E-08 | 2.67E-02 | 6.66E-04 | 3.54E-10 |
| | S5087 | 6 | 3 | 3 | 1 | 20565333 | 20744926 | 179.594 | 501 | *PINK1* | 7.44E-24 | 4.00E-02 | 4.00E-04 | 5.72E-26 |
| | S5104 | 6 | 2 | 4 | 3 | 29133845 | 29670933 | 537.089 | 1291 | *LINC00693 RBMS3* | 2.77E-09 | 2.67E-02 | 5.33E-04 | 2.13E-11 |
| | S5165 | 6 | 2 | 4 | 10 | 120198681 | 120609072 | 410.392 | 701 | *BAG3 INPP5F* | 2.77E-09 | 2.67E-02 | 5.33E-04 | 2.13E-11 |

| | | | | | | | | | | | | | | |
|---|---|---|---|---|---|---|---|---|---|---|---|---|---|---|
| | S5187 | 6 | 2 | 4 | 13 | 108842523 | 109533802 | 691.28 | 1574 | *RN7SKP10* | 2.77E-09 | 2.67E-02 | 5.33E-04 | 2.13E-11 |
| | S5244 | 5 | 4 | 1 | 3 | 182675426 | 182843221 | 167.796 | 275 | *MCCC1* | 7.78E-55 | 5.33E-02 | 1.33E-04 | 5.98E-57 |
| | S5287 | 5 | 2 | 3 | 8 | 21757132 | 22574864 | 817.733 | 1395 | *BIN3 BIN3* | 5.52E-11 | 2.67E-02 | 4.00E-04 | 4.25E-13 |
| | S5330 | 5 | 3 | 2 | 15 | 88871064 | 89515313 | 644.25 | 1064 | *POLG1* | 1.64E-28 | 4.00E-02 | 2.66E-04 | 1.26E-30 |
| | S796 | 75 | 7 | 68 | 10 | 103044198 | 103193308 | 149.111 | 260 | *GBF1* | 1.47E-11 | 9.33E-02 | 9.06E-03 | 1.13E-13 |
| | S881 | 64 | 6 | 58 | 1 | 92419450 | 93018831 | 599.382 | 859 | *MTF2* | 6.73E-10 | 8.00E-02 | 7.73E-03 | 5.17E-12 |
| | S933 | 61 | 8 | 53 | 5 | 60612700 | 61170442 | 557.743 | 801 | *ELOVL7* | 3.33E-19 | 1.07E-01 | 7.06E-03 | 2.56E-21 |
| | S953 | 59 | 5 | 54 | 3 | 161193918 | 161461944 | 268.027 | 332 | *SPTSSB* | 2.32E-07 | 6.67E-02 | 7.20E-03 | 1.79E-09 |
| | S970 | 58 | 5 | 53 | 3 | 160982708 | 161090052 | 107.345 | 163 | *SPTSSB* | 1.71E-07 | 6.67E-02 | 7.06E-03 | 1.31E-09 |
| **SAS** | S541 | 6 | 3 | 3 | 7 | 55649184 | 57334218 | 1685.035 | 315 | *CHCHD2* | 1.16E-06 | 3.75E-01 | 1.52E-02 | 3.88E-08 |
| | S892 | 2 | 2 | 0 | 8 | 128064327 | 129470564 | 1406.238 | 522 | *FAM49B* | 1.68E-07 | 2.50E-01 | 0.00E+00 | 5.62E-09 |

Legend: AAC = African Admixed, AFR = African, AJ = Ashkenazi Jewish, AMR = American Admixed, CAS = Central Asian, CHR = chromosome, EAS = East Asian, EUR = European, KB = kilobase, NSNP = number of single nucleotide polymorphisms, ROH = runs of homozygosity, SAS = South Asian, SNP = single nucleotide polymorphism. The Bonferroni threshold was adjusted by dividing 0.05 by the number of enriched ROHs to account for multiple comparisons.

***Supplementary Table 22:*** *Runs of homozygosity pools enriched in early-onset cases that pass Bonferroni correction*

| Ancestry | ROH POOL | Total participants | Total cases | Total controls | CHR | Base position 1 | Base position 2 | ROH KB | NSNP | p-value | Proportion cases | Proportion controls | Bonferroni correction |
|---|---|---|---|---|---|---|---|---|---|---|---|---|---|
| AFR | S471 | 10 | 2 | 8 | 6 | 56159013 | 56285898 | 126.886 | 336 | 1.01E-04 | 0.095 | 4.90E-03 | 3.55E-07 |
| | S483 | 10 | 2 | 8 | 8 | 50110122 | 50371754 | 261.633 | 619 | 1.01E-04 | 0.095 | 4.90E-03 | 3.55E-07 |
| | S563 | 9 | 2 | 7 | 8 | 50929511 | 51047399 | 117.889 | 321 | 3.56E-05 | 0.095 | 4.29E-03 | 1.25E-07 |
| | S590 | 9 | 2 | 7 | 13 | 83852625 | 84244091 | 391.467 | 829 | 3.56E-05 | 0.095 | 4.29E-03 | 1.25E-07 |
| | S591 | 9 | 2 | 7 | 13 | 84346393 | 84466830 | 120.438 | 322 | 3.56E-05 | 0.095 | 4.29E-03 | 1.25E-07 |
| | S740 | 7 | 2 | 5 | 2 | 21445013 | 22189250 | 744.238 | 1196 | 1.84E-06 | 0.095 | 3.07E-03 | 6.42E-09 |
| | S741 | 7 | 2 | 5 | 2 | 22323717 | 22483764 | 160.048 | 331 | 1.84E-06 | 0.095 | 3.07E-03 | 6.42E-09 |
| | S812 | 7 | 2 | 5 | 6 | 65338009 | 65627749 | 289.741 | 601 | 1.84E-06 | 0.095 | 3.07E-03 | 6.42E-09 |
| | S861 | 7 | 2 | 5 | 13 | 82670299 | 82855364 | 185.066 | 478 | 1.84E-06 | 0.095 | 3.07E-03 | 6.42E-09 |
| | S935 | 6 | 2 | 4 | 6 | 36751302 | 37009125 | 257.824 | 672 | 2.02E-07 | 0.095 | 2.45E-03 | 7.05E-10 |
| | S960 | 6 | 2 | 4 | 8 | 65359477 | 65773338 | 413.862 | 785 | 2.02E-07 | 0.095 | 2.45E-03 | 7.05E-10 |
| | S1000 | 6 | 2 | 4 | 11 | 97985598 | 98466352 | 480.755 | 1494 | 2.02E-07 | 0.095 | 2.45E-03 | 7.05E-10 |

| | | | | | | | | | | | | | |
|---|---|---|---|---|---|---|---|---|---|---|---|---|---|
| | S1020 | 6 | 2 | 4 | 13 | 83291736 | 83567001 | 275.266 | 625 | 2.02E-07 | 0.095 | 2.45E-03 | 7.05E-10 |
| | S1118 | 5 | 2 | 3 | 6 | 86802921 | 87341282 | 538.362 | 1289 | 9.30E-09 | 0.095 | 1.84E-03 | 3.25E-11 |
| | S1157 | 5 | 2 | 3 | 8 | 96157210 | 97227476 | 1070.267 | 2080 | 9.30E-09 | 0.095 | 1.84E-03 | 3.25E-11 |
| | S1376 | 4 | 2 | 2 | 11 | 95311792 | 95427262 | 115.471 | 215 | 9.44E-11 | 0.095 | 1.23E-03 | 3.30E-13 |
| **EUR** | S4130 | 10 | 6 | 4 | 1 | 67749103 | 68044287 | 295.185 | 545 | 3.47E-05 | 0.006 | 5.33E-04 | 5.19E-08 |

Legend: AAC = African Admixed, AFR = African, CHR = chromosome, EUR = European, KB = kilobase, NSNP = number of single nucleotide polymorphisms, ROH = runs of homozygosity, SNP = single nucleotide polymorphism. The Bonferroni threshold was adjusted by dividing 0.05 by the number of enriched ROHs to account for multiple comparisons.

**Banner Authors: Global Parkinson's Genetics Program (GP2)**

| Country | Name | Institution | Funders and Disclosures |
|---|---|---|---|
| **Algeria** | Yasser Mecheri | Centre Hospitalo-Universitaire Dr Benbadis Constantine | Nothing to declare |
| **Argentina** | Emilia M Gatto | Sanatorio de la Trinidad Mitre- INEBA | Nothing to declare |
| | Marcelo Kauffman | Hospital JM Ramos Mejia | Nothing to declare |
| | Federico Capparelli | Centro de Educación Médica e Investigaciones Clínicas Norberto Quirno | Nothing to declare |
| **Armenia** | Samson Khachatryan | Somnus Neurology Clinic | Nothing to declare |
| | Zaruhi Tavadyan | Somnus Neurology Clinic | Nothing to declare |
| | Mariam Isayan | Somnus Neurology Clinic | Nothing to declare |
| **Australia** | Claire E Shepherd | Neuroscience Research Australia | The Sydney Brain Bank is located at and supported by Neuroscience Research Australia |
| | Kishore Kumar | Garvan Institute of Medical Research and Concord Repatriation General Hospital | Paul Ainsworth Family Foundation |
| | Melina Ellis | Concord Hospital | Nothing to declare |
| | Miguel E. Rentería | QIMR Berghofer Medical Research Institute | The Australian Parkinson's Genetics Study is supported by the Shake It Up Australia Foundation and The Michael J. Fox Foundation for Parkinson's Research |
| | Sulev Koks | Murdoch University | Nothing to declare |
| | Simon Rowe | Neuroscience Research Australia | Nothing to declare |
| | Laura Rudaks | Neuroscience Research Australia | Nothing to declare |
| | Dennis Yeow | Neuroscience Research Australia | Nothing to declare |
| | Carolyn Sue | Neuroscience Research Australia | Nothing to declare |

| | | | |
|---|---|---|---|
| | Victor Flores Ocampo | QIMR Berghofer Medical Research Institute | Nothing to declare |
| **Austria** | Alexander Zimprich | Medical University Vienna Austria | Nothing to declare |
| **Azerbaijan** | Kanan Jafarov | Istanbul Klinik | Nothing to declare |
| **Belgium** | David Crosiers | University of Antwerp | Nothing to declare |
| **Brazil** | Artur F. Schumacher-Schuh | Universidade Federal do Rio Grande do Sul / Hospital de Clínicas de Porto Alegre | Nothing to declare |
| | Carlos Rieder | Federal University of Health Sciences of Porto Alegre | Nothing to declare |
| | Paula Saffie Awad | Universidade Federal do Rio Grande do Sul | Global Parkinson's Genetics Program |
| | Vitor Tumas | University of São Paulo | Nothing to declare |
| | Sarah Camargos | Universidade Federal de Minas Gerais | Nothing to declare |
| | Lucas Faria Costa | Universidade Federal de Minas Gerais | Nothing to declare |
| **Canada** | Edward A. Fon | Montreal Neurological Institute | Nothing to declare |
| | Oury Monchi | Institut universitaire de gériatrie de Montréal | CIHR, Brain Canada, Parkinson Canada |
| | Edward Fon | McGill University | Nothing to declare |
| | Robert Thibault | Aligning Science Across Parkinson's | Nothing to declare |
| | Ziv Gan-Or | McGill University | Nothing to declare |
| | Anthony Lang | University of Toronto | Nothing to declare |
| | Konstantin Senkevich | McGill University | Nothing to declare |
| **Chile** | Benjamin Pizarro Galleguillos | Universidad de Chile | Nothing to declare |
| | Marcelo Miranda | Fundación Diagnosis | Nothing to declare |
| | Maria Leonor Bustamante | Faculty of Medicine Universidad de Chile | Nothing to declare |
| | Patricio Olguin | Universidad de Chile | Nothing to declare |

| | | | |
|---|---|---|---|
| | Pedro Chana | CETRAM | Nothing to declare |
| | Elias Fernandez | Hospital Regional de Concepción | Nothing to declare |
| **China** | Beisha Tang | Central South University | Nothing to declare |
| | Huifang Shang | West China Hospital Sichuan University | Nothing to declare |
| | Jifeng Guo | Xiangya Hospital | Nothing to declare |
| | Piu Chan | Capital Medical University | Nothing to declare |
| | Wei Luo | Zhejiang University | Nothing to declare |
| | Zhenhua Liu | Xiangya Hospital, Central South University | Nothing to declare |
| **Colombia** | Gonzalo Arboleda | Universidad Nacional de Colombia | Nothing to declare |
| | Jorge Orozco | Fundación Valle del Lili | Nothing to declare |
| | Marlene Jimenez del Rio | University of Antioquia | Nothing to declare |
| **Costa Rica** | Alvaro Hernandez | University of Costa Rica | Nothing to declare |
| **Denmark** | Per Borghammer | Aarhus University | Nothing to declare |
| **Egypt** | Mohamed Salama | The American University in Cairo | The AUC/ ASRT/ DAAD |
| | Walaa A. Kamel | Beni-Suef University | Nothing to declare |
| **Ethiopia** | Yared Z. Zewde | Addis Ababa University | Nothing to declare |
| **France** | Alexis Brice | Paris Brain Institute | Nothing to declare |
| | Jean-Christophe Corvol | Sorbonne Université | Nothing to declare |
| | Mari Vidailhet | Salpêtriere Hospital (AP-HP), Sorbonne Université | Nothing to declare |
| **Georgia** | Mariam Kekenadze | Tbilisi State Medical University | Nothing to declare |
| | Irine Khatiashvili | S. Khechinashvili University Hospital | Nothing to declare |
| **Germany** | Ana Westenberger | University of Lübeck | Nothing to declare |

| | | |
|---|---|---|
| Anastasia Illarionova | Deutsches Zentrum für Neurodegenerative Erkrankungen | Nothing to declare |
| Brit Mollenhauer | University Medical Center Göttingen | Nothing to declare |
| Christine Klein | University of Lübeck | CK serves as a medical Advisor to Centogene on genetic testing reports in the field of movement disorders, except Parkinson's disease, and is a member of the Scientific Advisory Board of Retromer Therapeutics |
| Eva-Juliane Vollstedt | University of Lübeck | Nothing to declare |
| Franziska Hopfner | Department of Neurology, University Hospital, LMU Munich | Nothing to declare |
| Günter Höglinger | Department of Neurology, University Hospital, LMU Munich | Nothing to declare |
| Harutyun Madoev | University of Lübeck | Nothing to declare |
| Joanne Trinh | University of Lübeck | Nothing to declare |
| Katja Lohmann | University of Lübeck | Nothing to declare |
| Lara M. Lange | University of Lübeck and University Medical Center Schleswig-Holstein | Nothing to declare |
| Manu Sharma | University of Tubingen | Dr. Sharma is further funded by the Michael J Fox Foundation, USA Genetic Diversity in PD Program: GAP-India Grant ID: 009411. |
| Sergiu Groppa | University of Mainz | Nothing to declare |
| Thomas Gasser | University of Tübingen | Nothing to declare |
| Zih-Hua Fang | The German Center for Neurodegenerative Diseases | Nothing to declare |
| Karl Heilbron | Charité - Universitätsmedizin Berlin | Former employee of 23andMe, Inc. Owns stock and/or stock options in 23andMe, Inc. |
| Theresa Luth | University of Luebeck | Nothing to declare |
| Wenhua Sun | University of Teubingen | Nothing to declare |

| | | | |
|---|---|---|---|
| | Inke König | University of Lübeck | Nothing to declare |
| | Daniela Berg | University Medical Center Schleswig-Holstein | Nothing to declare |
| | Bernhard Haslinger | Technical University of Munich | Nothing to declare |
| | Teresa Kleinz | University of Lübeck | Nothing to declare |
| | Norbert Brüggemann | University of Lübeck | Dr. Brüggemann received honaria from Abbvie, Esteve, Ipsen, Merz, Teva and Zambon |
| **Ghana** | Albert Akpalu | University of Ghana Medical School | Nothing to declare |
| **Greece** | Georgia Xiromerisiou | University of Thessaly | Nothing to declare |
| | Georgios Hadjigorgiou | University of Thessaly | Nothing to declare |
| | Ioannis Dagklis | Aristotle University of Thessaloniki | Nothing to declare |
| | Ioannis Tarnanas | Ionian University | Nothing to declare |
| | Leonidas Stefanis | Biomedical research Foundation of the Academy of Athens | PPMI2 (funded by MJFF), ALAMEDA (H2020 grant), funding by HFRI |
| | Maria Stamelou | Diagnostic and Therapeutic Centre HYGEIA Hospital | The non-profit organisation for scientific research in Parkinson’s disease and related disorders |
| | Efthymios Dadiotis | University of Thessaly | Nothing to declare |
| **Honduras** | Alex Medina | Hospital San Felipe | Nothing to declare |
| **Hong Kong** | Germaine Hiu-Fai Chan | Queen Elizabeth Hospital | Nothing to declare |
| | Nancy Ip | The Hong Kong University of Science and Technology | Nothing to declare |
| | Nelson Yuk-Fai Cheung | Queen Elizabeth Hospital | Nothing to declare |
| | Phillip Chan | The Hong Kong University of Science and Technology | Nothing to declare |

| | | | |
|---|---|---|---|
| | Xiaopu Zhou | The Hong Kong University of Science and Technology | Nothing to declare |
| **India** | Asha Kishore | Aster Medcity | Michael J. Fox Foundation |
| | Divya KP | Sree Chitra Tirunal Institute for Medical Sciences and Technology | Nothing to declare |
| | Pramod Pal | National Institute of Mental Health & Neurosciences | Nothing to declare |
| | Prashanth Lingappa Kukkle | Manipal Hospital | Nothing to declare |
| | Roopa Rajan | All India Institute of Medical Sciences | Nothing to declare |
| | Rupam Borgohain | Nizam's Institute Of Medical Sciences | Nothing to declare |
| **Iran** | Mehri Salari | Shahid Beheshti University of Medical Science | Nothing to declare |
| **Israel** | Tamara Shiner | Tel Aviv Sourasky Medical Center | Nothing to declare |
| | Avner Thaler | Tel Aviv Sourasky Medical Center | Nothing to declare |
| **Italy** | Andrea Quattrone | Magna Græcia University of Catanzaro | Nothing to declare |
| | Enza Maria Valente | University of Pavia | Nothing to declare |
| | Grazia Annesi | National Research Council | Nothing to declare |
| | Lucilla Parnetti | University of Perugia | Nothing to declare |
| | Micol Avenali | University of Pavia | Nothing to declare |
| | Monica Gagliardi | Magna Graecia University | Nothing to declare |
| | Tommaso Schirinzi | University of Rome Tor Vergata | Nothing to declare |
| | Caterina Galandra | IRCCS Mondino Foundation | Nothing to declare |
| | Anna De Rosa | University of Naples Federico II | Nothing to declare |
| **Japan** | Manabu Funayama | Juntendo University | Nothing to declare |
| | Nobutaka Hattori | Juntendo University faculty of medicine | Nothing to declare |

| | | | |
|---|---|---|---|
| | Tomotaka Shiraishi | Jikei University School of Medicine | Nothing to declare |
| **Kazakhstan** | Altynay Karimova | Institute of Neurology and Neurorehabilitation | Nothing to declare |
| | Gulnaz Kaishibayeva | Institute of Neurology and Neurorehabilitation | Nothing to declare |
| | Aigerim Utegenova | West Kazakhstan Marat Ospanov State Medical University | Nothing to declare |
| | Aigul Yemagambetova | West Kazakhstan Marat Ospanov State Medical University | Nothing to declare |
| | Vadim Akhmetzhanov | Medline medical center | Nothing to declare |
| | Seitzhan Aidarov | National Center for Neurosurgery | Nothing to declare |
| | Tautanova Raushan | Astana Medical University | Nothing to declare |
| | Bagyzhan Syzdykova Syzdykova | City multi-field hospital No. 1 | Nothing to declare |
| | Dinara Alzhanova | Astana Medical University | Nothing to declare |
| | Zhanybek Myrzayev | International University of Postgraduate Education | Nothing to declare |
| | Saltanat Abdraimova | South Kazakhstan Medical Academy | Nothing to declare |
| **Kyrgyzstan** | Cholpon Shambetova | Kyrgyz State Medical Academy | Nothing to declare |
| **Luxembourg** | Rejko Krüger | University of Luxembourg | Nothing to declare |
| | Patrick May | University of Luxembourg | Nothing to declare |
| **Malaysia** | Ai Huey Tan | University of Malaya | Nothing to declare |
| | Azlina Ahmad-Annuar | University of Malaya | Nothing to declare |
| | Mohamed Ibrahim Norlinah | Universiti Kebangsaan Malaysia | Nothing to declare |
| | Nor Azian Abdul Murad | UKM Medical Molecular Biology Institute | Nothing to declare |

| | | | |
|---|---|---|---|
| | Shahrul Azmin | Universiti Kebangsaan Malaysia Medical Centre | Nothing to declare |
| | Shen-Yang Lim | University of Malaya | Nothing to declare |
| | Wael Mohamed | International Islamic University | Nothing to declare |
| | Yi Wen Tay | University of Malaya | Nothing to declare |
| | Azalea Tenerife Pajo | University of Malaya | Nothing to declare |
| **Mexico** | Daniel Martinez-Ramirez | Tecnologico de Monterrey | Nothing to declare |
| | Mayela Rodriguez-Violante | Instituto Nacional de Neurologia y Neurocirugia | Nothing to declare |
| | Paula Reyes-Pérez | Universidad Nacional Autónoma de México | Global Parkinson's Genetics Program |
| | Alejandra Medina Rivera | Universidad Nacional Autónoma de México | Nothing to declare |
| | Nancy Monroy Jaramillo | Instituto Nacional de Neurología y Neurocirugía | Nothing to declare |
| **Mongolia** | Bayasgalan Tserensodnom | Mongolian National University of Medical Sciences | Nothing to declare |
| **Nepal** | Rajeev Ojha | Tribhuvan University | Nothing to declare |
| **Netherlands** | Wilma Van De Berg | Vanderbilt University Medical Center | Nothing to declare |
| | Bas Bleom | Radboud University | Nothing to declare |
| | Bart Van de Warrenburg | Radboud University Medical Center | Nothing to declare |
| | Lisette Charbonnier | Brain Research and Innovation Center | Nothing to declare |
| **New Zealand** | Tim J. Anderson | University of Otago | Health Research Council of New Zealand; Ministry of Business Innovation and Employment (New Zealand), Neurological Foundation of New Zealand |
| | Toni L. Pitcher | University of Otago | Funding - Health Research Council of New Zealand |

| | | | |
|---|---|---|---|
| **Nigeria** | Arinola Sanyaolu | University of Lagos | Nothing to declare |
| | Njideka Okubadejo | University of Lagos | Michael J Fox Foundation; Tertiary Education Trust Fund (TETFUND) National Research Fund |
| | Oluwadamilola Ojo | College of Medicine of the University of Lagos | Nothing to declare |
| | Simon Izuchukwu Ozomma | University of Calabar Teaching Hospital | Nothing to declare |
| | Kolawole Wahab | University of Ilorin | Nothing to declare |
| **Norway** | Lasse Pihlstrøm | Oslo University Hospital | Southeastern Regional Health Authority, Norway and Michael J. Fox Foundation |
| | Manuela Tan | Oslo University Hospital | Southeastern Norway Regional Health Authority Norway and Michael J. Fox Foundation |
| | Ingeborg Haugesag Lie | Oslo University Hospital | Nothing to declare |
| | Jodi Maple-Grødem | Stavanger University Hospital | Nothing to declare |
| **Pakistan** | Shoaib Ur-Rehman | University of Science and Technology Bannu | Nothing to declare |
| **Peru** | Mario Cornejo-Olivas | Universidad Cientifica del Sur | Michael J. Fox Foundation for Parkinson's Research and Aligning Science Across Parkinson's Initiative |
| **Philippines** | Maria Leila Doquenia | Metropolitan Medical Center | Nothing to declare |
| | Raymond Rosales | Metropolitan Medical Center | Nothing to declare |
| **Puerto Rico** | Angel Vinuela | University of Puerto Rico | Nothing to declare |
| **Russia** | Elena Iakovenko | Research Center of Neurology | Nothing to declare |
| | Anna Gareeva | Ufa Federal Research Center | Nothing to declare |
| | Gulnara Akhmadeeva | Ufa Scientific Center | Nothing to declare |

| | | | |
|---|---|---|---|
| | Irina Gilyazova | Russian Academy of Sciences / Bashkir State Medical University | Nothing to declare |
| **Saudi Arabia** | Bashayer Al Mubarak | King Faisal Specialist Hospital and Research Center | Nothing to declare |
| | Muhammad Umair | King Abdullah International Medical Research Center | Nothing to declare |
| **Singapore** | Eng-King Tan | National Neuroscience Institute | Singapore National Medical Research Council (MOH-OFLCG-000207) |
| | Jia Nee Foo | Nanyang Technological University | Singapore National Medical Research Council (MOH-000559) |
| | Elaine Chew | Nanyang Technological University | Nothing to declare |
| **Slovenia** | Vesna van Midden | Ljubljana University Medical Centre | Nothing to declare |
| **South Africa** | Ferzana Amod | University of KwaZulu-Natal | Nothing to declare |
| | Jonathan Carr | University of Stellenbosch | Nothing to declare |
| | Soraya Bardien | Stellenbosch University | National Research Foundation of South Africa[Grant Number 129249] |
| | Nikita Pillay | University of the Western Cape | Nothing to declare |
| | Kathryn Step | Stellenbosch University | Nothing to declare |
| **South Korea** | Beomseok Jeon | Seoul National University Hospital | Nothing to declare |
| | Yun Joong Kim | Yongin Severance Hospital | Nothing to declare |
| | Jung Hwan Shin | Seoul National University | Nothing to declare |
| | Joowon Jang | Seoul National University | Nothing to declare |
| **Spain** | Esther Cubo | Hospital Universitario Burgos | Nothing to declare |
| | Ignacio Alvarez | University Hospital Mutua Terrassa | Nothing to declare |

| | | | |
|---|---|---|---|
| | Janet Hoenicka | Institut de Recerca Sant Joan de Deu | Fondo de Investigación Sanitaria, Instituto Salud Carlos III, Grant PI019/00126 |
| | Katrin Beyer | Research Institute Germans Trias i Pujol | Nothing to declare |
| | Maria Teresa Periñan | Instituto de Biomedicina de Sevilla | Nothing to declare |
| | Pau Pastor | University Hospital Germans Trias i Pujol | Nothing to declare |
| | Ruben Fernandez-Santiago | Hospital Clínic de Barcelona | Nothing to declare |
| | Pilar Gómez Garre | Instituto de Biomedicina de Sevilla | Nothing to declare |
| | Pablo Mir | Instituto de Biomedicina de Sevilla | Nothing to declare |
| **Sudan** | Sarah El-Sadig | Faculty of medicine university of Khartoum | Nothing to declare |
| **Sweden** | Kajsa Brolin | Lund University | Nothing to declare |
| | Per Svenningsson | Karolinska Institute | Nothing to declare |
| | Maria Swanberg | Lund University | Nothing to declare |
| **Switzerland** | Christiane Zweier | Inselspital Bern, University of Bern | Nothing to declare |
| | Gerd Tinkhauser | University Hospital Bern | Nothing to declare |
| | Paul Krack | Inselspital Bern, University of Bern | Nothing to declare |
| **Taiwan** | Chin-Hsien Lin | National Taiwan University Hospital | Nothing to declare |
| | Hsiu-Chuan Wu | Chang Gung Memorial Hospital | Nothing to declare |
| | Pin-Jui Kung | National Taiwan University | Global Parkinson's Genetics Program |
| | Ruey-Meei Wu | National Taiwan University Hospital | Minister of Science and Technology, Taiwan Government; National Taiwan University; Parkinson foundation (USA); Michael J. Fox Foundation |
| | Yihru Wu | Chang Gung Memorial Hospital | Nothing to declare |

| | | | |
|---|---|---|---|
| **Tajikistan** | Ganieva Manizha | Avicenna Tajik State Medical University | Nothing to declare |
| **Tunisia** | Rim Amouri | National Institute Mongi Ben Hamida of Neurology | Nothing to declare |
| | Samia Ben Sassi | Mongi Ben Hmida National Institute of Neurology | Nothing to declare |
| **Turkey** | A. Nazlı Başak | Koç University | Kirac Foundation and Koc Univ. |
| | Gencer Genc | Şişli Etfal Training and Research Hospital | Nothing to declare |
| | Özgür Öztop Çakmak | Koç University | Nothing to declare |
| | Sibel Ertan | Koç University | Nothing to declare |
| **United Kingdom** | Alastair Noyce | Queen Mary University of London | Prof. Noyce reports grants from Parkinson's UK, Barts Charity, Cure Parkinson's, NIHR, Innovate UK, Virginia Keiley benefaction, Alchemab, Aligning Science Across Parkinson's and Michael J Fox Foundation. Consultancy and personal fees from Astra Zeneca, AbbVie, Profile, Roche, Biogen, UCB, Bial, Charco Neurotech, uMedeor and Britannia. |
| | Alejandro Martínez-Carrasco | University College London | Global Parkinson's Genetics Program |
| | Anette Schrag | University College London | NIHR UCL/H Biomedical Research Centre |
| | Anthony Schapira | University College London | ASAP |
| | Camille Carroll | University of Plymouth | C Carroll receives salary from University of Plymouth, University Hospitals Plymouth NHS Trust and National Institute of Health Research; she has received advisory, consulting, and/or lecture fees from AbbVie, |

| | | | |
|---|---|---|---|
| | | | Bial, Lundbeck, Global Kinetics, Britannia and Medscape, and research funding from Parkinson's UK, Edmond J Safra Foundation, National Institute of Health Research and Cure Parkinson’s |
| | Donald Grosset | University of Glasgow | Tracking Parkinson's (J-1101) is funded by Parkinson's UK |
| | Eleanor J. Stafford | University College London | Nothing to declare |
| | Henry Houlden | University College London | Nothing to declare |
| | Huw R Morris | University College London | Dr Morris is employed by UCL. In the last 12 months he reports paid consultancy from Roche and Amylyx ; lecture fees/honoraria - BMJ, Kyowa Kirin, Movement Disorders Society. Research Grants from Parkinson’s UK, Cure Parkinson’s Trust, PSP Association, CBD Solutions, Drake Foundation, Medical Research Council, Michael J Fox Foundation. Dr Morris is a co-applicant on a patent application related to C9ORF72 - Method for diagnosing a neurodegenerative disease (PCT/GB2012/052140) |
| | John Hardy | University College London | Nothing to declare |
| | Kin Ying Mok | Univeristy College London | Nothing to declare |
| | Mie Rizig | University College London | Nothing to declare |
| | Nicholas Wood | University College London | ASAP- CRN |
| | Nigel Williams | Cardiff University | Parkinson's UK |

| | | | |
|---|---|---|---|
| | Olaitan Okunoye | University College London | Nothing to declare |
| | Rauan Kaiyrzhanov | University College London | Nothing to declare |
| | Rimona Weil | University College London | Nothing to declare |
| | Seth Love | University of Bristol | Medical Research Council (UK) MR/T018569/1 |
| | Simona Jasaityte | University College London | Nothing to declare |
| | Sumit Dey | Queen Mary University of London | The Michael J. Fox Foundation for Parkinson's Research |
| | Vida Obese | University College London | Nothing to declare |
| | Spencer Finch | Queen Mary University of London | Nothing to declare |
| | Valentina Escott-Price | Cardiff University | Nothing to declare |
| | Hamin Lee | St George's, University of London | Nothing to declare |
| | Roger Barker | University of Cambridge | Nothing to declare |
| | Mina Ryten | University College London | Nothing to declare |
| | Michele Hu | University of Oxford | Nothing to declare |
| | Laura Parkkinen | University of Oxford | Nothing to declare |
| | Kailash Bhatia | University College London | Nothing to declare |
| | Richard Walker | Northumbria Healthcare at NHS Foundation Trust | Nothing to declare |
| | Steve Gentleman | Imperial College London | Nothing to declare |
| | Thomas Warner | University College London | Nothing to declare |
| | David Burn | Newcastle University | Nothing to declare |
| | Christian Lambert | Imperial College London | Nothing to declare |

| | | | |
|---|---|---|---|
| | Caroline Williams-Gray | University of Cambridge | Nothing to declare |
| | Chris Morris | Newcastle University | Newcastle University, Alzheimer's Society, Alzheimer's Research UK, Medical Research Council |
| | Deborah Attuah | YLD | Nothing to declare |
| | Raquel Real | University College London | Nothing to declare |
| | Yen Tai | Imperial College London | Nothing to declare |
| **USA** | Alberto Espay | University of Cincinnati | Nothing to declare |
| | Alyssa O'Grady | The Michael J. Fox Foundation for Parkinson's Research | Nothing to declare |
| | Andrew B Singleton | National Institute on Aging | Michael J Fox Foundation for Parkinson's disease Research and Aligning Science Across Parkinson's Initiative |
| | Andrew K. Sobering | Augusta University / University of Georgia Medical Partnership | Nothing to declare |
| | Bernadette Siddiqi | The Michael J. Fox Foundation for Parkinson's Research | Nothing to declare |
| | Bradford Casey | The Michael J. Fox Foundation for Parkinson's Research | Nothing to declare |
| | Brian Fiske | The Michael J. Fox Foundation for Parkinson's Research | Nothing to declare |
| | Cabell Jonas | Mid-Atlantic Permanente Medical Group | Nothing to declare |
| | Carlos Cruchaga | Washington University | National Institutes of Health (R01AG044546 (CC), P01AG003991(CC, JCM), RF1AG053303 (CC), RF1AG058501 (CC), U01AG058922 (CC), RF1AG074007 (YJS)), the Chuck Zuckerberg Initiative (CZI), the Michael J. Fox Foundation (LI, CC), and the |

| | | | |
|---|---|---|---|
| | | | Department of Defense (LI-W81XWH2010849). The recruitment and clinical characterization of research participants at Washington University were supported by NIH P30AG066444 (JCM), P01AG03991(JCM), and P01AG026276(JCM). |
| | Caroline B. Pantazis | National Institutes of Health | Nothing to declare |
| | Charisse Comart | The Michael J. Fox Foundation for Parkinson's Research | Nothing to declare |
| | Claire Wegel | Indiana University | Nothing to declare |
| | Cornelis Blauwendraat | National Institutes of Health | Nothing to declare |
| | Dan Vitale | National Institutes of Health | Nothing to declare |
| | Deborah Hall | Rush University | Nothing to declare |
| | Dena Hernandez | National Institutes of Health | Nothing to declare |
| | Ejaz Shiamim | Kaiser Permanente | Nothing to declare |
| | Ekemini Riley | Coalition for Aligning Science | Nothing to declare |
| | Faraz Faghri | National Institutes of Health | F.F.'s participation in this research was supported in part by the Intramural Research Program of the NIH, National Institute on Aging (NIA), National Institutes of Health, Department of Health and Human Services; project number ZO1 AG000535, as well as the National Institute of Neurological Disorders and Stroke. F.F.'s participation in this project was part of a competitive contract awarded to Data |

| | | | |
|---|---|---|---|
| | | | Tecnica International LLC by the National Institutes of Health to support open science research. |
| | Geidy E. Serrano | Banner Sun Health Research Institute | Banner Sun Health Research Institute Brain and Body Donation Program of Sun City, Arizona for the provision of human biological materials. The Brain and Body Donation Program has been supported by the National Institute of Neurological Disorders and Stroke (U24 NS072026 National Brain and Tissue Resource for Parkinson's Disease and Related Disorders), the National Institute on Aging (P30 AG19610 and P30AG072980, Arizona Alzheimer's Disease Center), the Arizona Department of Health Services (contract 211002, Arizona Alzheimer's Research Center), the Arizona Biomedical Research Commission (contracts 4001, 0011, 05-901 and 1001 to the Arizona Parkinson's Disease Consortium) and the Michael J. Fox Foundation for Parkinson's Research |
| | Hampton Leonard | National Institute on Aging/National Institutes of Health | H.L.L is supported by a competitive contract awarded to Data Tecnica International LLC by the National |

| | | | |
|---|---|---|---|
| | | | Institutes of Health to support open science research |
| | Hirotaka Iwaki | Data Tecnica International | H.I is supported by a competitive contract awarded to Data Tecnica International LLC by the National Institutes of Health to support open science research |
| | Honglei Chen | Michigan State University | NIH/DoD/Parkinson Foundation/MSU Foundation/Gibby vs. Parky Foundation - No COI to disclose |
| | Ignacio F. Mata | Cleveland Clinic | Funding from MJFF and NIH |
| | Ignacio Juan Keller Sarmiento | Northwestern University | Nothing to declare |
| | Jared Williamson | Kaiser Permanente | Nothing to declare |
| | Jonggeol Jeff Kim | National Institutes of Health | Nothing to declare |
| | Joseph Jankovic | Baylor College of Medicine | Nothing to declare |
| | Joshua Shulman | Baylor College of Medicine / Texas Children's Hospital | Collection of samples and data at Baylor College of Medicine was supported by Huffington Foundation. |
| | Justin C. Solle | The Michael J. Fox Foundation for Parkinson's Research | Nothing to declare |
| | Kaileigh Murphy | The Michael J. Fox Foundation for Parkinson's Research | Nothing to declare |
| | Kamalini Ghosh Galvelis | Parkinson's Foundation | Nothing to declare |
| | Karen Nuytemans | University of Miami Miller School of Medicine | This work has been supported by the American Parkinson Disease Association and the Margaret Q. Landenberger Research Foundation. |

| | | | |
|---|---|---|---|
| | Karl Kieburtz | Beth Israel Deaconess Medical Center | Nothing to declare |
| | Kate Andersh | National Institute on Aging | Nothing to declare |
| | Katerina Markopoulou | North Shore University Health System | Nothing to declare |
| | Kenneth Marek | Institute for Neurodegenerative Disorders | Consultant for Michael J Fox Foundation, GE Healthcare, Roche, UCB, BIAL, Denali, Takeda, , Cerapsir, UCB, Biohaven, Neuron23, Aprinoia, Astellas, Calico, Genentech, Invicro |
| | Kristin S. Levine | Data Tecnica International | Nothing to declare |
| | Lana M. Chahine | University of Pittsburgh | Nothing to declare |
| | Laura Ibanez | Washington University | Nothing to declare |
| | Laurel Screven | National Institute on Aging | Nothing to declare |
| | Lauren Ruffrage | University of Alabama at Birmingham | Nothing to declare |
| | Lisa Shulman | University of Maryland | Nothing to declare |
| | Luca Marsili | University of Cincinnati | Nothing to declare |
| | Maggie Kuhl | The Michael J. Fox Foundation for Parkinson's Research | Nothing to declare |
| | Marissa Dean | University of Alabama at Birmingham | Dr. Dean is an investigator in studies funded by Abbvie, Inc., Hoffmann-La Roche, CHDI Foundation, Inc., Annexon, Inc., Retrophin, Inc, Neurocrine Biosciences, UniQure Biopharma B.V., Praxis Precision Medicines, Neuraly, Inc., Michael J. Fox Foundation for Parkinson’s Research, and US Army Medical Research and Material Command (grant#W81XWH-18-1- |

| | | | |
|---|---|---|---|
| | | | 0508). In addition, Dr. Dean receives support through the Huntington's Disease Society of American Centers of Excellence program. |
| | Mary B Makarious | National Institutes of Health | Nothing to declare |
| | Matthew Farrer | University of Florida - Neurology | Nothing to declare |
| | Mathew Koretsky | National Institutes of Health | Nothing to declare |
| | Megan J. Puckelwartz | Northwestern University | Nothing to declare |
| | Miguel Inca-Martinez | Cleveland Clinic | Nothing to declare |
| | Mike A. Nalls | National Institutes of Health | M.A.N.'s participation in this project was part of a competitive contract awarded to Data Tecnica International LLC by the National Institutes of Health to support open science research. M.A.N. also currently serves as an advisor for Clover Therapeutics and Neuron23 Inc. |
| | Naomi Louie | The Michael J. Fox Foundation for Parkinson's Research | Nothing to declare |
| | Niccolò Emanuele Mencacci | Northwestern University | Nothing to declare |
| | Roger Albin | Universit of Michigan | P50NS123067; Parkinson's Foundation |
| | Roy Alcalay | Columbia University | Dr. Alcalay is funded by the Micharl J. Fox Foundation and the Parkinson's Foundation. He received consultation fees from Avrobio, Caraway, GSK, Merck, Sanofi, Ono Therapeutics and Takeda |

| | | | |
|---|---|---|---|
| | Ruth Walker | James J. Peters Veterans Affairs Medical Center | Research funding from the Department of Veterans Affairs (CSRD Merit Award CX002342), consulted for Teladoc, Inc. and received an honorarium from New York University (NYU) Medical Center. |
| | Sara Bandres-Ciga | National Institutes of Health | Nothing to declare |
| | Sohini Chowdhury | The Michael J. Fox Foundation for Parkinson's Research | Nothing to declare |
| | Sonya Dumanis | Aligning Science Across Parkinson's | Nothing to declare |
| | Steven Lubbe | Northwestern University | Nothing to declare |
| | Tao Xie | University of Chicago | Nothing to declare |
| | Tatiana Foroud | Indiana University School of Medicine | The Michael J. Fox Foundation for Parkinson's Research |
| | Thomas Beach | Sun Health Research Institution | Banner Sun Health Research Institute Brain and Body Donation Program of Sun City, Arizona for the provision of human biological materials. The Brain and Body Donation Program has been supported by the National Institute of Neurological Disorders and Stroke (U24NS072026 National Brain and Tissue Resource for Parkinson’s Disease and Related Disorders), the National Institute on Aging (P30 AG19610 and P30AG072980, Arizona Alzheimer’s Disease Center), the Arizona Department of Health Services (contract 211002, Arizona Alzheimer’s |

| | | | |
|---|---|---|---|
| | | | Research Center), the Arizona Biomedical Research Commission (contracts 4001, 0011, 05-901 and 1001 to the Arizona Parkinson's Disease Consortium) and the Michael J. Fox Foundation for Parkinson’s Research |
| | Todd Sherer | The Michael J Fox Foundation for Parkinson's Research | Nothing to declare |
| | Yeajin Song | National Institutes of Health | Nothing to declare |
| | Dana Lewis | Aligning Science Across Parkinson's | Nothing to declare |
| | Shreya Menon | Gladstone Institutes | Nothing to declare |
| | Melissa Nirenberg | Icahn School of Medicine at Mount Sinai | Nothing to declare |
| | Spencer Grant | National Institutes of Health | Nothing to declare |
| | Shannon Ballard | National Institutes of Health | Nothing to declare |
| | Chad Shaw | Baylor College of Medicine | Nothing to declare |
| | Sidra Aslam | Banner Health | Nothing to declare |
| | Geidy Serrano | Banner Sun Health Research Institute | Nothing to declare |
| | Devin Sharp | Aligning Science Across Parkinson's | Nothing to declare |
| | Kensuke Daida | National Institute of Aging | Nothing to declare |
| | Rachel Saunders-Pullman | Icahn School of Medicine at Mount Sinai | Nothing to declare |
| | Michiko Kimura Bruno | The Queen's Medical Center | Nothing to declare |
| | Matt Farrer | University of Florida College of Medicine | Nothing to declare |
| | Haydeh Payami | The University of Alabama at Birmingham Heersink School of Medicine | Nothing to declare |
| | Ryan Pfingst | The Michael J Fox Foundation | Nothing to declare |

| | | | |
|---|---|---|---|
| | James B Leverenz | Cleveland Clinic | Nothing to declare |
| | Elizabeth Disbrow | LSU Health Shreveport | Nothing to declare |
| | Debi Brooks | The Michael J Fox Foundation | Nothing to declare |
| | Randy Schekman | University of California, Berkeley | Nothing to declare |
| | Un Kang | NYU Grossman School of Medicine | Nothing to declare |
| | Zbigniew K. Wszolek | Mayo Clinic College of Medicine | Nothing to declare |
| | Cyrus Zabetian | VA Puget Sound Health Care System | Nothing to declare |
| | Zach Chaney | The Michael J Fox Foundation | Nothing to declare |
| | Mark Cookson | National Institute of Health | Nothing to declare |
| | Christine Swanson-Fischer | National Institute of Health | Nothing to declare |
| | Conor Hennessey | The Michael J Fox Foundation | Nothing to declare |
| | Cassandra Barrett | The Michael J Fox Foundation | Nothing to declare |
| | Beate Ritz | University of California, Los Angeles | Nothing to declare |
| | Bradley Boeve | Mayo Clinic | Nothing to declare |
| | Ashley Rawls | University of Florida College of Medicine | Nothing to declare |
| **Vietnam** | Duan Nguyen | Hue University | Nothing to declare |
| | Toan Nguyen | Hue University | Nothing to declare |
| **Zambia** | Masharip Atadzhanov | University of Zambia | Nothing to declare |
